# Evidence of a causal relationship between genetic tendency to gain muscle mass and uterine leiomyomata

**DOI:** 10.1101/2021.08.20.21262098

**Authors:** Eeva Sliz, Jaakko Tyrmi, Nilufer Rahmioglu, Krina T. Zondervan, Christian M. Becker, FinnGen, Outi Uimari, Johannes Kettunen

## Abstract

Uterine leiomyomata (UL) are the most common benign tumours of the female genital tract with an estimated lifetime incidence of up to 70%^1^. To date, 7 genome-wide association studies (GWASs) have identified 35 loci predisposing to UL. To improve the understanding of the underlying genetic pathways, we conducted the largest genetic association study of UL to date in 426,558 European women from FinnGen and a previously published UL meta-GWAS^2^. We identified 36 novel and replicated 31 previously reported loci. Annotations of the potential candidate genes suggest involvement of smooth muscle cell (SMC) differentiation and/or proliferation-regulating genes in modulating UL risk. Our results further advocate that genetic predisposition to increased fat-free mass may be causally related to UL risk, underscoring the involvement of altered muscle tissue biology in UL pathophysiology. Overall, our findings provide novel insights into genetic risk factors related to UL, which may aid in developing novel treatment strategies.

---

UL are hormone-driven benign neoplasms of the uterus composed mostly of SMC and fibroblasts with a profound component of extracellular matrix (ECM). UL are present in single or multiple numbers, with size ranging from millimeters to 20 cm or more in diameter^3^. In 25-50% of women with ULs, the enlarged and deformed uterus can cause symptoms such as heavy or prolonged menstrual bleeding resulting in anemia, reduced fertility, and pregnancy complications^4^. Major risk factors for UL are family history, African ancestry, increasing age up to menopause, and nulliparity^5^. Also, metabolic factors, including hypertension and increased body mass index (BMI), have been reported in association with UL susceptibility^6^. Familial aggregation, twin studies, and disparity in prevalence between different ethnic groups suggest that genetic factors modulate UL risk^7-10^.

Until recently, the focus in the genetics of UL has been on somatic rearrangements, and key driver variations in *MED12, FH, HMGA2* and *COL4A6*-*COL4A5* have been reported^11^. To add understanding to the heritable genetic underpinnings, we conducted two sets of fixed-effect, inverse-variance weighted meta-analyses with data from FinnGen (18,060 cases and 105,519 female controls) and 1) a previously published UL meta-GWAS^2^ totaling 53,534 cases and 373,024 female controls in ‘META-1’ and 2) the UK Biobank totaling 33,244 cases and 311,271 controls in ‘META-2’. META-1 was restricted to publicly available 10,000 variants from the previous meta-GWAS^2^, wheras the META-2 included 10,693,588 variants across the genome. In META-1, we identified 63 loci associating with UL at *p*<5×10^−8^ (Figure 1, Table S1); of these, 32 were novel (Table 1). In META-2, we identified 51 loci out of which 4 had not been associated with UL risk in prior GWASs or in META-1 (Figure 1, Table 1, Table S2). Regional association plots of the novel loci are presented in Figures S1-S36. There was no evidence of inflation in the test statistics (Figure S37). LD score (LDSC) regression-derived SNP-based heritability was estimated to be 0.099 (standard error=0.01) on the liability scale. Characterization of the genome-wide results of META-2 suggested that the key UL associated variants were mostly intronic (Figure 2A). Genes associated with UL risk (Figure 2B) were significantly enriched on pathways related to gonad development, positive regulation of growth, and cellular senescence (Figure 2C) and they were mostly expressed in uterus, cervix, and fallopian tube (Figure 2D). In two of the novel loci, we found evidence that altered gene expression mediates UL association (Figures S38-S40).

**Table 1.**
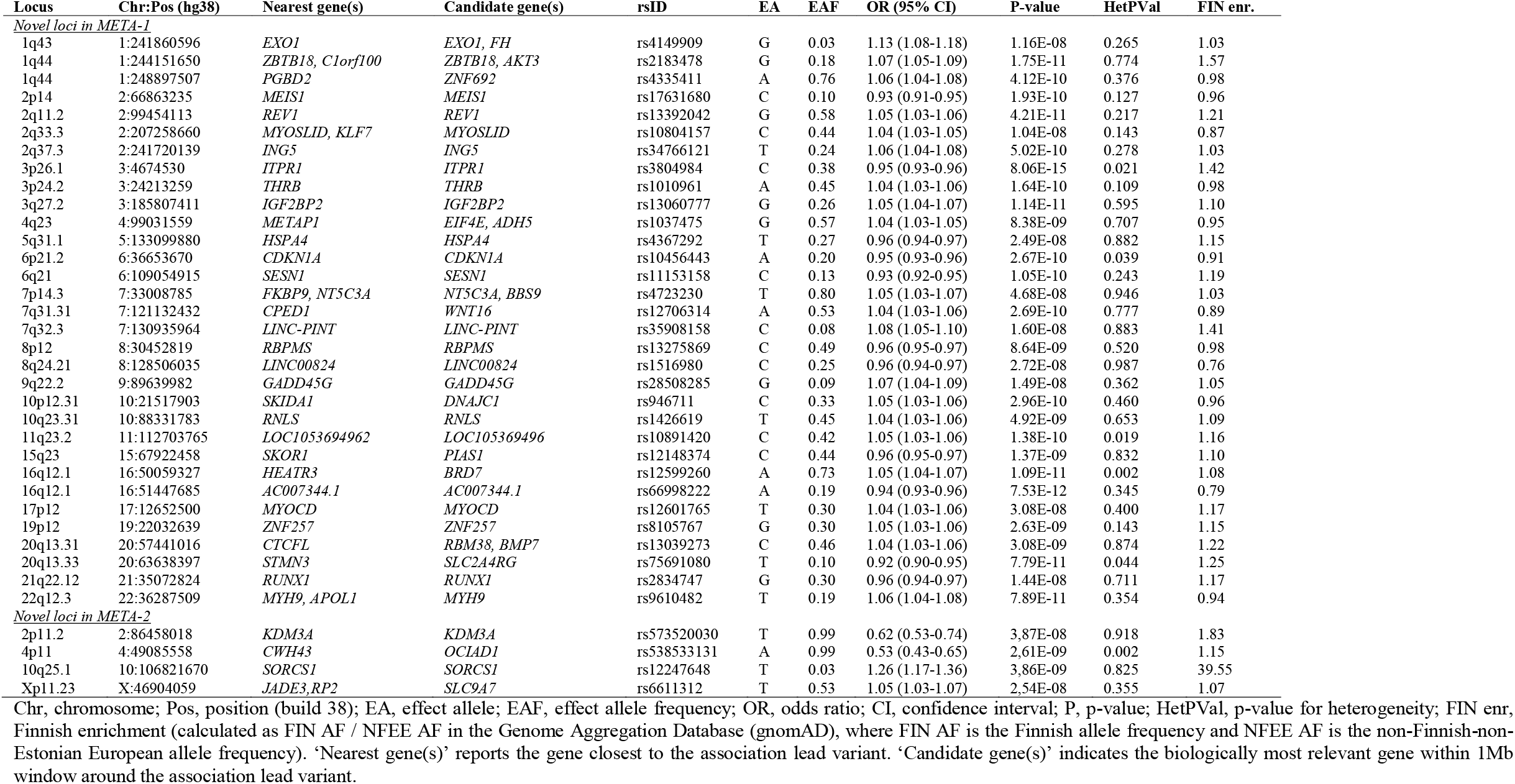
Novel UL risk loci. The table reports novel distinct loci (more than 1Mb apart) containing at least one variant identified to be associated with UL at *p*<5×10^−8^ in a meta-analysis of 53,534 UL cases and 373,024 female controls from FinnGen and a previously published meta-GWAS of UL^2^ (META-1) and in a meta-analysis of 33,244 UL cases and 311,271 female controls from FinnGen and the UK Biobank (META-2).

**Figure 1.**
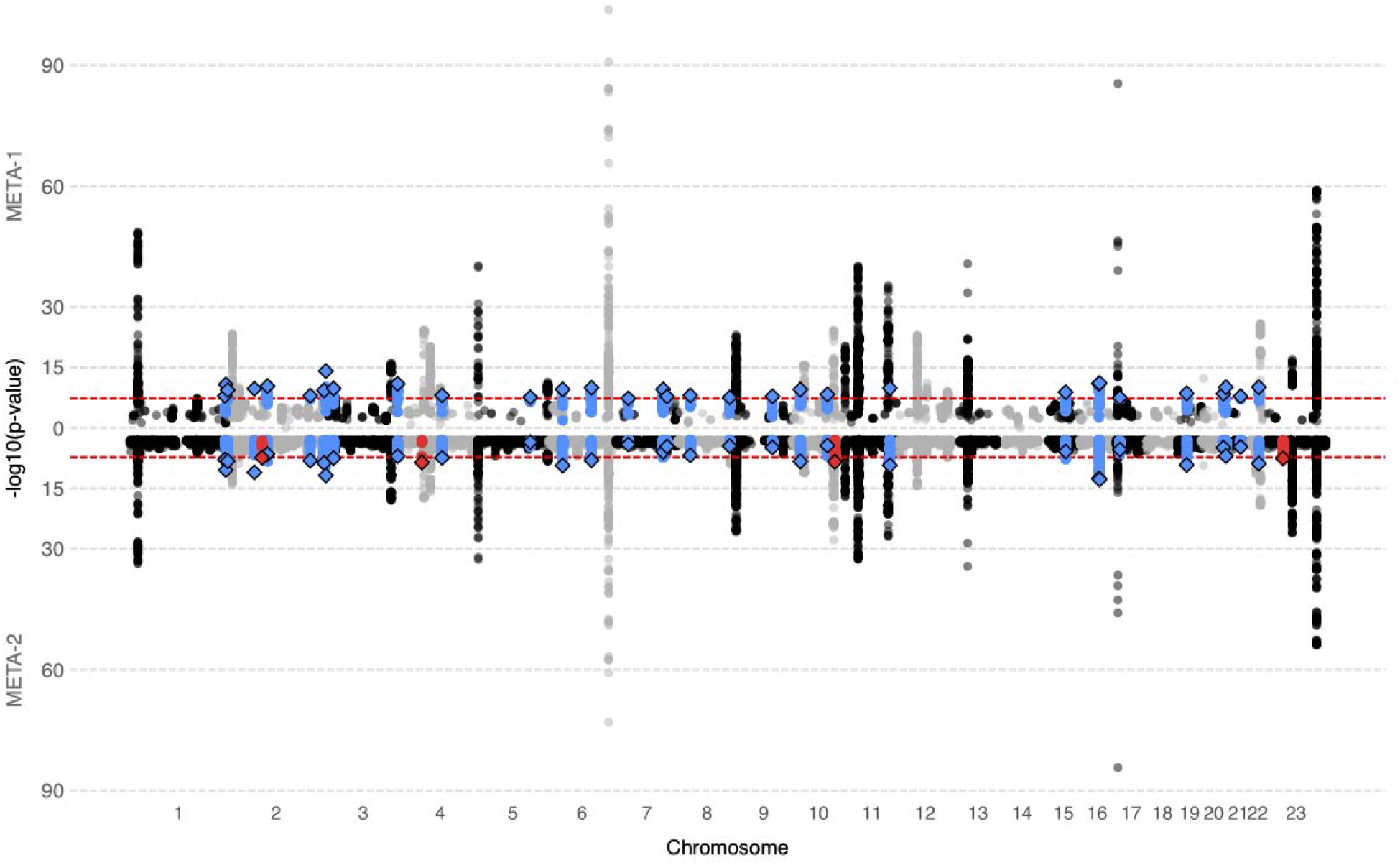
A combined Manhattan plot of UL associations in META-1 and META-2. We conducted UL GWAS in FinnGen and, subsequently, two sets of meta-analyses with data from a previously published UL meta-analysis^2^ and the UK Biobank, adding up to 53,534 UL cases and 373,024 female controls in META-1 (top) and 33,244 UL cases and 311,271 female controls in META-2 (bottom), respectively. Blue color denotes novel UL risk loci identified in META-1, and red color indicates novel loci identified in META-2 that were not associated previously with UL risk in prior GWASs or META-1. Black and grey colours indicate odd and even chromosome numbers, respectively. The red dashed lines correspond to the threshold for genome-wide significance (*p*<5×10^−8^).

**Figure 2.**
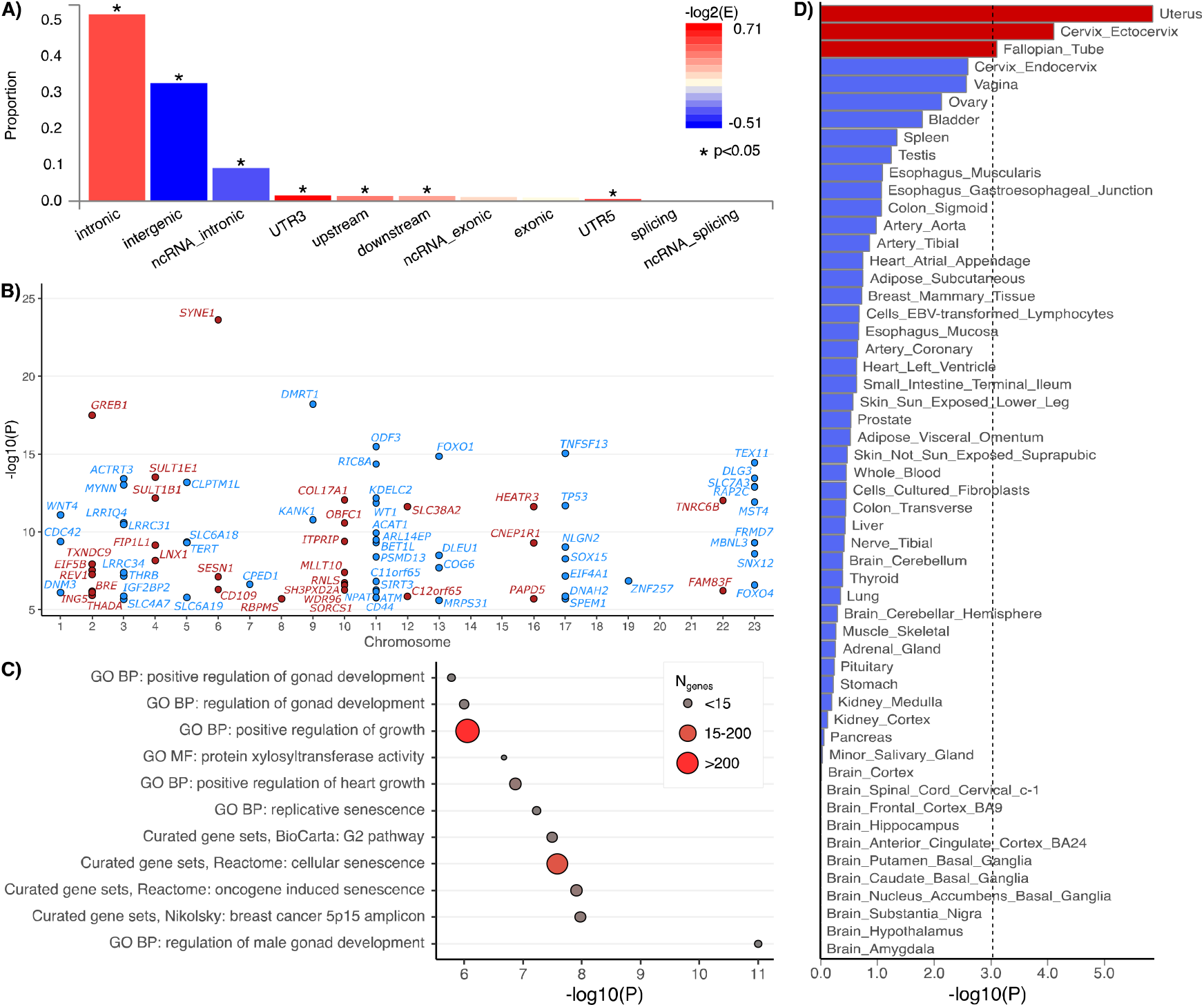
Variant summary and gene set-based results using summary statistics from META-2. A) The proportions of ‘independent genome-wide significant variants’ and ‘variants in LD with independent significant variants’ having corresponding functional annotation. Bars are colored according to -log_2_(enrichment) relative to all variants in the reference panel. B) A Manhattan plot of the gene-based test computed by MAGMA^54^. The input variants were mapped to 19,920 protein coding genes and, thus, significance was considered at *p*<2.51×10^−6^ (0.05/19,920); only 87 genes with p-values below this threshold are plotted. Blue and red colours indicate odd and even chromosome numbers, respectively. C) MAGMA^54^ gene-set enrichment analysis was performed for curated gene sets and GO terms available at MsigDB^55^. The plot shows the results for significantly enriched pathways (*p*_Bonferroni_<0.05). D) Results of MAGMA^54^ tissue expression analysis testing a positive relationship between tissue-specific gene expression profiles and disease-gene associations. Tissue-specific gene-expression data were from GTEx (v8)^56^. All data plotted in Figure 2 A-D were produced using FUMA^57^.

Previous GWAS findings^2,12-17^ have indicated that genetic factors altering pathways involved in estrogen signalling, Wnt signalling, transforming growth factor (TGF)-β signalling, and cell cycle progression are associated with UL risk^18^. The novel loci identified in this study further underscore the involvement of pathways regulating SMC proliferation in modulation of UL risk. Many of these pathways are interrelated: for example, both estrogen and progesterone increase the secretion of Wnt ligands from myometrial or leiomyoma SMC, which promotes cell proliferation and tumorigenesis via activation of β-catenin^19^. Steroid hormones also influence the production of ECM via signalling through the TGF-β family of ligands and receptors that are highly expressed in multiple fibrotic conditions^20-26^ and contribute to the fibrotic phenotype seen in UL^24,25,27-29^. We identified multiple loci with potential candidate genes functioning in one or more of these pathways, and, in the following, we describe some of our key findings with the focus on loci involved in regulation of SMC proliferation.

A central finding is the novel association at 17p12 harbouring myocardin (*MYOCD*; Figure S27). Myocardin is a transcription factor expressed in smooth muscle tissues, including most prominently arteries and colon, but also uterus (Figure S41), and it is required for SMC differentiation^30^. The expression of myocardin has been shown to be downregulated in UL tissue compared with normal myometrium^31^. Also, it has been proposed that the loss of myocardin function may be a key factor in driving SMC proliferation in UL^31^; however, there are no reported GWAS associations implicating myocardin until now. The lead variants near *MYOCD* are intergenic variants with no strong evidence of altered regulatory consequences (Table S3) and, thus, a possible association-driving mechanism remains inconclusive. We identified another myocardin-related novel UL risk association near ‘myocardin-induced smooth muscle cell lncRNA, inducer of differentiation’ (*MYOSLID*), a transcriptional target of myocardin^32^ (Figure S6). RegulomeDB^33^ annotation provided robust evidence that the association lead variant is a regulatory variant (Table S4) altering binding of multiple transcription factors (Table S5), including Fos proto-oncogene (FOS) that has been shown to be downregulated in UL^34^. To add yet another example of a myocardin-related UL risk loci, a well-established UL risk association at 22q13.1^2,12,16,35,36^ locates near ‘myocardin related transcription factor A’ (*MRTFA*; also known as *MKL1*), a gene interacting with myocardin^37^. Also, expression of *MRTFA/MKL1* has been shown to be downregulated in UL-related deletions at 22q^38^.

Others have suggested that loss of myocardin function may account for the differentiation defects of human leiomyosarcoma cells during malignant transformation^39^: downregulation of myocardin resulted in lower expression of cyclin dependent kinase inhibitor 1A (*CDKN1A*; also known as p21), a mediator of cell cycle G1 phase arrest, which facilitated cell cycle progression. We identified a novel UL association at 6p21.2 near *CDKN1A* (Figure S13); the association lead variants locate on intergenic region with possible regulatory consequences (Table S6). Previous evidence suggests that *CDKN1A* is among the genes, the expression of which correlates with UL size^40^. Another novel UL association at 20q13.31 harbours RNA binding motif protein 38 (*RBM38*; Figure S29) that specifically binds to and regulates the stability of *CDKN1A* transcripts^37^. In this locus, the UL risk-increasing rs13039273-C associates with lower *RBM38* expression in ovary (*p*=8.7×10^−6^)(Table S7, Figure S42; eQTL in uterus *p*=2.2×10^−3^). Relevant to UL, a variant in the same locus (rs760640162) has been previously associated with ‘pregnancy examination and test’ (*p*=5.97×10^−15^) in the UK Biobank^41,42^. Interestingly, estrogen receptor (ER)α has been shown to inhibit expression of myocardin^31^, suggesting that the ability of myocardin-CDKN1A-signalling to inhibit cell cycle progression may be impaired in tissues enriched with ERα. Taken together our findings and previous evidence, it seems highly probable that downregulation of myocardin-CDKN1A signalling increases the risk of UL.

We used an automated LDSC regression pipeline^43^ to evaluate the genetic correlations (r_g_) of UL with other traits (Figure 2; Table S8). The most robustly correlated traits were seen for other gynecological traits, such as bilateral oophorectomy (r_g_=0.74, *p*=1.05×10^−25^), hysterectomy (r_g_=0.62, *p*=3.24×10^−19^), age at menarche (r_g_=-0.18, *p*=1.40×10^−9^), excessive frequent and irregular menstruation (r_g_=0.51, *p*=5.66×10^−7^), having had menopause (r_g_=-0.33, *p*=2.31×10^−5^), and usage of hormonal replacement therapy (r_g_=0.20, p=5.54×10^−5^). Strong correlations were also seen between UL and anthropometric and metabolic measures, including whole body fat-free mass (r_g_=0.11, *p*=4.21×10^−5^), high blood pressure (r_g_=0.15, *p*=4.58×10^−6^), and serum triglyceride level (r_g_=0.16, *p*=4.80×10^−5^). Genetic susceptibility for UL was also correlated with genetic susceptibility for multiple psychiatric or mood-related traits, including depressive symptoms (r_g_=0.20, *p*=3.31×10^−5^) or neuroticism score (r_g_=0.14, *p*=3.88×10^−5^).

To estimate the causal relationships between UL and the key traits showing significant r_g_ with UL, we applied two-sample Mendelian randomization. Our results suggest that genetic tendency to accumulate fat-free mass may be causally linked to higher risk of UL (*p*=2.37×10^−3^; Figure 3; Table S9). In line with this, the higher impedance of whole body (i.e., a bioelectrical measure used for estimating body composition; lower muscle mass leads to higher impedance) was causally associated with a lower risk of UL (*p*=1.05×10^−3^). When considering the null causal effect of whole-body fat mass on UL risk (*p*=0.46), it seems apparent that the nominally significant (*p*=8.40×10^−3^) causal effect of BMI on UL in the present study and the previously reported associations of BMI with UL risk^44-46^ arise from the increased lean body mass rather than fat mass. Although we did not observe horizontal pleiotropy, the causal estimates were heterogenic for many key traits (Table S9), suggesting that all the underlying genetic pathways may not function in a consistent manner – thus, these results need to be interpreted with caution. However, in the leave-one-out sensitivity analyses all causal estimates were consistently positive (higher fat-free mass was causally associated with higher risk of UL) or negative (higher impedance was causally associated with lower risk of UL) suggesting that there is no single variant driving the causal associations (Figures S50-S63). Regarding most of the gynecological traits, the causal direction with UL was not clear (Table S9), suggesting that the shared molecular factors predispose to multiple gynecological complications simultaneously instead of one being causal to another. Of note, we replicated the previously reported finding of genetic predisposition to UL being causal to excessive menstrual bleeding^2^ (*p*=1.07×10^−4^).

**Figure 3.**
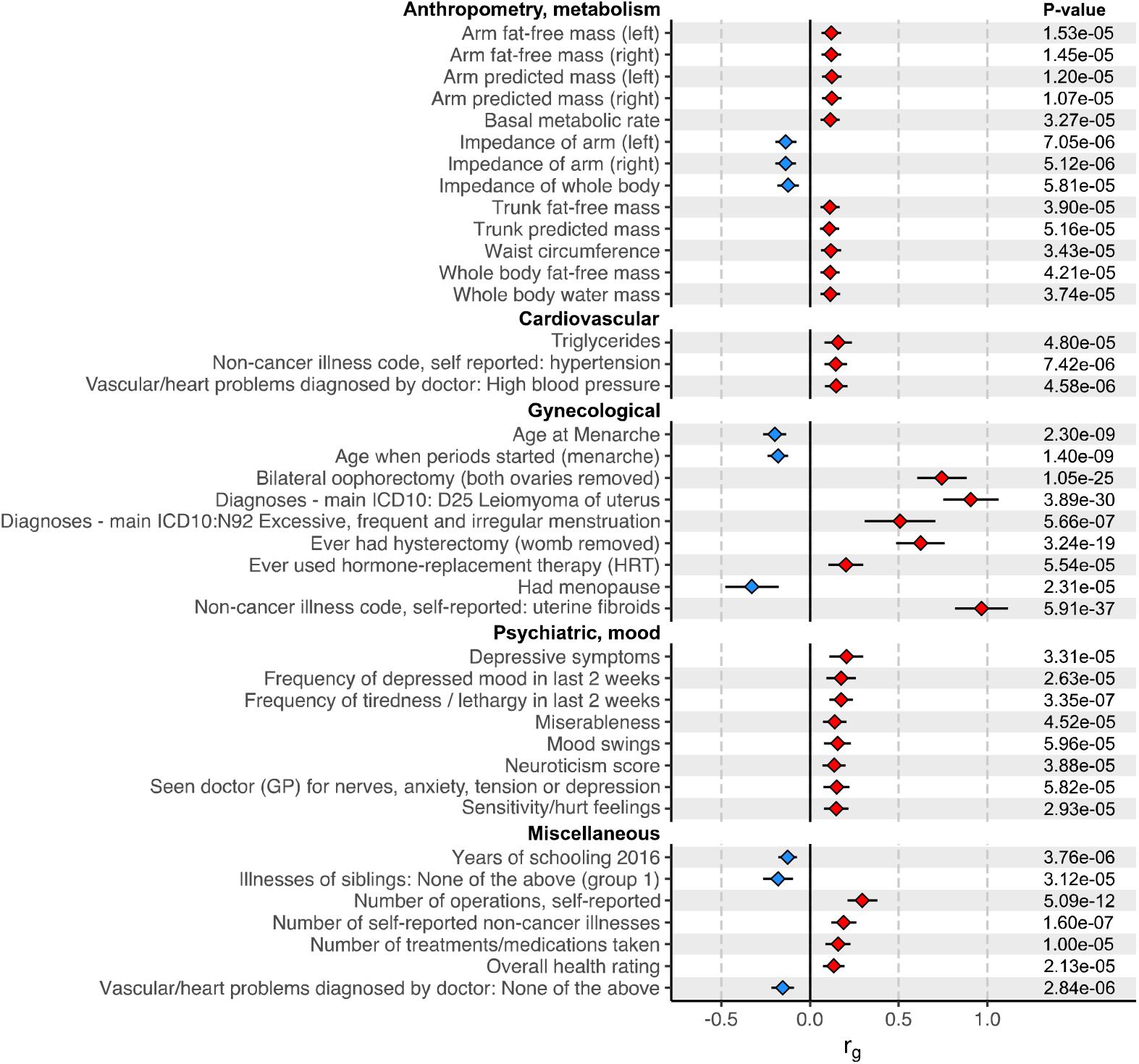
Genetic correlations (r_g_) of UL with other traits. The analyses were completed using an automated LD score regression pipeline available at http://ldsc.broadinstitute.org/^43^. Statistical significance was considered at *p*<6×10^−5^ (0.05/830, where 830 is the number of traits tested). The results of traits that showed significant r_g_ with UL are plotted (more results are provided in Table S8). Traits are named as reported in the LD Hub output.

**Figure 4.**
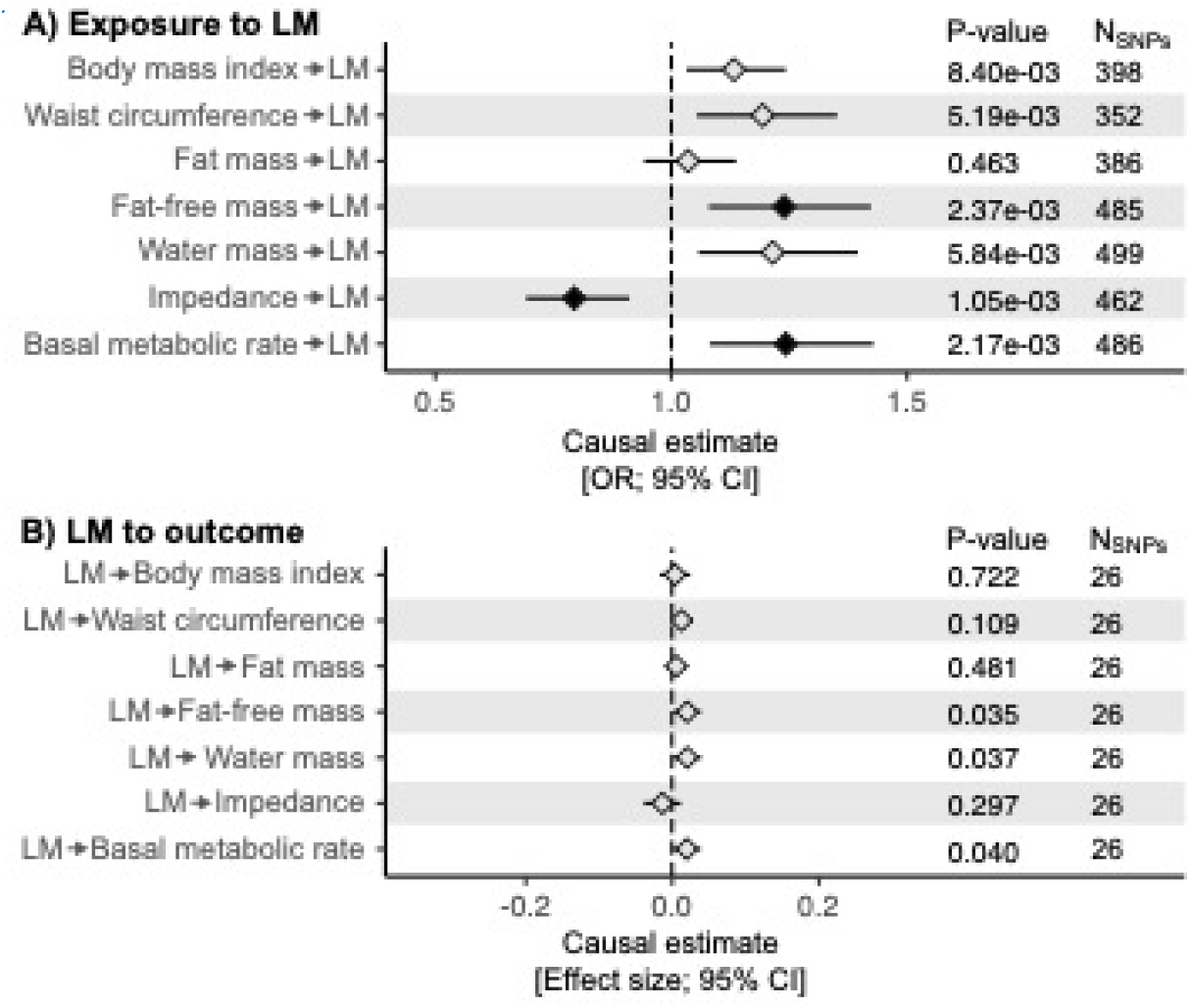
Causal relationships between UL and body composition measures. We estimated causal relationships between UL and the key traits showing significant genetic correlation with UL using two-sample Mendelian randomization implemented in ‘TwoSampleMR’ R library^58,59^. Bi-directional causal estimates were obtained by using summary statistics provided by the MRC Integrative Epidemiology Unit (IEU) GWAS database and UL GWAS results obtained in FinnGen in the present study to extract genetic instruments for other, mostly UKBB-based, traits and UL, respectively. LD pruning was completed using European population reference, threshold of r^2^=0.001, and clumping window of 10kb. Statistical significance was considered at *p*<3×10^−3^ (0.05/19, where 19 is the number of traits tested; black diamonds). The plot shows the causal estimates obtained using inverse variance weighted method for key body composition measures (more results are provided in Table S9; the scatter plots for the body composition measures can be found in Figures S43-S49, and the results of the leave-one-out sensitivity analyses are showin in Figures S50-S63).

The discovered causal relationship between genetic tendency to accumulate fat-free mass and UL risk provides a novel perspective on UL-related pathophysiology. UL are considered estrogen-dependent, and UL have higher ERα expression compared with normal uterine myometria^47,48^. ERs are expressed in a variety of tissues, including all musculoskeletal tissues^49^. In females, muscle mass and strength are closely coupled with estrogen status: girls begin to gain muscle mass after the onset of puberty^50^ whereas in older age during perimenopausal and postmenopausal periods, muscle strength declines considerably^51,52^. If estrogen enhances muscle growth^53^, the observed causal relationship between fat-free mass and UL risk could arise secondary to high estrogen contributing to the muscle growth. Many of the newly identified candidate genes, however, directly regulate cell proliferation – thus, it remains possible that the estrogen-rich environment, due to sexual maturity, may trigger excess SMC growth resulting in ULs in women who are genetically susceptible to build up muscle.

Currently, the only essentially curative treatment for UL is hysterectomy, which underscores the high demand for the development of alternative effective therapies. Our findings double the number of known UL risk loci and, thus, the herein presented results provide several potential targets for translational research to develop pharmacologic interventions for UL. Therapies targeted at myocardin-CDKN1A signalling or, considering the causal evidence, other factors regulating muscle growth may hold the greatest potential.

## Methods

### Study populations

**FinnGen** (www.finngen.fi/en) is a public-private partnership project launched in 2017 with an aim to improve human health through genetic research. The project utilizes genome information from a nationwide network of Finnish biobanks that are linked with digital health records from national hospital discharge (available from 1968), death (1969-), cancer (1953-), and medication reimbursement (1995-) registries using the unique national personal identification codes. Ultimately, the data resource will cover roughly 10% of the Finnish population. We studied data from 123,579 female participants (18,060 UL cases and 105,519 female controls) from FinnGen Preparatory Phase Data Freeze 5. UL cases were required to have an entry of ICD-10: D25, ICD-9: 218, or ICD-8: 21899, and participants who had no records of these entries were deemed as controls. The Coordinating Ethics Committee of the Hospital District of Helsinki and Uusimaa (HUS) approved the FinnGen study protocol Nr HUS/990/2017.

The FinnGen study is approved by Finnish Institute for Health and Welfare (THL), approval number THL/2031/6.02.00/2017, amendments THL/1101/5.05.00/2017, THL/341/6.02.00/2018, THL/2222/6.02.00/2018, THL/283/6.02.00/2019, THL/1721/5.05.00/2019, Digital and population data service agency VRK43431/2017-3, VRK/6909/2018-3, VRK/4415/2019-3 the Social Insurance Institution (KELA) KELA 58/522/2017, KELA 131/522/2018, KELA 70/522/2019, KELA 98/522/2019, and Statistics Finland TK-53-1041-17.

The Biobank Access Decisions for FinnGen samples and data utilized in FinnGen Data Freeze 5 include: THL Biobank BB2017_55, BB2017_111, BB2018_19, BB_2018_34, BB_2018_67, BB2018_71, BB2019_7, BB2019_8, BB2019_26, Finnish Red Cross Blood Service Biobank 7.12.2017, Helsinki Biobank HUS/359/2017, Auria Biobank AB17-5154, Biobank Borealis of Northern Finland_2017_1013, Biobank of Eastern Finland 1186/2018, Finnish Clinical Biobank Tampere MH0004, Central Finland Biobank 1-2017, and Terveystalo Biobank STB 2018001.

**FibroGENE** is consortium of conventional, population-based and direct-to-consumer cohorts that was assembled to replicate and identify UL risk variants. In the study by Gallagher *et al*., they studied data from 35,474 UL cases and 267,505 female controls including participants from four population-based cohorts (Women’s Genome Health Study, WGHS; Northern Finland Birth Cohort, NFBC; QIMR Berghofer Medical Research Institute, QIMR; the UK Biobank, UKBB) and one direct-to-consumer cohort (23andMe) Detailed descriptions of cohorts and sample selections are available in Supplementary Methods^2^.

**The UK Biobank** is a large national and international health resource following the health and wellbeing of 500,000 male and female volunteer participants, enrolled at ages from 40 to 69 ^60^. The study began in 2006 with the aim to follow the participants for at least 30 years thereafter. Information has been collected from participants during recruitment using questionnaires on socioeconomic status, lifestyle, family history and medical history. Participants have also been followed up for cause-specific morbidity and mortality through linkage to disease registries, death registries, and hospital admission records. For this study, altogether 220,936 women of European ancestry were considered. Based on both hospital-linked medical records and self-report (interview with research nurse), women with a history of UL were selected as cases (n=15,184), while controls (n=205,752) had no previous history of UL. Informed consent was obtained from all participants. The UKBB project is approved by the North West Multi-Centre Research Ethics Committee.

### Genotyping, imputation, and quality control

In FinnGen, genotyping of the samples was performed using Illumina and Affymetrix arrays (Illumina Inc., San Diego, and Thermo Fisher Scientific, Santa Clara, CA, USA). Sample quality control (QC) was performed to exclude individuals with high genotype missingness (>5%), ambiguous gender, excess heterozygosity (±4SD) and non-Finnish ancestry. Regarding variant QC, all variants with low Hardy-Weinberg equilibrium (HWE) p-value (<1e-6), high missingness (>2%) and minor allele count (MAC)<3 were excluded. Chip genotyped samples were pre-phased with Eagle 2.3.5 with the number of conditioning haplotypes set to 20,000. Genotype imputation was carried out by using the Finnish population-specific SISu v3 reference panel with Beagle 4.1 (version 08Jun17.d8b) as described in the following protocol: dx.doi.org/10.17504/protocols.io.nmndc5e. In post-imputation QC, variants with imputation INFO<0.6 were excluded.

In UK Biobank, genotyping of samples was performed either on the Affymetrix UK BiLEVE or Affymetrix UK Biobank Axiom array. The UK Biobank genotype data passed centralised quality control and was imputed by the UKBB team up to the HRC reference panel^61^.

### GWAS

The UL GWAS in FinnGen was completed using the Scalable and Accurate Implementation of Generalized (SAIGE) software^62^. The association models were adjusted for age, sex, the first 10 genetic principal components, and genotyping batch, and only variants with minimum allele count of 5 were included in the analysis.

Given the extensive relatedness in the UK Biobank data, a linear mixed model (LMM) was utilised for association testing that accounts for population structure and model the related individuals, including genotyping array, age and BMI as covariates as implemented in Bolt-LMM V2.3.

### Meta-analyses

Two sets of fixed-effect, inverse-variance weighted meta-analyses (implemented in METAL^63^) were performed: the results obtained in FinnGen were meta-analyzed with 1) the top 10,000 most significant variants associating with UL in a published GWAS^2^ (META-1) and 2) UL GWAS results obtained in the UKBB. Statistical significance was considered at the standard genome-wide significance level (*p*<5×10^−8^). Genomic inflation factor was estimated using an automated LD score (LDSC) regression pipeline^43^.

### Characterization of association signals

We used a web-based platform FUMA^57^ to perform functional annotations of the GWAS results: we completed functional gene mapping and gene-based association and enrichment tests using the genome-wide UL associations from META-2 (FinnGen and UKBB) and predefined lead variants as reported in Table S2. FUMA identifies variants showing genome-wide significant association (*p*<5×10^−8^) with the study trait and, among the significant variants, identifies variants in low LD (r^2^<0.6) as ‘independent significant variants’ and further identifies variants in LD (r^2^>0.6) with the ‘independent significant variants’; ANNOVAR^64^ annotations are performed for all these variants to obtain information on the functional consequences of the key variants. MAGMA^54^, also implemented in FUMA, was used to perform gene-based association testing and gene-set enrichment analyses: gene-based p-values were computed for protein-coding genes by mapping variants to genes and subsequent enrichment analyses were performed for the significant genes using 4728 curated gene sets and 6166 GO terms as reported in MsigDB^55^.

To further characterize the potential UL candidate gene(s) according to biological relevance, we annotated all genes within ±1Mb window from the association lead variant. We explored information provided by GenBank^37^ and UniProt^65^ to determine functions of the genes. To complement the information available in these databases, a broad literature search was performed to identify previous work published regarding to the genes of interest.

In each locus, we explored the associations of the lead variant with gene expression levels in the Genotype-Tissue Expression (GTEx) Portal; GTEx was accessed during 02/10/2021-02/11/2021. To further test if altered gene expression mediates UL risk associations, we used a method proposed by Zhu *et al*.^66^ as implemented in Complex Traits Genetics Virtual Lab (CTG-VL)^67^; we performed these tests using genome-wide UL results from META-2 and tissue-specific gene expression data (GTEx, V7) for uterus and whole blood. We further used RegulomeDB^33^ to discover regulatory elements overlapping with the intergenic variants in the novel loci showing genome-wide significant association with UL risk. Here, significant results for the novel loci are reported.

### SNP-based heritability and genetic correlations

The SNP-based heritability (h^2^_SNP_) of UL was estimated using LDSC regression implemented in LDSC software^68^. Population prevalence of 0.30 (as in ^2^) and sample prevalence of 0.11 were used to estimate h^2^_SNP_ on liability scale. We further applied LDSC to estimate genetic correlation (r_g_) of UL with other traits using LD Hub^43^, a web interface for performing automated LDSC regression. As recommended, we excluded the major histocompatilibity complex region from the analyses^43^. We tested r_g_ between UL and all 830 traits available in the LD Hub database and, thus, statistical significance was considered at *p*<6×10^−5^ (0.05/830).

### Mendelian randomization

To test for causal inferences between UL and the key traits showing significant r_g_ with UL, we performed bi-directional two-sample Mendelian randomization. These analyses were completed using ‘TwoSampleMR’ R library^58^ (https://mrcieu.github.io/TwoSampleMR/). To avoid possible bias from overlapping samples, we extracted genetic instruments for UL from the GWAS results obtained in FinnGen, and for other traits from the GWAS database provided by the MRC Integrative Epidemiology Unit (IEU) available at https://gwas.mrcieu.ac.uk/ and integrated in TwoSampleMR. LD pruning was completed using European population reference, threshold of r^2^=0.001, and clumping window of 10kb, as set as default in ‘clump_data’ function; the numbers of SNPs available for the analyses are listed in Table S9. Inverse variance weighted (IVW) method was considered as a primary analysis, and MR Egger estimates were derived in sensitivity analyses: the estimates were required to be to a matching direction with the IVW-estimates and Egger intercepts were evaluated to assess horizontal pleiotropy. Cochran’s Q statistics were derived using ‘mr_heterogeneity’ function to test for heterogeneity. To screen for highly influential variants that could drive the association for example due to horizontal pleiotropy, we performed leave-one-out analyses using ‘mr_leaveoneout’ function. Statistical significance for the causal effects was considered at *p*<3×10^−3^ (0.05/19) to correct for 19 traits tested.

## Supporting information

Supplementary Checklist

## Data Availability

Summary statistics will be made available through the NHGRI-EBI GWAS Catalog with GCSTxxxxxxx upon publication.

## Acknowledgements and funding

The work was supported through The Sigrid Juselius Foundation (JK) and funds from the Academy of Finland [grant numbers 297338 and 307247](JK), Novo Nordisk Foundation [grant number NNF17OC0026062] (JK), and The Finnish Medical Association (OU).

The FinnGen project is funded by two grants from Business Finland (HUS 4685/31/2016 and UH 4386/31/2016) and eleven industry partners (AbbVie Inc, AstraZeneca UK Ltd, Biogen MA Inc, Celgene Corporation, Celgene International II Sàrl, Genentech Inc, Merck Sharp & Dohme Corp, Pfizer Inc., GlaxoSmithKline, Sanofi, Maze Therapeutics Inc., Janssen Biotech Inc). Following biobanks are acknowledged for delivering biobank samples to Finngen: Auria Biobank (www.auria.fi/biopankki), THL Biobank (www.thl.fi/biobank), Helsinki Biobank (www.helsinginbiopankki.fi), Biobank Borealis of Northern Finland (https://www.ppshp.fi/Tutkimus-ja-opetus/Biopankki/Pages/Biobank-Borealis-briefly-in-English.aspx), Finnish Clinical Biobank Tampere (www.tays.fi/en-US/Research_and_development/Finnish_Clinical_Biobank_Tampere), Biobank of Eastern Finland (www.ita-suomenbiopankki.fi/en), Central Finland Biobank (www.ksshp.fi/fi-FI/Potilaalle/Biopankki), Finnish Red Cross Blood Service Biobank (www.veripalvelu.fi/verenluovutus/biopankkitoiminta) and Terveystalo Biobank (www.terveystalo.com/fi/Yritystietoa/Terveystalo-Biopankki/Biopankki/). All Finnish Biobanks are members of BBMRI.fi infrastructure (www.bbmri.fi).

This research has been conducted using data from UK Biobank, a major biomedical database, (http://www.ukbiobank.ac.uk/) under project ID 9637 (Gallagher et al., 2019).

The Genotype-Tissue Expression (GTEx) Project was supported by the Common Fund of the Office of the Director of the National Institutes of Health, and by NCI, NHGRI, NHLBI, NIDA, NIMH, and NINDS. The data used for the analyses described in this manuscript were obtained from the GTEx Portal on 02/10/2021-02/11/2021.

## Author contributions

OU conceptualized the study. ES, NR, KTZ, CMB, and JK contributed to the analysis plan. ES, JT and NR analyzed data and generated results. ES, JT, NR, KTZ, and OU interpreted the results. ES and OU wrote the original manuscript. JK supervised the study, obtained funding and provided additional study resources. All authors contributed to revising the content of the manuscript and approved the final version.

## Competing interests

KTZ: Competing financial interests: Scientific collaborations (grant funding) with Bayer AG, Roche Diagnostics Inc, MDNA Life Sciences, and Evotec. Competing non-financial interests: Board memberships of the World Endometriosis Society, World Endometriosis Research Foundation, and research advisory committee member of Wellbeing of Women UK.

CMB: Competing financial interests: Scientific collaborations (grant funding) with Bayer AG, Roche Diagnostics Inc, MDNA Life Sciences, and Evotec. Scientific board Myovant; IDDM Member ObsEva. Competing non-financial interests: Chair ESHRE Endometriosis Guideline Development Group.

## Contributors of FinnGen

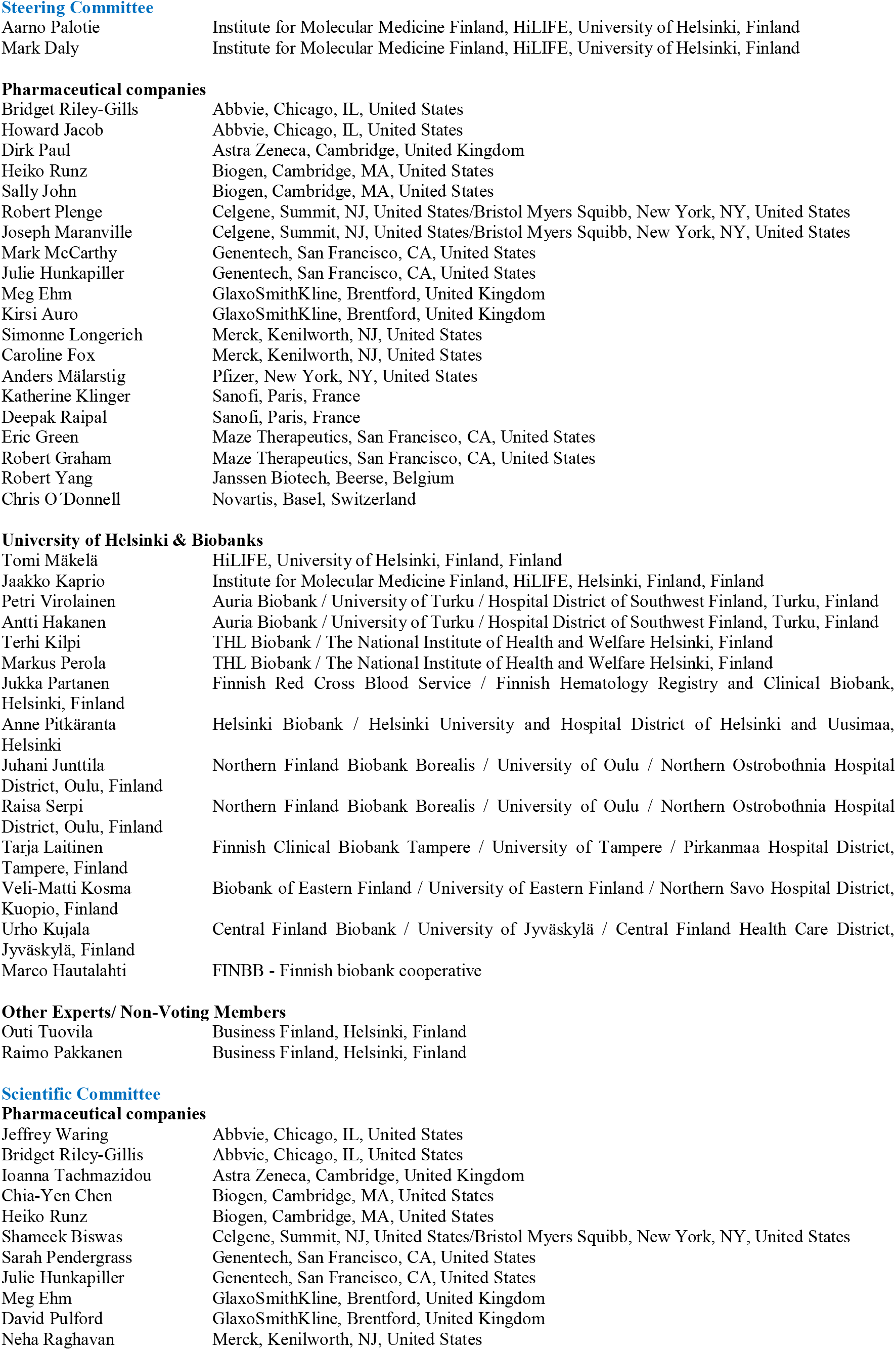
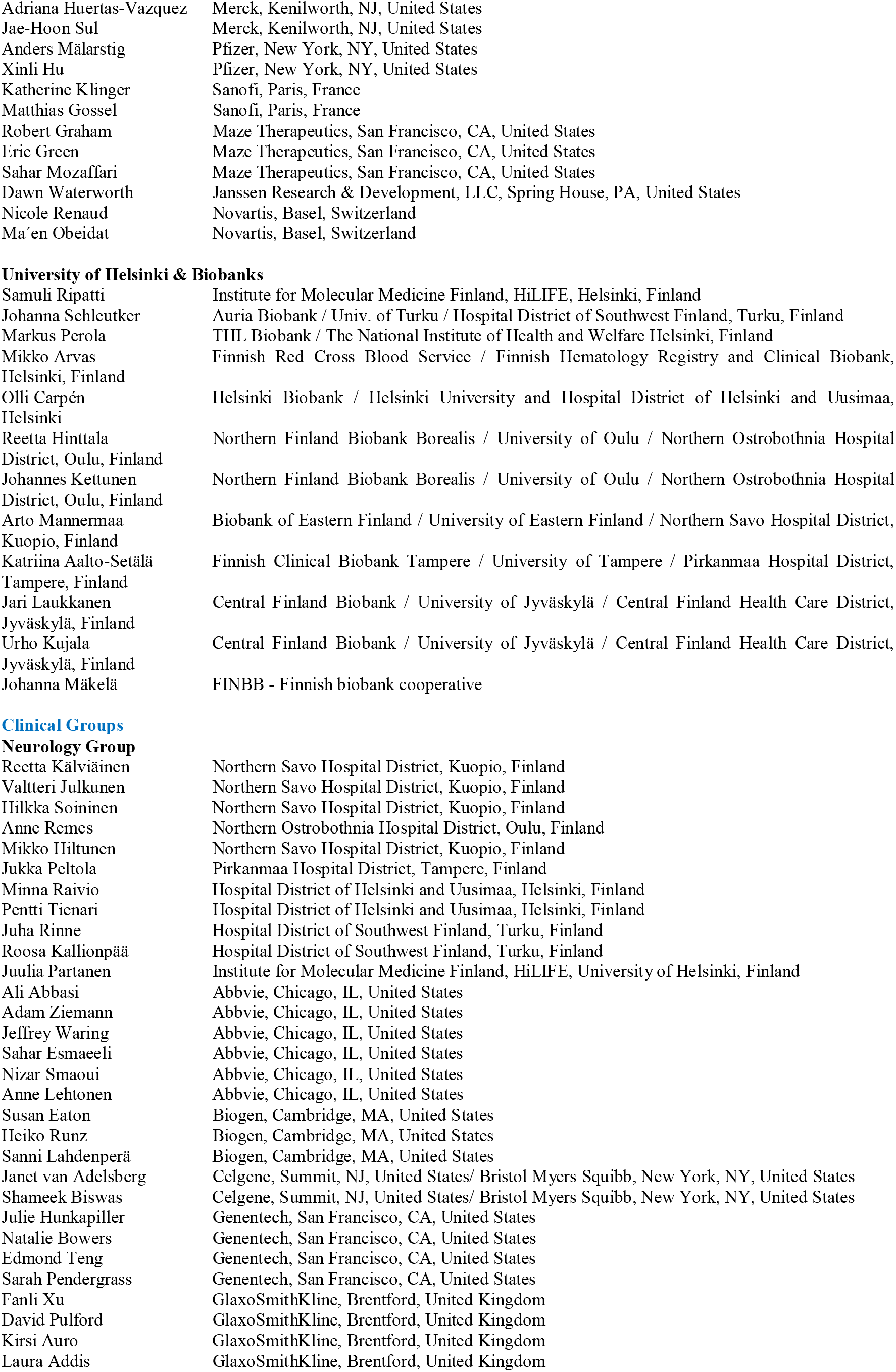
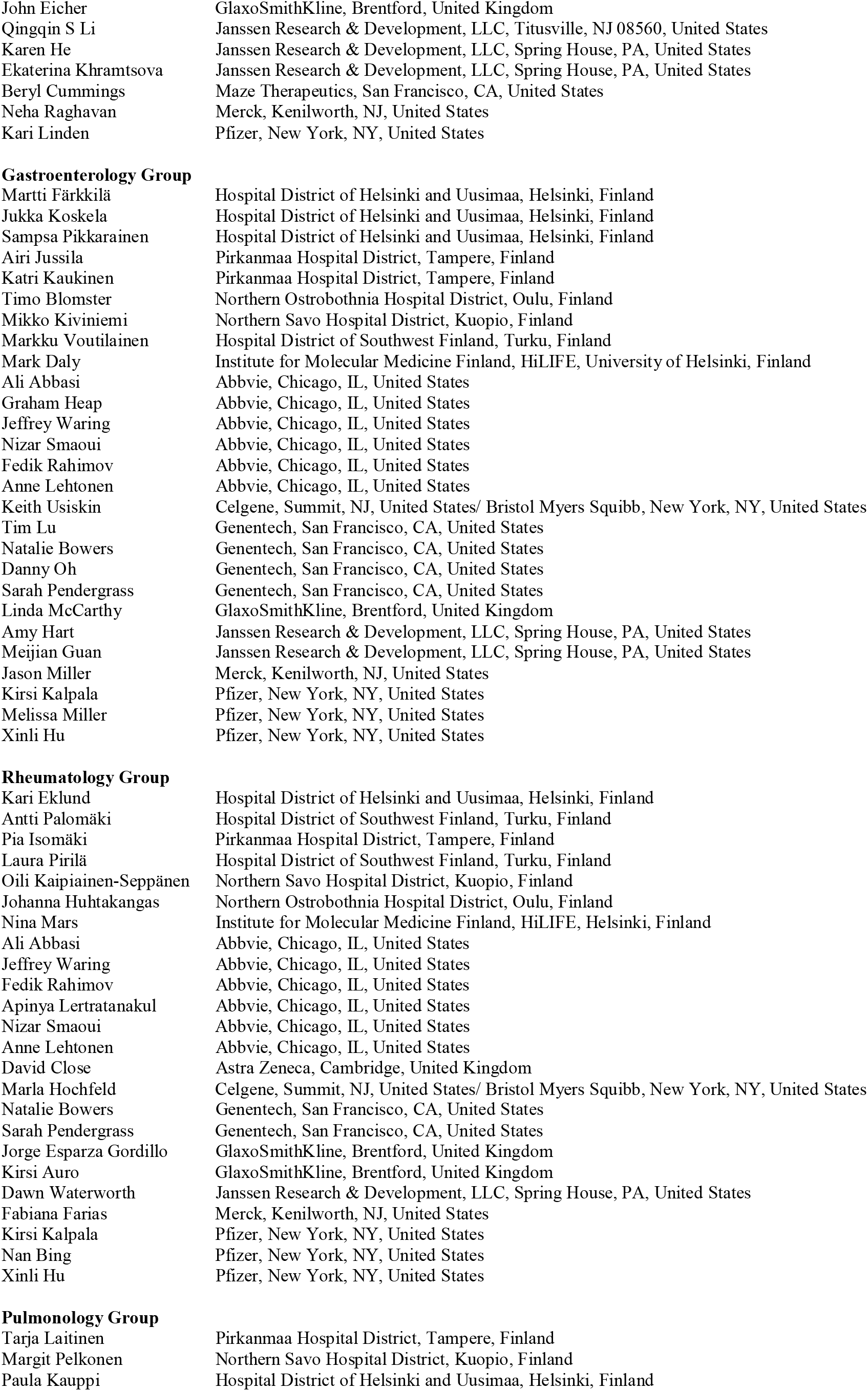
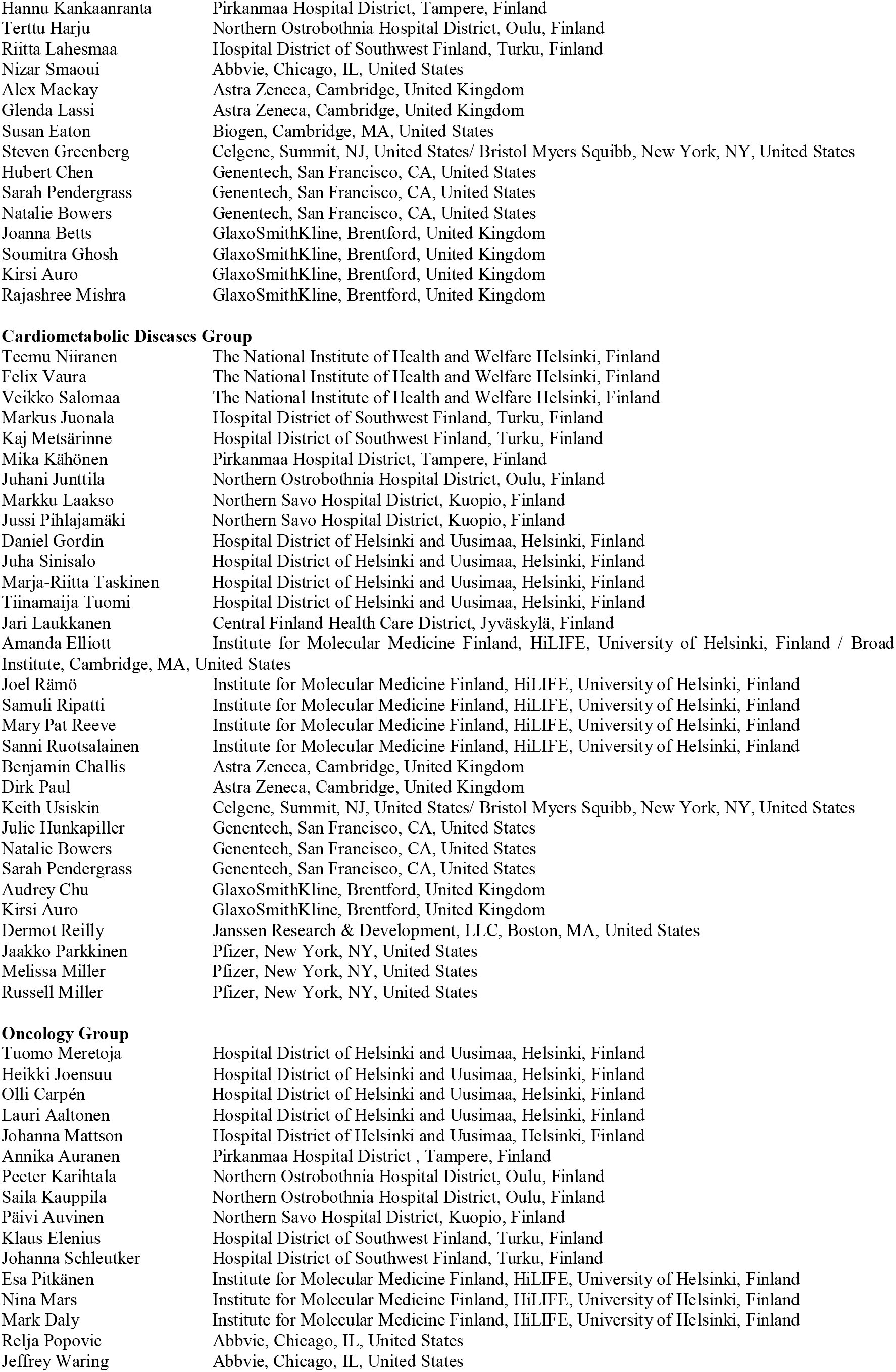
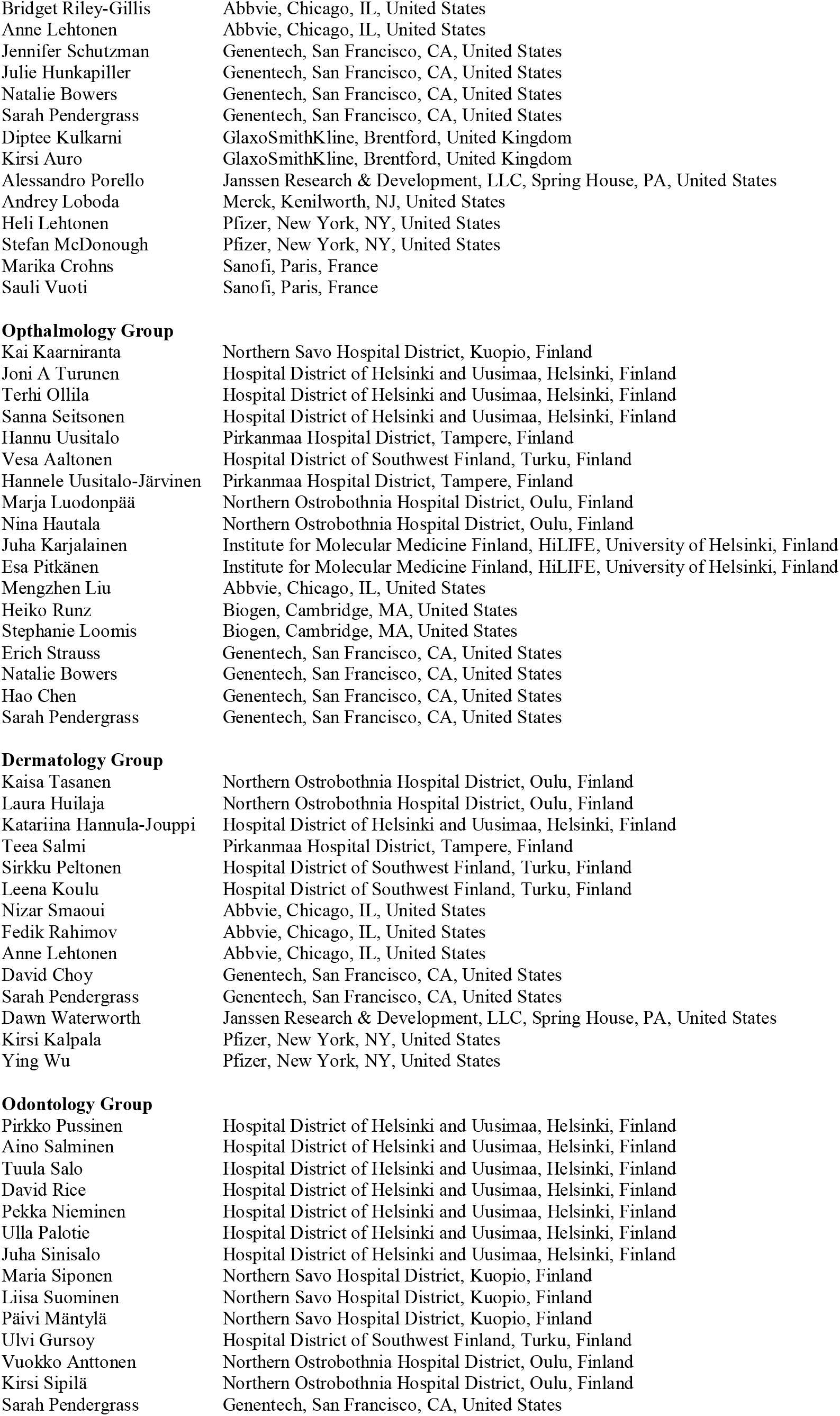
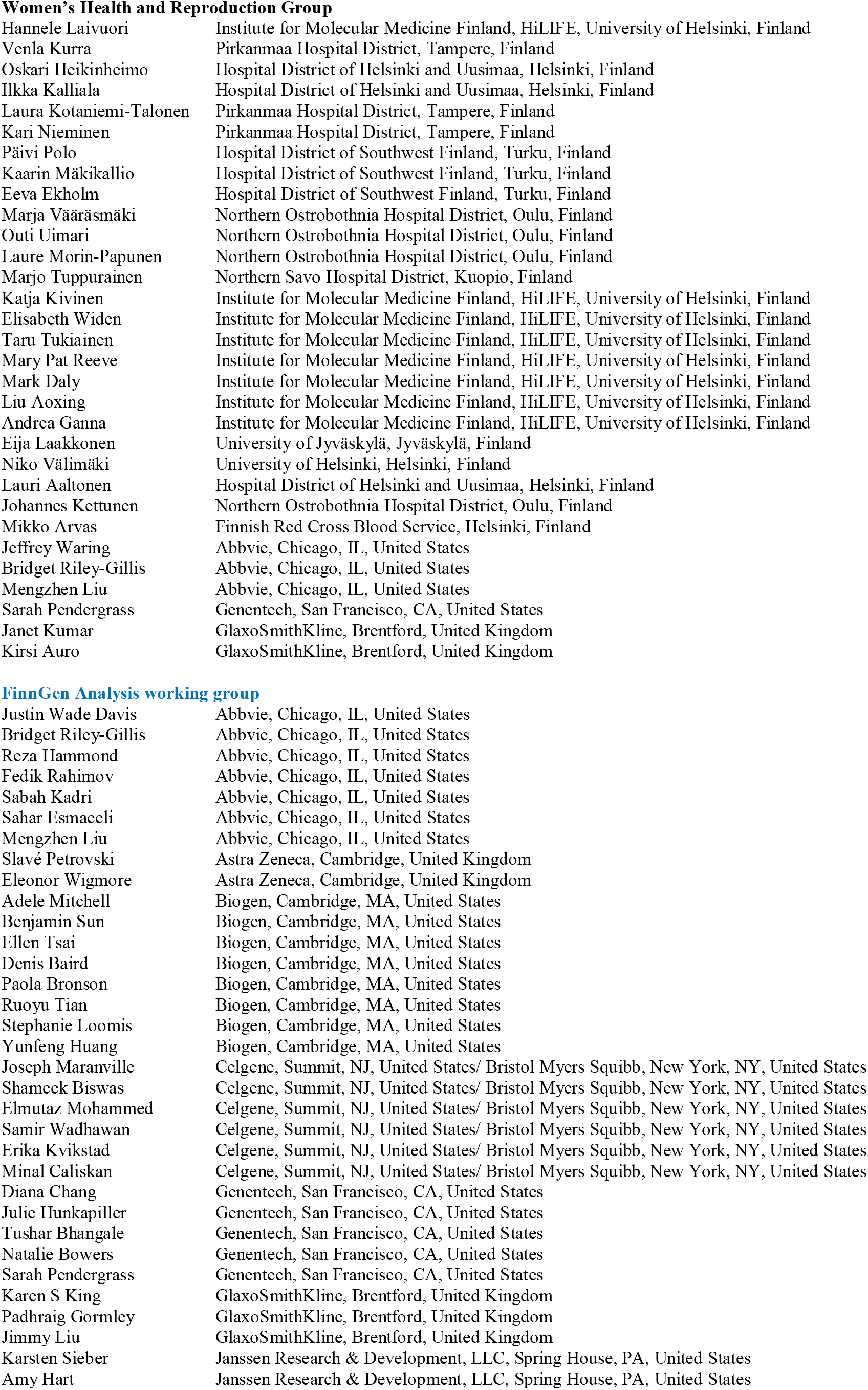
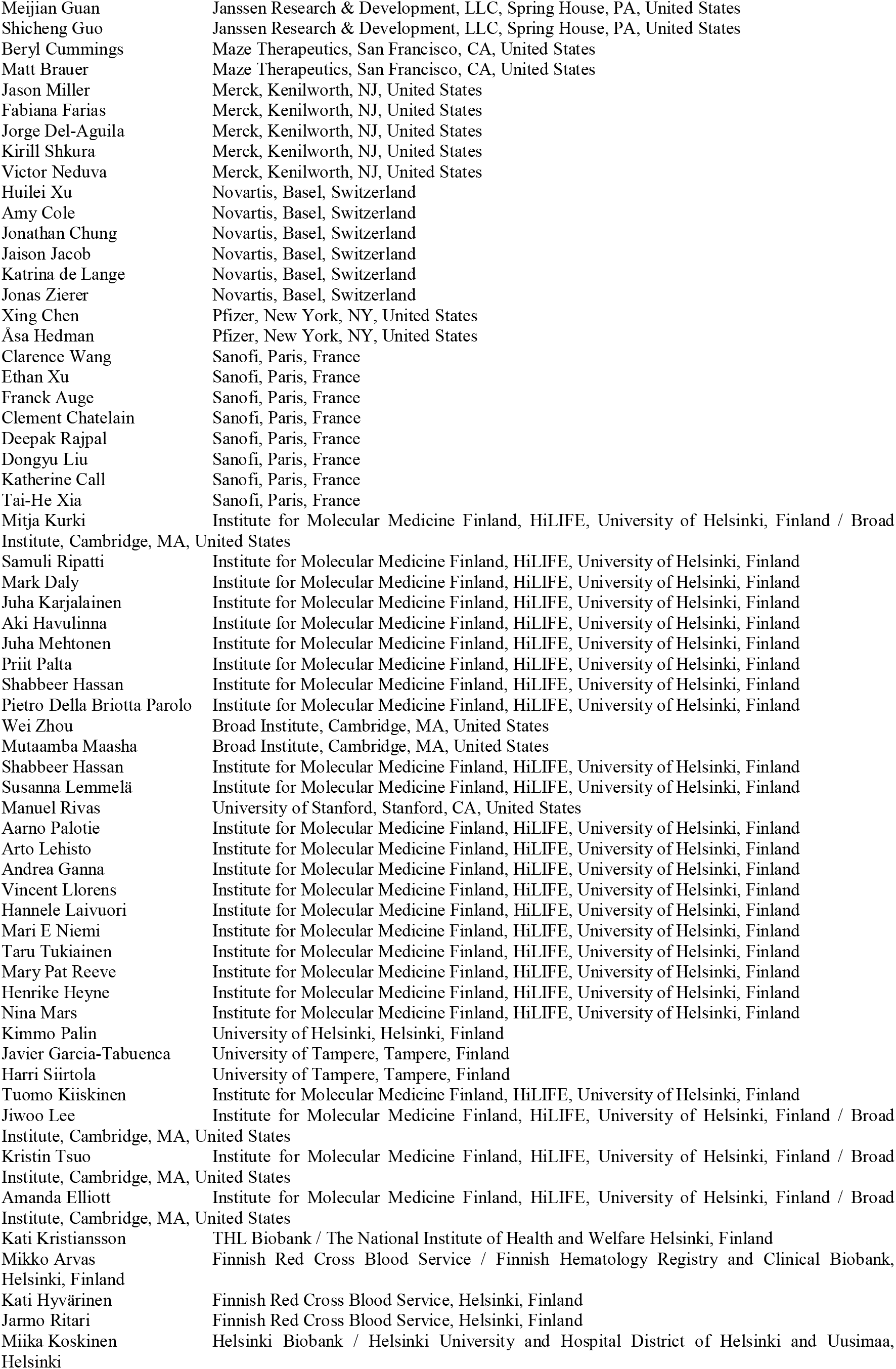
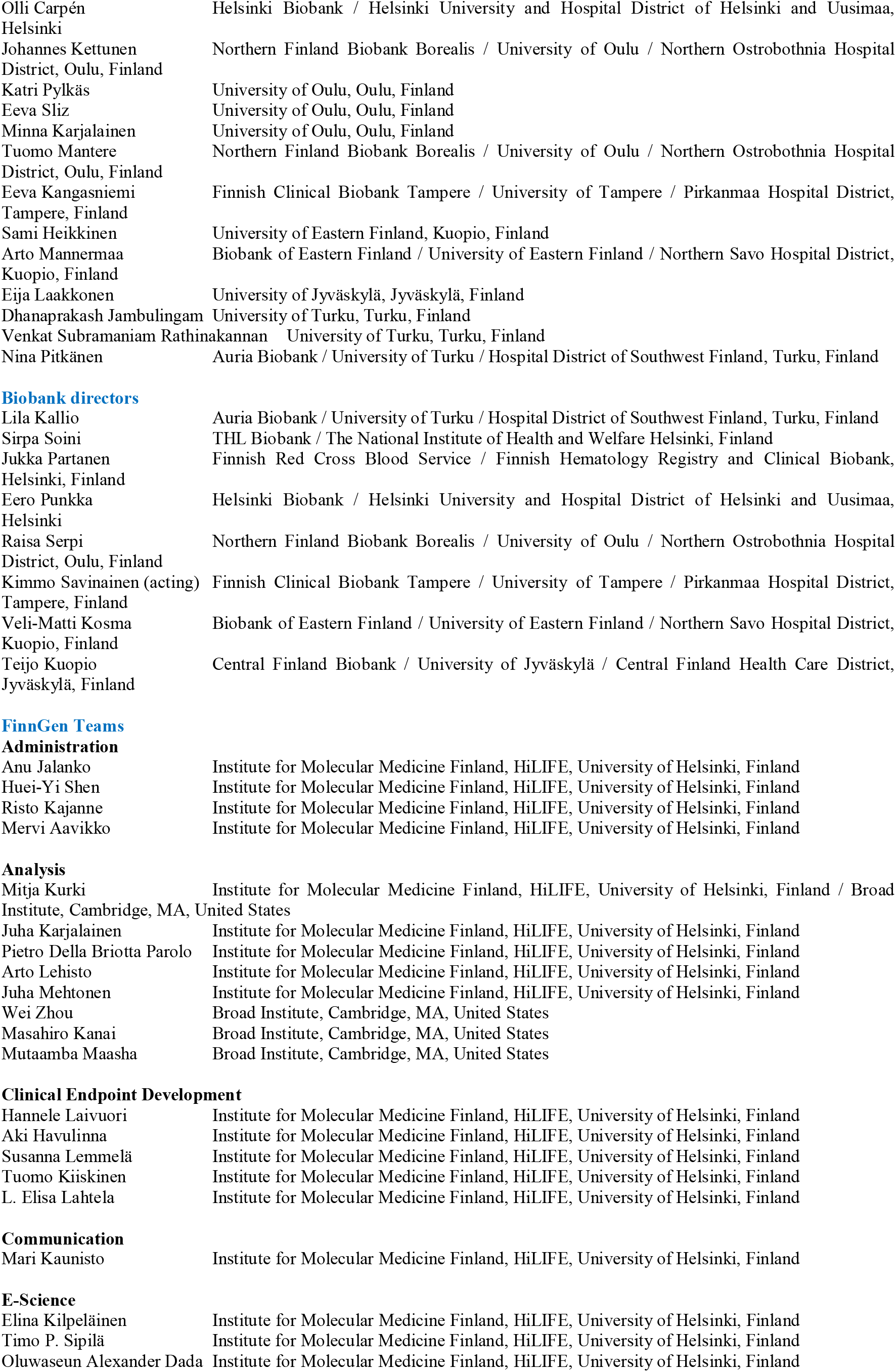
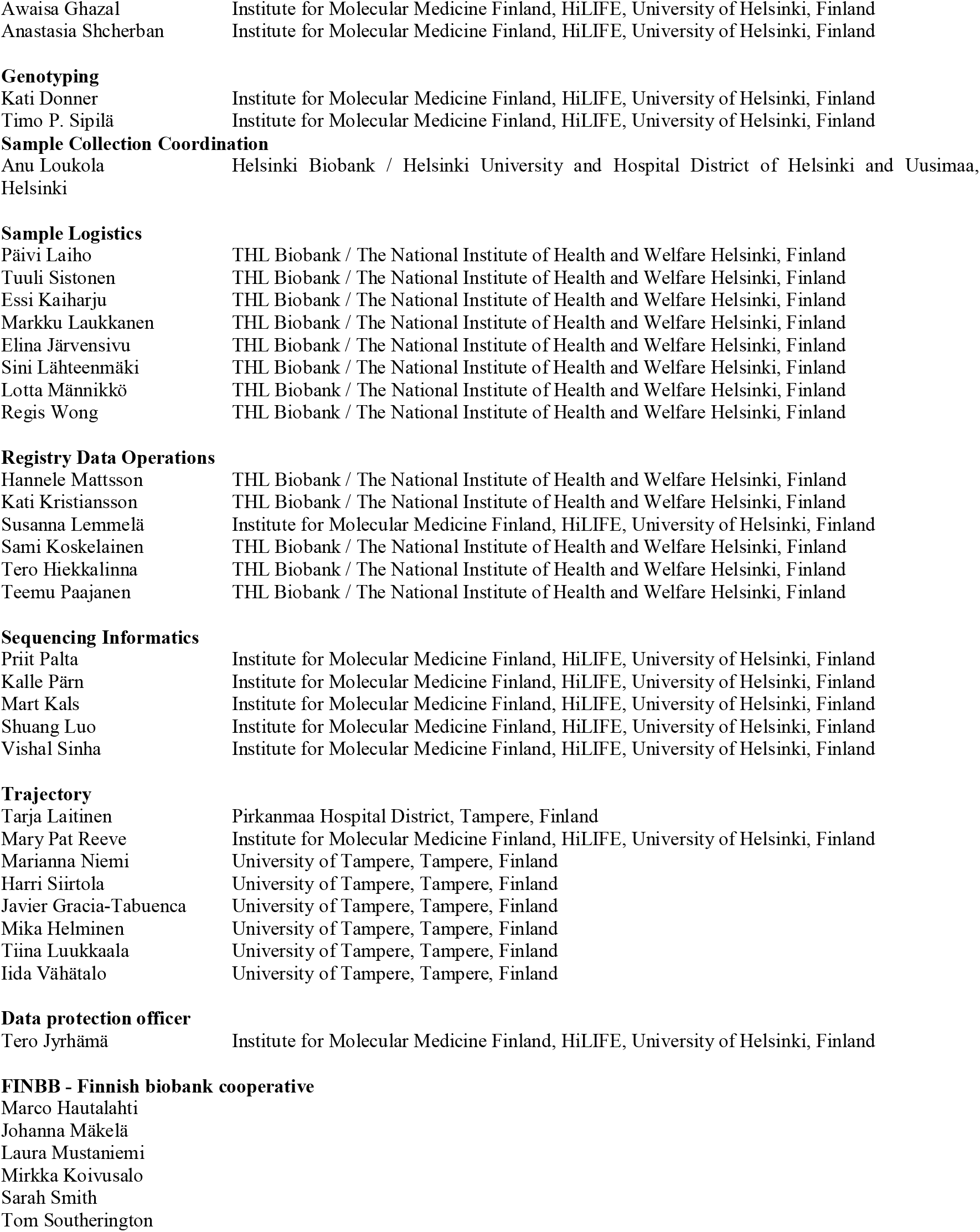

## Notes

### Author Declarations

The FinnGen study is approved by Finnish Institute for Health and Welfare (THL), approval number THL/2031/6.02.00/2017, amendments THL/1101/5.05.00/2017, THL/341/6.02.00/2018, THL/2222/6.02.00/2018, THL/283/6.02.00/2019, THL/1721/5.05.00/2019, Digital and population data service agency VRK43431/2017-3, VRK/6909/2018-3, VRK/4415/2019-3 the Social Insurance Institution (KELA) KELA 58/522/2017, KELA 131/522/2018, KELA 70/522/2019, KELA 98/522/2019, and Statistics Finland TK-53-1041-17. The Biobank Access Decisions for FinnGen samples and data utilized in FinnGen Data Freeze 5 include: THL Biobank BB2017_55, BB2017_111, BB2018_19, BB_2018_34, BB_2018_67, BB2018_71, BB2019_7, BB2019_8, BB2019_26, Finnish Red Cross Blood Service Biobank 7.12.2017, Helsinki Biobank HUS/359/2017, Auria Biobank AB17-5154, Biobank Borealis of Northern Finland_2017_1013, Biobank of Eastern Finland 1186/2018, Finnish Clinical Biobank Tampere MH0004, Central Finland Biobank 1-2017, and Terveystalo Biobank STB 2018001.

