## Supplemental Figures and Tables for "Evidence of a causal relationship between genetic tendency to gain muscle mass and uterine leiomyomata"

### Table of Contents

**FIGURE S1.** REGIONAL ASSOCIATION PLOT OF THE NOVEL UL ASSOCIATION IN CHR1 NEAR *EXO1*. 2
**FIGURE S2.** REGIONAL ASSOCIATION PLOT OF THE NOVEL UL ASSOCIATION IN CHR1 NEAR *ZBTB18*. 3
**FIGURE S3.** REGIONAL ASSOCIATION PLOT OF THE NOVEL UL ASSOCIATION IN CHR1 NEAR *PGBD2 (ZNF692)*. 4
**FIGURE S4.** REGIONAL ASSOCIATION PLOT OF THE NOVEL UL ASSOCIATION IN CHR2 NEAR *MEIS1*. 5
**FIGURE S5.** REGIONAL ASSOCIATION PLOT OF THE NOVEL UL ASSOCIATION IN CHR2 NEAR *REV1*. 6
**FIGURE S6.** REGIONAL ASSOCIATION PLOT OF THE NOVEL UL ASSOCIATION IN CHR2 NEAR *MYOSLID*. 7
**FIGURE S7.** REGIONAL ASSOCIATION PLOT OF THE NOVEL UL ASSOCIATION IN CHR2 NEAR *ING5*. 8
**FIGURE S8.** REGIONAL ASSOCIATION PLOT OF THE NOVEL UL ASSOCIATION IN CHR3 NEAR *ITPR1*. 9
**FIGURE S9.** REGIONAL ASSOCIATION PLOT OF THE NOVEL UL ASSOCIATION IN CHR3 NEAR *THRB*. 10
**FIGURE S10.** REGIONAL ASSOCIATION PLOT OF THE NOVEL UL ASSOCIATION IN CHR3 NEAR *IGF2BP2*. 11
**FIGURE S11.** REGIONAL ASSOCIATION PLOT OF THE NOVEL UL ASSOCIATION IN CHR4 NEAR *METAP1 (EIF4E, ADH5)*. 12
**FIGURE S12.** REGIONAL ASSOCIATION PLOT OF THE NOVEL UL ASSOCIATION IN CHR5 NEAR *HSPA4*. 13
**FIGURE S13.** REGIONAL ASSOCIATION PLOT OF THE NOVEL UL ASSOCIATION IN CHR6 NEAR *CDKN1A*. 14
**FIGURE S14.** REGIONAL ASSOCIATION PLOT OF THE NOVEL UL ASSOCIATION IN CHR6 NEAR *SESN1*. 15
**FIGURE S15.** REGIONAL ASSOCIATION PLOT OF THE NOVEL UL ASSOCIATION IN CHR7 NEAR *NT5C3A*. 16
**FIGURE S16.** REGIONAL ASSOCIATION PLOT OF THE NOVEL UL ASSOCIATION IN CHR7 NEAR *CPED1 (WNT16)*. 17
**FIGURE S17.** REGIONAL ASSOCIATION PLOT OF THE NOVEL UL ASSOCIATION IN CHR7 NEAR *LINC-PINT*. 18
**FIGURE S18.** REGIONAL ASSOCIATION PLOT OF THE NOVEL UL ASSOCIATION IN CHR8 NEAR *RBPMS*. 19
**FIGURE S19.** REGIONAL ASSOCIATION PLOT OF THE NOVEL UL ASSOCIATION IN CHR8 NEAR *LINC00824*. 20
**FIGURE S20.** REGIONAL ASSOCIATION PLOT OF THE NOVEL UL ASSOCIATION IN CHR9 NEAR *GADD45G*. 21
**FIGURE S21.** REGIONAL ASSOCIATION PLOT OF THE NOVEL UL ASSOCIATION IN CHR10 NEAR *SKIDA1 (DNAJC1)*. 22
**FIGURE S22.** REGIONAL ASSOCIATION PLOT OF THE NOVEL UL ASSOCIATION IN CHR10 NEAR *RNLS*. 23
**FIGURE S23.** REGIONAL ASSOCIATION PLOT OF THE NOVEL UL ASSOCIATION CHR11 NEAR *NCAM1*. 24
**FIGURE S24.** REGIONAL ASSOCIATION PLOT OF THE NOVEL UL ASSOCIATION IN CHR15 NEAR *SKOR1 (PIAS1)*. 25
**FIGURE S25.** REGIONAL ASSOCIATION PLOT OF THE NOVEL UL ASSOCIATION IN CHR16 NEAR *HEATR3*. 26
**FIGURE S26.** REGIONAL ASSOCIATION PLOT OF THE NOVEL UL ASSOCIATION IN CHR16 NEAR *AC007344.1*. 27
**FIGURE S27.** REGIONAL ASSOCIATION PLOT OF THE NOVEL UL ASSOCIATION IN CHR17 NEAR *MYOCD*. 28
**FIGURE S28.** REGIONAL ASSOCIATION PLOT OF THE NOVEL UL ASSOCIATION IN CHR19 NEAR *ZNF257*. 29
**FIGURE S29.** REGIONAL ASSOCIATION PLOT OF THE NOVEL UL ASSOCIATION IN CHR20 NEAR *CTCFL*. 30
**FIGURE S30.** REGIONAL ASSOCIATION PLOT OF THE NOVEL UL ASSOCIATION IN CHR20 NEAR *STMN3*. 31
**FIGURE S31.** REGIONAL ASSOCIATION PLOT OF THE NOVEL UL ASSOCIATION IN CHR21 NEAR *RUNX1*. 32
**FIGURE S32.** REGIONAL ASSOCIATION PLOT OF THE NOVEL UL ASSOCIATION IN CHR22 NEAR *MYH9*. 33
**FIGURE S33.** REGIONAL ASSOCIATION PLOT OF THE NOVEL UL ASSOCIATION IN CHR2 NEAR *KDM3A* [META-2]. 34
**FIGURE S34.** REGIONAL ASSOCIATION PLOT OF THE NOVEL UL ASSOCIATION IN CHR4 NEAR *CWH43* [META-2]. 35
**FIGURE S35.** REGIONAL ASSOCIATION PLOT OF THE NOVEL UL ASSOCIATION IN CHR10 NEAR *SORCS1* [META-2]. 36
**FIGURE S36.** REGIONAL ASSOCIATION PLOT OF THE NOVEL UL ASSOCIATION IN CHRX NEAR *JADE3* [META-2]. 37
**FIGURE S37.** QUANTILE-QUANTILE PLOTS OF THE P-VALUES IN META-1 (UPPER) AND META-2 (LOWER). 38
**FIGURE S38.** SMR RESULT: *HEATR3* AT 16Q12.1 IN UTERUS. 39
**FIGURE S39.** SMR RESULT: *HEATR3* AT 16Q12.1 IN WHOLE BLOOD. 40
**FIGURE S40.** SMR RESULT: *ZNF257* AT 19Q12 IN WHOLE BLOOD. 41
**FIGURE S41.** *MYOCD* EXPRESSION IN TISSUES. 42
**FIGURE S42.** THE EFFECT OF RS13039273 ON *RBM38* EXPRESSION. 43
**FIGURE S43.** MR SCATTER PLOT: UL & BMI. 44
**FIGURE S44.** MR SCATTER PLOT: UL & WAIST CIRCUMFERENCE. 45
**FIGURE S45.** MR SCATTER PLOT: UL & WHOLE BODY FAT MASS. 46
**FIGURE S46.** MR SCATTER PLOT: UL & WHOLE BODY FAT-FREE MASS. 47
**FIGURE S47.** MR SCATTER PLOT: UL & WHOLE BODY WATER MASS. 48
**FIGURE S48.** MR SCATTER PLOT: UL & IMPEDANCE OF WHOLE BODY. 49
**FIGURE S49.** MR SCATTER PLOT: UL & BASAL METABOLIC RATE. 50
**FIGURE S50.** LEAVE-ONE-OUT: EXPOSURE BMI, OUTCOME UL. 51
**FIGURE S51.** LEAVE-ONE-OUT: EXPOSURE WAIST CIRCUMFERENCE, OUTCOME UL. 52
**FIGURE S52.** LEAVE-ONE-OUT: EXPOSURE WHOLE BODY FAT MASS, OUTCOME UL. 53

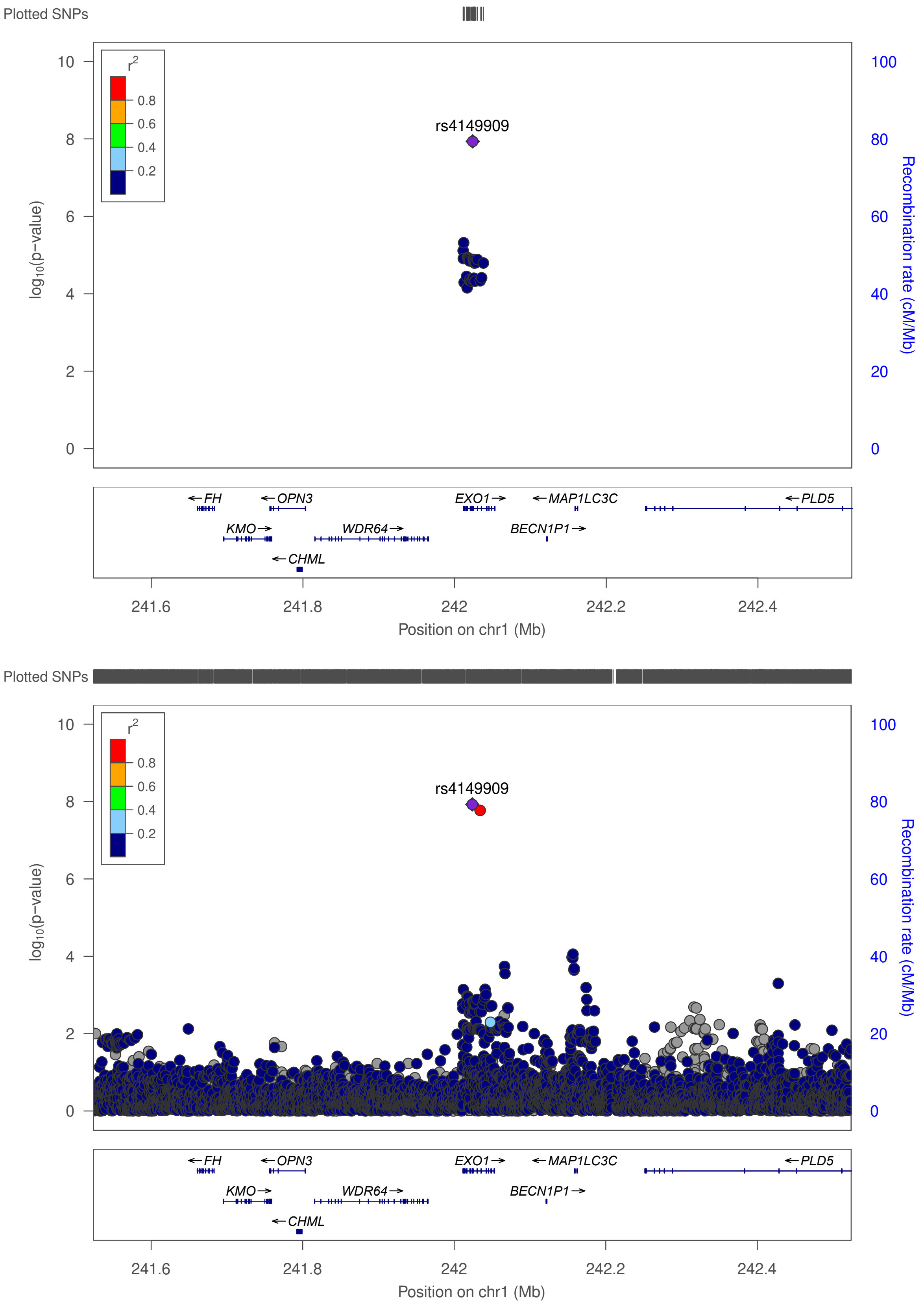

**Figure S1. Regional association plot of the novel UL association in chr1 near *EXO1*.**
The plot on top shows the result in META-1, and the plot on bottom shows the result in META-2.

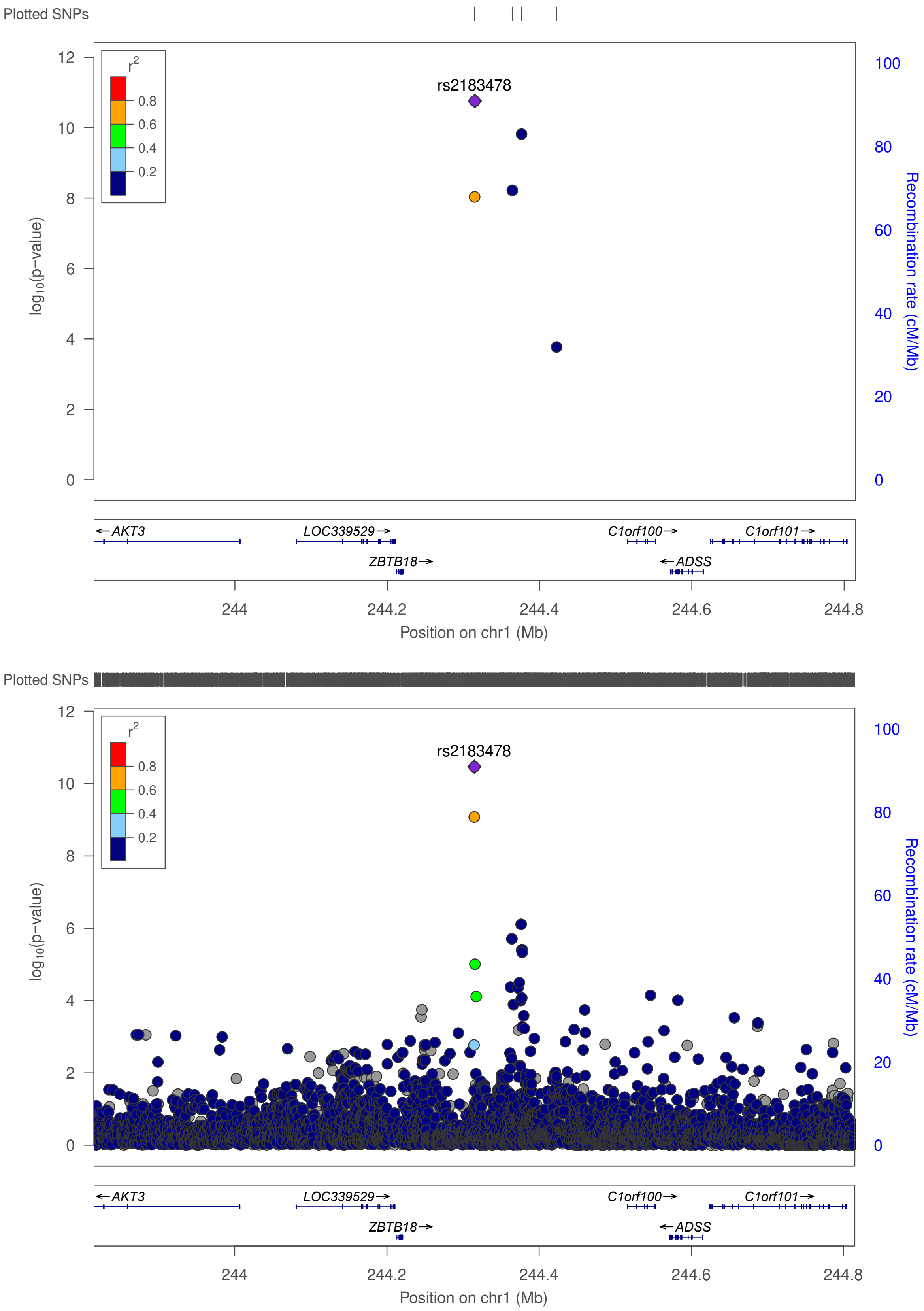

**Figure S2. Regional association plot of the novel UL association in chr1 near *ZBTB18*.**
The plot on top shows the result in META-1, and the plot on bottom shows the result in META-2.

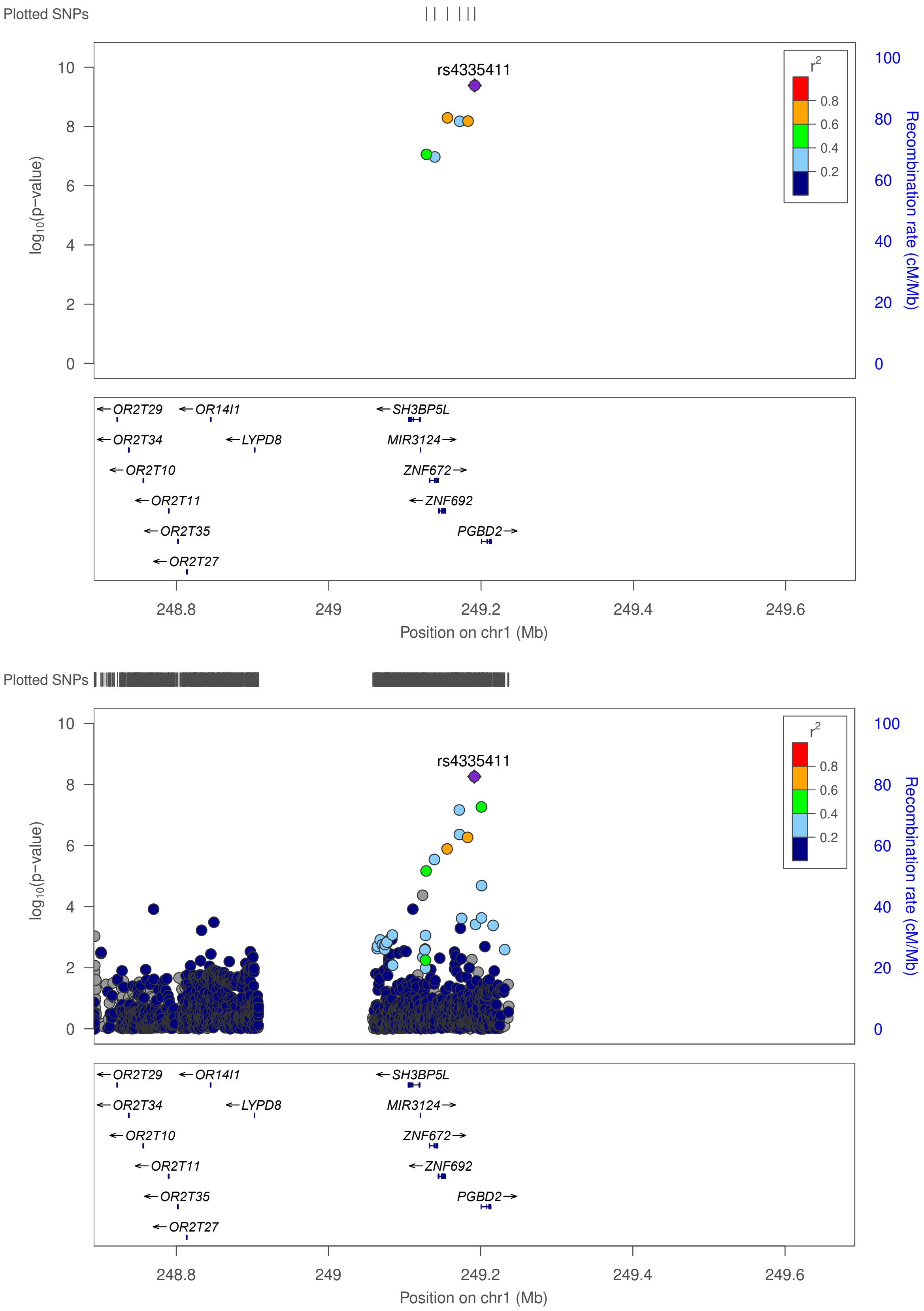

**Figure S3. Regional association plot of the novel UL association in chr1 near *PGBD2 (ZNF692)*.** The plot on top shows the result in META-1, and the plot on bottom shows the result in META-2.

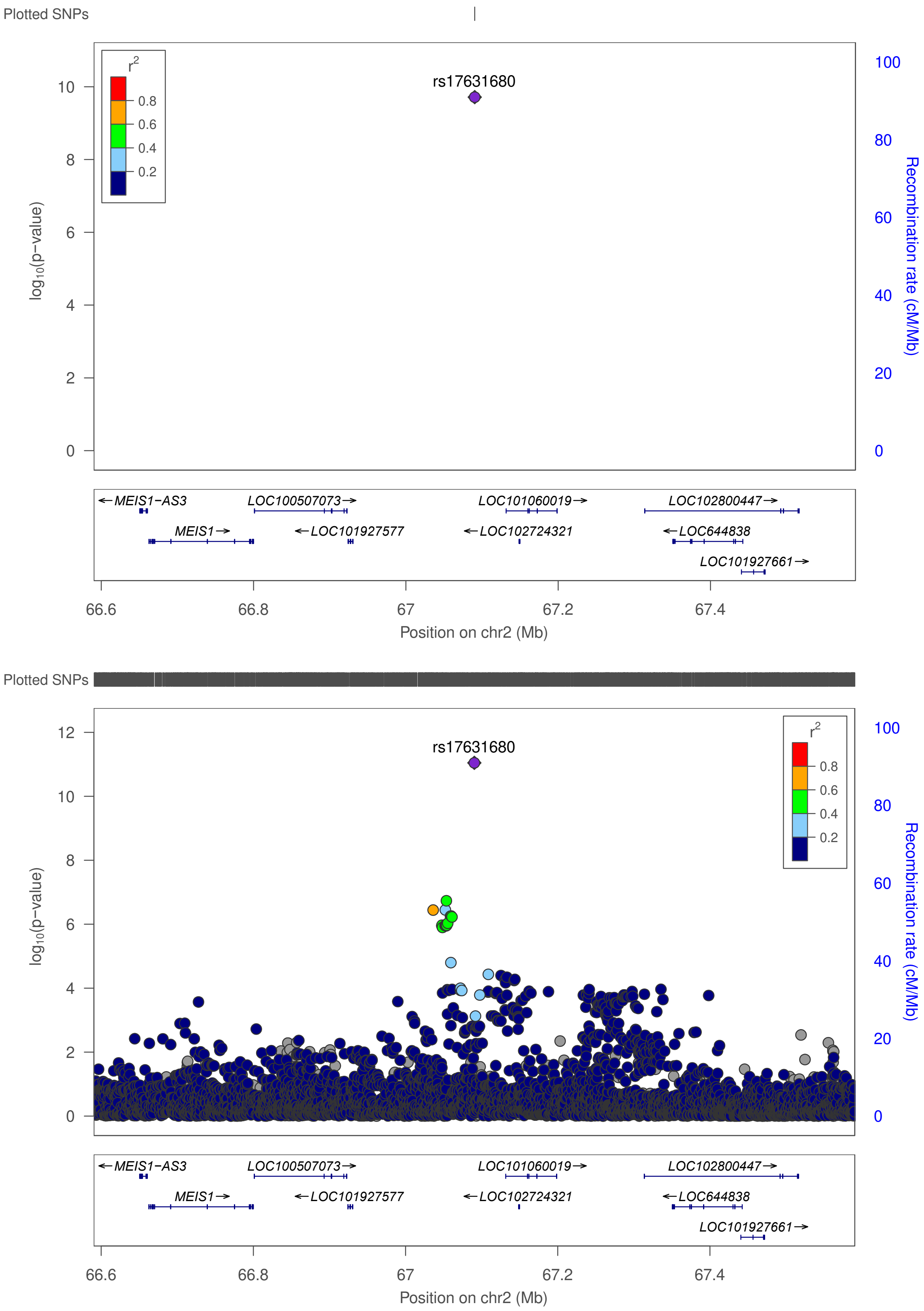

**Figure S4. Regional association plot of the novel UL association in chr2 near *MEIS1*.**
The plot on top shows the result in META-1, and the plot on bottom shows the result in META-2.

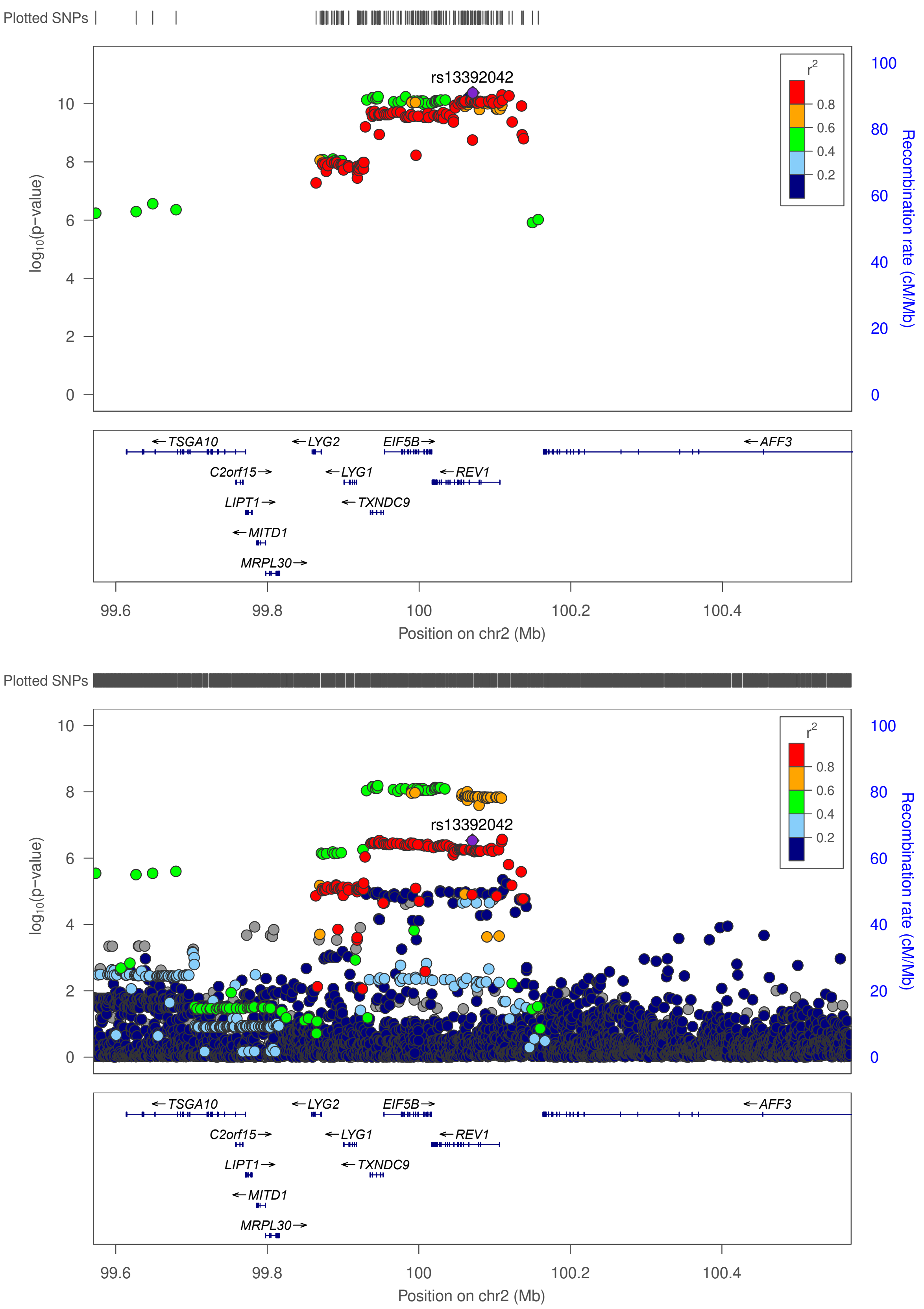

**Figure S5. Regional association plot of the novel UL association in chr2 near *REV1*.**
The plot on top shows the result in META-1, and the plot on bottom shows the result in META-2.

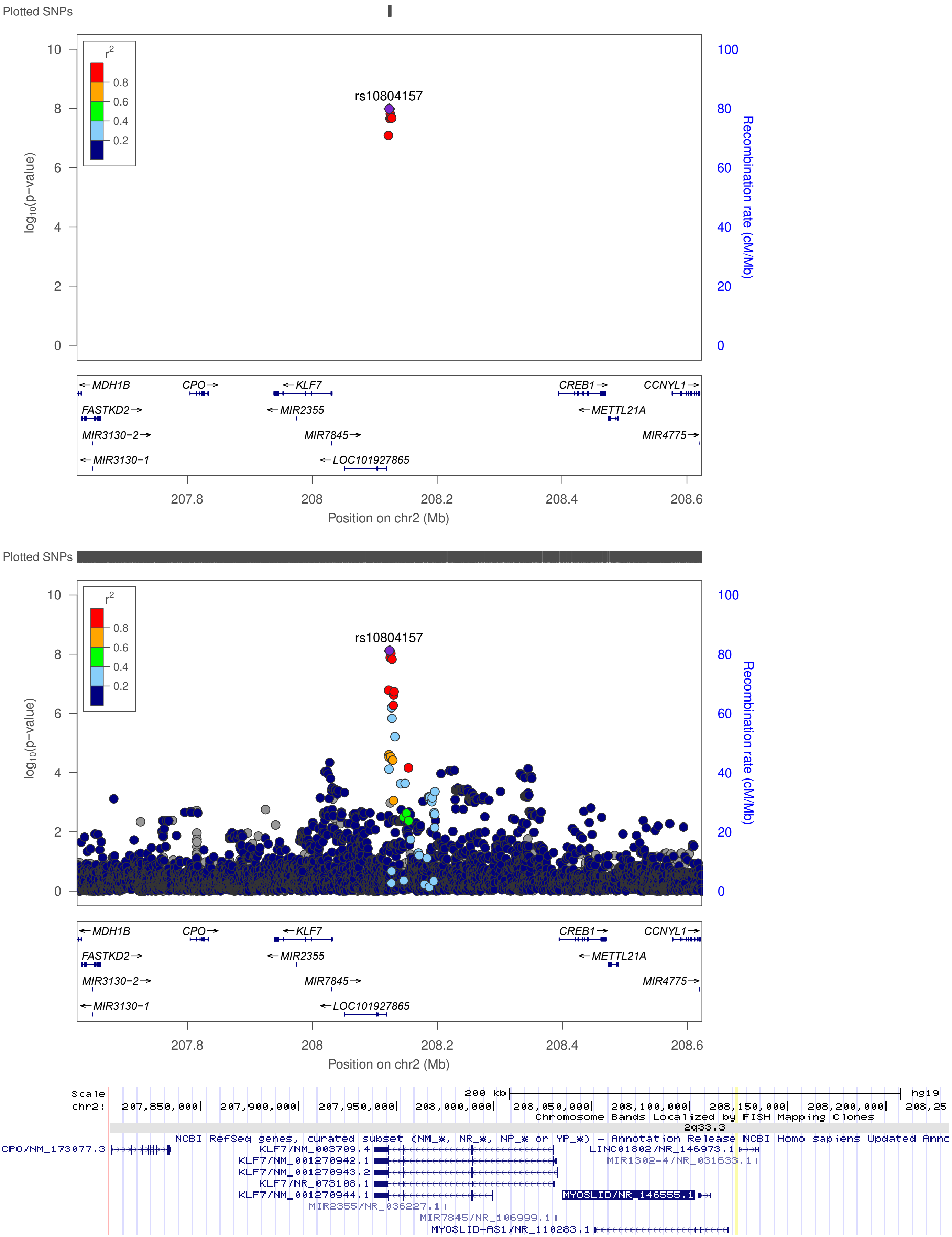

**Figure S6. Regional association plot of the novel UL association in chr2 near *MYOSLID*.**
The plot on top shows the result in META-1, and the plot in the middle shows the result in META-2. *MYOSLID* location was not available for LocusZoom: it locates ~78kb downstream from *KLF7*, as indicated in the image extracted from UCSC Genome Browser (bottom).

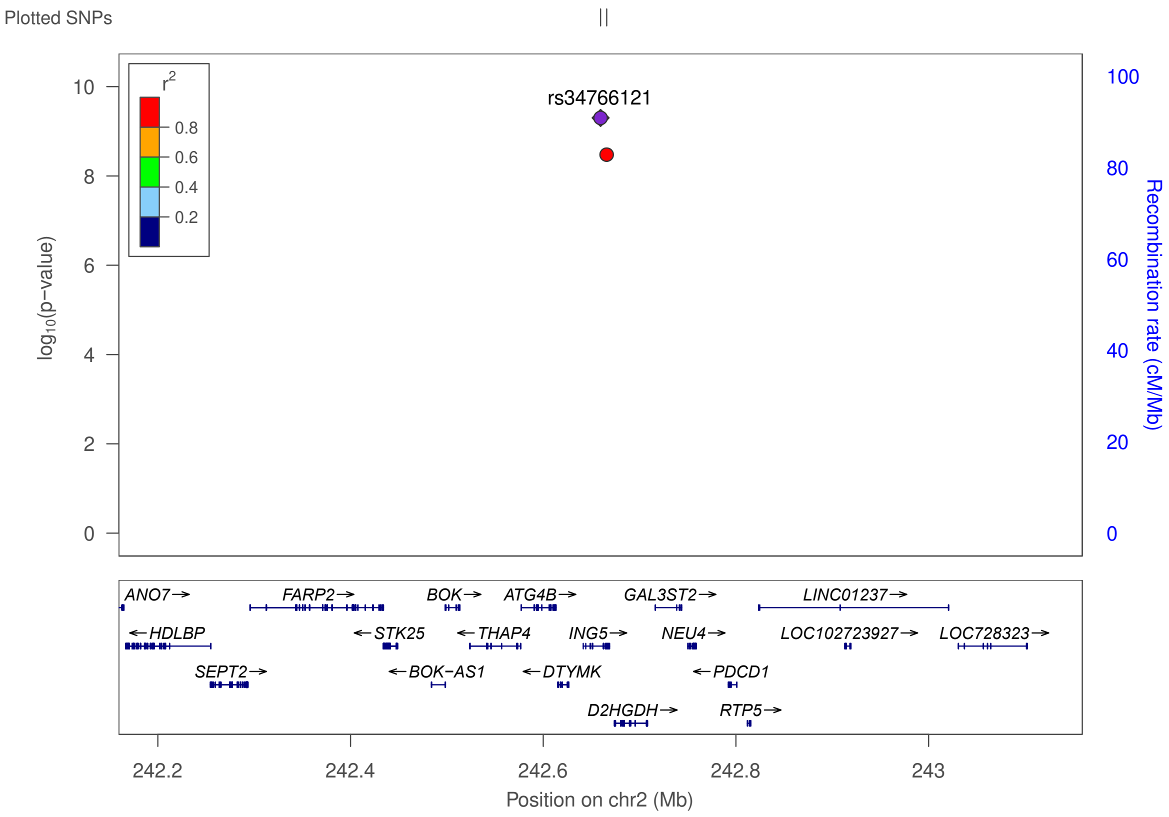

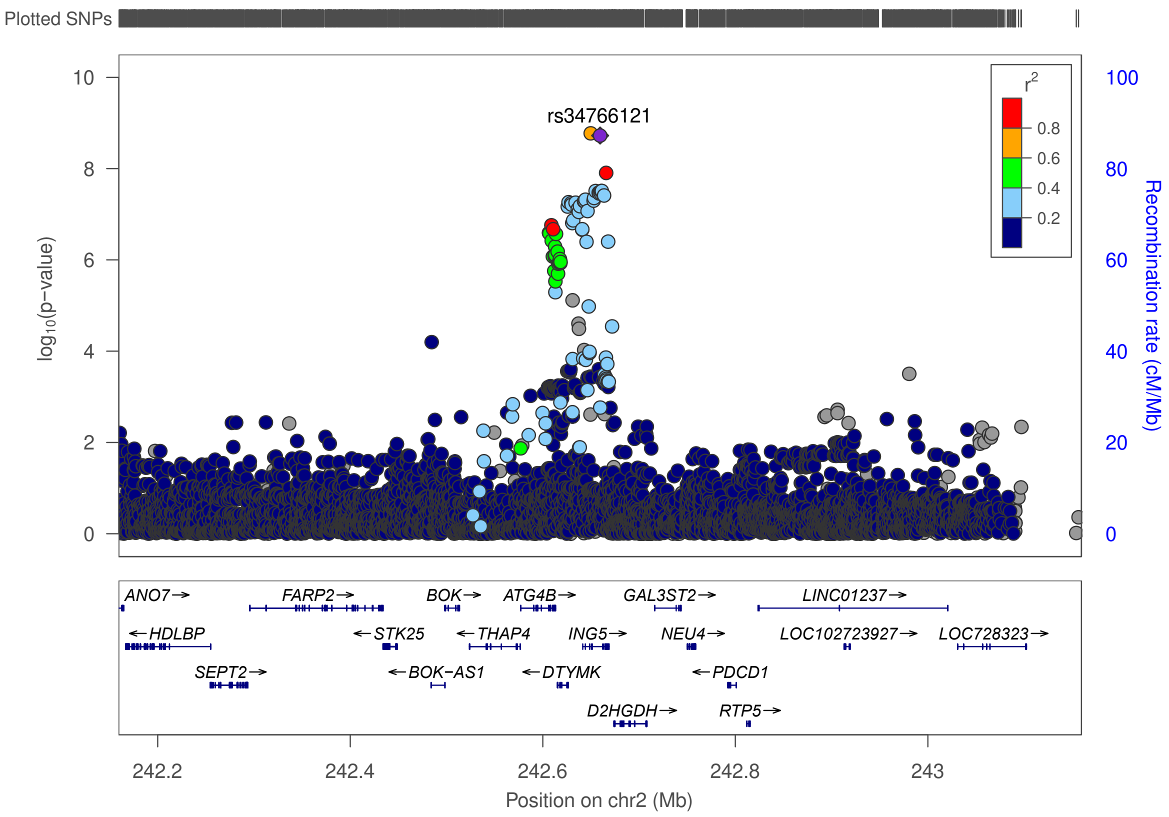

**Figure S7. Regional association plot of the novel UL association in chr2 near *ING5*.**
The plot on top shows the result in META-1, and the plot on bottom shows the result in META-2.

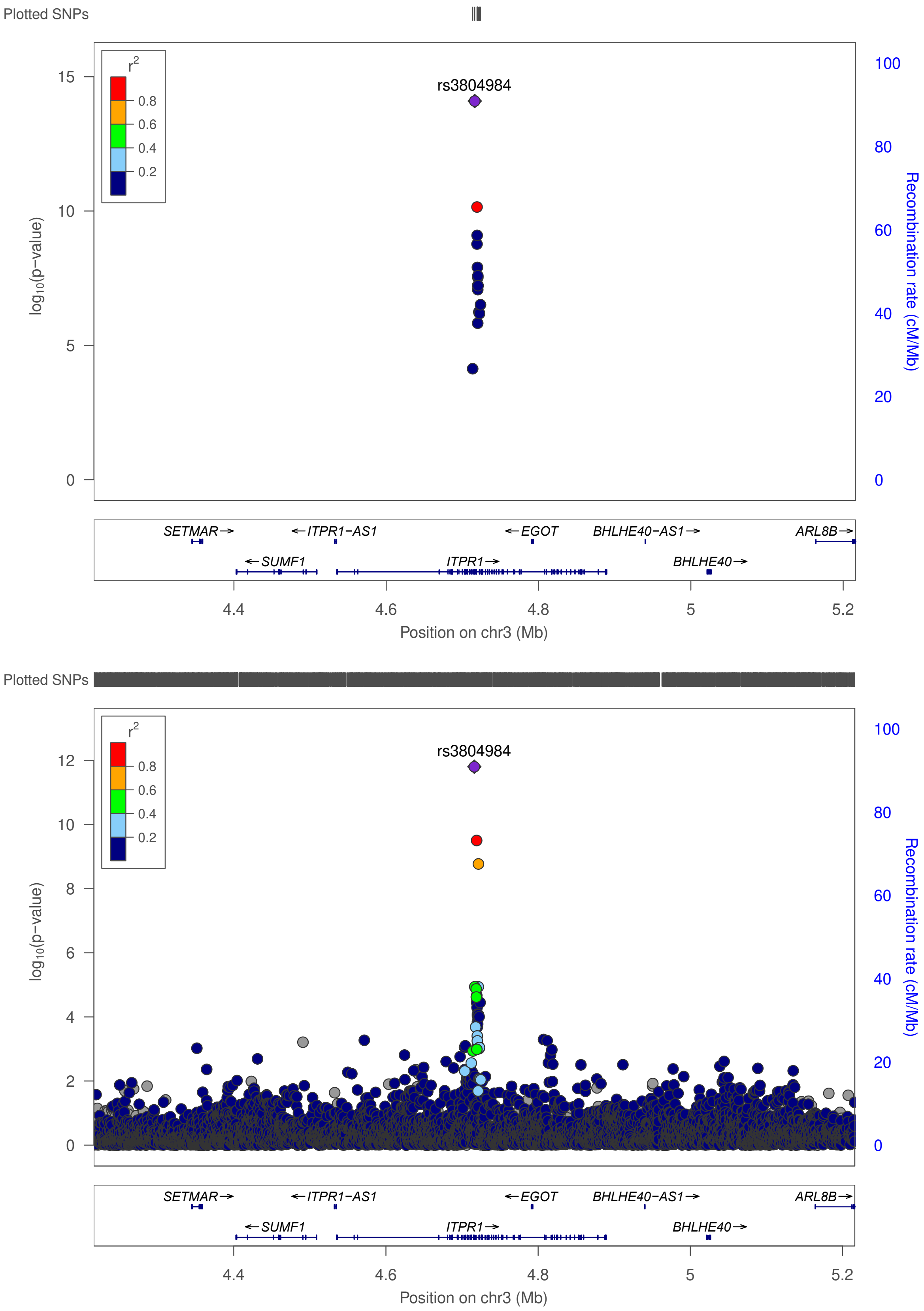

**Figure S8. Regional association plot of the novel UL association in chr3 near *ITPR1*.**
The plot on top shows the result in META-1, and the plot on bottom shows the result in META-2.

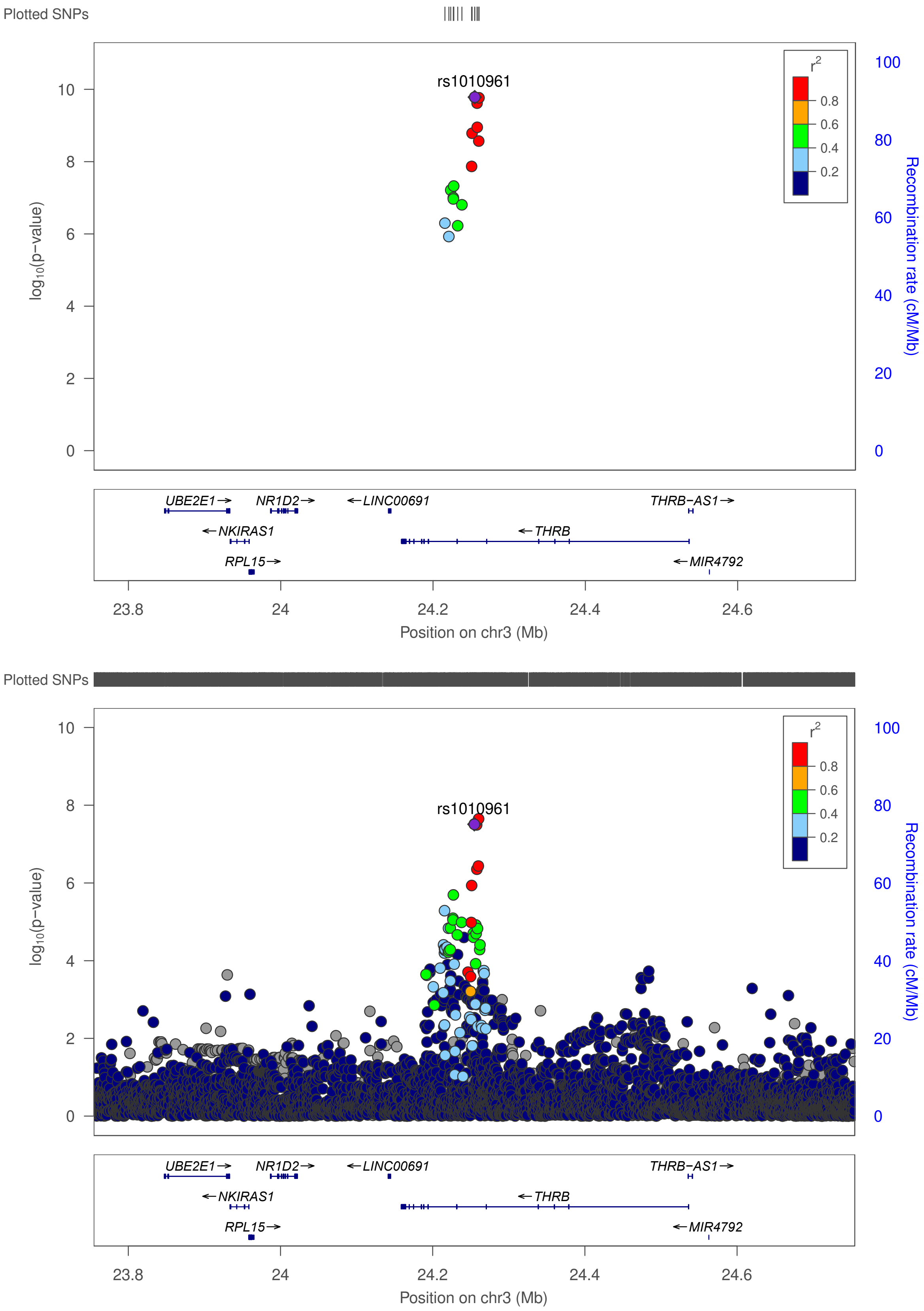

**Figure S9. Regional association plot of the novel UL association in chr3 near *THRB*.**
The plot on top shows the result in META-1, and the plot on bottom shows the result in META-2.

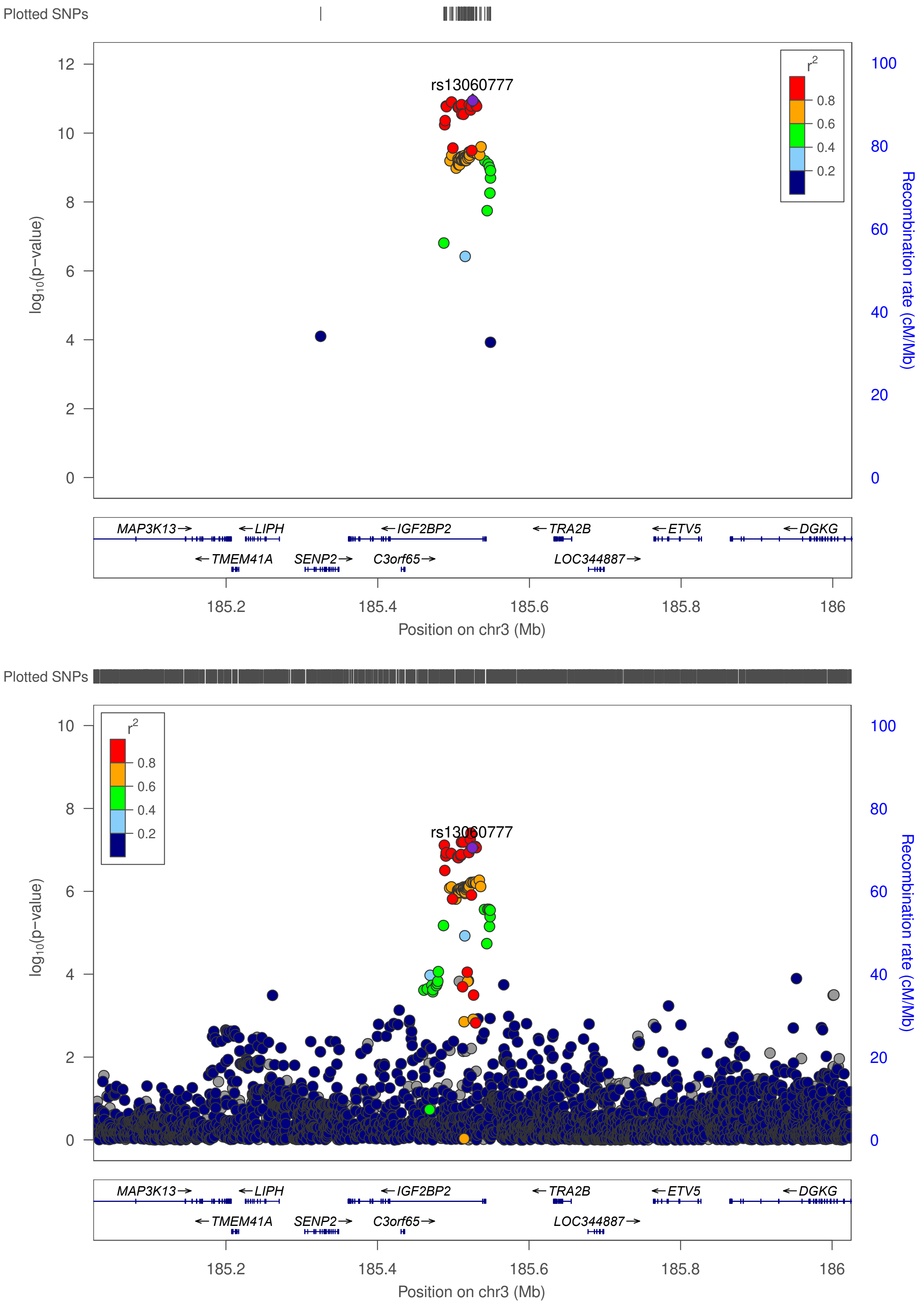

**Figure S10. Regional association plot of the novel UL association in chr3 near *IGF2BP2*.** The plot on top shows the result in META-1, and the plot on bottom shows the result in META-2.

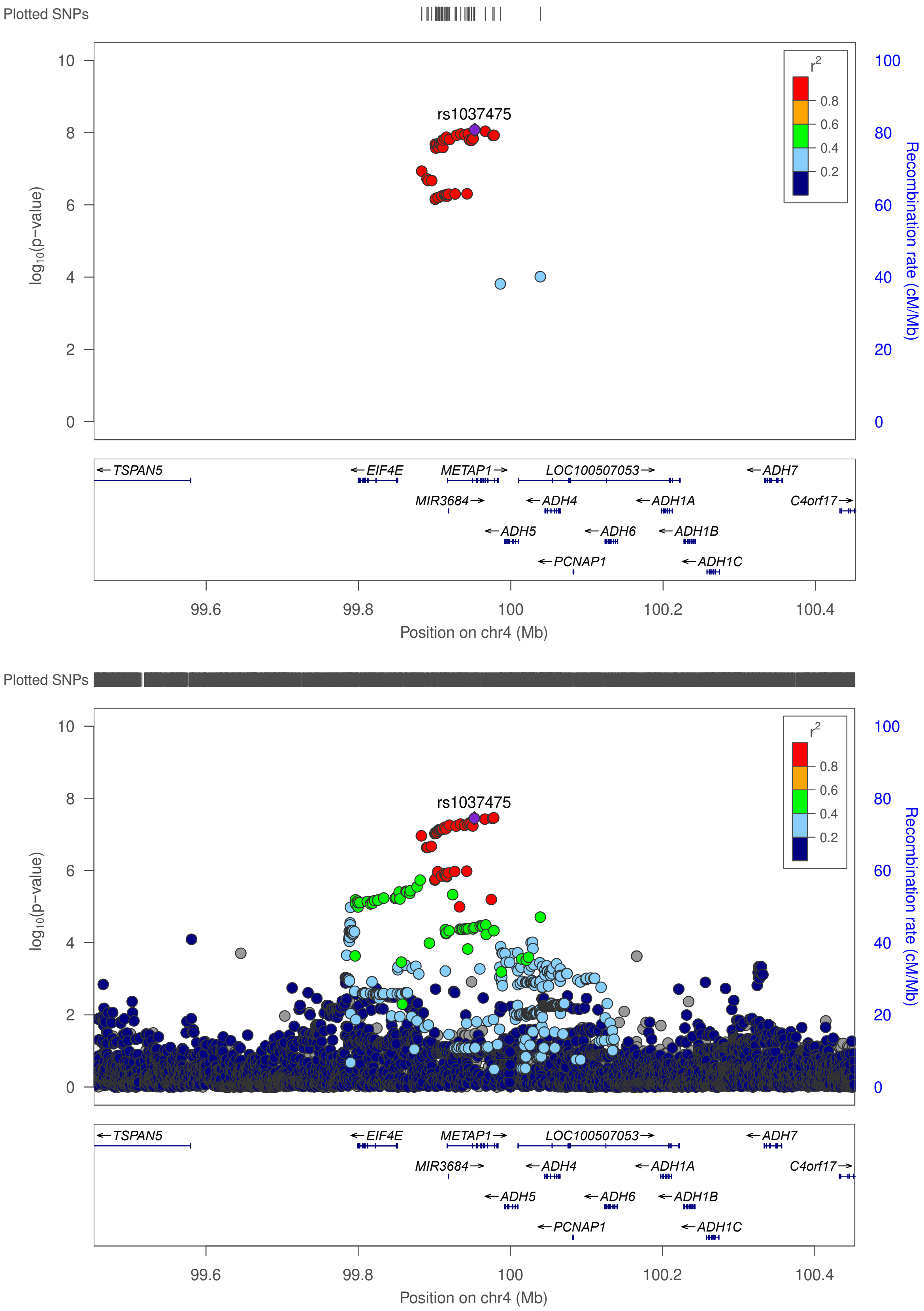

**Figure S11. Regional association plot of the novel UL association in chr4 near *METAP1 (EIF4E, ADH5)*.** The plot on top shows the result in META-1, and the plot on bottom shows the result in META-2.

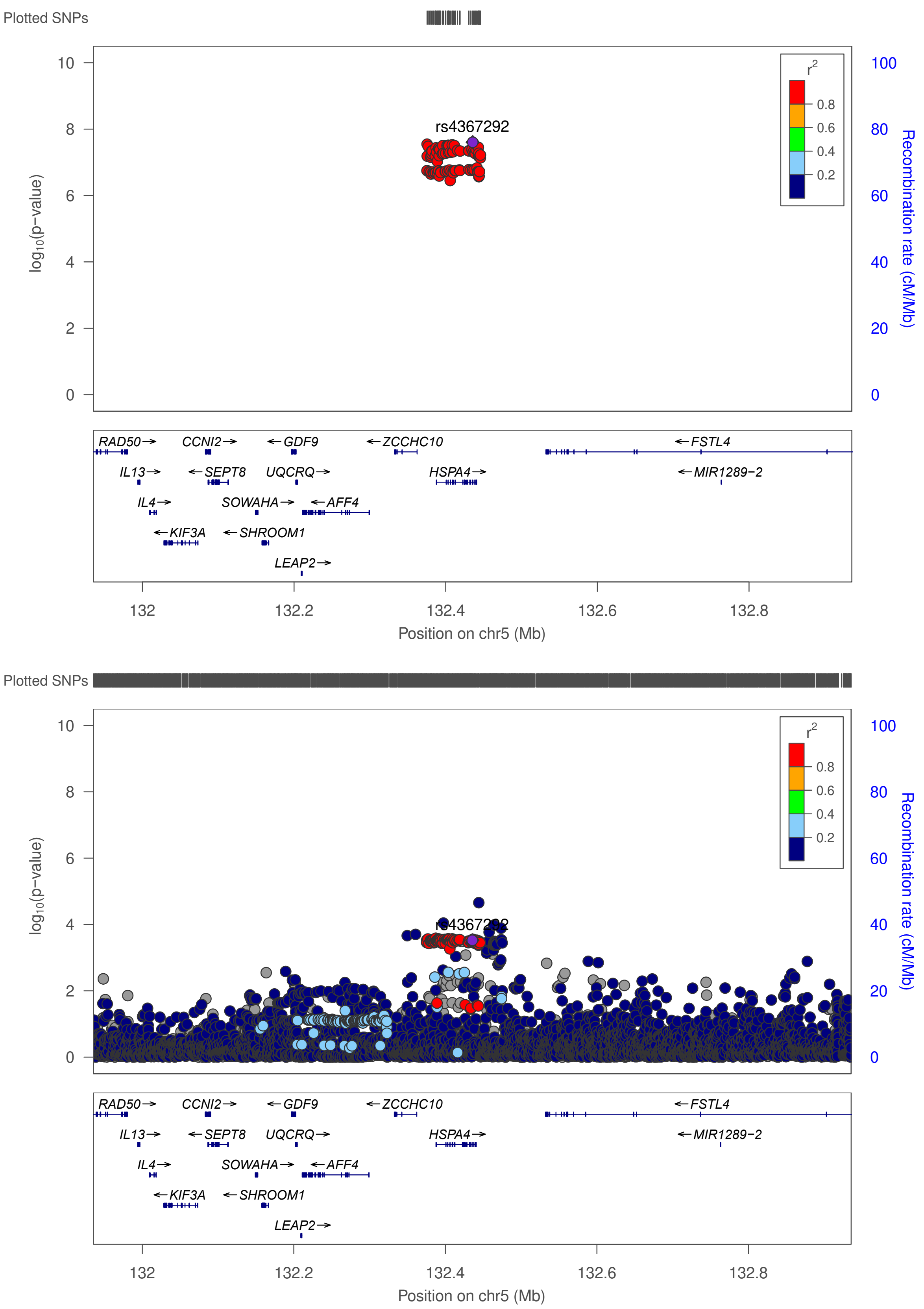

**Figure S12. Regional association plot of the novel UL association in chr5 near *HSPA4*.**
The plot on top shows the result in META-1, and the plot on bottom shows the result in META-2.

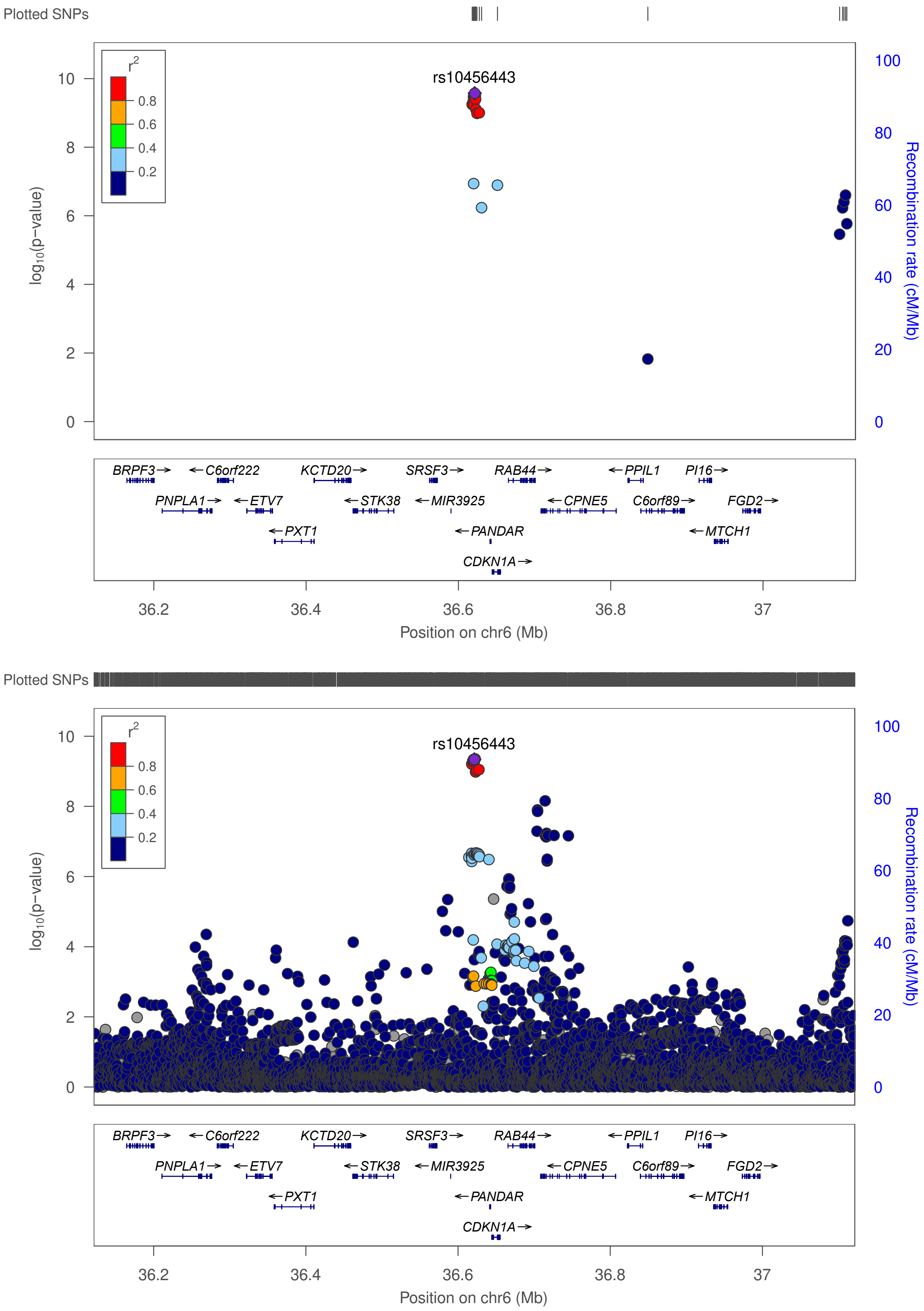

**Figure S13. Regional association plot of the novel UL association in chr6 near *CDKN1A*.** The plot on top shows the result in META-1, and the plot on bottom shows the result in META-2.

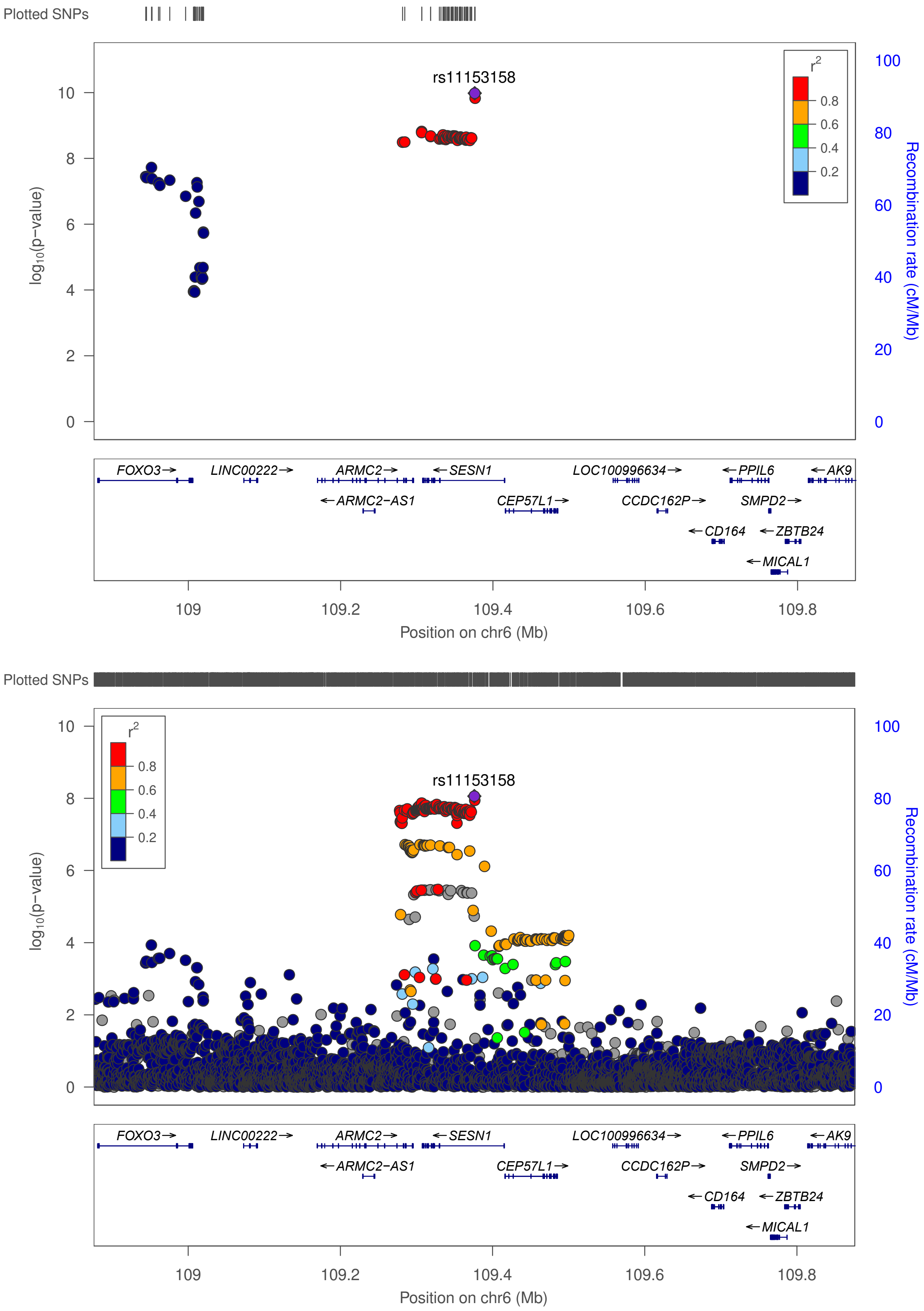

**Figure S14. Regional association plot of the novel UL association in chr6 near *SESN1*.**
The plot on top shows the result in META-1, and the plot on bottom shows the result in META-2.

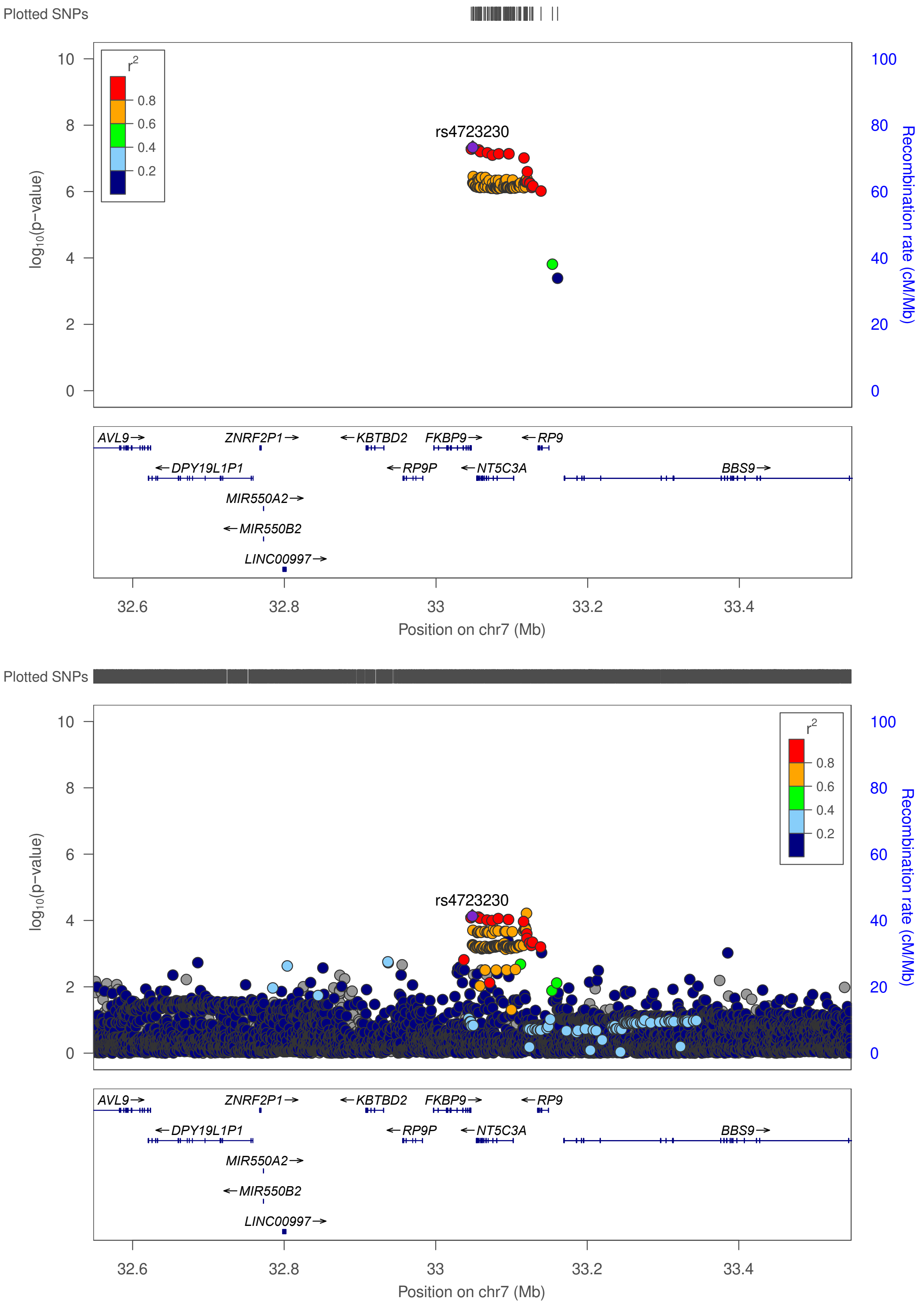

**Figure S15. Regional association plot of the novel UL association in chr7 near *NT5C3A*.**
The plot on top shows the result in META-1, and the plot on bottom shows the result in META-2.

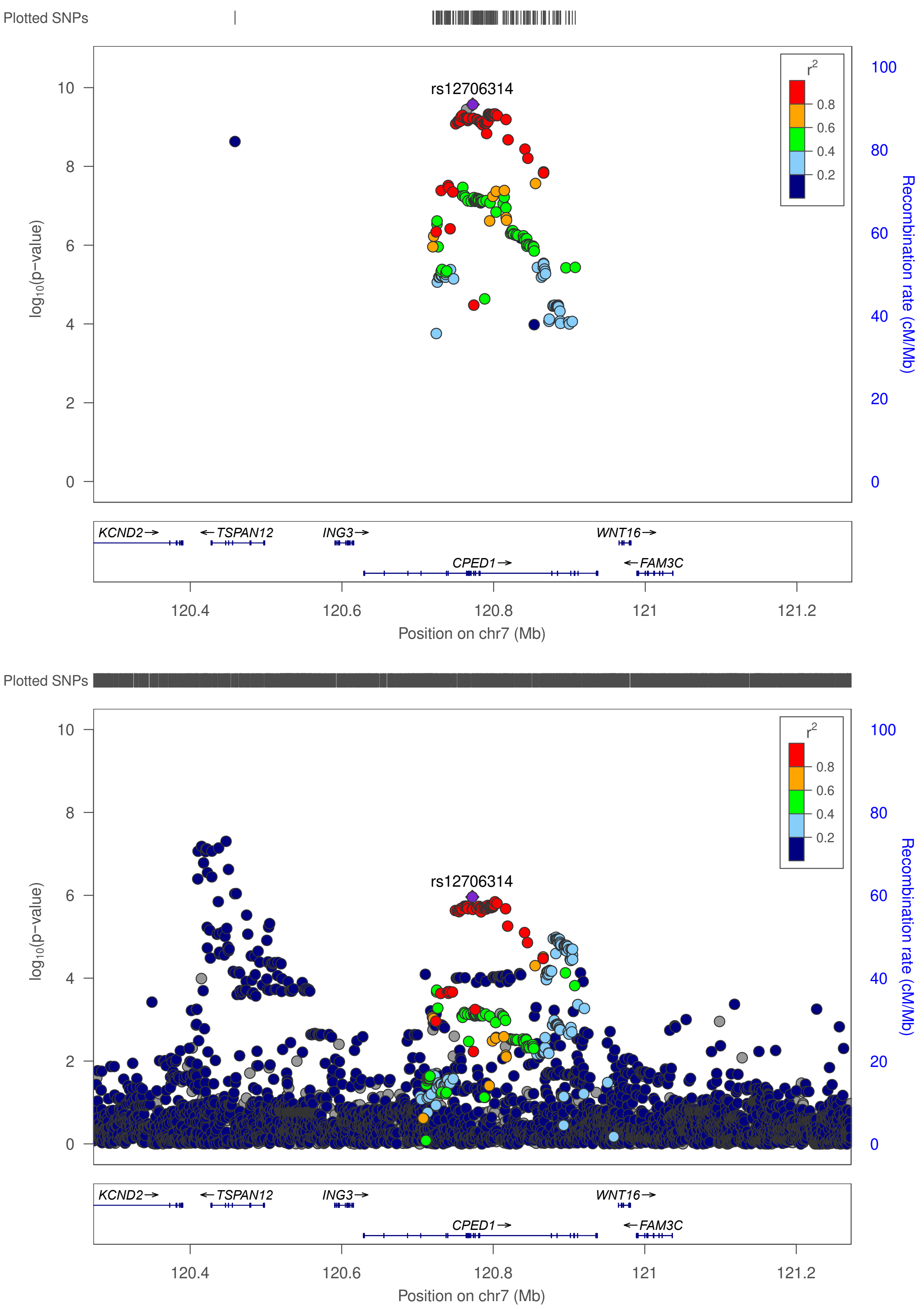

**Figure S16. Regional association plot of the novel UL association in chr7 near *CPED1 (WNT16)*.** The plot on top shows the result in META-1, and the plot on bottom shows the result in META-2.

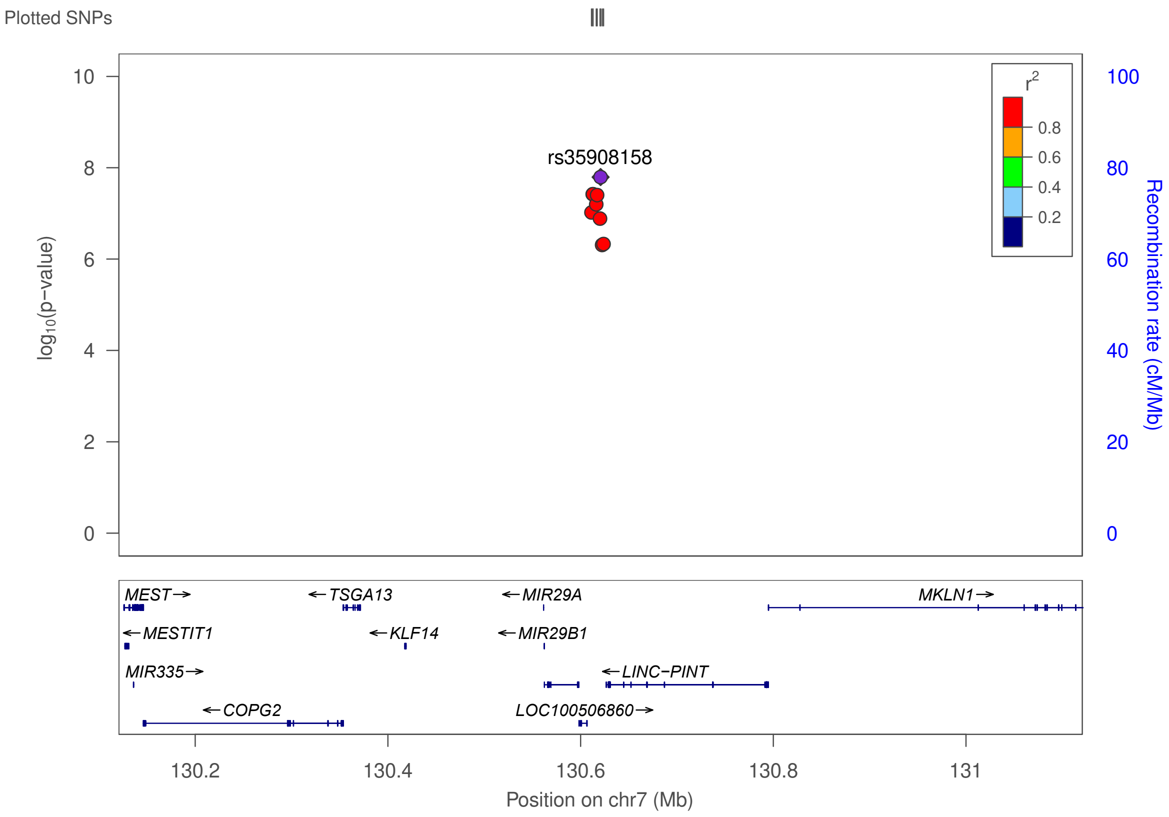

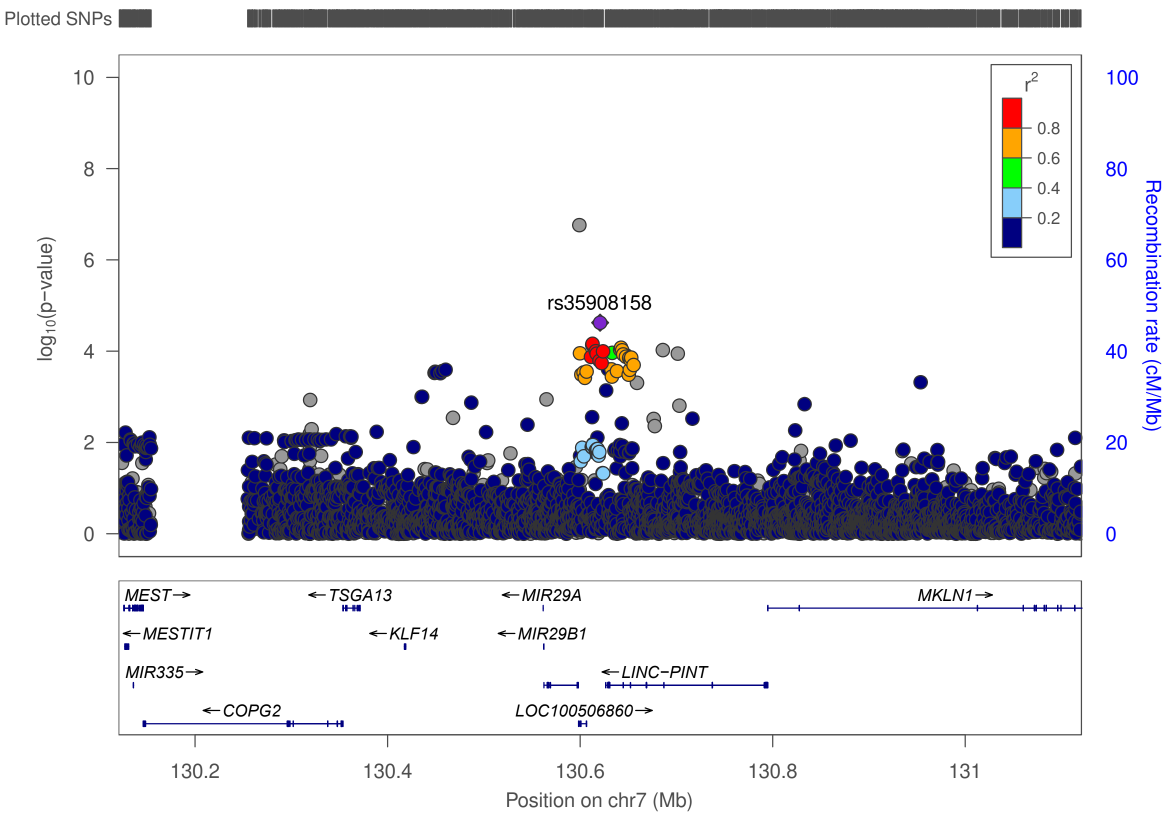

**Figure S17. Regional association plot of the novel UL association in chr7 near *LINC-PINT*.** The plot on top shows the result in META-1, and the plot on bottom shows the result in META-2.

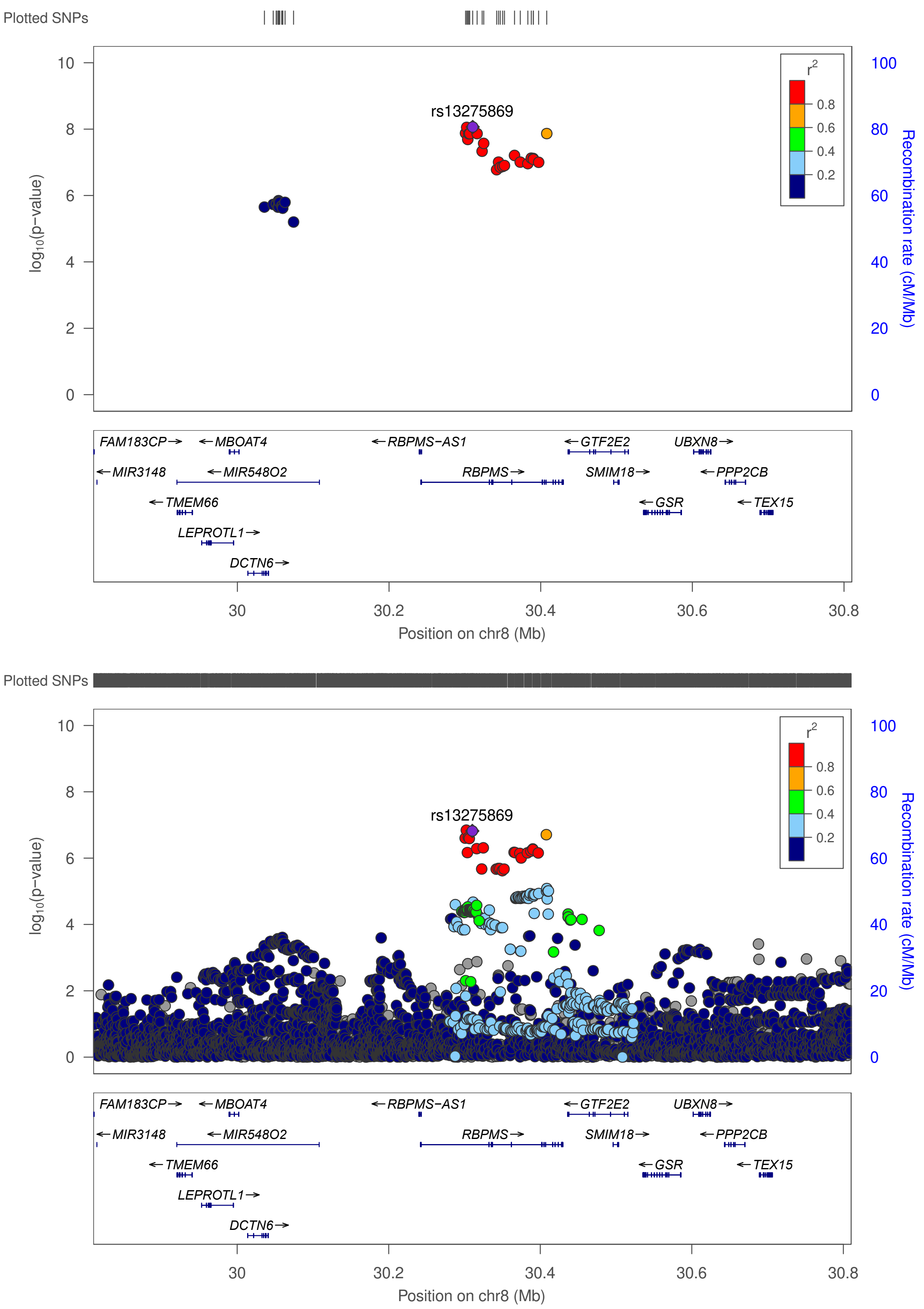

**Figure S18. Regional association plot of the novel UL association in chr8 near *RBPMS*.** The plot on top shows the result in META-1, and the plot on bottom shows the result in META-2.

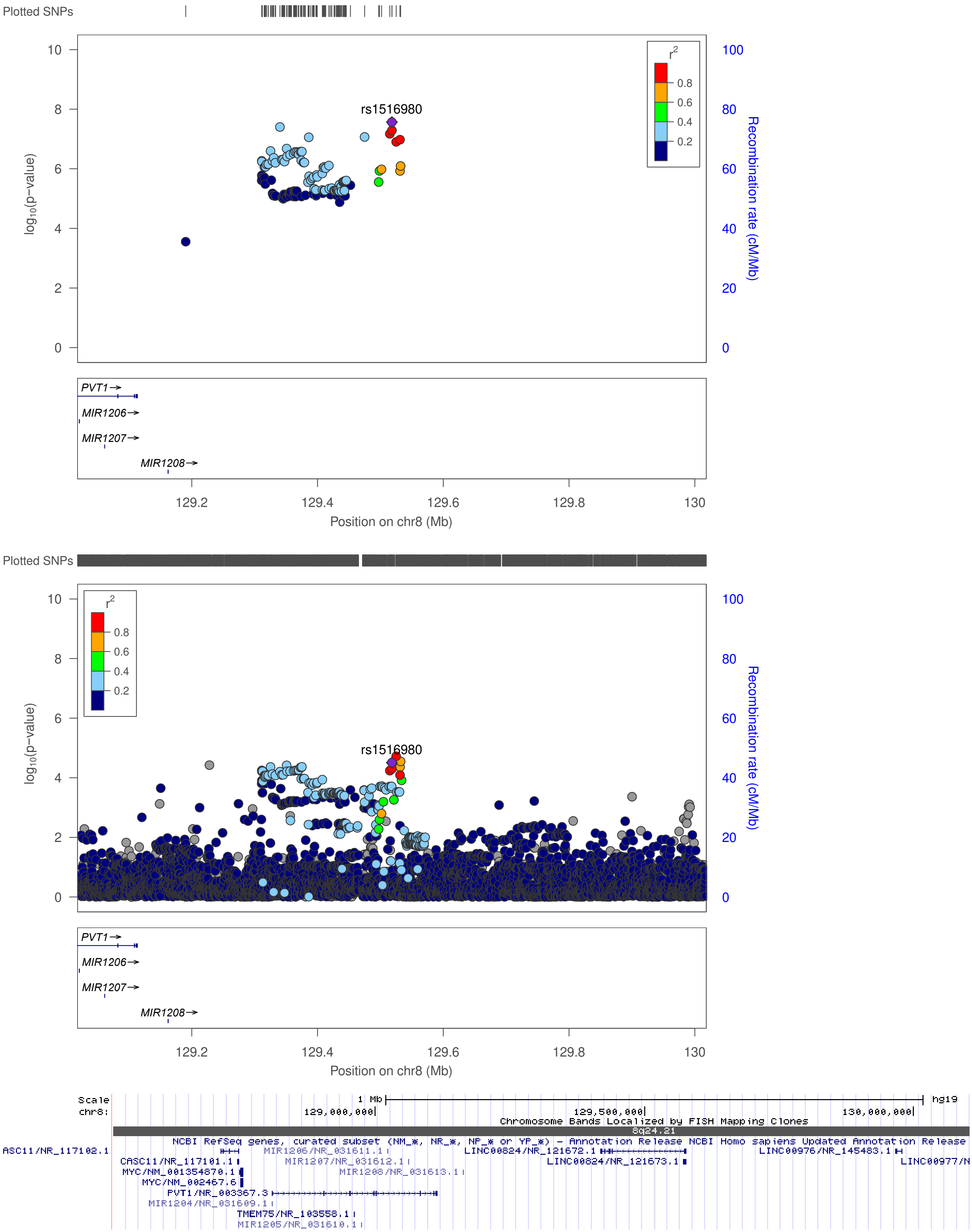

**Figure S19. Regional association plot of the novel UL association in chr8 near *LINC00824*.**
The plot on top shows the result in META-1, and the plot in the middle shows the result in META-2. *LINC00824* location was not available for LocusZoom: it locates ~300kb downstream from *PVT1*, as indicated in the image extracted from UCSC Genome Browser (bottom).

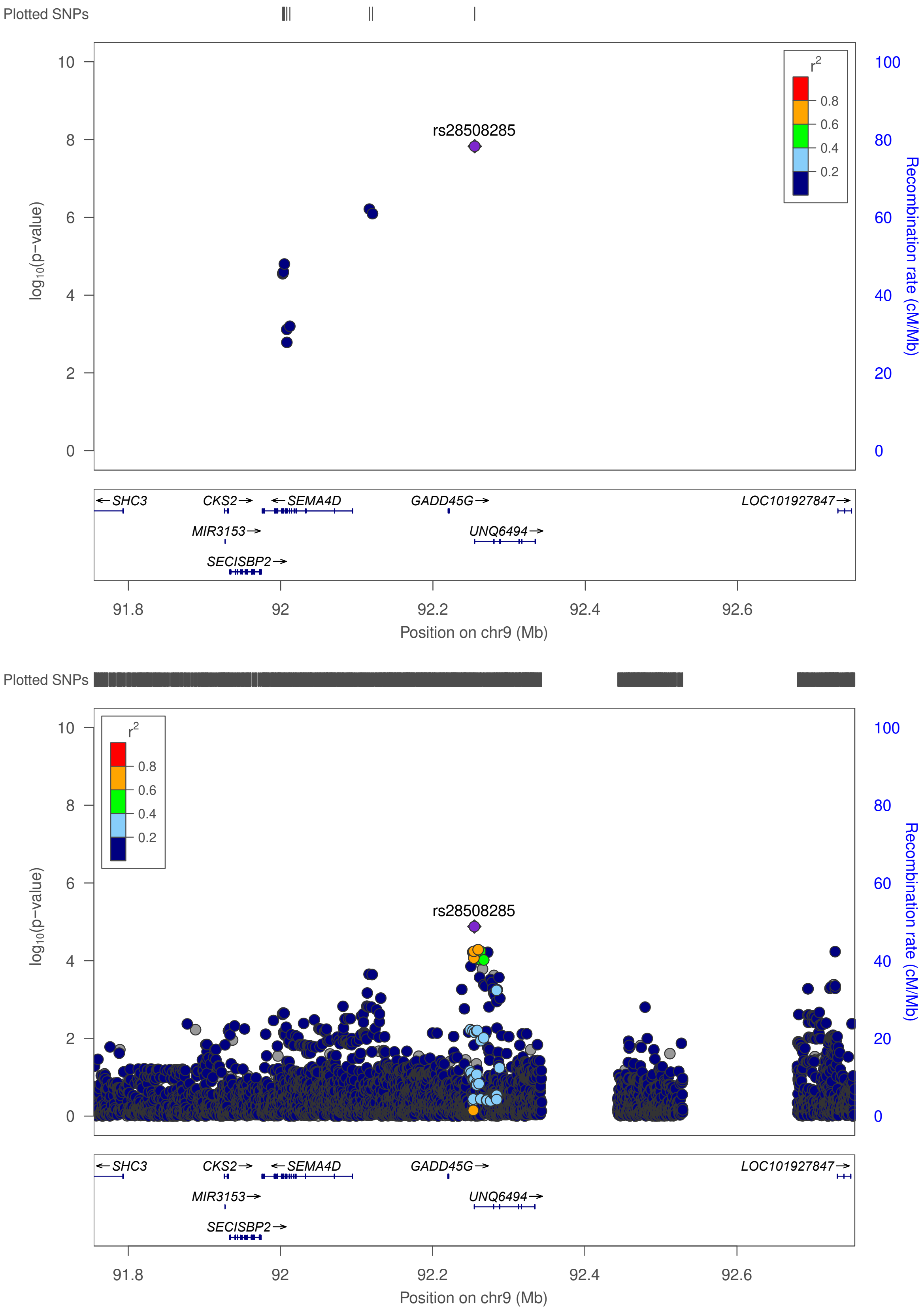

**Figure S20. Regional association plot of the novel UL association in chr9 near *GADD45G*.** The plot on top shows the result in META-1, and the plot on bottom shows the result in META-2.

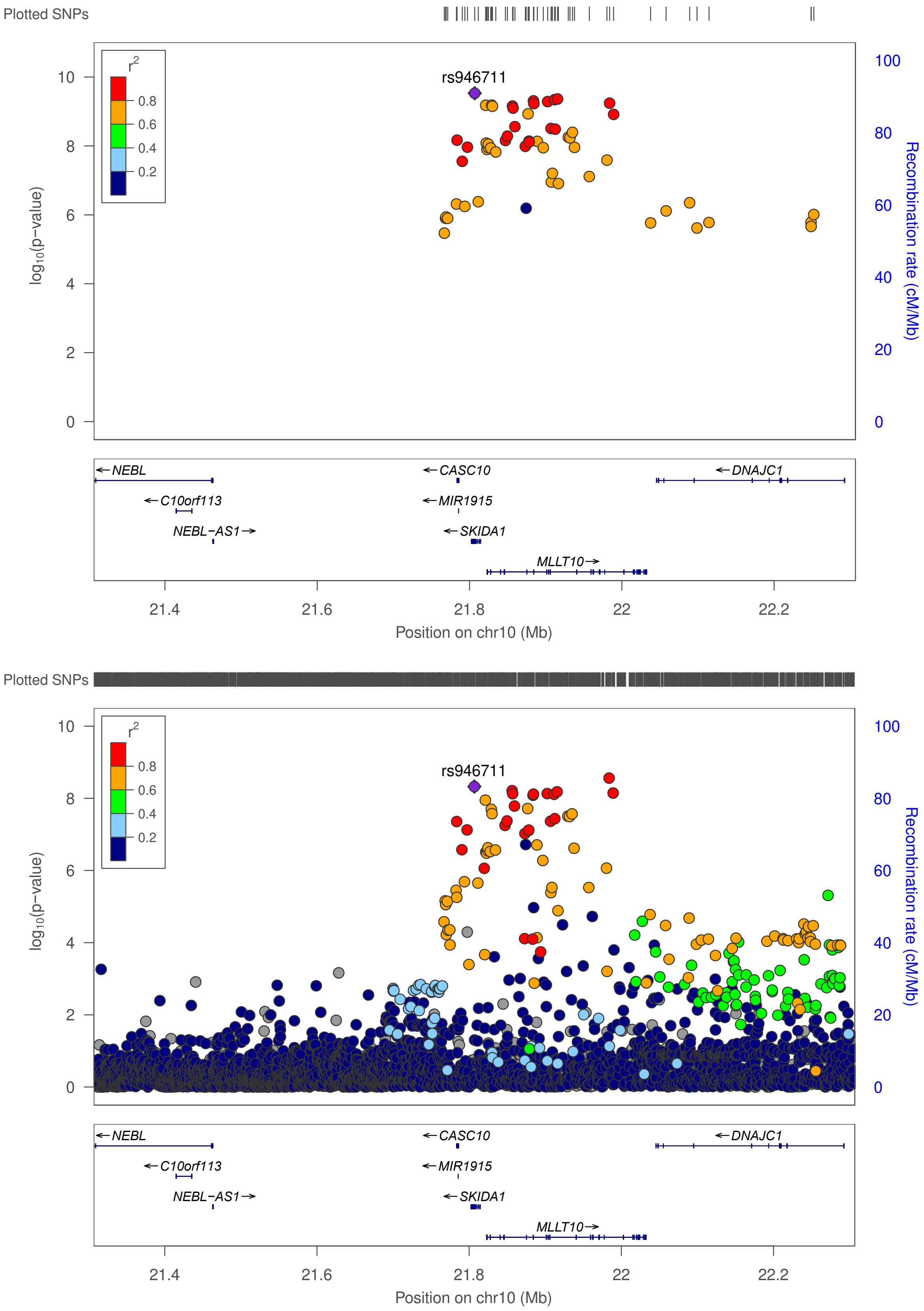

**Figure S21. Regional association plot of the novel UL association in chr10 near *SKIDA1 (DNAJC1)*.** The plot on top shows the result in META-1, and the plot on bottom shows the result in META-2.

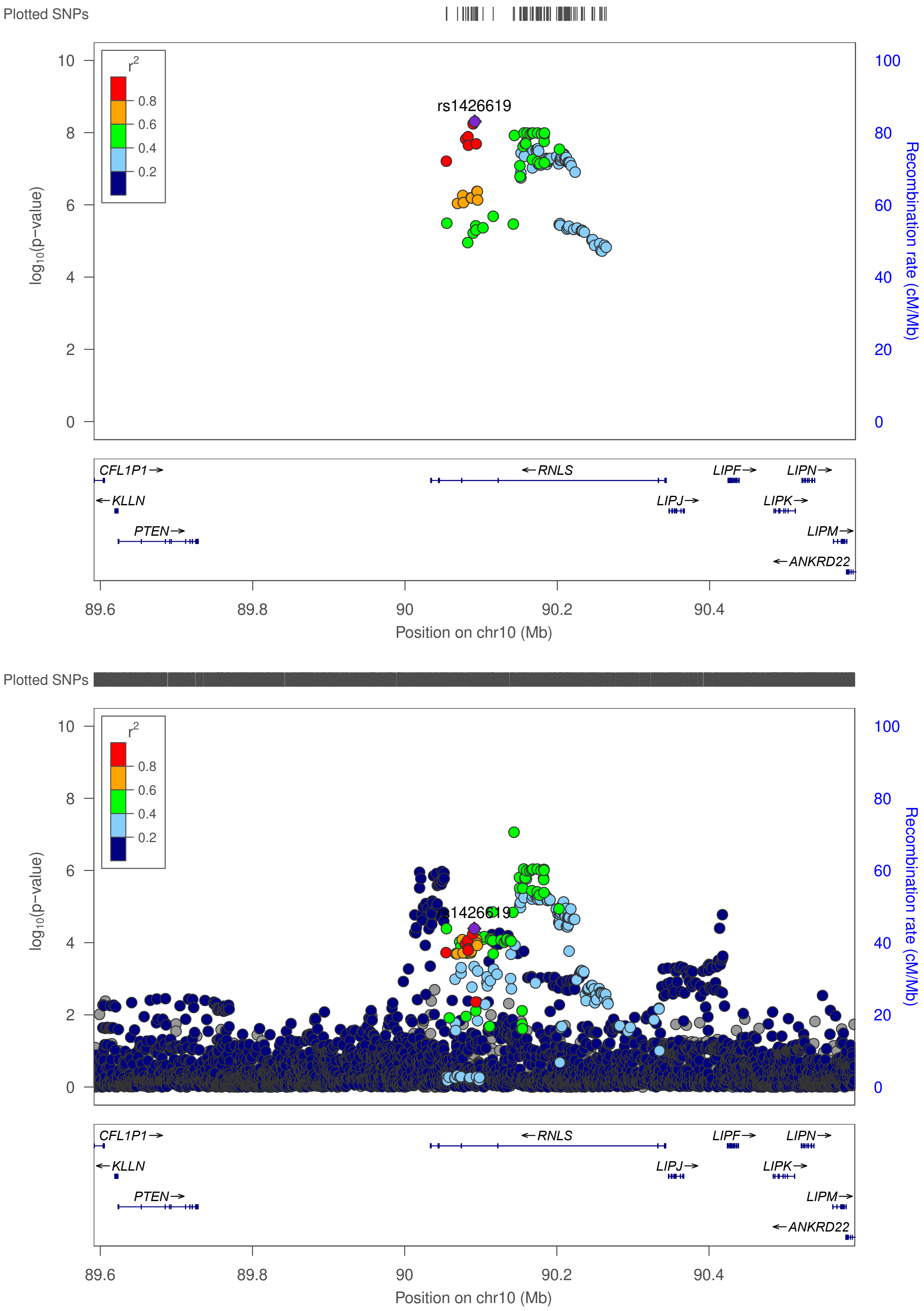

**Figure S22. Regional association plot of the novel UL association in chr10 near *RNLS*.**
The plot on top shows the result in META-1, and the plot on bottom shows the result in META-2.

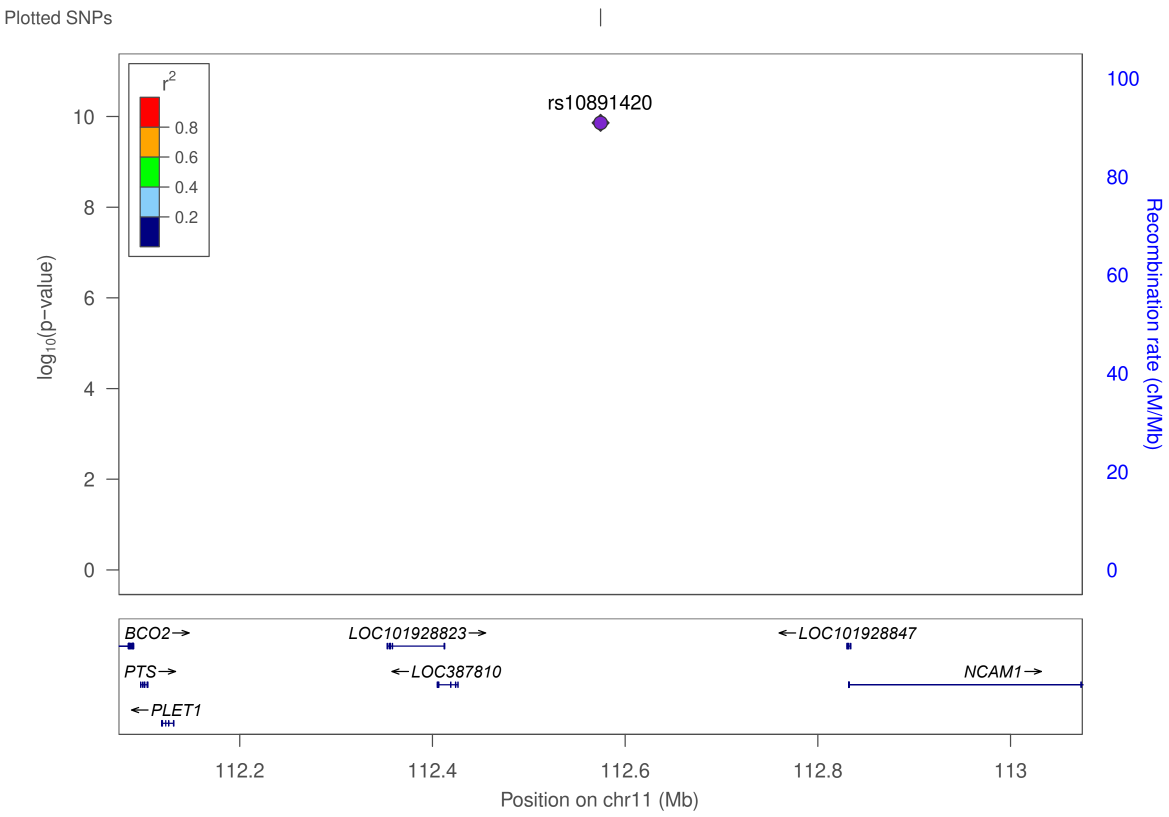

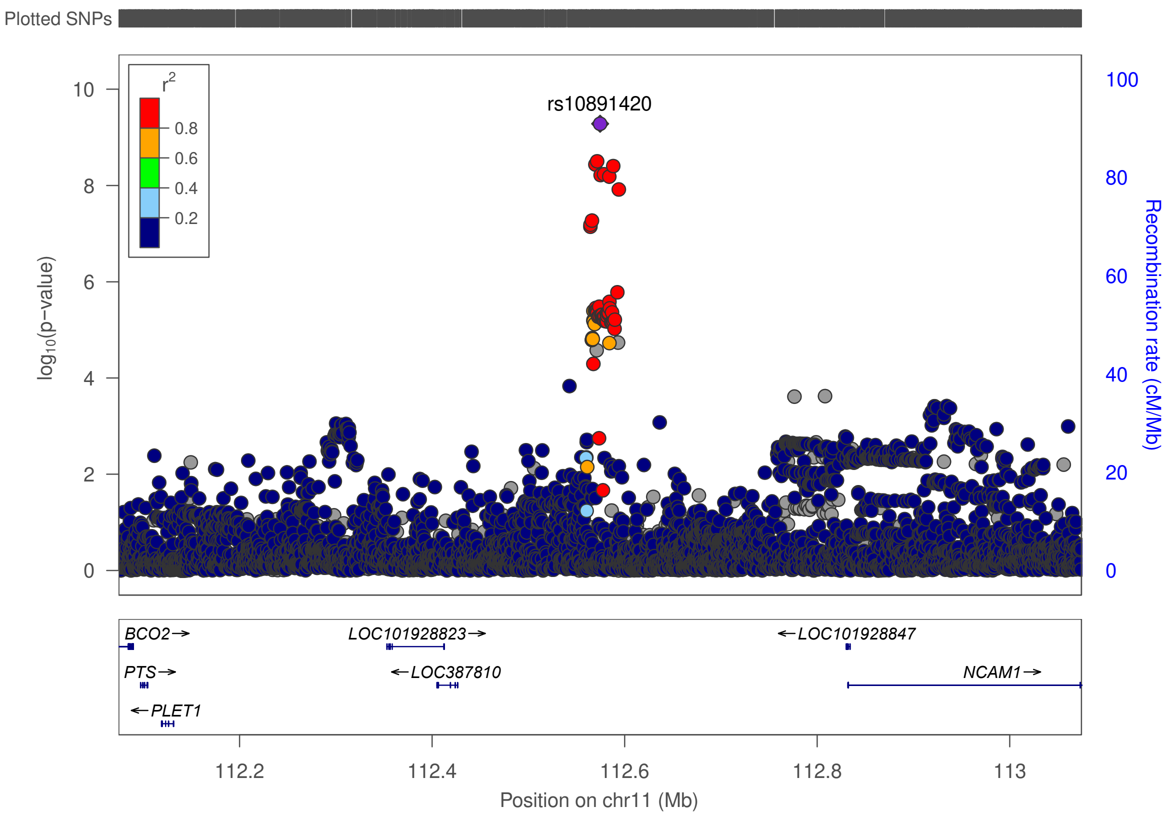

**Figure S23. Regional association plot of the novel UL association chr11 near *NCAM1*.**
The plot on top shows the result in META-1, and the plot on bottom shows the result in META-2.

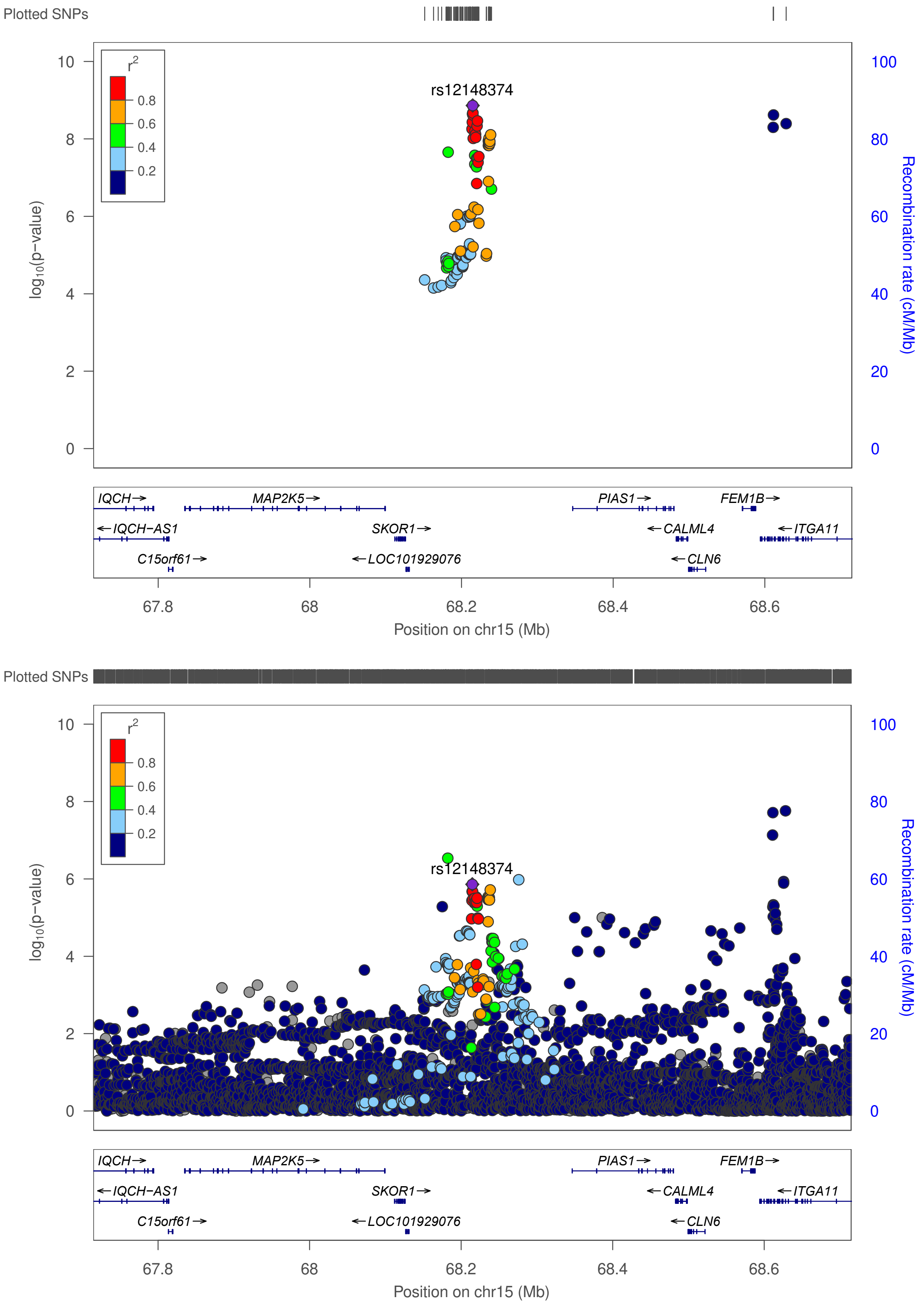

**Figure S24. Regional association plot of the novel UL association in chr15 near *SKOR1 (PIAS1)*.** The plot on top shows the result in META-1, and the plot on bottom shows the result in META-2.

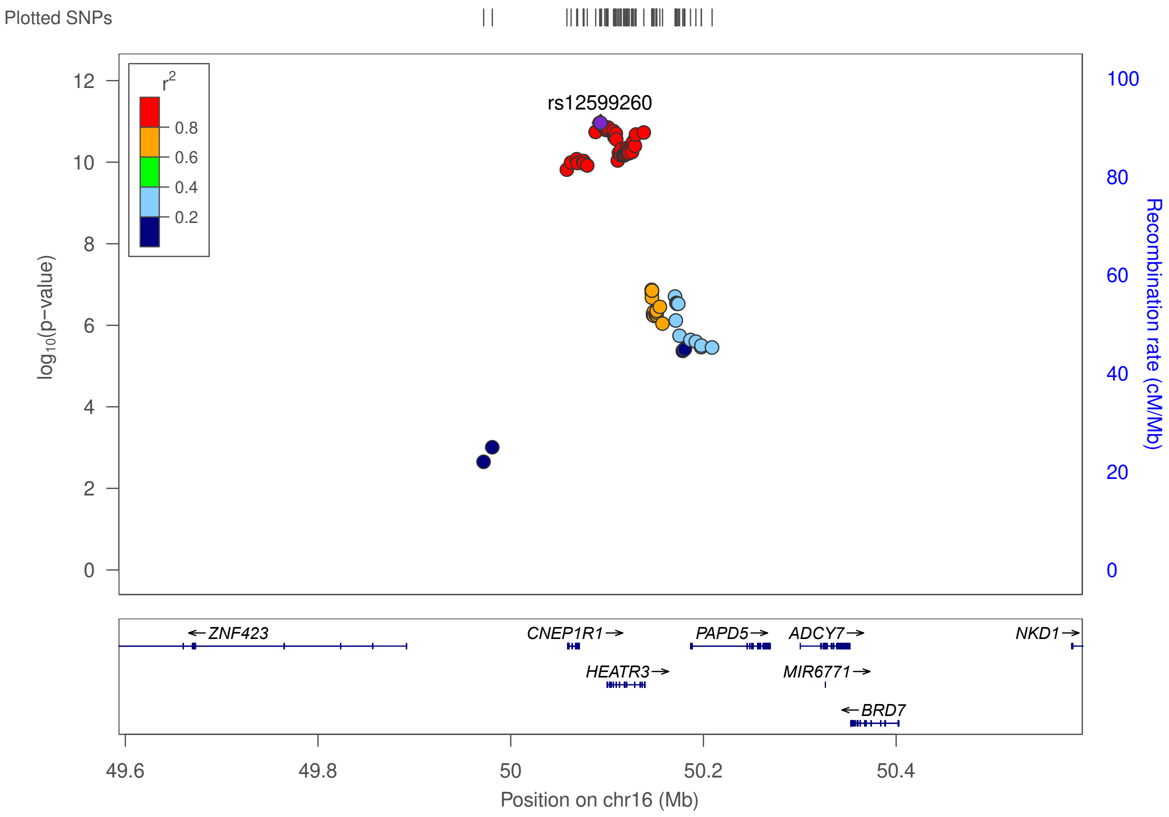

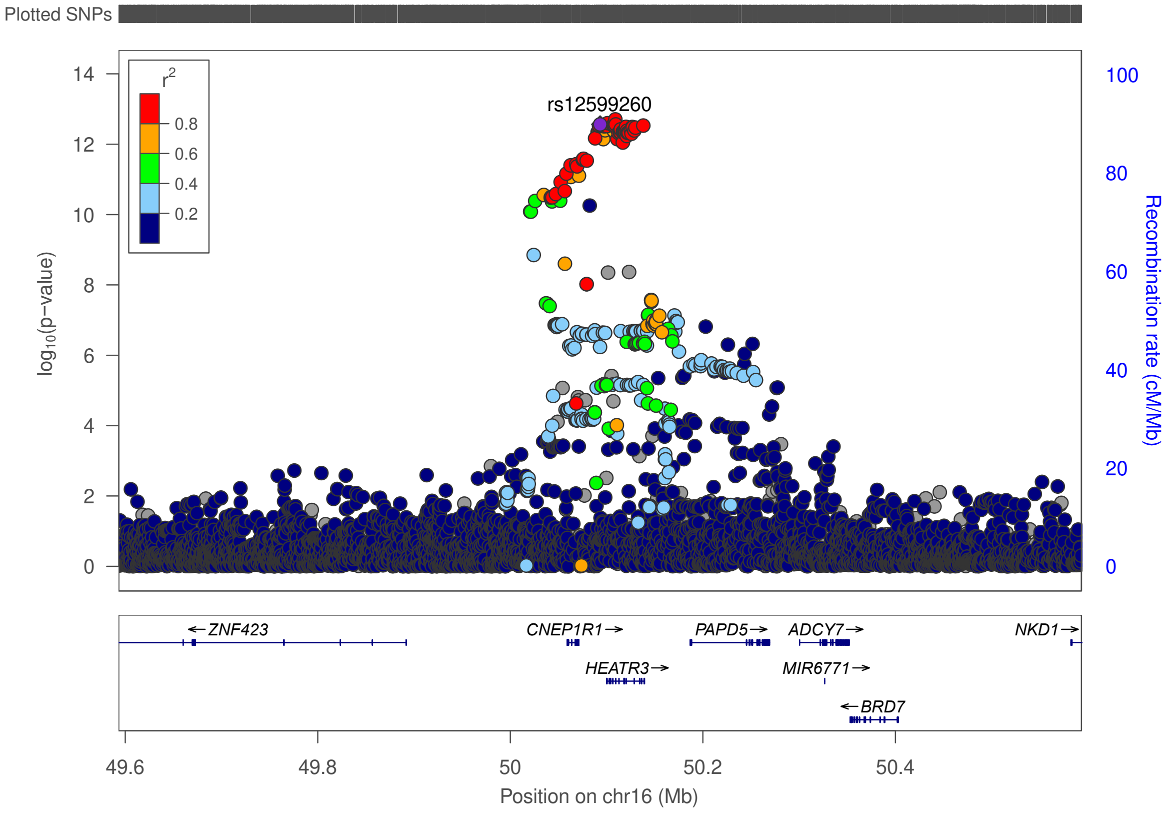

**Figure S25. Regional association plot of the novel UL association in chr16 near *HEATR3*.** The plot on top shows the result in META-1, and the plot on bottom shows the result in META-2.

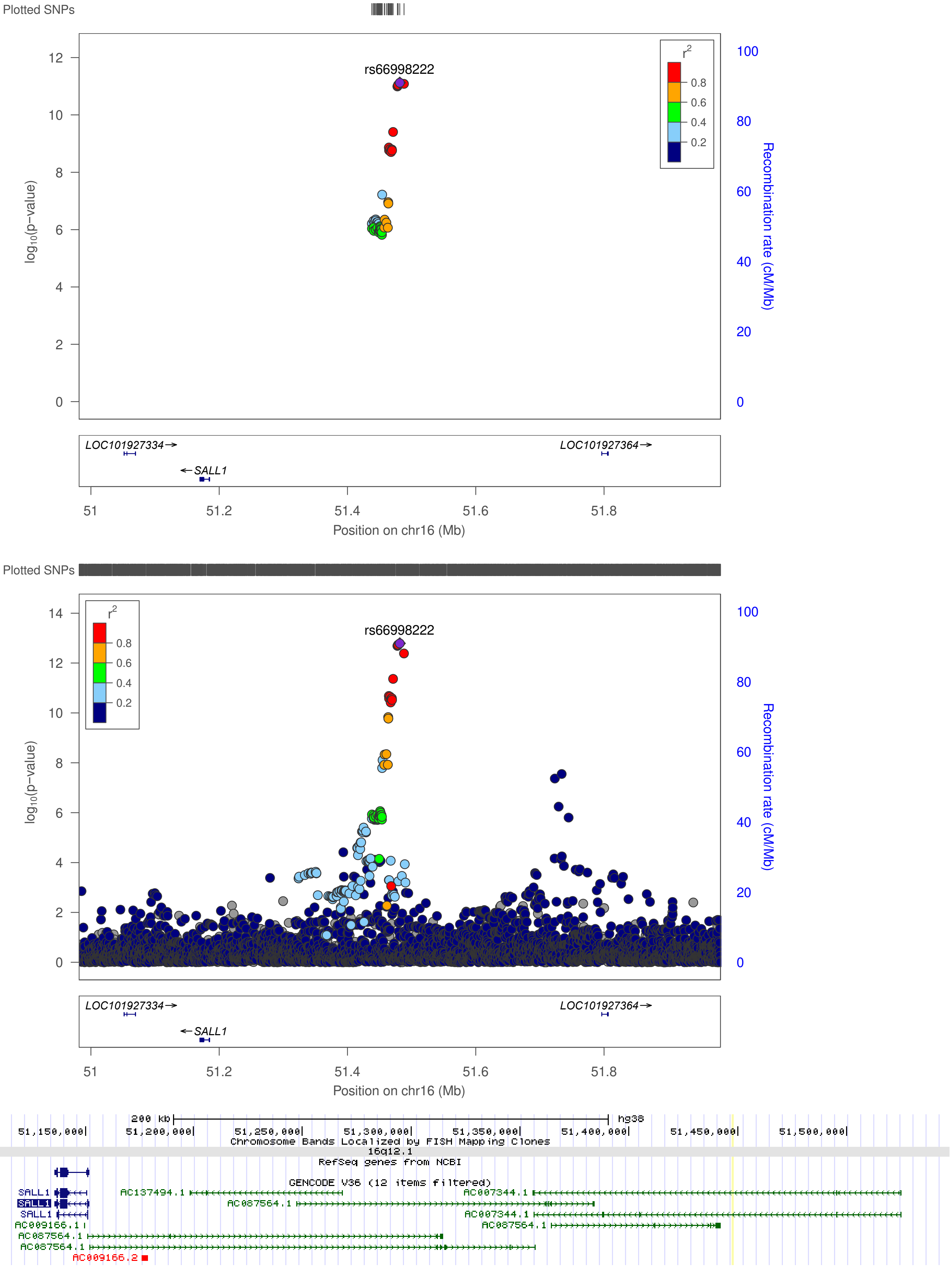

**Figure S26. Regional association plot of the novel UL association in chr16 near *AC007344.1*.**
The plot on top shows the result in META-1, and the plot on bottom shows the result in META-2. *AC007344.1* location was not available for LocusZoom: it locates ~200kb downstream from *SALL1*, as indicated in the image extracted from UCSC Genome Browser (bottom).

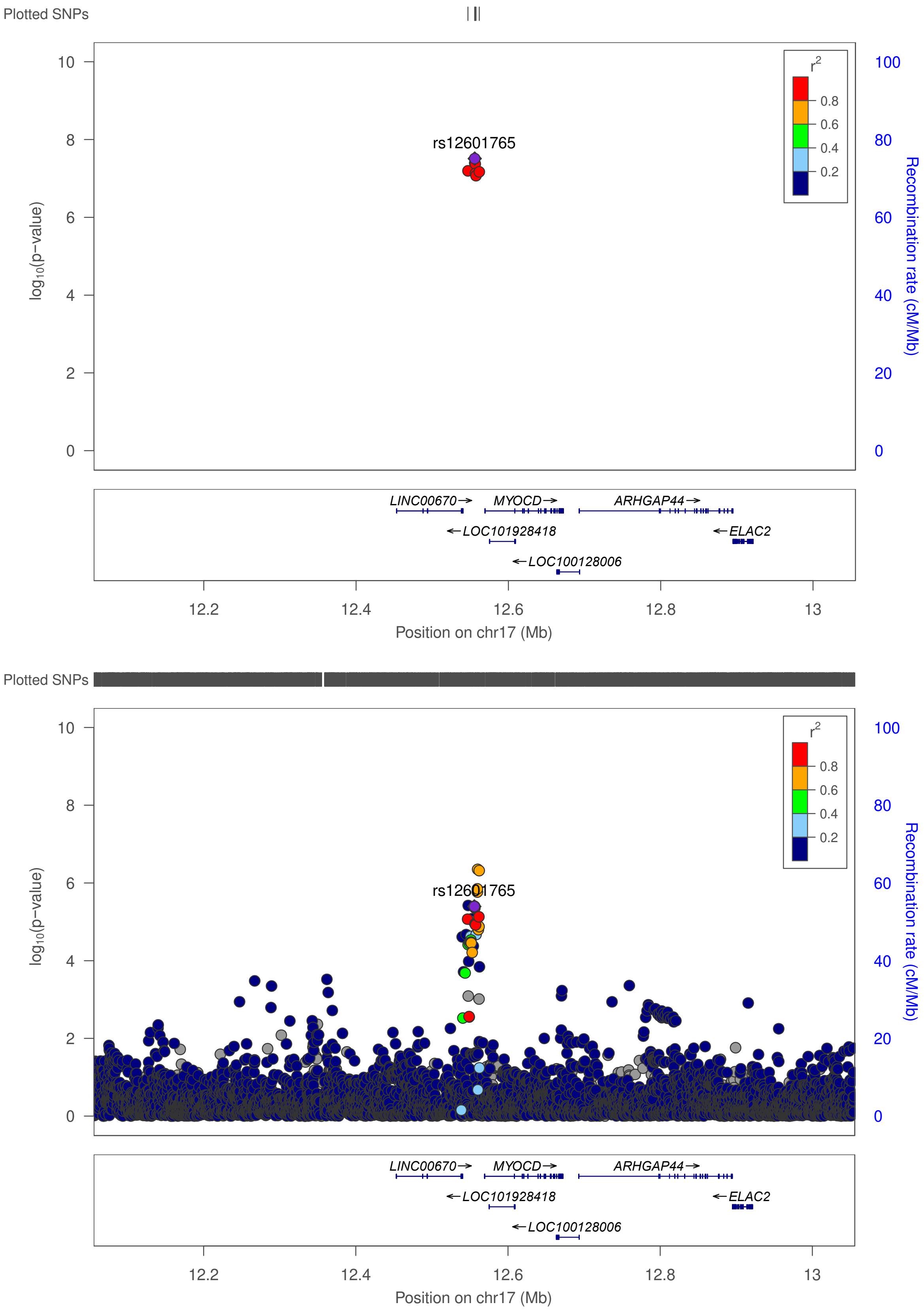

**Figure S27. Regional association plot of the novel UL association in chr17 near *MYOCD*.** The plot on top shows the result in META-1, and the plot on bottom shows the result in META-2.

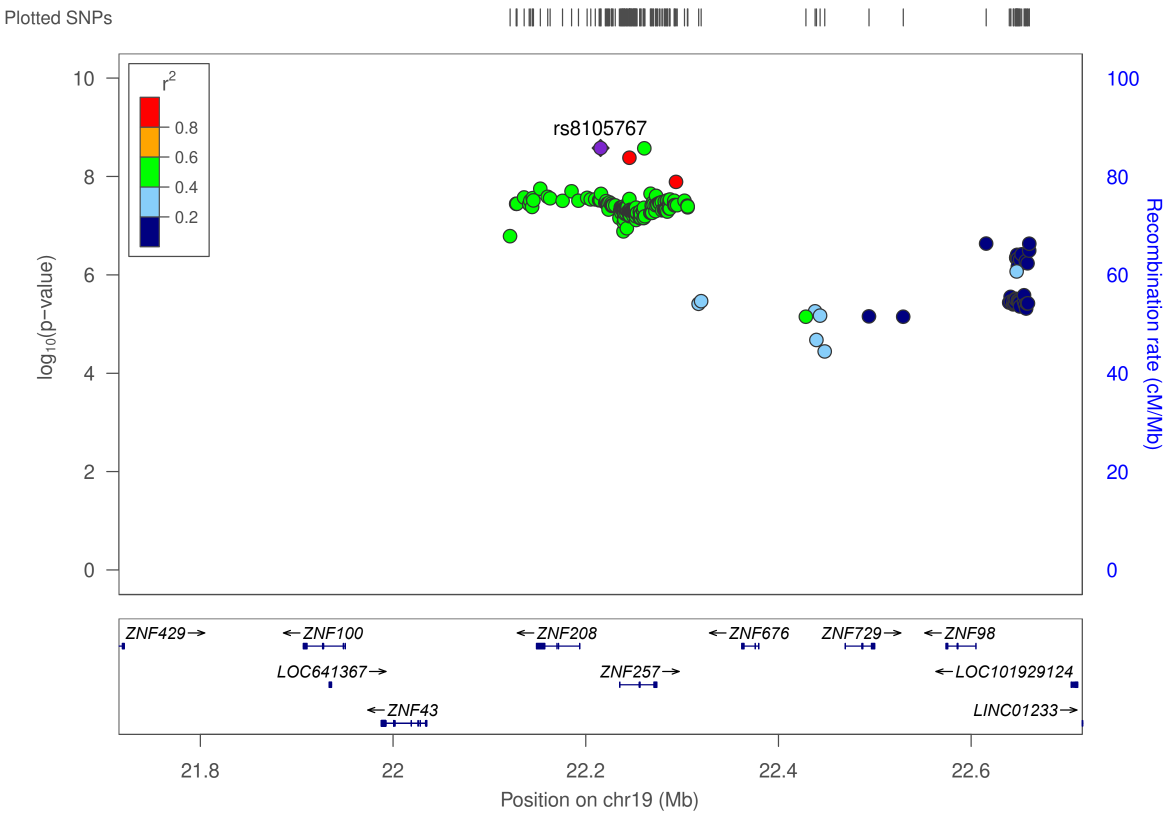

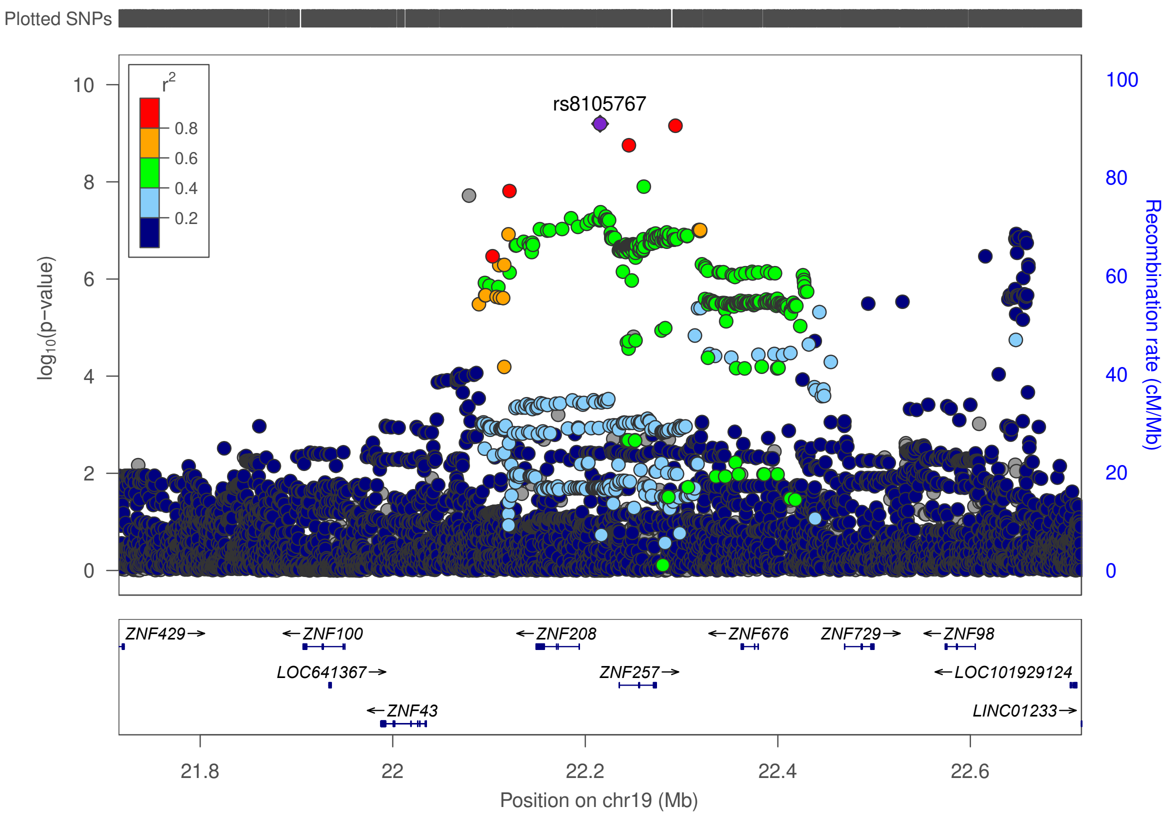

**Figure S28. Regional association plot of the novel UL association in chr19 near *ZNF257*.** The plot on top shows the result in META-1, and the plot on bottom shows the result in META-2.

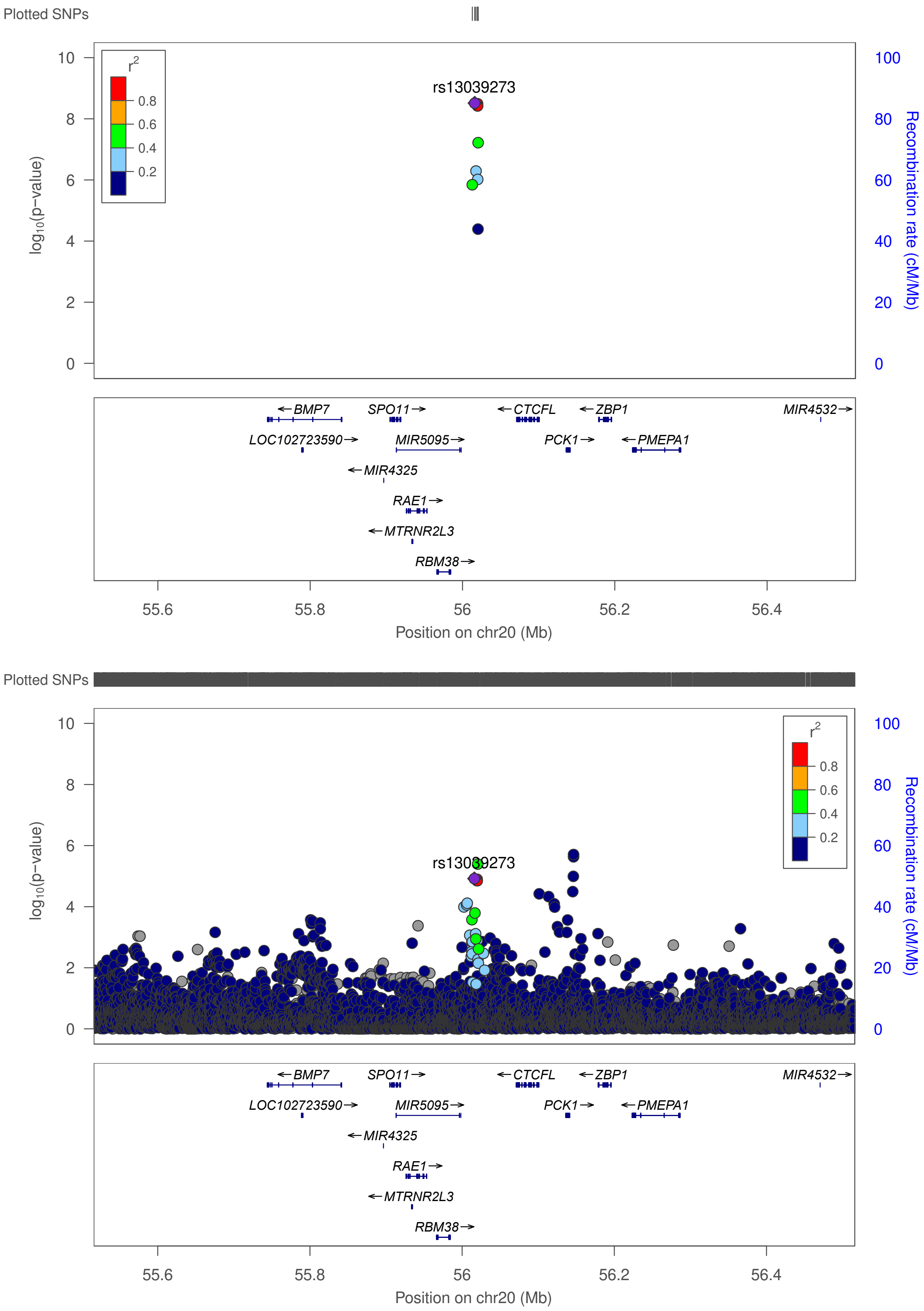

**Figure S29. Regional association plot of the novel UL association in chr20 near *CTCFL*.**
The plot on top shows the result in META-1, and the plot on bottom shows the result in META-2.

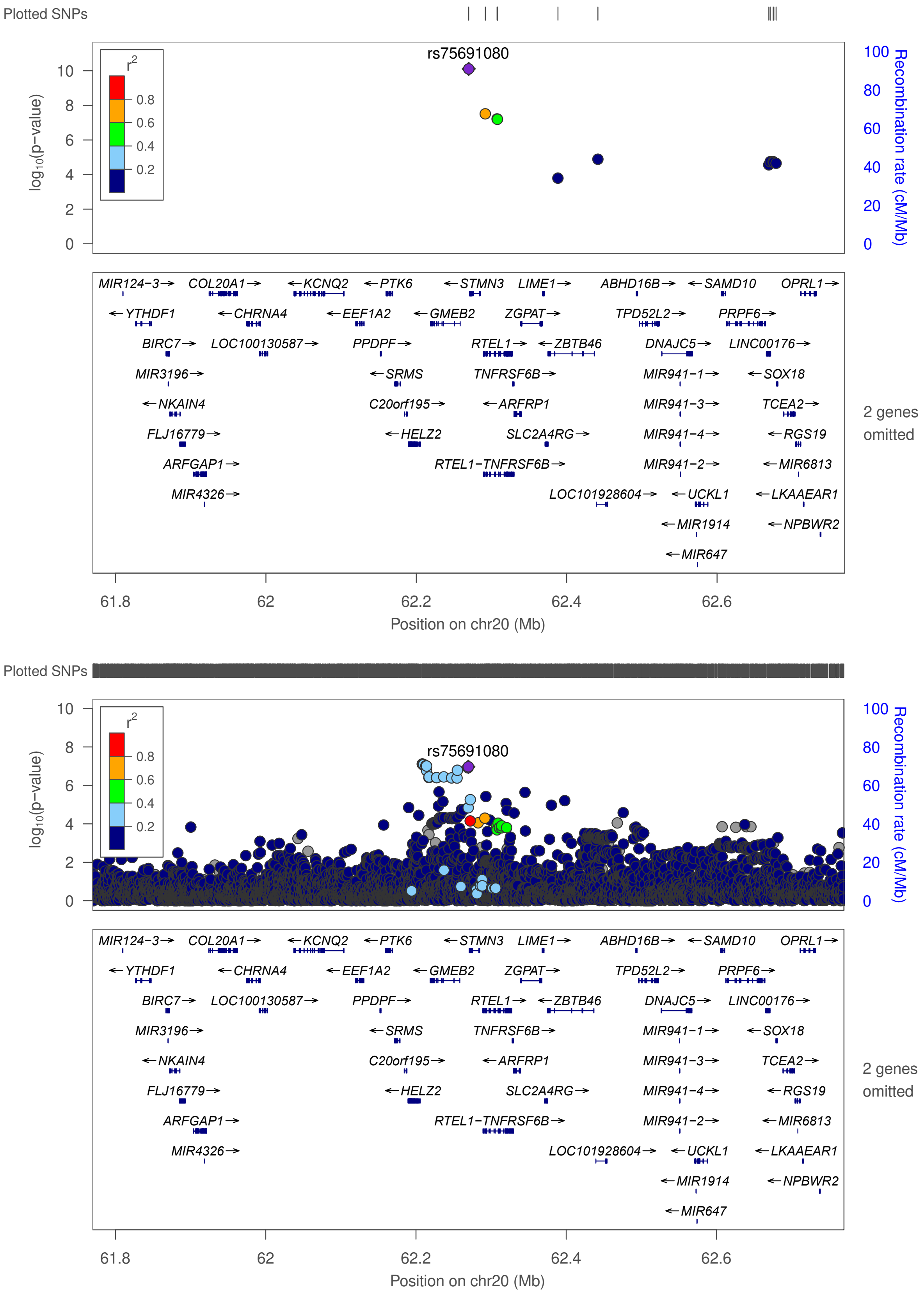

**Figure S30. Regional association plot of the novel UL association in chr20 near *STMN3*.** The plot on top shows the result in META-1, and the plot on bottom shows the result in META-2.

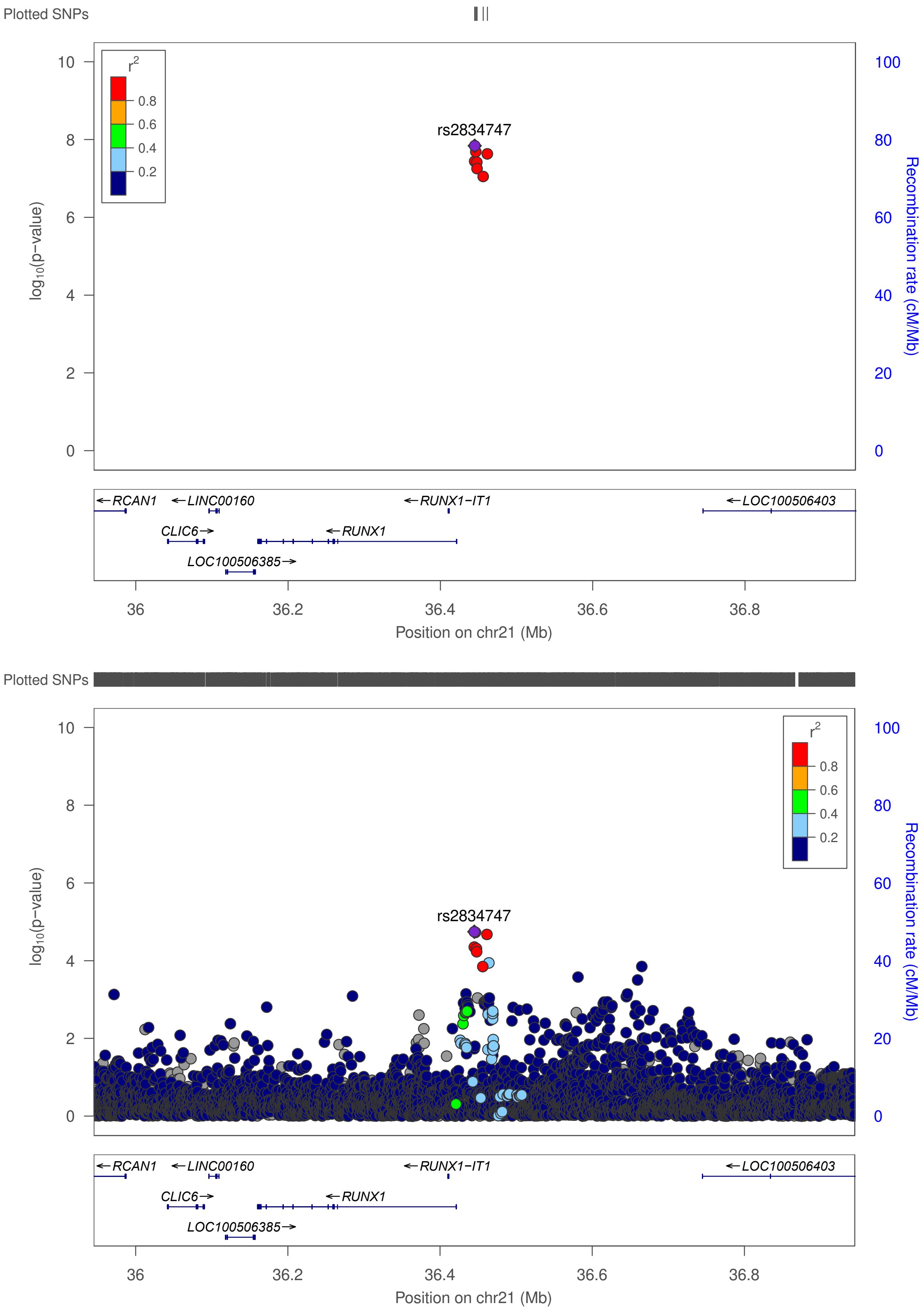

**Figure S31. Regional association plot of the novel UL association in chr21 near *RUNX1*.**
The plot on top shows the result in META-1, and the plot on bottom shows the result in META-2.

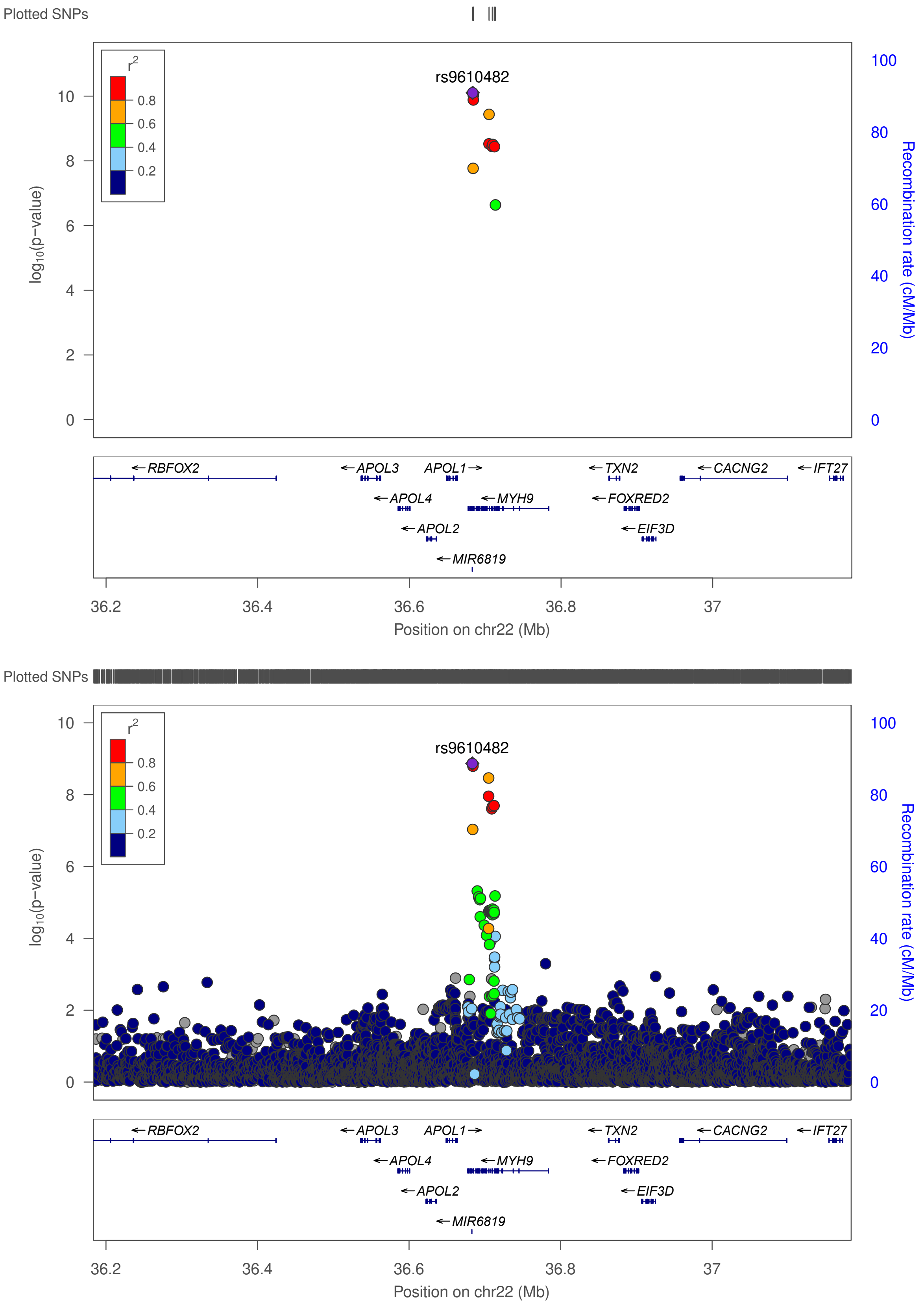

**Figure S32. Regional association plot of the novel UL association in chr22 near *MYH9*.** The plot on top shows the result in META-1, and the plot on bottom shows the result in META-2.

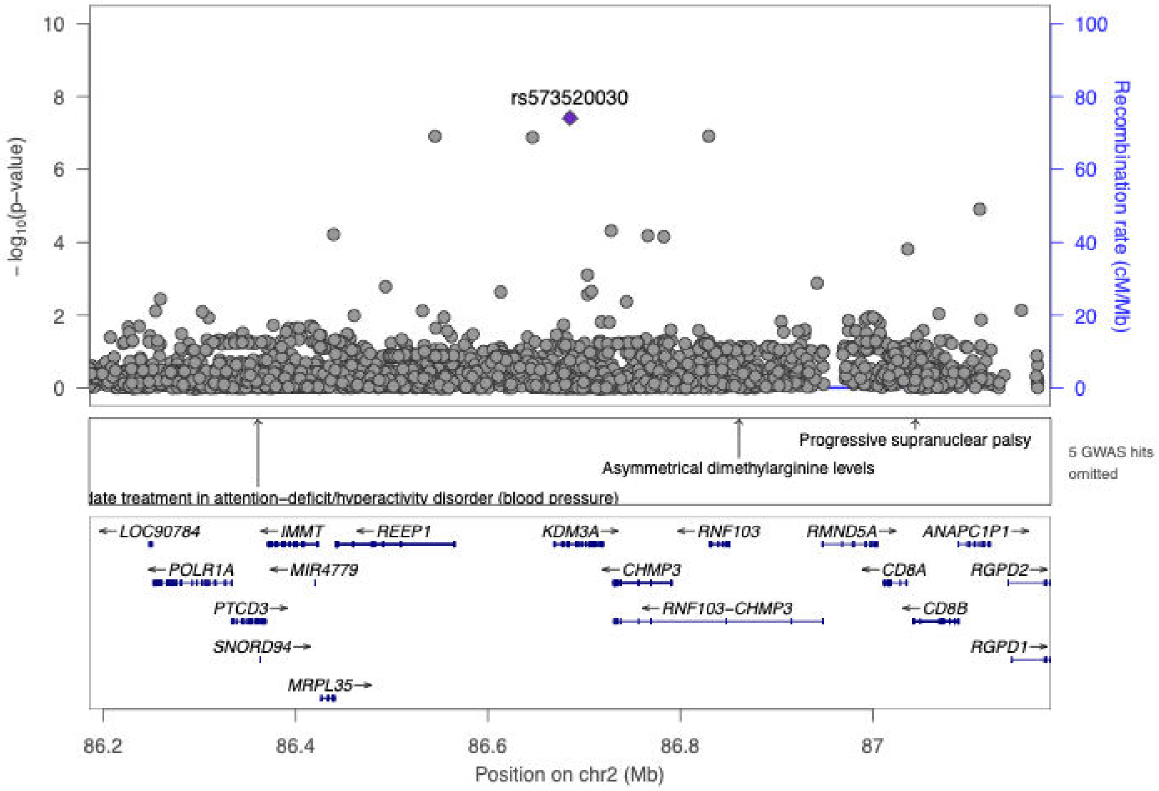

**Figure S33. Regional association plot of the novel UL association in chr2 near *KDM3A* [META-2].**

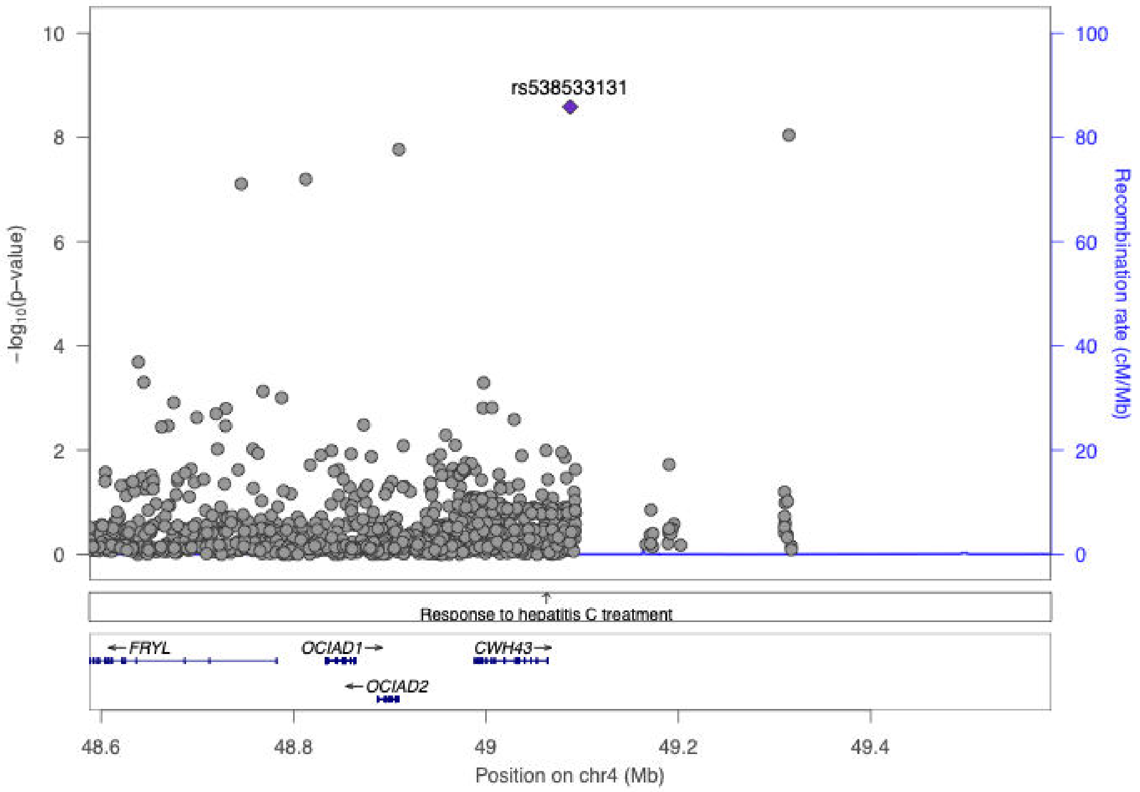

**Figure S34. Regional association plot of the novel UL association in chr4 near *CWH43* [META-2].**

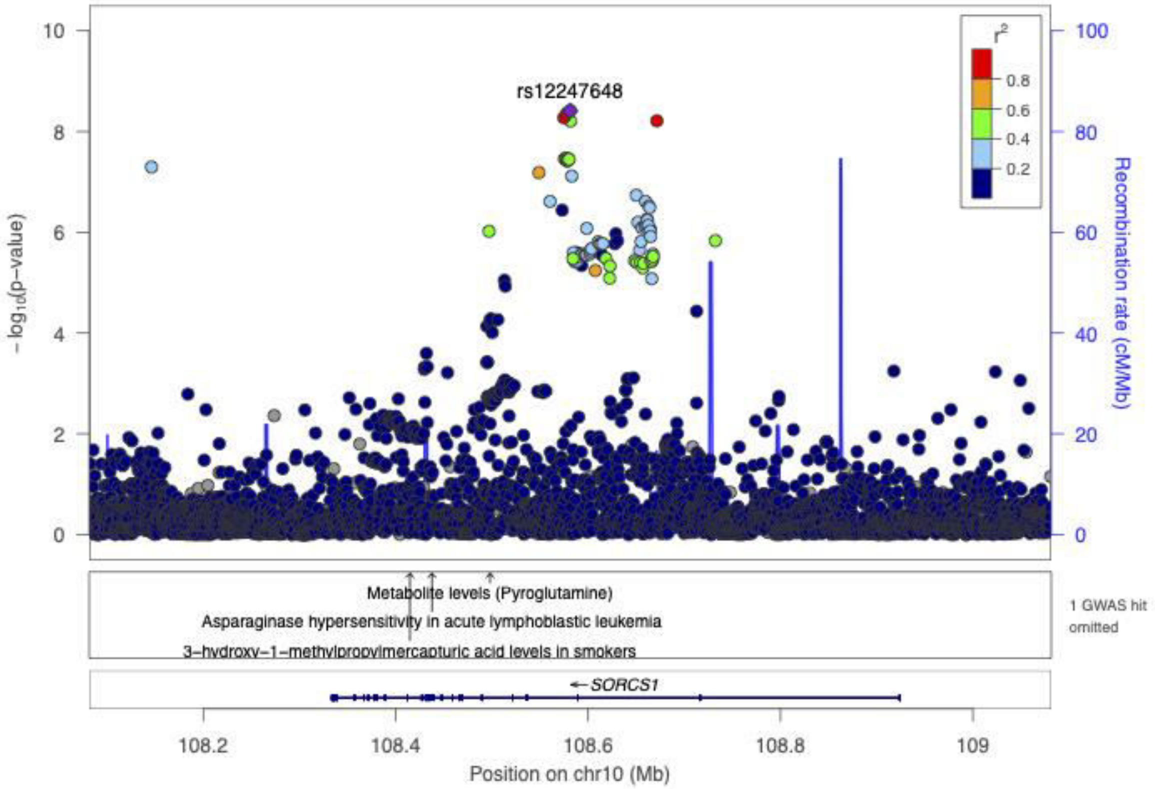

**Figure S35. Regional association plot of the novel UL association in chr10 near *SORCS1* [META-2].**

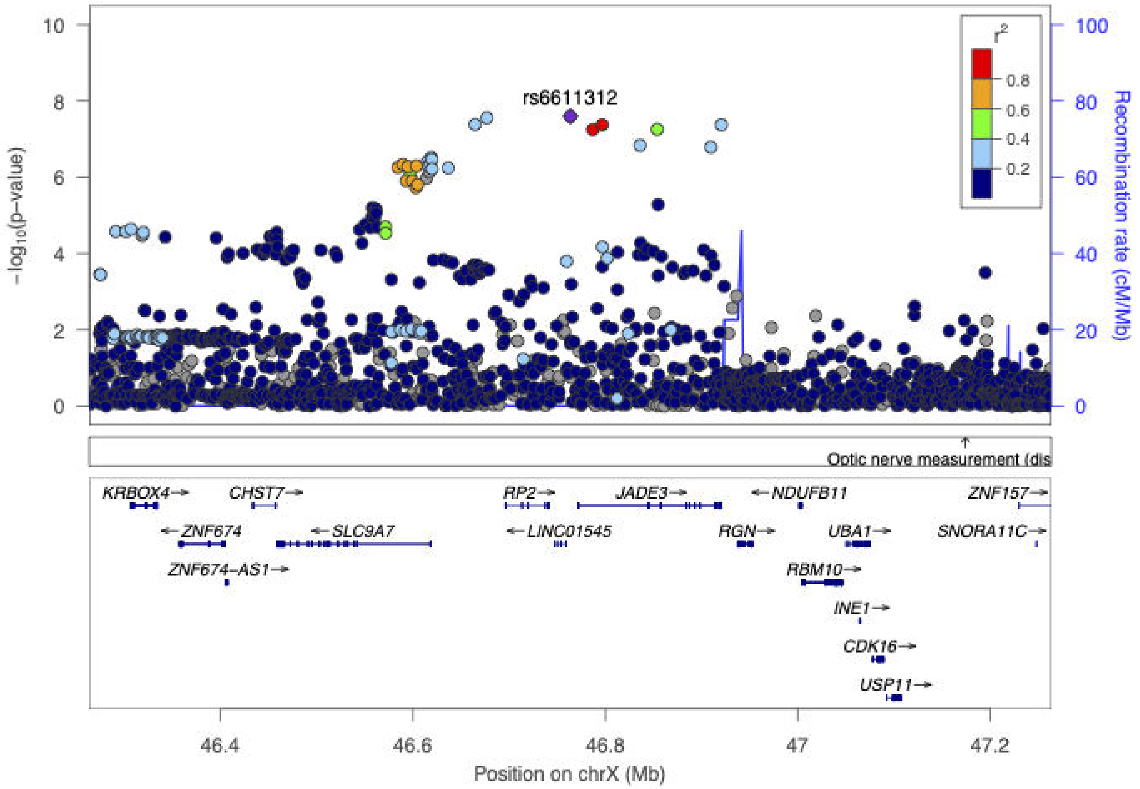

**Figure S36. Regional association plot of the novel UL association in chrX near *JADE3* [META-2].**

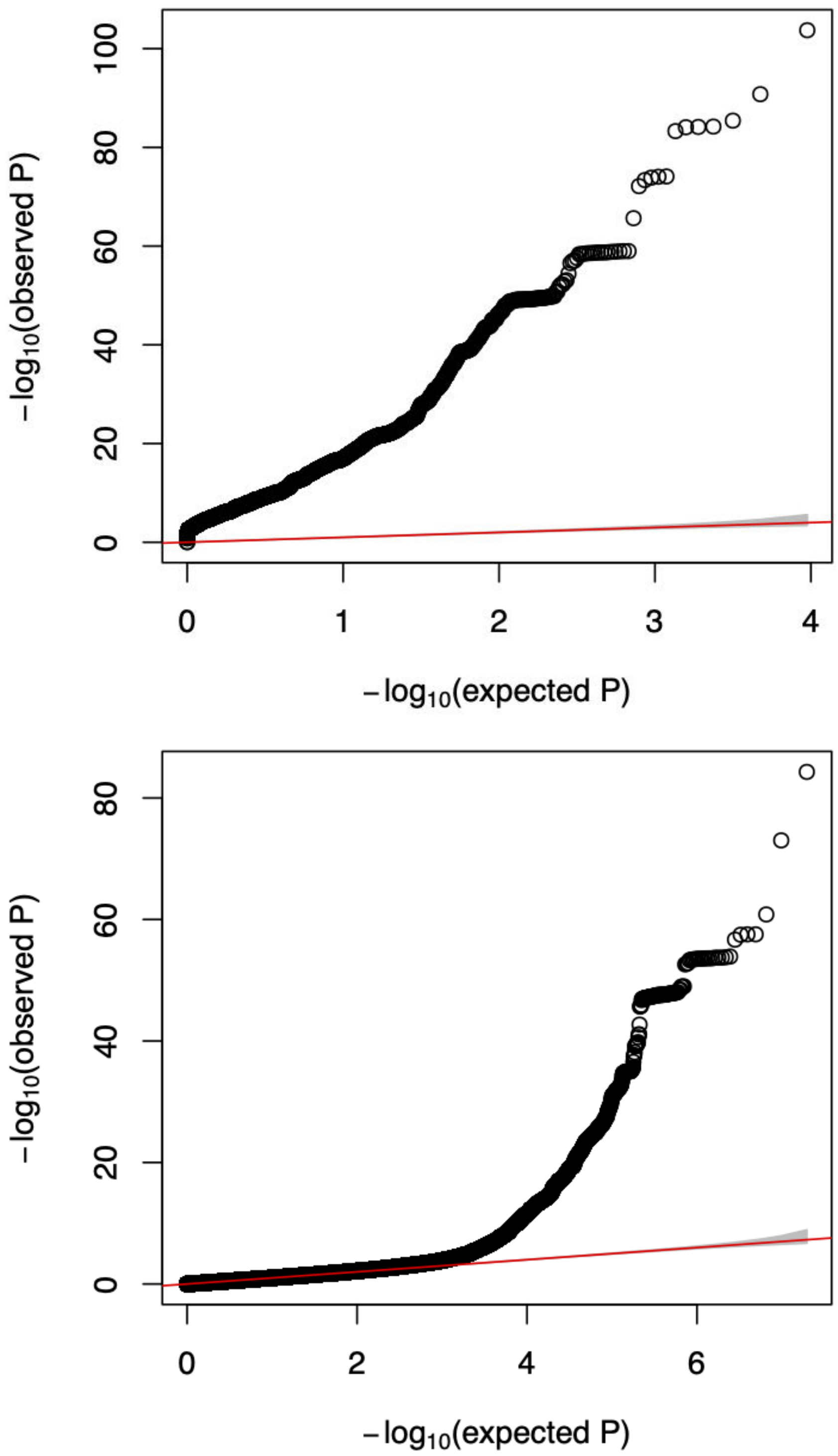

**Figure S37. Quantile-quantile plots of the p-values in META-1 (upper) and META-2 (lower).**
In META-1, the results of UL GWAS completed in FinnGen were meta-analysed data from a prior UL meta-analysis[1]; here, meta-analysis was limited to the top 10,000 variants the previous publication resulting in an atypical distribution of p-values compared with standard genome-wide analyses. In META-2, genome-wide results obtained in FinnGen results were meta-analysed with genome-wide data from the UK Biobank. Genomic inflation factor obtained using LDSC software[2] and genome-wide association results from META-2 was 1.07 suggested no inflation in the test statistics.

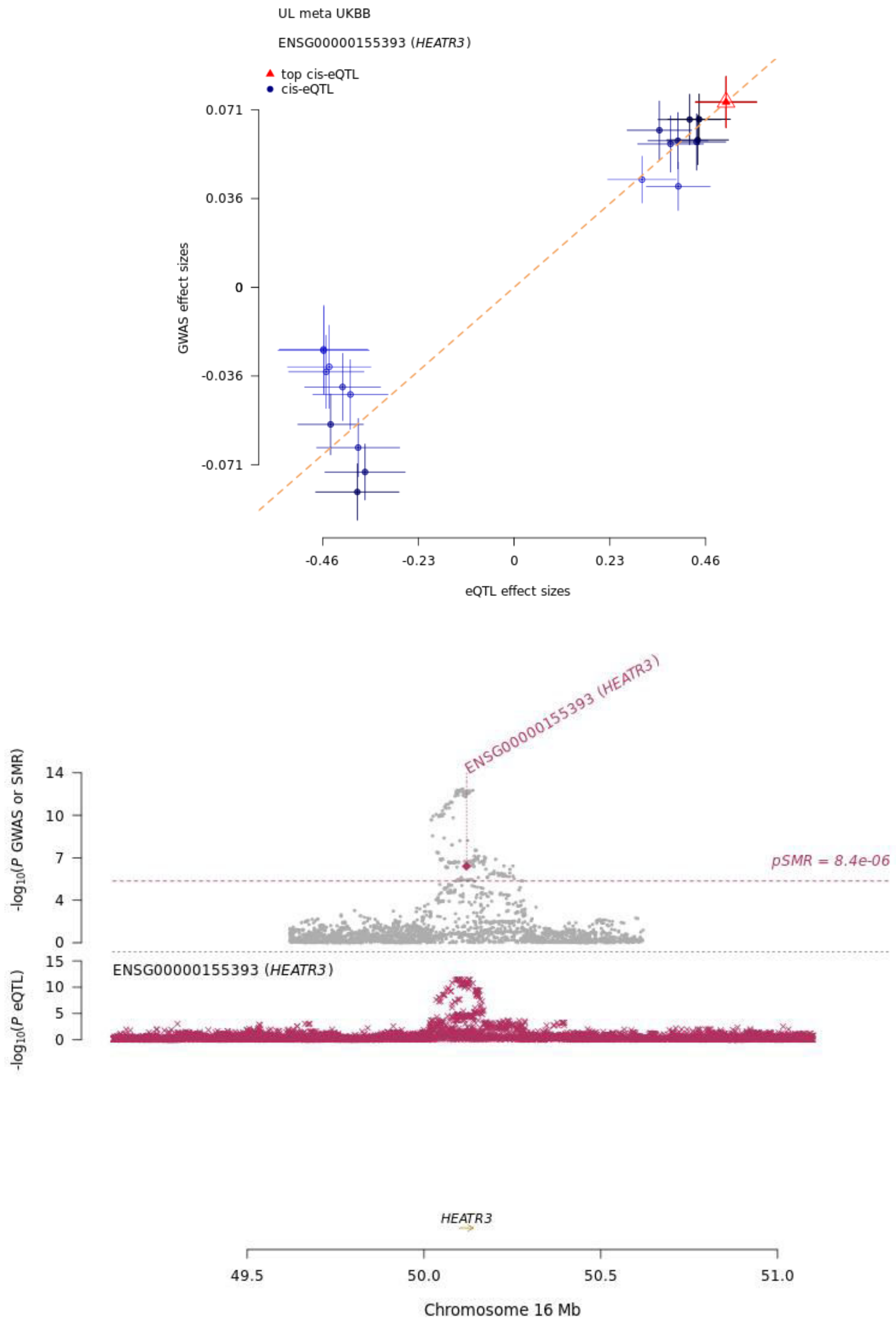

**Figure S38. SMR result: *HEATR3* at 16q12.1 in uterus.**
We tested if altered gene expression mediates UL risk associations using a method proposed by Zhu *et al.*[3] as implemented in CTG-VL[4]. Gray dots represent the p-values from the UL GWAS (META-2; meta-analysis of results obtained using FinnGen and UKBB data), pink diamond represents the p-value for the SMR test, and pink crosses represent the p-values from eQTL analysis for *HEATR3* (data from GTEx V7). The pink dashed line represents the threshold for statistical significance after correction for 5,967 gene-expression probes[3] ($p$=0.05/5,967=8.4x$10^{-6}$).

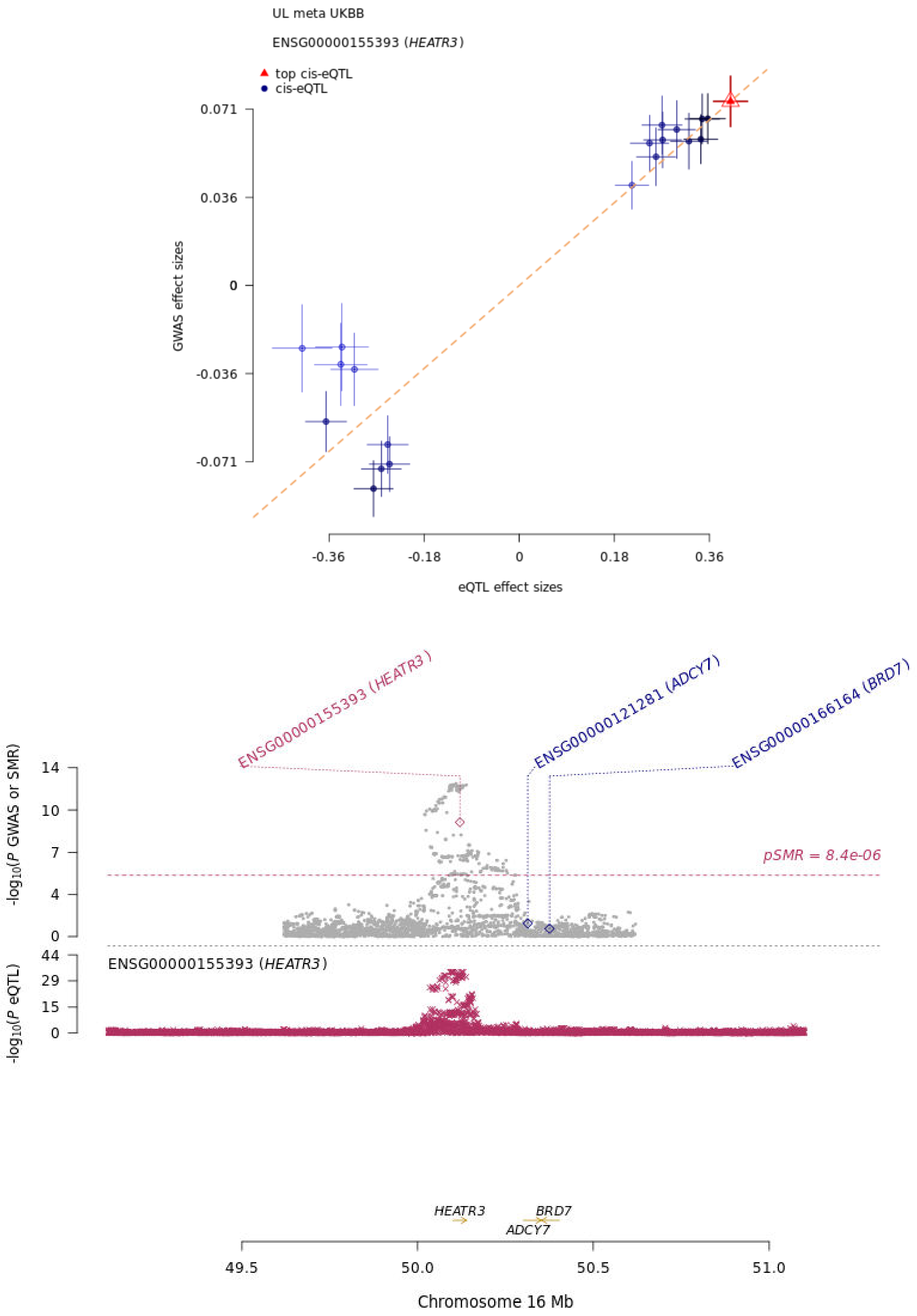

**Figure S39. SMR result: *HEATR3* at 16q12.1 in whole blood.**
We tested if altered gene expression mediates UL risk associations using a method proposed by Zhu *et al*.[3] as implemented in CTG-VL[4]. Gray dots represent the p-values from the UL GWAS (META-2; meta-analysis of results obtained using FinnGen and UKBB data), pink diamond represents the p-value for the SMR test, and pink crosses represent the p-values from eQTL analysis for *HEATR3* (data from GTEx V7). The pink dashed line represents the threshold for statistical significance after correction for 5,967 gene-expression probes[3] ($p$=0.05/5,967=8.4x10$^{-6}$).

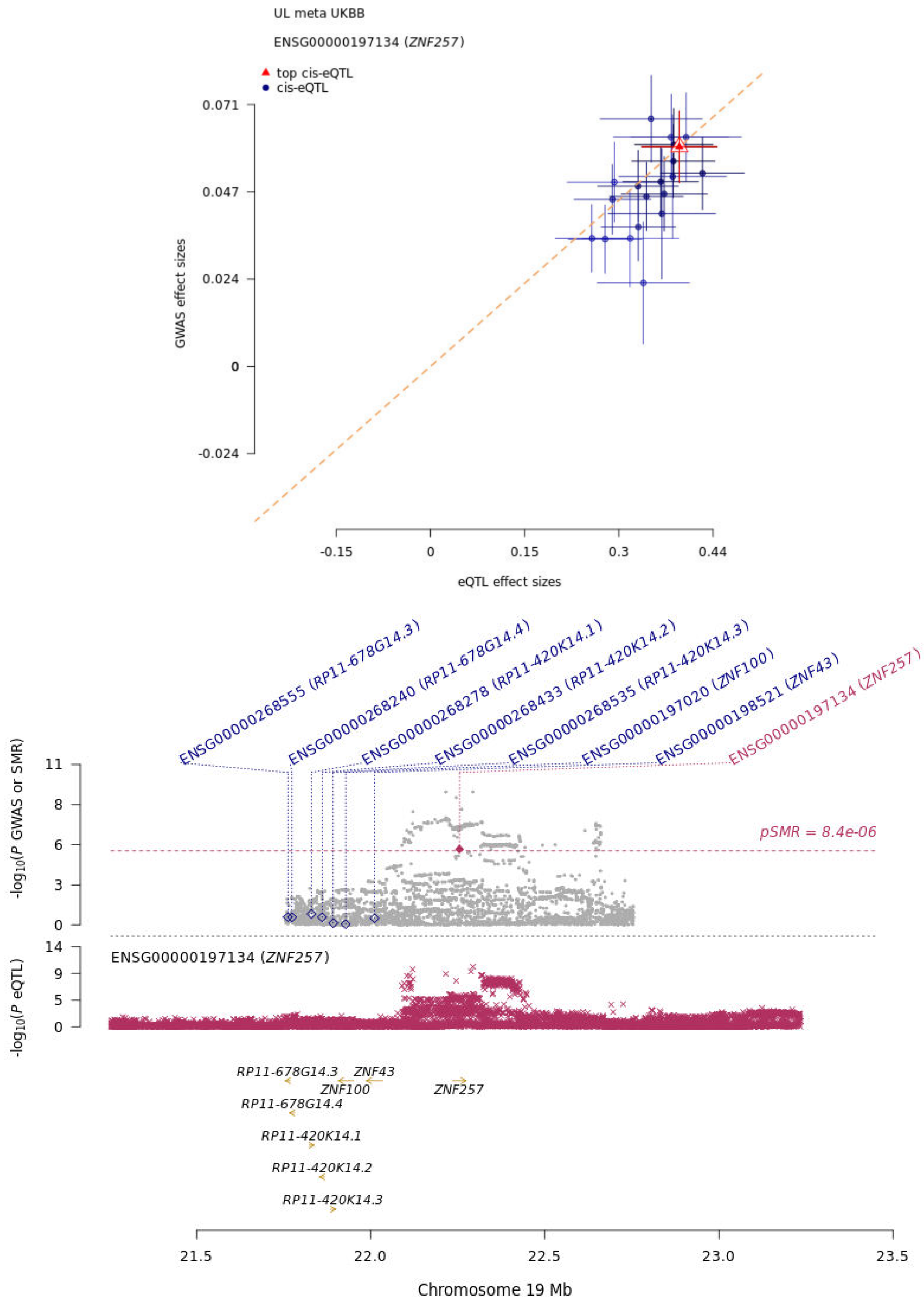

**Figure S40. SMR result: *ZNF257* at 19q12 in whole blood.**
We tested if altered gene expression mediates UL risk associations using a method proposed by Zhu *et al.*[3] as implemented in CTG-VL[4]. Gray dots represent the p-values from the UL GWAS (META-2; meta-analysis of results obtained using FinnGen and UKBB data), pink diamond represents the p-value for the SMR test, and pink crosses represent the p-values from eQTL analysis for *ZNF257* (data from GTEx V7). The pink dashed line represents the threshold for statistical significance after correction for 5,967 gene-expression probes[3] ($p$=0.05/5,967=8.4x$10^{-6}$).

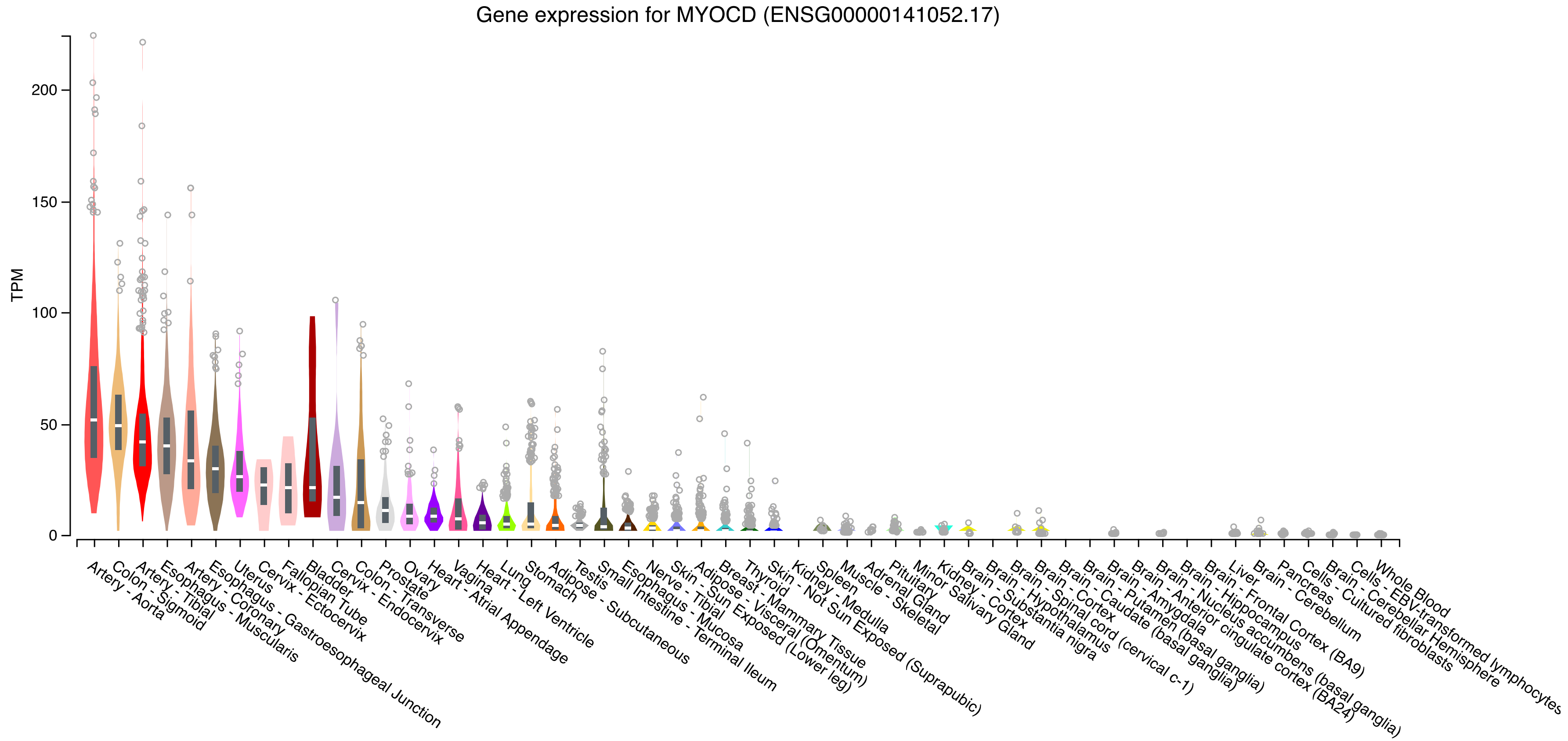

**Figure S41. *MYOCD* expression in tissues.**
The data was extracted from the GTEx Portal[5] on 02/10/2021.

**Figure S42. The effect of rs13039273 on *RBM38* expression.**
'Samples' indicate the number of RNA-seq samples with genotype. Normalized effect size ('NES') is the slope of the linear regression of the normalized data versus the three genotype categories using single-tissue expression quantitative trait (eQTL) analysis, representing eQTL effect size. 'P-value' originates from a t-test that compares observed NES from single-tissue eQTL analysis to a null NES of 0 ($p$=1.38x$10^{-19}$ for the meta-analyzed effects). 'M-value' indicates the posterior probability that an eQTL effect exists in each tissue tested in the cross-tissue meta-analysis. NES are given for the C allele that was the effect allele in the present study.

| Tissue | Samples | NES | p-value | m-value |
|---|---|---|---|---|
| Kidney - Cortex | 73 | 0.184 | 0.1 | 0.00 |
| Artery - Coronary | 213 | 0.174 | 3.7e-3 | 0.00 |
| Artery - Aorta | 387 | 0.101 | 0.01 | 0.00 |
| Adrenal Gland | 233 | 0.0778 | 0.2 | 0.00 |
| Breast - Mammary Tissue | 396 | 0.0751 | 0.03 | 0.00 |
| Brain - Anterior cingulate cortex (BA24) | 147 | 0.0730 | 0.1 | 0.00 |
| Brain - Spinal cord (cervical c-1) | 126 | 0.0681 | 0.4 | 0.00 |
| Spleen | 227 | 0.0600 | 0.3 | 0.00 |
| Adipose - Visceral (Omentum) | 469 | 0.0595 | 0.1 | 0.00 |
| Brain - Hypothalamus | 170 | 0.0519 | 0.3 | 0.00 |
| Brain - Substantia nigra | 114 | 0.0423 | 0.5 | 0.00 |
| Muscle - Skeletal | 706 | 0.0376 | 0.09 | 0.00 |
| Nerve - Tibial | 532 | 0.0364 | 0.3 | 0.00 |
| Brain - Hippocampus | 165 | 0.0304 | 0.6 | 0.00 |
| Artery - Tibial | 584 | 0.0277 | 0.3 | 0.00 |
| Liver | 208 | 0.0239 | 0.6 | 0.00 |
| Minor Salivary Gland | 144 | 0.0231 | 0.8 | 0.00900 |
| Adipose - Subcutaneous | 581 | 0.0228 | 0.5 | 0.00 |
| Whole Blood | 670 | 0.0227 | 0.2 | 0.00 |
| Heart - Left Ventricle | 386 | 0.0203 | 0.6 | 0.00 |
| Brain - Cortex | 205 | 0.0151 | 0.7 | 0.00 |
| Testis | 322 | 0.0120 | 0.6 | 0.00 |
| Stomach | 324 | 0.00944 | 0.8 | 0.00 |
| Brain - Cerebellum | 209 | 0.00199 | 1 | 0.00300 |
| Brain - Caudate (basal ganglia) | 194 | 0.000202 | 1 | 0.00 |
| Brain - Frontal Cortex (BA9) | 175 | 0.000182 | 1 | 0.00 |
| Skin - Not Sun Exposed (Suprapubic) | 517 | -0.00388 | 0.9 | 0.00 |
| Skin - Sun Exposed (Lower leg) | 605 | -0.0108 | 0.7 | 0.00 |
| Brain - Putamen (basal ganglia) | 170 | -0.0131 | 0.7 | 0.00 |
| Vagina | 141 | -0.0153 | 0.8 | 0.00 |
| Pituitary | 237 | -0.0259 | 0.6 | 0.00 |
| Colon - Sigmoid | 318 | -0.0261 | 0.3 | 0.00 |
| Colon - Transverse | 368 | -0.0334 | 0.1 | 0.00400 |
| Cells - Cultured fibroblasts | 483 | -0.0405 | 0.2 | 0.00700 |
| Brain - Amygdala | 129 | -0.0451 | 0.5 | 0.0120 |
| Small Intestine - Terminal Ileum | 174 | -0.0463 | 0.2 | 0.0260 |
| Esophagus - Mucosa | 497 | -0.0490 | 0.1 | 0.0250 |
| Prostate | 221 | -0.0507 | 0.2 | 0.0240 |
| Brain - Nucleus accumbens (basal ganglia) | 202 | -0.0598 | 0.1 | 0.0390 |
| Lung | 515 | -0.0600 | 0.05 | 0.0470 |
| Esophagus - Muscularis | 465 | -0.0660 | 6.9e-4 | 0.0830 |
| Thyroid | 574 | -0.0678 | 0.02 | 0.0350 |
| Esophagus - Gastroesophageal Junction | 330 | -0.0776 | 1.5e-3 | 0.0960 |
| Brain - Cerebellar Hemisphere | 175 | -0.0873 | 0.1 | 0.0450 |
| Cells - EBV-transformed lymphocytes | 147 | -0.132 | 0.2 | 0.258 |
| Heart - Atrial Appendage | 372 | -0.165 | 3.1e-7 | 1.00 |
| Ovary | 167 | -0.226 | 8.7e-6 | 1.00 |
| Uterus | 129 | -0.242 | 2.2e-3 | 0.908 |
| Pancreas | 305 | -0.586 | 2.8e-16 | 1.00 |

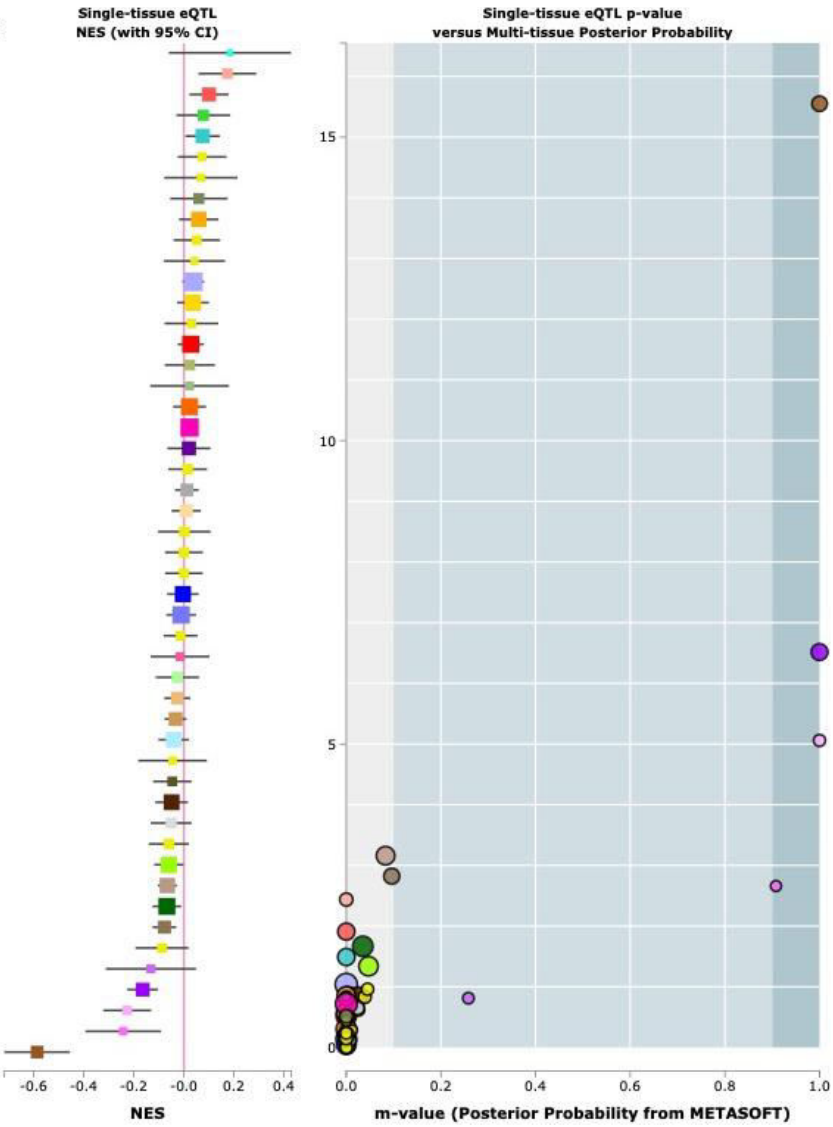

**Figure S43. MR scatter plot: UL & BMI.**
Scatter plot showing the relationship of SNP effects on BMI against SNP effects on UL risk (left) and vice versa (right).

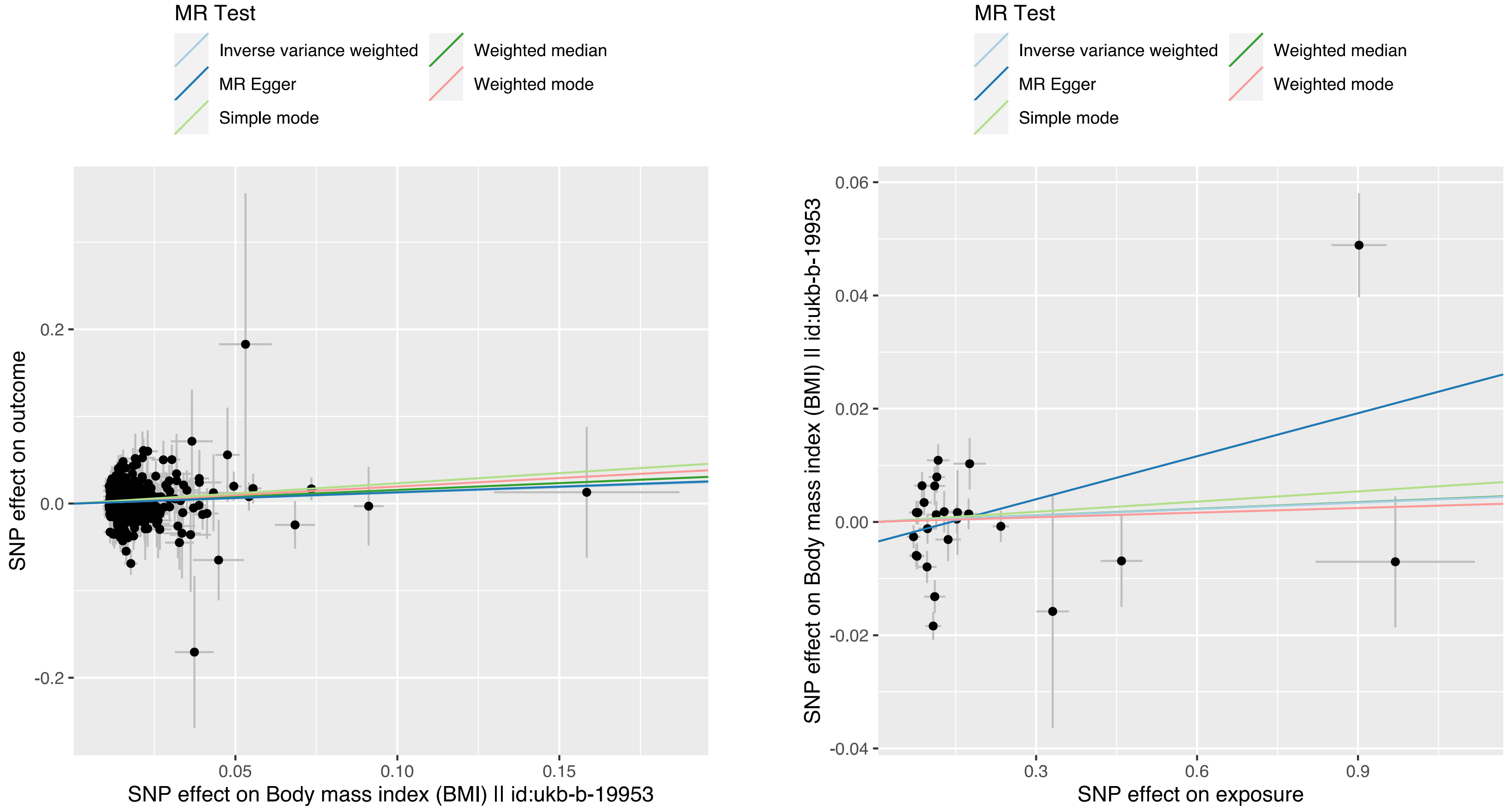

**Figure S44. MR scatter plot: UL & waist circumference.**
Scatter plot showing the relationship of SNP effects on waist circumference against SNP effects on UL risk (left) and vice versa (right).

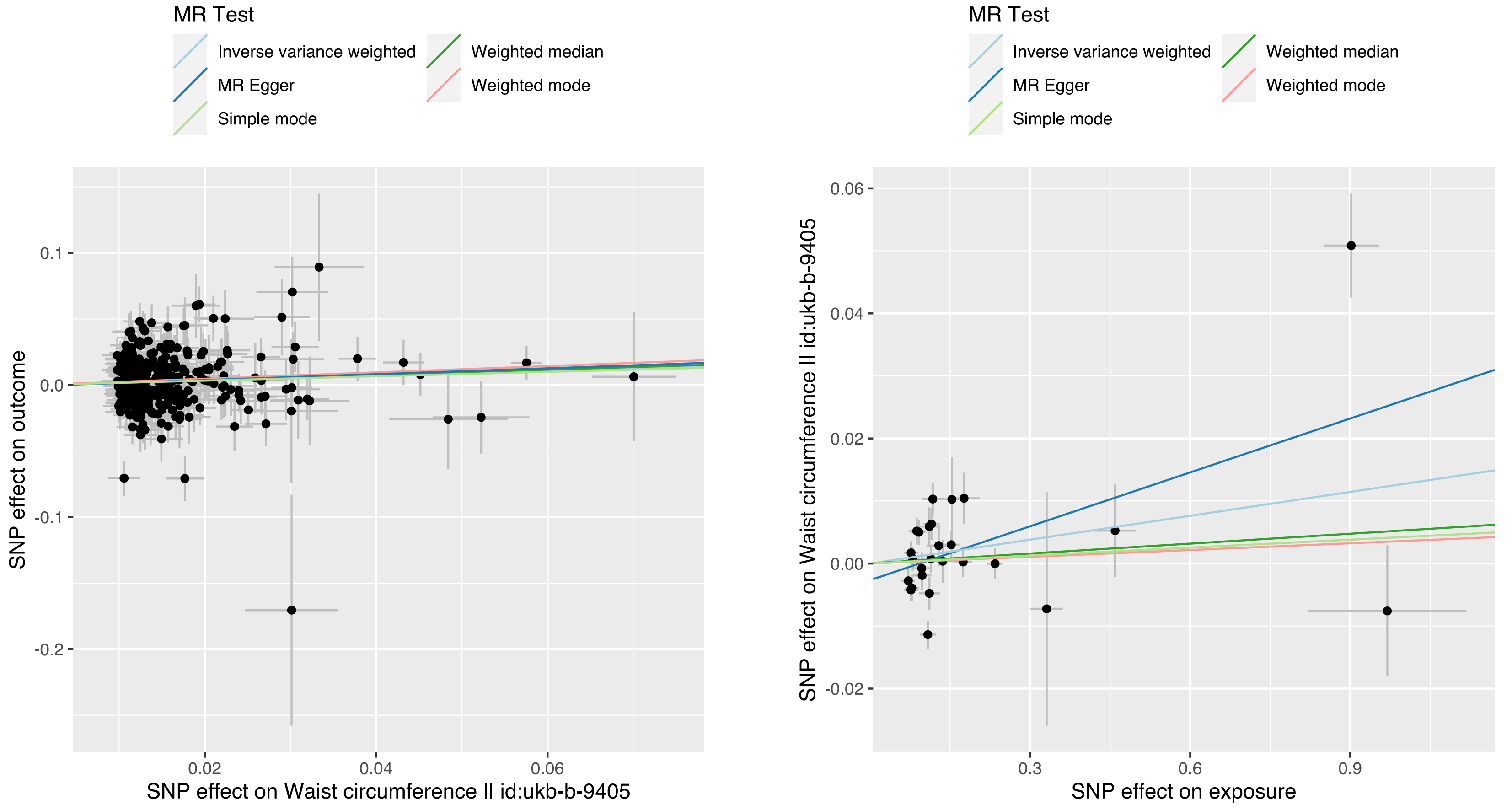

**Figure S45. MR scatter plot: UL & whole body fat mass.**
Scatter plot showing the relationship of SNP effects on whole body fat mass against SNP effects on UL risk (left) and vice versa (right).

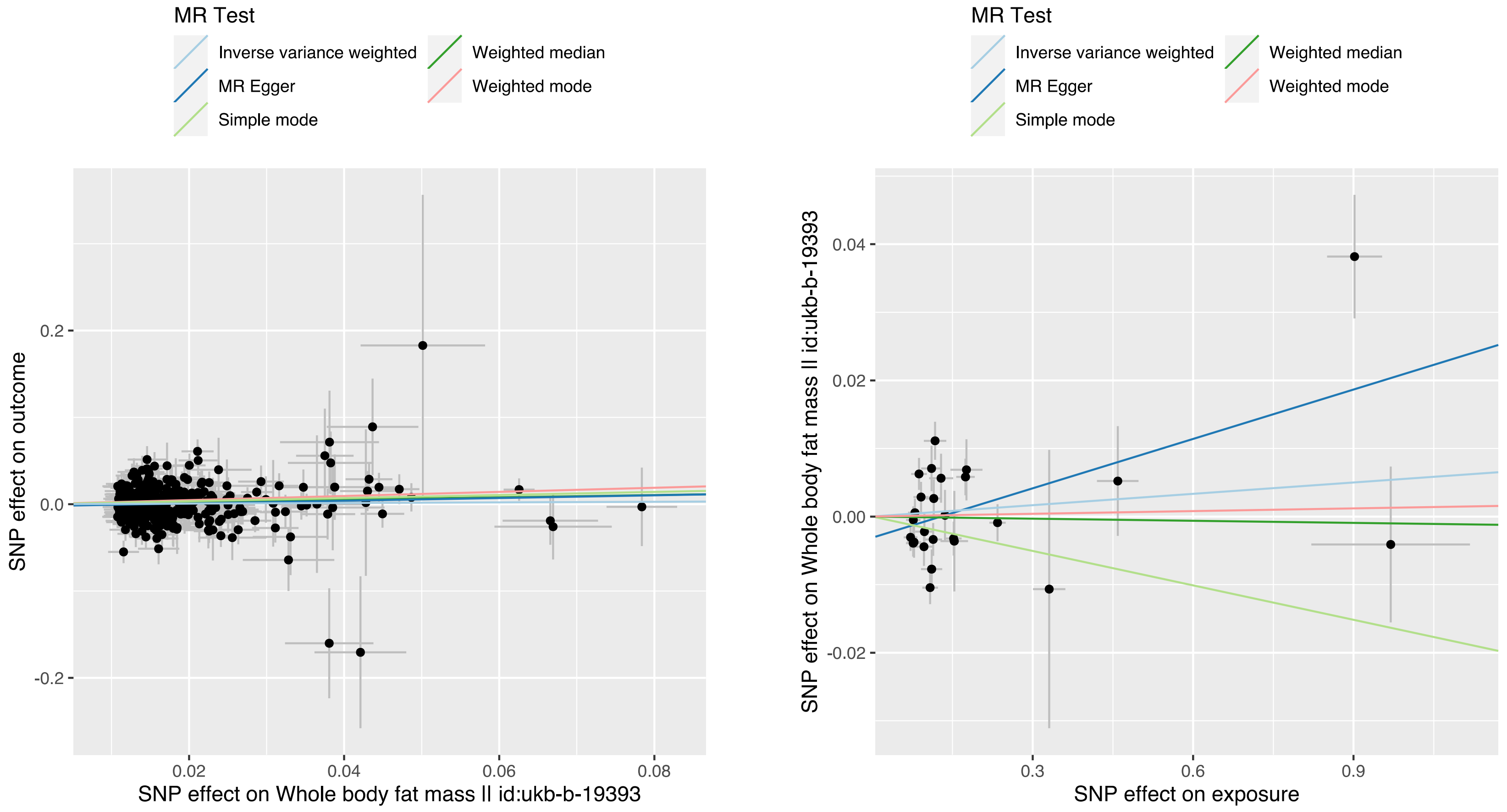

**Figure S46. MR scatter plot: UL & whole body fat-free mass.**
Scatter plot showing the relationship of SNP effects on whole body fat-free mass against SNP effects on UL risk (left) and vice versa (right).

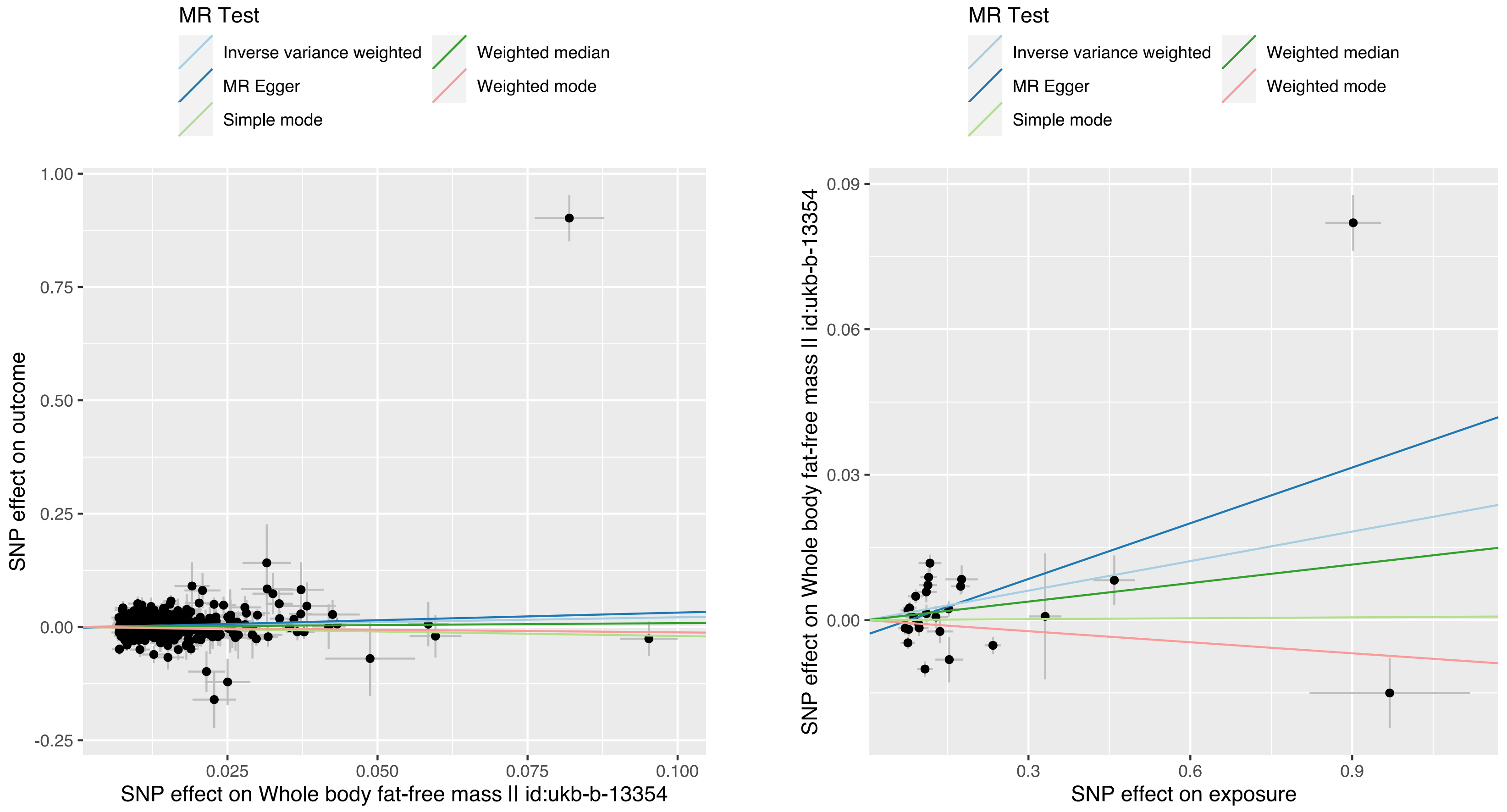

**Figure S47. MR scatter plot: UL & whole body water mass.**
Scatter plot showing the relationship of SNP effects on whole body water mass against SNP effects on UL risk (left) and vice versa (right).

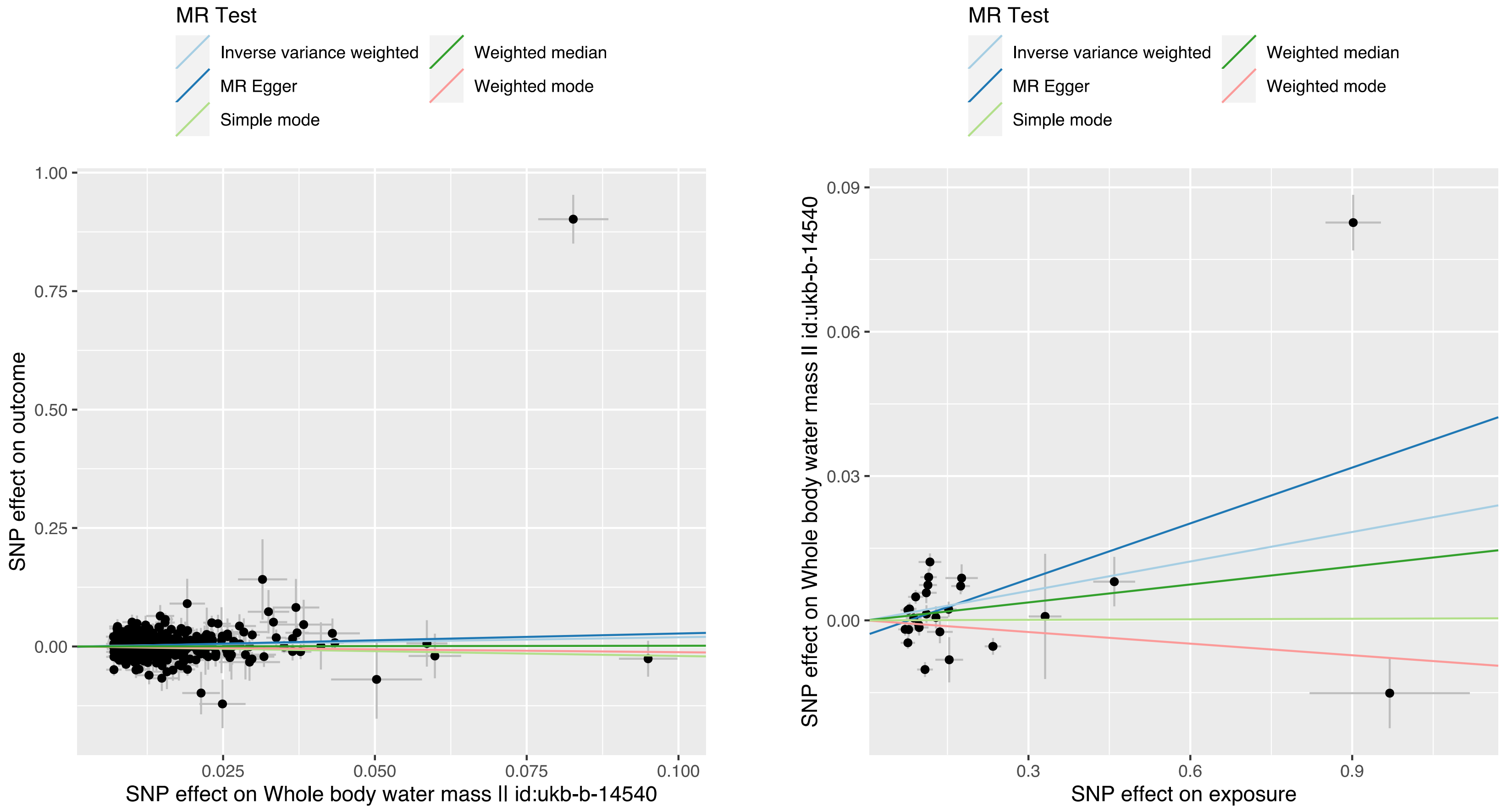

**Figure S48. MR scatter plot: UL & impedance of whole body.**
Scatter plot showing the relationship of SNP effects on impedance of whole body against SNP effects on UL risk (left) and vice versa (right).

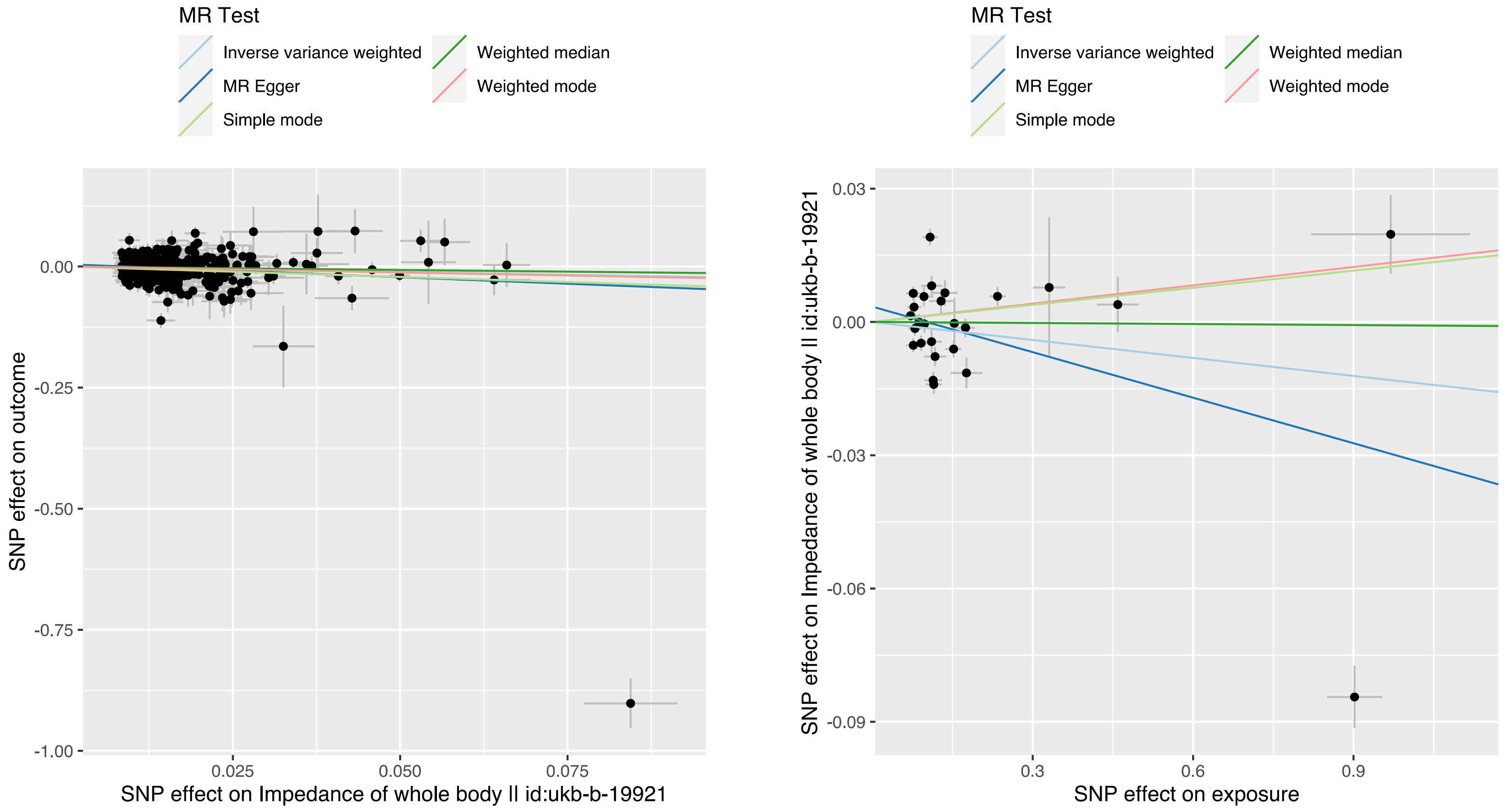

**Figure S49. MR scatter plot: UL & basal metabolic rate.**
Scatter plot showing the relationship of SNP effects on basal metabolic rate against SNP effects on UL risk (left) and vice versa (right).

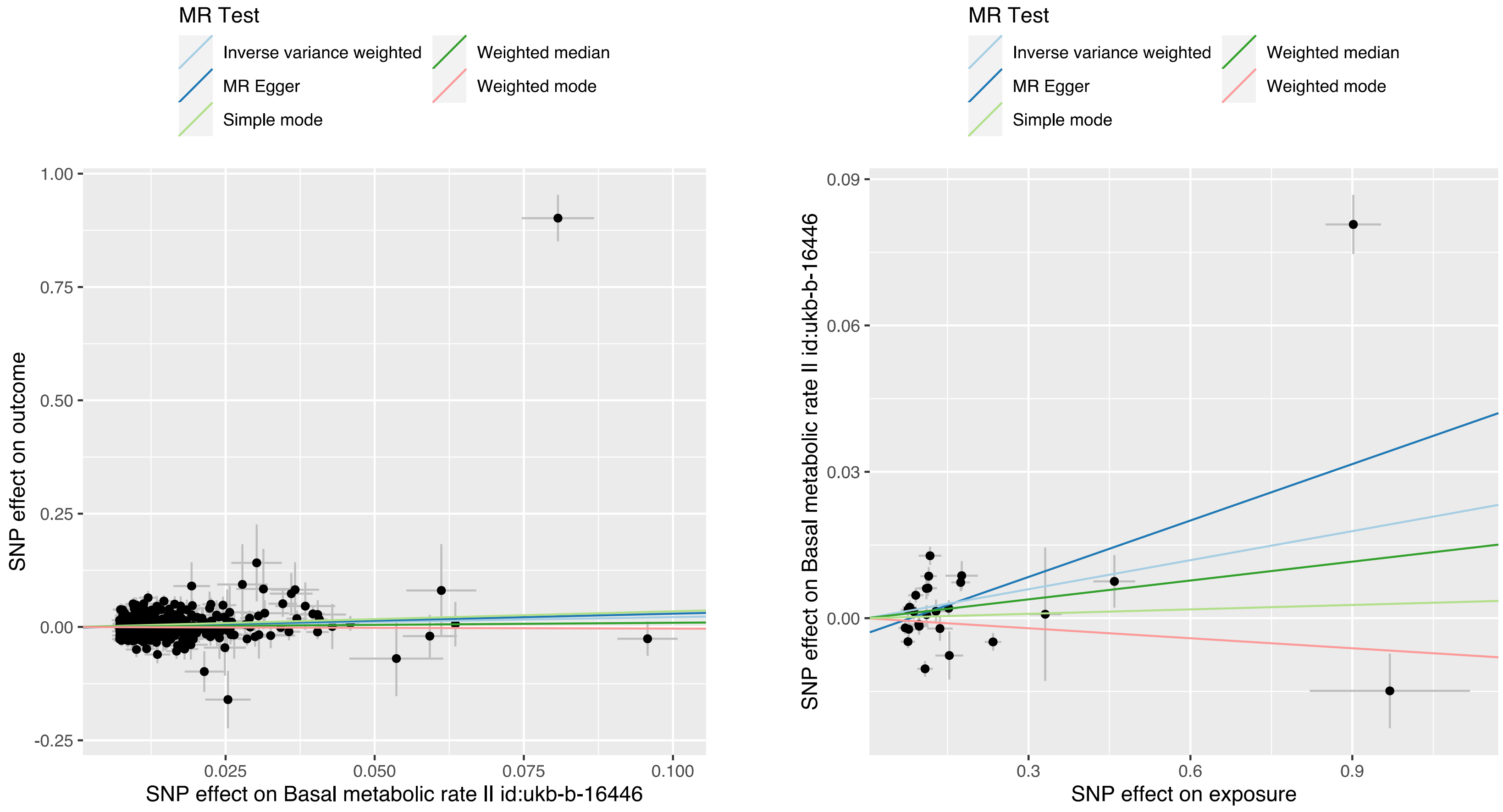

**Figure S50. Leave-one-out: exposure BMI, outcome UL.**

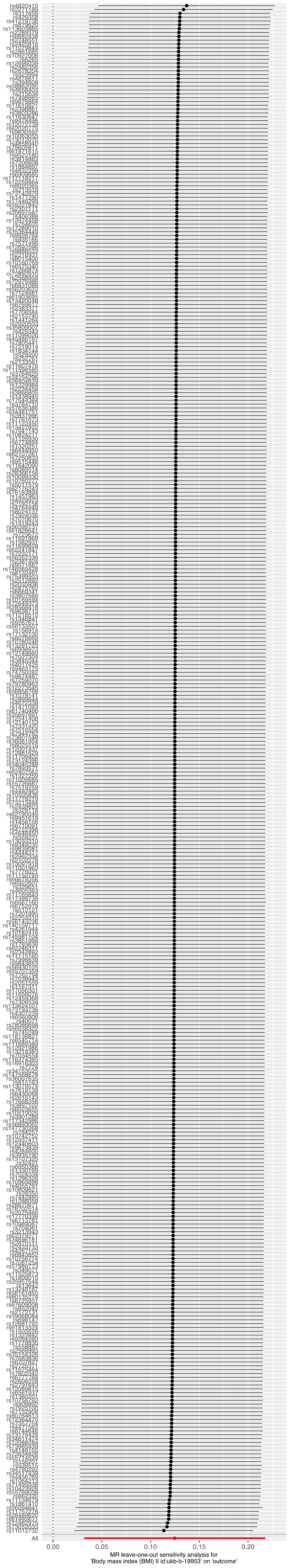

**Figure S51. Leave-one-out: exposure waist circumference, outcome UL.**

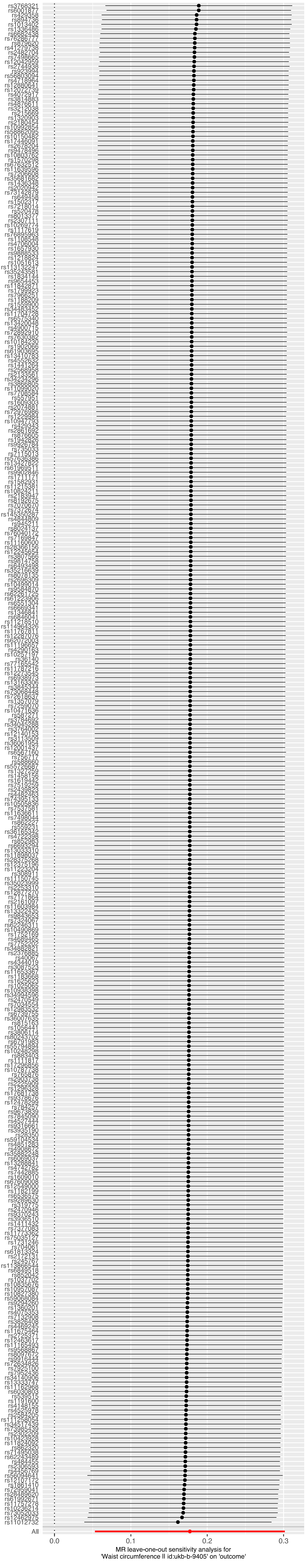

**Figure S52. Leave-one-out: exposure whole body fat mass, outcome UL.**

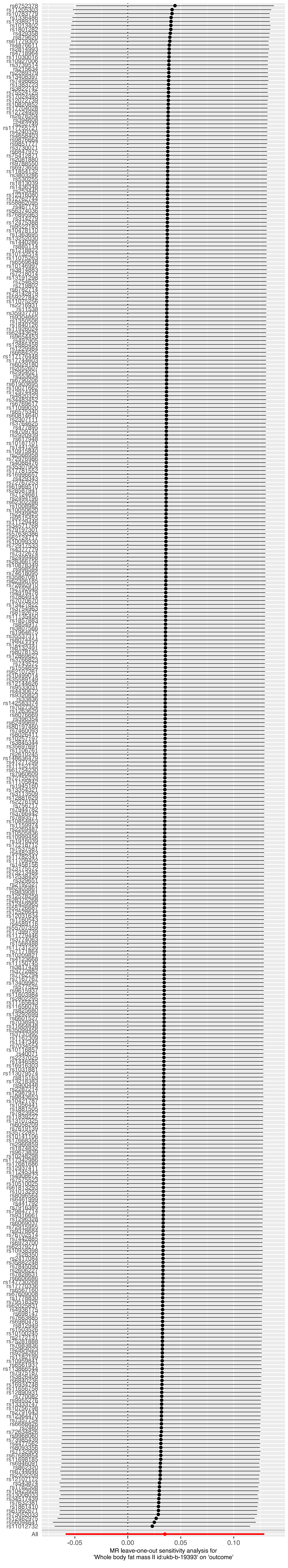

**Figure S53. Leave-one-out: exposure whole body fat-free mass, outcome UL.**

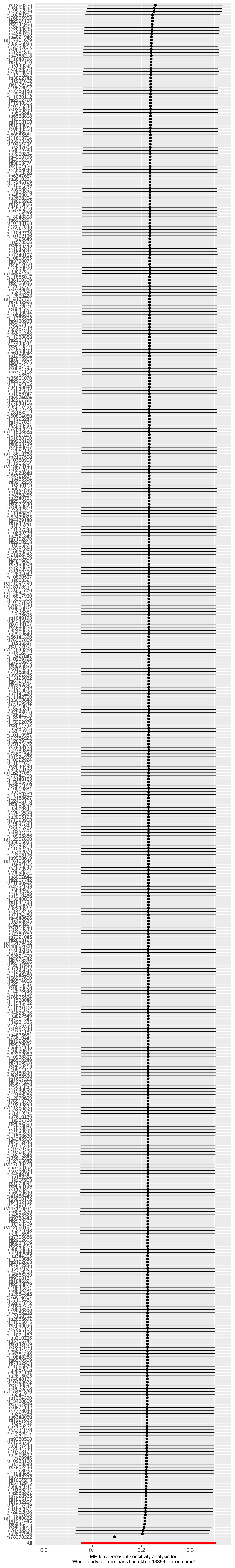

**Figure S54. Leave-one-out: exposure whole body water mass, outcome UL.**

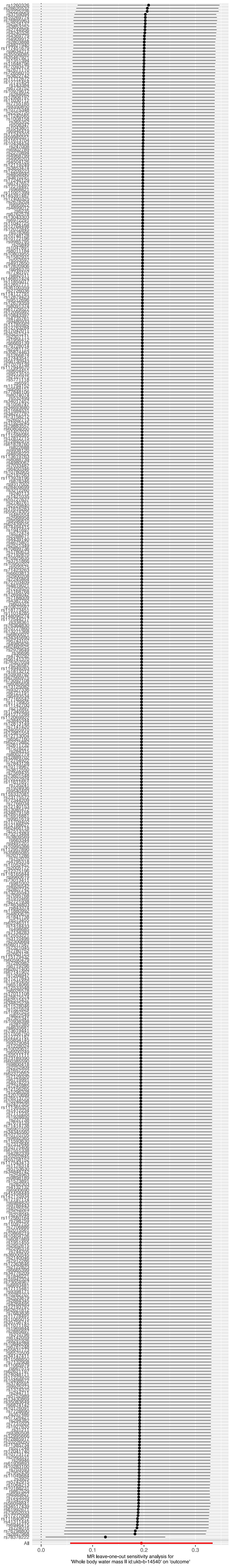

**Figure S55. Leave-one-out: exposure impedance of whole body, outcome UL.**

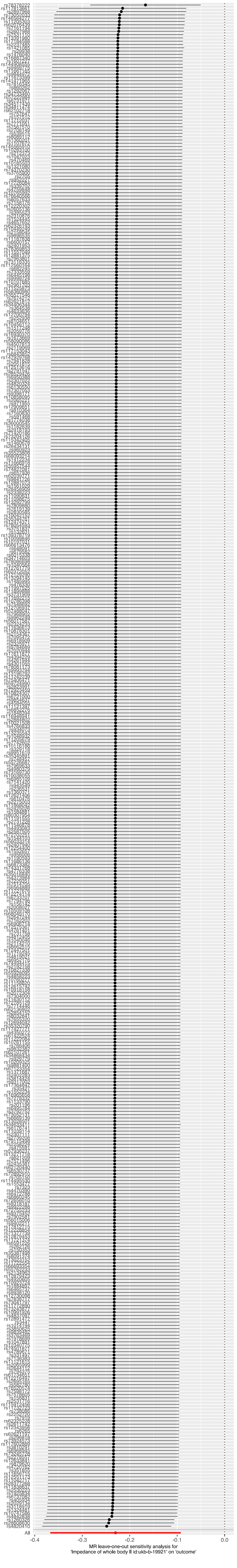

**Figure S56. Leave-one-out: exposure basal metabolic rate, outcome UL.**

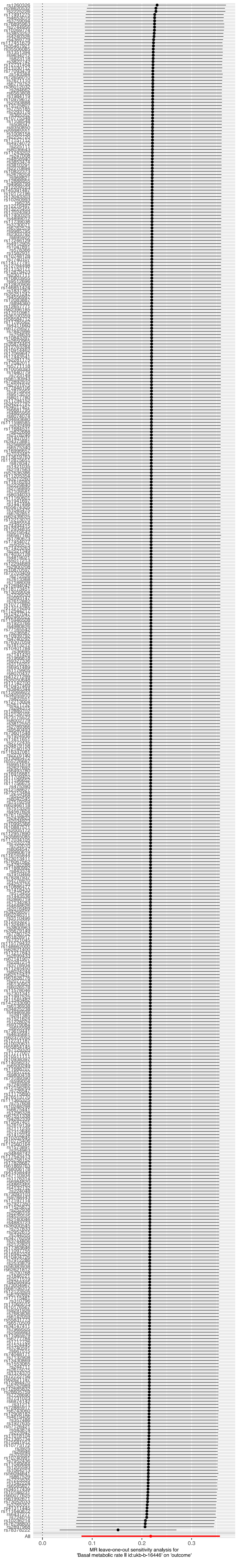

**Figure S57. Leave-one-out: exposure UL, outcome BMI.**

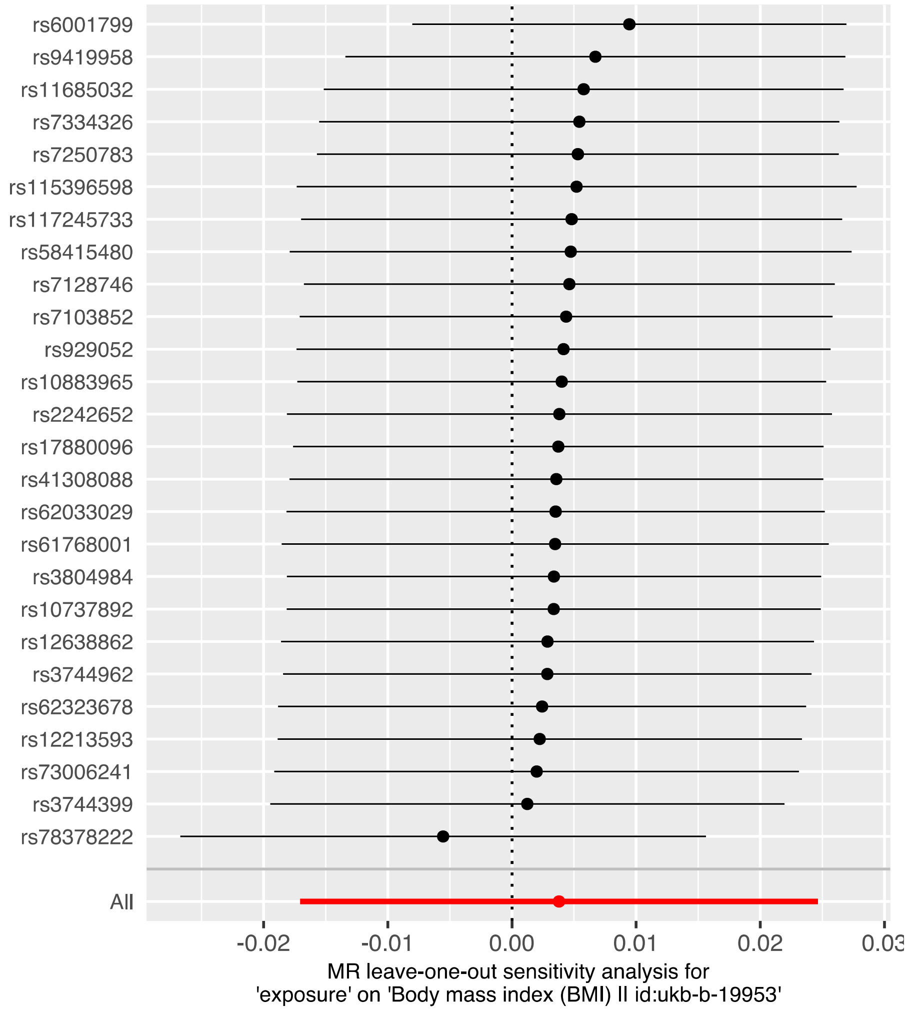

**Figure S58. Leave-one-out: exposure UL, outcome waist circumference.**

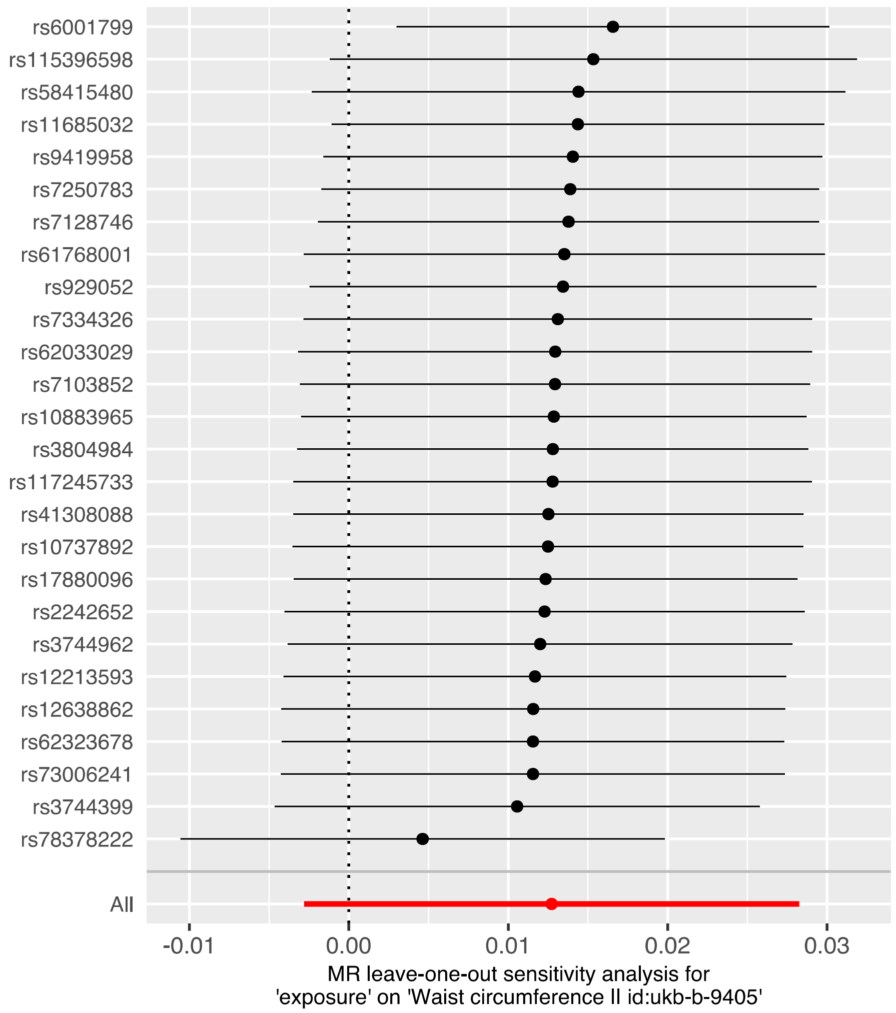

**Figure S59. Leave-one-out: exposure UL, outcome whole body fat mass.**

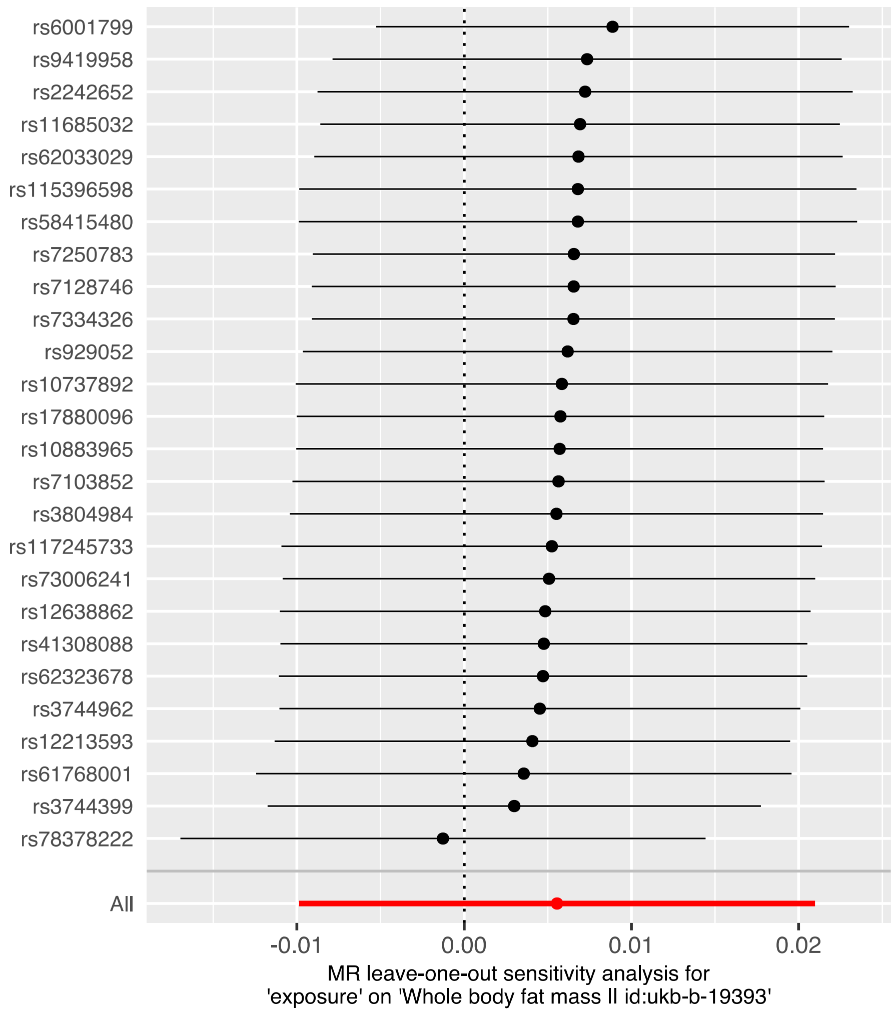

**Figure S60. Leave-one-out: exposure UL, outcome whole body fat-free mass.**

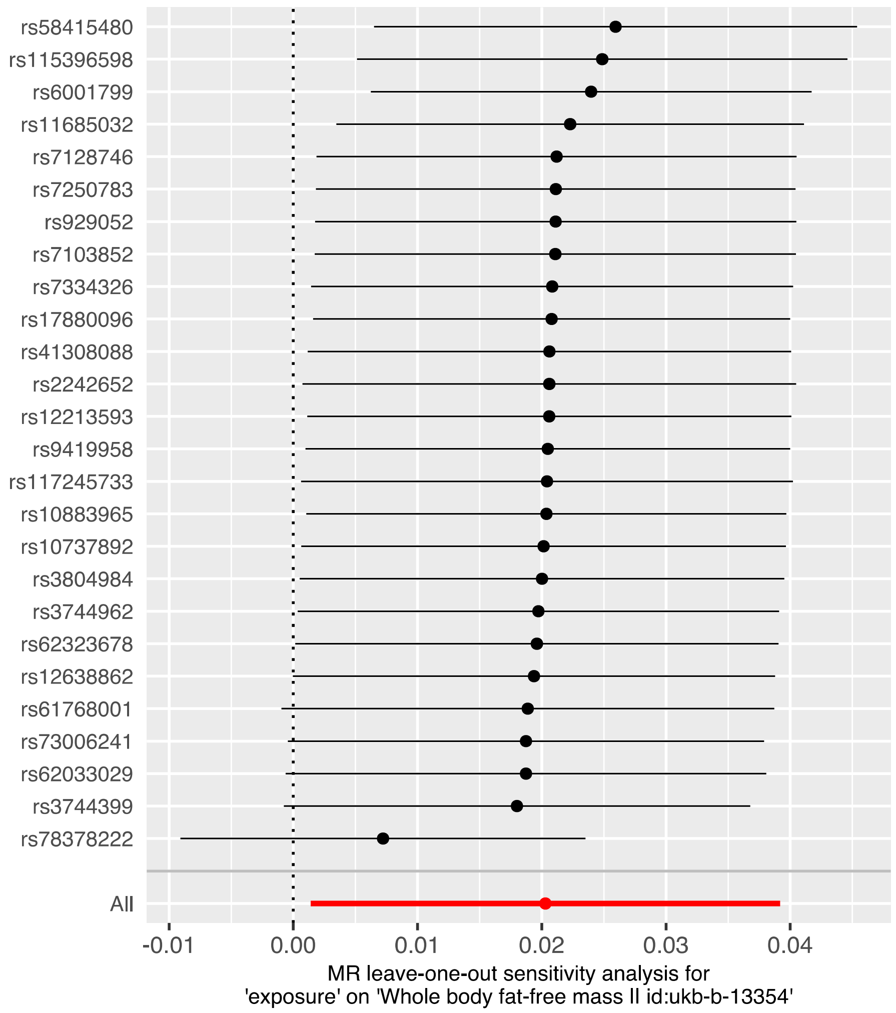

**Figure S61. Leave-one-out: exposure UL, outcome whole body water mass.**

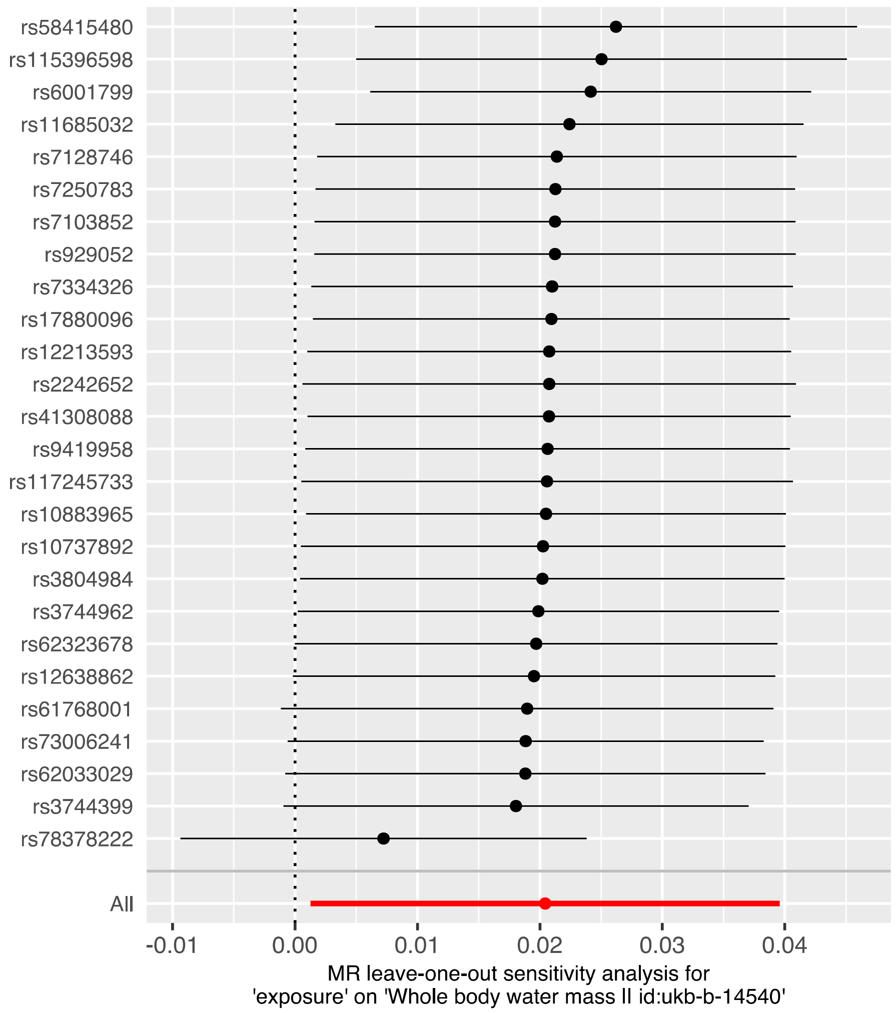

**Figure S62. Leave-one-out: exposure UL, outcome impedance of whole body.**

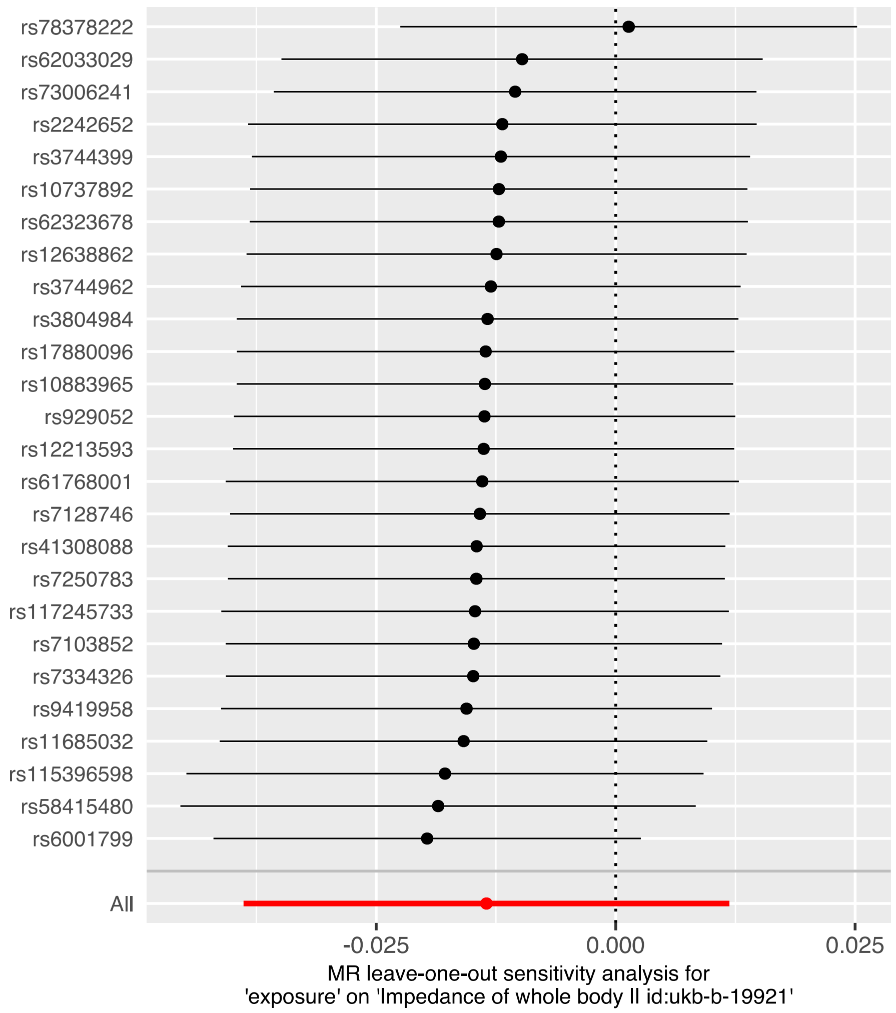

**Figure S63. Leave-one-out: exposure UL, outcome basal metabolic rate.**

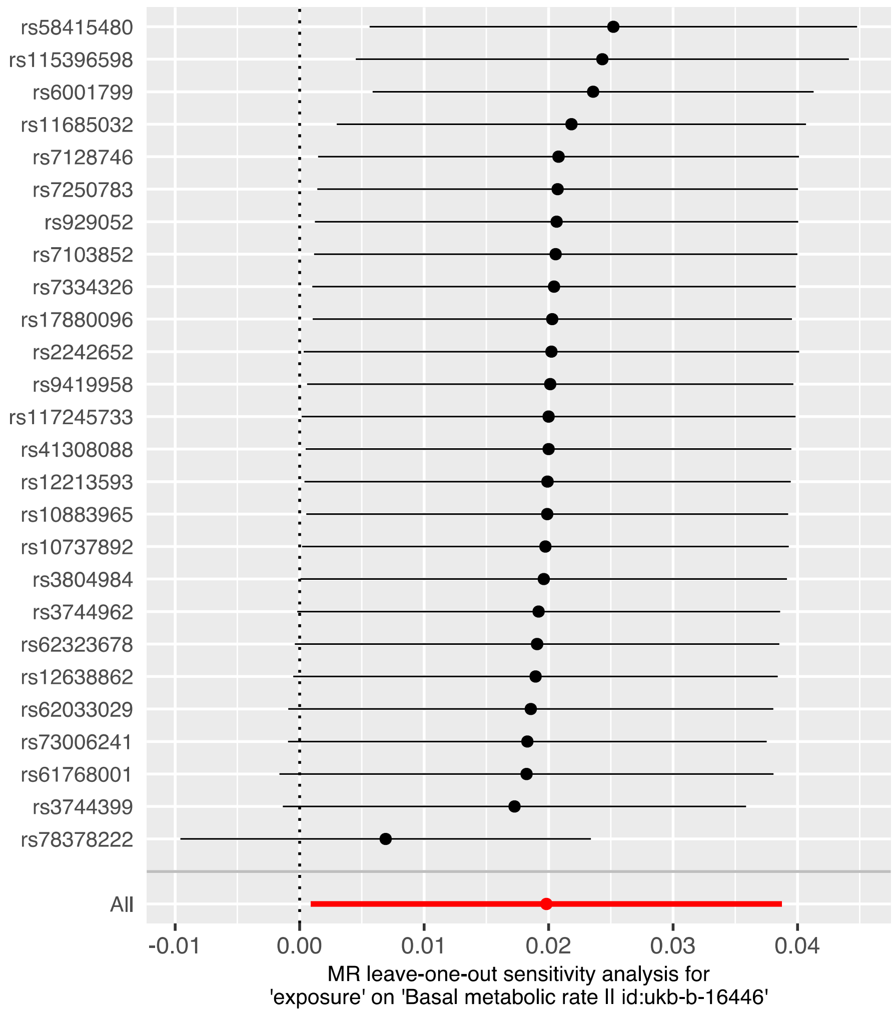

**Table S1. Genome-wide significant ($p$<5x10$^{-8}$) loci in META-1.**

| Locus | Chr:Pos (hg38) | Nearest gene | Candidate gene | rsID | EA | OR (95% CI) | P-value |
|---|---|---|---|---|---|---|---|
| 1p36.12 | 1:22141722 | *WNT4* | *WNT4* | rs3820282 | T | 1.15 (1.13-1.17) | 2.60E-49 |
| 1q24.3 | 1:172162145 | *DNM3* | *DNM3* | rs61807787 | T | 1.04 (1.03-1.06) | 3.89E-08 |
| 1q43 | 1:241860596 | *EXO1* | *EXO1,FH* | rs4149909 | G | 1.13 (1.08-1.18) | 1.16E-08 |
| 1q44 | 1:244151650 | *ZBTB18,C1orf100* | *ZBTB18* | rs2183478 | G | 1.07 (1.05-1.09) | 1.75E-11 |
| 1q44 | 1:248897507 | *PGBD2* | *ZNF692* | rs4335411 | A | 1.06 (1.04-1.08) | 4.12E-10 |
| 2p25.1 | 2:11540277 | *GREB1* | *GREB1* | rs35417544 | T | 1.08 (1.07-1.10) | 3.94E-24 |
| 2p23.2 | 2:28106534 | *BABAM2* | *BABAM2* | rs74576866 | G | 1.09 (1.06-1.11) | 7.74E-11 |
| 2p14 | 2:66863235 | *MEIS1* | *MEIS1* | rs17631680 | C | 0.93 (0.91-0.95) | 1.93E-10 |
| 2q11.2 | 2:99454113 | *REV1* | *REV1* | rs13392042 | G | 1.05 (1.03-1.06) | 4.21E-11 |
| 2q33.3 | 2:207258660 | *MYOSLID,KLF7* | *MYOSLID* | rs10804157 | C | 1.04 (1.03-1.05) | 1.04E-08 |
| 2q37.3 | 2:241720139 | *ING5* | *ING5* | rs34766121 | T | 1.06 (1.04-1.08) | 5.02E-10 |
| 3p26.1 | 3:4674530 | *ITPR1* | *ITPR1* | rs3804984 | C | 0.95 (0.93-0.96) | 8.06E-15 |
| 3p24.2 | 3:24213259 | *THRB* | *THRB* | rs1010961 | A | 1.04 (1.03-1.06) | 1.64E-10 |
| 3p24.1 | 3:27488262 | *SLC4A7* | *NEK10* | rs35701251 | A | 1.05 (1.03-1.06) | 1.59E-08 |
| 3q26.2 | 3:169768720 | *ACTRT3,TERC* | *TERC* | rs35446936 | A | 0.94 (0.92-0.95) | 1.01E-16 |
| 3q27.2 | 3:185807411 | *IGF2BP2* | *IGF2BP2* | rs13060777 | G | 1.05 (1.04-1.07) | 1.14E-11 |
| 4q12 | 4:53684007 | *LNX1* | *LNX1* | rs62323682 | C | 1.16 (1.13-1.19) | 5.67E-25 |
| 4q13.3 | 4:69735020 | *SULT1B1* | *SULT1E1* | rs12640488 | G | 1.07 (1.05-1.08) | 6.21E-21 |
| 4q22.3 | 4:94572095 | *PDLIM5* | *BMPR1B* | rs2452597 | G | 1.05 (1.03-1.06) | 9.42E-11 |
| 4q23 | 4:99031559 | *METAP1,ADH5* | *ADH5* | rs1037475 | G | 1.04 (1.03-1.05) | 8.38E-09 |
| 5p15.33 | 5:1279913 | *TERT* | *TERT* | rs2242652 | A | 1.12 (1.11-1.14) | 5.86E-41 |
| 5q31.1 | 5:133099880 | *HSPA4* | *HSPA4* | rs4367292 | T | 0.96 (0.94-0.97) | 2.49E-08 |
| 5q35.2 | 5:177023836 | *ZNF346* | *UIMC1,FGFR4* | rs2456181 | G | 1.05 (1.04-1.07) | 4.20E-12 |
| 6p21.31 | 6:34240996 | *HMGA1* | *HMGA1* | rs41269026 | A | 1.11 (1.07-1.15) | 4.02E-09 |
| 6p21.2 | 6:36653670 | *CDKN1A* | *CDKN1A* | rs10456443 | A | 0.95 (0.93-0.96) | 2.67E-10 |
| 6q21 | 6:109054915 | *SESN1* | *SESN1* | rs11153158 | C | 0.93 (0.92-0.95) | 1.05E-10 |
| 6q25.2 | 6:152241136 | *SYNE1* | *ESR1* | rs58415480 | G | 1.22 (1.19-1.24) | 1.86E-104 |
| 7p14.3 | 7:33008785 | *FKBP9,NT5C3A* | *BBS9* | rs4723230 | T | 1.05 (1.03-1.07) | 4.68E-08 |
| 7q31.2 | 7:117273513 | *WNT2* | *WNT2* | rs2270206 | A | 1.06 (1.04-1.08) | 1.24E-09 |
| 7q31.31 | 7:121132432 | *CPED1,WNT16* | *WNT16* | rs12706314 | A | 1.04 (1.03-1.06) | 2.69E-10 |
| 7q32.3 | 7:130935964 | *LINC-PINT* | *LINC-PINT* | rs35908158 | C | 1.08 (1.05-1.10) | 1.60E-08 |
| 8p12 | 8:30452819 | *RBPMS* | *RBPMS* | rs13275869 | C | 0.96 (0.95-0.97) | 8.64E-09 |
| 8q24.21 | 8:128506035 | *LINC00824* | *LINC00824* | rs1516980 | C | 0.96 (0.94-0.97) | 2.72E-08 |
| 9p24.3 | 9:680714 | *KANK1,ANKRD15* | *KANK1* | rs10815466 | A | 1.10 (1.08-1.12) | 9.51E-24 |
| 9q22.2 | 9:89639982 | *GADD45G,SEMA4D* | *GADD45G* | rs28508285 | G | 1.07 (1.04-1.09) | 1.49E-08 |
| 10p12.31 | 10:21517903 | *SKIDA1* | *DNAJC1* | rs946711 | C | 1.05 (1.03-1.06) | 2.96E-10 |
| 10p11.22 | 10:31678920 | *ZEB1,ARHGAP12* | *ZEB1* | rs72784785 | C | 0.93 (0.92-0.95) | 2.35E-16 |
| 10q23.31 | 10:88331783 | *RNLS* | *RNLS* | rs1426619 | T | 1.04 (1.03-1.06) | 4.92E-09 |
| 10q24.33 | 10:103918139 | *STN1* | *SH3PXD2A* | rs4387287 | C | 0.91 (0.89-0.92) | 5.61E-25 |
| 11p15.5 | 11:197557 | *ODF3,BET1L* | *PKP3* | rs7103852 | G | 1.13 (1.10-1.16) | 5.10E-21 |
| 11p14.1 | 11:30204981 | *FSHB* | *FSHB* | rs11031006 | A | 0.91 (0.89-0.93) | 1.62E-22 |
| 11p13 | 11:32342641 | *WT1* | *WT1* | rs2057178 | A | 1.14 (1.12-1.17) | 7.32E-41 |
| 11p13 | 11:35062086 | *PDHX* | *CD44* | rs2553773 | G | 1.07 (1.06-1.09) | 1.19E-23 |
| 11q22.3 | 11:108444879 | *C11orf65* | *ATM* | rs149934734 | T | 1.36 (1.29-1.42) | 4.74E-36 |
| 11q23.2 | 11:112703765 | *LOC105369496* | *LOC105369496* | rs10891420 | C | 1.05 (1.03-1.06) | 1.38E-10 |
| 12q13.11 | 12:46402739 | *SLC38A2* | *SLC38A2* | rs2131371 | C | 1.08 (1.06-1.09) | 1.03E-23 |
| 12q15 | 12:70756878 | *PTPRR* | *PTPRR* | rs11178393 | C | 0.92 (0.90-0.95) | 7.46E-12 |
| 12q24.31 | 12:123379073 | *KMT5A,PITPNM2* | *KMT5A* | rs28583837 | A | 0.95 (0.93-0.96) | 9.67E-10 |
| 13q14.11 | 13:40149807 | *FOXO1* | *FOXO1* | rs117245733 | A | 1.42 (1.35-1.49) | 1.78E-41 |
| 15q23 | 15:67922458 | *SKOR1,PIAS1* | *PIAS1* | rs12148374 | C | 0.96 (0.95-0.97) | 1.37E-09 |
| 16q12.1 | 16:50059327 | *HEATR3* | *BRD7* | rs12599260 | A | 1.05 (1.04-1.07) | 1.09E-11 |
| 16q12.1 | 16:51447685 | *AC007344.1* | *AC007344.1* | rs66998222 | A | 0.94 (0.93-0.96) | 7.53E-12 |
| 17p13.1 | 17:7668434 | *TP53* | *TP53* | rs78378222 | G | 1.81 (1.71-1.92) | 3.88E-86 |
| 17p12 | 17:12652500 | *MYOCD* | *MYOCD* | rs12601765 | T | 1.04 (1.03-1.06) | 3.08E-08 |
| 19p12 | 19:22032639 | *ZNF257* | *ZNF257* | rs8105767 | G | 1.05 (1.03-1.06) | 2.63E-09 |
| 20p12.3 | 20:5967581 | *MCM8* | *MCM8* | rs16991615 | A | 1.12 (1.09-1.15) | 5.15E-13 |
| 20q13.31 | 20:57441016 | *CTCFL* | *RBM38* | rs13039273 | C | 1.04 (1.03-1.06) | 3.08E-09 |
| 20q13.33 | 20:63638397 | *STMN3* | *SLC2A4RG* | rs75691080 | T | 0.92 (0.90-0.95) | 7.79E-11 |
| 21q22.12 | 21:35072824 | *RUNX1* | *RUNX1* | rs2834747 | G | 0.96 (0.94-0.97) | 1.44E-08 |
| 22q12.3 | 22:36287509 | *MYH9,APOL1* | *MYH9* | rs9610482 | T | 1.06 (1.04-1.08) | 7.89E-11 |
| 22q13.1 | 22:40269221 | *TNRC6B* | *MKL1* | rs112251865 | T | 1.09 (1.07-1.11) | 1.21E-26 |
| Xq13.1 | X:70928740 | *SLC7A3,MED12* | *FOXO4* | rs5936604 | C | 0.93 (0.91-0.94) | 1.00E-17 |
| Xq26.2 | X:132178061 | *FRMD7,RAP2C* | *RAP2C* | rs5930554 | C | 1.16 (1.14-1.18) | 1.04E-59 |

**Table S2. Genome-wide significant ($p$<5x10$^{-8}$) loci in META-2.**

| Locus | Chr:Pos (hg19) | Nearest gene | Candidate gene | rsID | EA | OR (95% CI) | P-value |
|---|---|---|---|---|---|---|---|
| 1p36.12 | 1:22468215 | *WNT4* | *WNT4* | rs3820282 | T | 1.16 (1.13-1.19) | 2,63E-34 |
| 1q43 | 1:242023898 | *EXO1* | *EXO1,FH* | rs4149909 | A | 0.87 (0.82-0.91) | 1,18E-08 |
| 1q44 | 1:244314952 | *ZBTB18,C1orf100* | *ZBTB18* | rs2183478 | A | 0.93 (0.91-0.95) | 3,44E-11 |
| 1q44 | 1:249191706 | *PGBD2* | *ZNF692* | rs4335411 | A | 1.07 (1.04-1.09) | 5,53E-09 |
| 2p25.1 | 2:11670305 | *GREB1* | *GREB1* | rs10929753 | A | 0.93 (0.91-0.95) | 9,25E-15 |
| 2p23.2 | 2:27730940 | *GCKR,BABAM2* | *BABAM2* | rs1260326 | T | 1.06 (1.04-1.08) | 1,36E-09 |
| 2p14 | 2:67090367 | *MEIS1* | *MEIS1* | rs17631680 | T | 1.11 (1.07-1.14) | 9,01E-12 |
| 2p11.2 | 2:86685141 | *KDM3A* | *KDM3A* | rs573520030 | T | 0.62 (0.53-0.74) | 3,87E-08 |
| 2q11.2 | 2:99945794 | *TXNDC9,REV1* | *REV1* | rs58086269 | A | 0.95 (0.93-0.97) | 6,50E-09 |
| 2q33.3 | 2:208123384 | *MYOSLID,KLF7* | *MYOSLID* | rs10804157 | T | 0.95 (0.93-0.97) | 7,65E-09 |
| 2q37.3 | 2:242649793 | *ING5* | *ING5* | rs6437284 | C | 0.94 (0.92-0.96) | 1,68E-09 |
| 3p26.1 | 3:4716214 | *ITPR1* | *ITPR1* | rs3804984 | T | 1.07 (1.05-1.09) | 1,58E-12 |
| 3p24.2 | 3:24260363 | *THRB* | *THRB* | rs4858590 | T | 1.05 (1.03-1.07) | 2,24E-08 |
| 3q26.2 | 3:169482335 | *TERC* | *TERC* | rs2293607 | T | 1.09 (1.07-1.12) | 1,25E-18 |
| 3q27.2 | 3:185523370 | *IGF2BP2* | *IGF2BP2* | rs66513933 | T | 0.95 (0.93-0.97) | 3,94E-08 |
| 4p11 | 4:49087575 | *CWH43* | *OCIAD1* | rs538533131 | A | 0.53 (0.43-0.65) | 2,61E-09 |
| 4q12 | 4:54550174 | *LNX1* | *LNX1* | rs62323682 | T | 0.85 (0.82-0.88) | 4,42E-18 |
| 4q13.3 | 4:70647778 | *SULTIB1* | *SULT1E1* | rs35487626 | T | 1.08 (1.06-1.10) | 1,03E-16 |
| 4q23 | 4:99978183 | *METAP1* | *ADH5* | rs12512420 | A | 0.95 (0.93-0.97) | 3,46E-08 |
| 5p15.3 | 5:1279790 | *TERT* | *TERT* | rs10069690 | T | 1.13 (1.11-1.15) | 2,40E-33 |
| 5q35.2 | 5:176450837 | *ZNF346* | *UIMC1,FGFR4* | rs2456181 | C | 0.95 (0.93-0.97) | 6,71E-09 |
| 6p21.2 | 6:36622549 | *CDKN1A* | *CDKN1A* | rs12528913 | A | 0.93 (0.91-0.95) | 4,45E-10 |
| 6q21 | 6:109376118 | *SESN1* | *SESN1* | rs11153158 | T | 1.08 (1.05-1.11) | 8,72E-09 |
| 6q25.2 | 6:152562271 | *SYNE1* | *ESR1* | rs58415480 | C | 0.81 (0.79-0.83) | 9,54E-74 |
| 7q31.31 | 7:120447460 | *TSPAN12,WNT16* | *WNT16* | rs4730982 | T | 0.95 (0.93-0.97) | 4,98E-08 |
| 8q24.21 | 8:130479731 | *LINC00824* | *LINC00824* | rs11786929 | T | 0.94 (0.92-0.96) | 1,09E-09 |
| 9p24.3 | 9:802228 | *DMRT1,KANK1* | *KANK1* | rs7027685 | A | 0.91 (0.89-0.92) | 1,67E-26 |
| 10p12.31 | 10:21983960 | *MLLT10,SKIDA1* | *DNAJC1* | rs1243192 | A | 1.06 (1.04-1.08) | 2,75E-09 |
| 10p11.22 | 10:31913890 | *ZEB1* | *ZEB1* | rs11008551 | T | 0.93 (0.91-0.95) | 7,26E-12 |
| 10q24.32 | 10:104548027 | *WBP1L,STN1* | *SH3PXD2A* | rs75731980 | T | 1.23 (1.17-1.30) | 1,43E-14 |
| 10q25.1 | 10:108581428 | *SORCS1* | *SORCS1* | rs12247648 | T | 1.26 (1.17-1.36) | 3,86E-09 |
| 11p15.5 | 11:197557 | *ODF3,BET1L* | *PKP3* | rs7103852 | A | 0.86 (0.84-0.89) | 8,06E-18 |
| 11p14.1 | 11:30338842 | *ARL14EP,FSHB* | *FSHB* | rs7947350 | A | 1.08 (1.06-1.11) | 1,61E-11 |
| 11p13 | 11:32365430 | *WT1* | *WT1* | rs11031731 | A | 1.18 (1.15-1.21) | 2,90E-33 |
| 11p13 | 11:35096995 | *CD44,PDHX* | *CD44* | rs429503 | T | 1.08 (1.06-1.10) | 6,99E-18 |
| 11q22.3 | 11:108149207 | *ATM,C11orf65* | *ATM* | rs141379009 | T | 0.70 (0.65-0.74) | 1,12E-27 |
| 11q23.2 | 11:112574488 | *LOC105369496* | *LOC105369496* | rs10891420 | T | 0.95 (0.93-0.96) | 5,21E-10 |
| 12q13.11 | 12:46831129 | *SLC38A2* | *SLC38A2* | rs12832777 | T | 0.93 (0.91-0.94) | 4,01E-15 |
| 12q24.31 | 12:123890276 | *KMT5A* | *KMT5A* | rs28576953 | T | 0.94 (0.92-0.96) | 4,45E-08 |
| 13q14.11 | 13:40723944 | *FOXO1* | *FOXO1* | rs117245733 | A | 1.48 (1.39-1.57) | 4,36E-35 |
| 15q23 | 15:68628163 | *ITGA11,SKOR1* | *PIAS1* | rs2306022 | T | 1.09 (1.06-1.13) | 1,73E-08 |
| 16q12.1 | 16:50109100 | *HEATR3* | *BRD7* | rs9939688 | T | 0.93 (0.91-0.95) | 1,96E-13 |
| 16q12.1 | 16:51481596 | *AC007344.1* | *AC007344.1* | rs66998222 | A | 0.92 (0.90-0.94) | 1,63E-13 |
| 17p13.1 | 17:7571752 | *TP53* | *TP53* | rs78378222 | T | 0.47 (0.43-0.51) | 5,10E-85 |
| 19p12 | 19:22215441 | *ZNF257* | *ZNF257* | rs8105767 | A | 0.94 (0.92-0.96) | 6,38E-10 |
| 20p12.3 | 20:5948227 | *MCM8* | *MCM8* | rs16991615 | A | 1.14 (1.10-1.19) | 4,72E-10 |
| 22q12.3 | 22:36683555 | *MYH9,APOL1* | *MYH9* | rs9610482 | T | 1.07 (1.05-1.10) | 1,35E-09 |
| 22q13.1 | 22:40665225 | *TNRC6B* | *MKL1* | rs112251865 | T | 1.10 (1.08-1.13) | 5,77E-20 |
| Xp11.23 | 23:46763494 | *JADE3,RP2* | *SLC9A7* | rs6611312 | T | 1.05 (1.03-1.07) | 2,54E-08 |
| Xq13.1 | 23:70108889 | *TEX11, SLC7A3* | *FOXO4* | rs13441059 | A | 1.11 (1.09-1.13) | 8,95E-27 |
| Xq26.2 | 23:131312089 | *FRMD7,RAP2C* | *RAP2C* | rs5930554 | T | 0.86 (0.84-0.88) | 1,26E-54 |

**Table S3. RegulomeDB annotation of the association lead variants near *MYOCD*.**

The RegulomeDB probability score is ranging from 0 to 1, with 1 being most likely to be a regulatory variant. 'Ranking' refers to following evidence: 1a, eQTL + TF binding + matched TF motif + matched DNase Footprint + DNase peak; 1b, eQTL + TF binding + any motif + DNase Footprint + DNase peak; 1c, eQTL + TF binding + matched TF motif + DNase peak; 1d, eQTL + TF binding + any motif + DNase peak; 1e, eQTL + TF binding + matched TF motif; 1f, eQTL + TF binding / DNase peak; 2a, TF binding + matched TF motif + matched DNase Footprint + DNase peak; 2b, TF binding + any motif + DNase Footprint + DNase peak; 2c, TF binding + matched TF motif + DNase peak; 3a, TF binding + any motif + DNase peak; 3b, TF binding + matched TF motif; 4, TF binding + DNase peak; 5, TF binding or DNase peak; 6, Motif hit; 7, Other. Data was downloaded from RegulomeDB 02/10/2021.

| Rsids | Probability | Ranking | ChIP | DNase | Footprint | Footprint matched | IC matched max | IC max | PWM | PWM matched | QTL |
|---|---|---|---|---|---|---|---|---|---|---|---|
| rs11871444 | 0.60906 | 4 | TRUE | TRUE | FALSE | FALSE | 0.0 | 0.0 | FALSE | FALSE | FALSE |
| rs12601765 | 0.0 | 5 | FALSE | TRUE | FALSE | FALSE | 0.0 | 0.780 | TRUE | FALSE | FALSE |
| rs12601724 | 0.13454 | 5 | FALSE | TRUE | FALSE | FALSE | 0.0 | 0.0 | FALSE | FALSE | FALSE |
| rs4792271 | 0.00167 | 6 | FALSE | FALSE | FALSE | FALSE | 0.0 | 0.270 | TRUE | FALSE | FALSE |
| rs4792272 | 0.13454 | 5 | FALSE | TRUE | FALSE | FALSE | 0.0 | 0.0 | FALSE | FALSE | FALSE |
| rs11870377 | 0.58955 | 5 | TRUE | FALSE | FALSE | FALSE | 0.0 | 0.0 | FALSE | FALSE | FALSE |
| rs72811248 | 0.58955 | 5 | TRUE | FALSE | FALSE | FALSE | 0.0 | 0.0 | FALSE | FALSE | FALSE |

**Table S4. RegulomeDB annotation of the association lead variants near *MYOSLID*.**

The RegulomeDB probability score is ranging from 0 to 1, with 1 being most likely to be a regulatory variant. 'Ranking' refers to following evidence: 1a, eQTL + TF binding + matched TF motif + matched DNase Footprint + DNase peak; 1b, eQTL + TF binding + any motif + DNase Footprint + DNase peak; 1c, eQTL + TF binding + matched TF motif + DNase peak; 1d, eQTL + TF binding + any motif + DNase peak; 1e, eQTL + TF binding + matched TF motif; 1f, eQTL + TF binding / DNase peak; 2a, TF binding + matched TF motif + matched DNase Footprint + DNase peak; 2b, TF binding + any motif + DNase Footprint + DNase peak; 2c, TF binding + matched TF motif + DNase peak; 3a, TF binding + any motif + DNase peak; 3b, TF binding + matched TF motif; 4, TF binding + DNase peak; 5, TF binding or DNase peak; 6, Motif hit; 7, Other. Data was downloaded from RegulomeDB 02/10/2021.

| Rsids | Probability | Ranking | ChIP | DNase | Footprint | Footprint matched | IC matched max | IC max | PWM | PWM matched | QTL |
|---|---|---|---|---|---|---|---|---|---|---|---|
| rs6758102 | 0.60906 | 4 | TRUE | TRUE | FALSE | FALSE | 0.0 | 0.0 | FALSE | FALSE | FALSE |
| rs10804157 | 1.0 | 2b | TRUE | TRUE | TRUE | FALSE | 0.0 | 0.720 | TRUE | FALSE | FALSE |
| rs6709320 | 0.18412 | 7 | FALSE | FALSE | FALSE | FALSE | 0.0 | 0.0 | FALSE | FALSE | FALSE |
| rs2111595 | 0.60906 | 4 | TRUE | TRUE | FALSE | FALSE | 0.0 | 0.0 | FALSE | FALSE | FALSE |
| rs11694764 | 0.18412 | 7 | FALSE | FALSE | FALSE | FALSE | 0.0 | 0.0 | FALSE | FALSE | FALSE |
| rs56167178 | 0.13454 | 5 | FALSE | TRUE | FALSE | FALSE | 0.0 | 0.0 | FALSE | FALSE | FALSE |

**Table S5. ChIP-seq data of rs10804157.**

The variant rs10804157 showed the highest probability (1.0) of being a regulatory variant among the variants showing genome-wide significant ($p<5x10^{-8}$) association with UL at 2q33.3. ChIP-seq data for this variant was extracted from RegulomeDB on 02/10/2021.

| Method | Peak location | Biosample | Targets | Organ | Dataset | File | Value | Strand |
|---|---|---|---|---|---|---|---|---|
| ChIP-seq | chr2:208123084..208123749 | A549 | SP1 | lung | ENCSR000BPE | ENCFF348RKC | 326.58420 | - |
| ChIP-seq | chr2:208123107..208123712 | A549 | EP300 | lung | ENCSR000BPW | ENCFF002CFV | 287.620948909819 | - |
| ChIP-seq | chr2:208123262..208123522 | A549 | NR3C1 | lung | ENCSR000BHF | ENCFF648ZNE | 244.46238 | - |
| ChIP-seq | chr2:208123094..208123709 | A549 | TCF12 | lung | ENCSR000BQQ | ENCFF672FQU | 221.07448 | - |
| ChIP-seq | chr2:208123378..208123656 | MCF-7 | FOS | mammary gland | ENCSR569XNP | ENCFF965AIZ | 206.29520 | - |
| ChIP-seq | chr2:208123332..208123669 | A549 | FOSL2 | lung | ENCSR000BQO | ENCFF374ZCG | 187.18388 | - |
| ChIP-seq | chr2:208123364..208123572 | A549 | FOXA1 | lung | ENCSR000BRD | ENCFF755AXA | 170.22291 | - |
| ChIP-seq | chr2:208123171..208123507 | MCF-7 | ESRRA | mammary gland | ENCSR954WVZ | ENCFF044NEB | 156.03072 | - |
| ChIP-seq | chr2:208123287..208123667 | A549 | MAFK | lung | ENCSR541WQI | ENCFF530ZTE | 139.13663 | - |
| ChIP-seq | chr2:208123361..208123595 | HEK293 | ZBTB17 | kidney | ENCSR631WAA | ENCFF047LUI | 114.24409 | - |
| ChIP-seq | chr2:208123179..208123669 | MCF-7 | ZNF217 | mammary gland | ENCSR465XQW | ENCFF567MGV | 99.76437 | - |
| ChIP-seq | chr2:208123231..208123661 | MCF-7 | ZNF579 | mammary gland | ENCSR018MQH | ENCFF854CXA | 98.09751 | - |
| ChIP-seq | chr2:208123265..208123545 | A549 | NR3C1 | lung | ENCSR000BHG | ENCFF913WFD | 97.82031 | - |
| ChIP-seq | chr2:208123181..208123801 | A549 | SIN3A | lung | ENCSR513XQX | ENCFF172VGB | 94.53579 | - |
| ChIP-seq | chr2:208123210..208123666 | MCF-7 | MNT | mammary gland | ENCSR663ZZZ | ENCFF628SBD | 84.80959 | - |
| ChIP-seq | chr2:208123135..208123675 | A549 | BCL3 | lung | ENCSR000BQH | ENCFF325WAV | 84.05348 | - |
| ChIP-seq | chr2:208123311..208123581 | IMR-90 | MAFK | lung | ENCSR000EFH | ENCFF593MEB | 83.58383 | - |
| ChIP-seq | chr2:208123232..208123622 | MCF-7 | FOXA1 | mammary gland | ENCSR126YEB | ENCFF596OJV | 82.25613 | - |
| ChIP-seq | chr2:208123208..208123618 | MCF-7 | CTBP1 | mammary gland | ENCSR636EYA | ENCFF785ZQF | 81.59685 | - |
| ChIP-seq | chr2:208123147..208123591 | liver | RXRA | liver | ENCSR098XMN | ENCFF189HPG | 75.54675 | - |
| ChIP-seq | chr2:208123227..208123697 | A549 | ZBTB33 | lung | ENCSR000BPZ | ENCFF639NGJ | 68.14892 | - |
| ChIP-seq | chr2:208123237..208123541 | A549 | NR3C1 | lung | ENCSR000BJR | ENCFF080RMP | 63.97341 | - |
| ChIP-seq | chr2:208123188..208123618 | HEK293 | KLF1 | kidney | ENCSR859BMR | ENCFF620ZDF | 61.18339 | - |
| ChIP-seq | chr2:208123153..208123417 | A549 | ESRRA | lung | ENCSR473SUA | ENCFF266ORV | 60.61061 | - |
| ChIP-seq | chr2:208123318..208123702 | MCF-7 | MAZ | mammary gland | ENCSR288IJC | ENCFF492DZG | 58.62570 | - |
| ChIP-seq | chr2:208123059..208123459 | endothelial cell of umbilical vein | FOS | vein, vasculature, epithelium, blood vessel | ENCSR000EVU | ENCFF390MXI | 56.59256 | - |
| ChIP-seq | chr2:208123235..208123645 | MCF-7 | NFIB | mammary gland | ENCSR702BYX | ENCFF817RST | 56.19887 | - |
| ChIP-seq | chr2:208123294..208123578 | IMR-90 | BHLHE40 | lung | ENCSR957KYB | ENCFF942FBW | 53.93919 | - |
| ChIP-seq | chr2:208122889..208123613 | K562 | FOSL1 | blood, bodily fluid | ENCSR239ZLZ | ENCFF692PVV | 52.28088 | - |
| ChIP-seq | chr2:208123124..208123544 | liver | NR2F2 | liver | ENCSR338MMB | ENCFF266KLW | 50.28680 | - |
| ChIP-seq | chr2:208123248..208123738 | A549 | REST | lung | ENCSR000BQP | ENCFF136RBA | 49.93059 | - |
| ChIP-seq | chr2:208123340..208123630 | A549 | ZFP36 | lung | ENCSR294JWV | ENCFF418DRY | 49.62579 | - |
| ChIP-seq | chr2:208123096..208123416 | SK-N-SH | JUND | brain | ENCSR000BSK | ENCFF598ZLV | 49.00620 | - |
| ChIP-seq | chr2:208123233..208123757 | A549 | YY1 | lung | ENCSR000BPM | ENCFF713ISJ | 48.03768 | - |
| ChIP-seq | chr2:208123332..208124008 | 22Rv1 | CTCF | prostate gland | ENCSR857PBV | ENCFF030BPR | 44.66346 | - |
| ChIP-seq | chr2:208123056..208123476 | liver | JUND | liver | ENCSR837GTK | ENCFF804UDG | 44.66249 | - |
| ChIP-seq | chr2:208123048..208123638 | MCF-7 | FOSL2 | mammary gland | ENCSR546KCN | ENCFF331KNS | 43.40140 | - |
| ChIP-seq | chr2:208123246..208123730 | A549 | ATF3 | lung | ENCSR000BPS | ENCFF913MUA | 43.04867 | - |

| Method | Peak location | Biosample | Targets | Organ | Dataset | File | Value | Strand |
|---|---|---|---|---|---|---|---|---|
| ChIP-seq | chr2:208123340..208124040 | 22Rv1 | CTCF | prostate gland | ENCSR847XGE | ENCFF259XYR | 41.50506 | - |
| ChIP-seq | chr2:208123167..208123671 | liver | SP1 | liver | ENCSR386YIH | ENCFF017YUI | 41.36437 | - |
| ChIP-seq | chr2:208123257..208123587 | T47D | EP300 | mammary gland | ENCSR000BLM | ENCFF002CNZ | 39.636132175516 | - |
| ChIP-seq | chr2:208123176..208123640 | liver | RXRA | liver | ENCSR352QSB | ENCFF651LIG | 39.78583 | - |
| ChIP-seq | chr2:208123138..208123628 | liver | NR2F2 | liver | ENCSR168SMX | ENCFF168INE | 38.88749 | - |
| ChIP-seq | chr2:208123233..208123877 | HEK293 | TRIM28 | kidney | ENCSR618HNF | ENCFF034GYQ | 37.81934 | - |
| ChIP-seq | chr2:208123305..208123735 | MCF-7 | ZNF687 | mammary gland | ENCSR899BKM | ENCFF955YVT | 37.18623 | - |
| ChIP-seq | chr2:208123250..208123560 | MCF-7 | MYC | mammary gland | ENCSR000DMQ | ENCFF695IQU | 37.15063 | - |
| ChIP-seq | chr2:208123277..208123697 | A549 | ELF1 | lung | ENCSR000BPT | ENCFF533NIV | 37.00036 | - |
| ChIP-seq | chr2:208122967..208123727 | MCF-7 | KLF4 | mammary gland | ENCSR265WJC | ENCFF463ZWH | 34.95441 | - |
| ChIP-seq | chr2:208123205..208123815 | IMR-90 | POLR2A | lung | ENCSR000EFK | ENCFF002CVJ | 33.0223703871219 | - |
| ChIP-seq | chr2:208123269..208123669 | endothelial cell of umbilical vein | FOS | vein, vasculature, epithelium, blood vessel | ENCSR000EVU | ENCFF390MXI | 32.41105 | - |
| ChIP-seq | chr2:208123383..208123623 | IMR-90 | NFE2L2 | lung | ENCSR197WGI | ENCFF641RNZ | 31.19285 | - |
| ChIP-seq | chr2:208123247..208123678 | Caco-2 | HNF4A | intestine, large intestine | TSTSR342350 | TSTFF265946 | 30 | - |
| ChIP-seq | chr2:208123293..208123583 | T47D | GATA3 | mammary gland | ENCSR000BMX | ENCFF058XAD | 29.56353 | - |
| ChIP-seq | chr2:208123281..208123601 | A549 | NR3C1 | lung | ENCSR000BJT | ENCFF495YRW | 29.31897 | - |
| ChIP-seq | chr2:208123261..208123531 | A549 | NR3C1 | lung | ENCSR000BHE | ENCFF841IFL | 29.14627 | - |
| ChIP-seq | chr2:208123248..208123838 | MCF-7 | FOSL2 | mammary gland | ENCSR546KCN | ENCFF331KNS | 29.01817 | - |
| ChIP-seq | chr2:208123298..208123598 | MCF 10A | MYC | mammary gland | ENCSR000DOM | ENCFF002CZA | 24.1612783298454 | - |
| ChIP-seq | chr2:208123121..208123685 | IMR-90 | CHD1 | lung | ENCSR000EFC | ENCFF441IFB | 24.73066 | - |
| ChIP-seq | chr2:208123247..208123517 | A549 | USF1 | lung | ENCSR000BHX | ENCFF002CGC | 23.8430557308026 | - |
| ChIP-seq | chr2:208123084..208123749 | A549 | SP1 | lung | ENCSR000BPE | ENCFF348RKC | 23.40951 | - |
| ChIP-seq | chr2:208123079..208123399 | HeLa-S3 | JUND | uterus | ENCSR000EDH | ENCFF002CSP | 22.3246370112007 | - |
| ChIP-seq | chr2:208123359..208123703 | GM12878 | BCL3 | blood, bodily fluid | ENCSR000BNQ | ENCFF625SHY | 20.58068 | - |
| ChIP-seq | chr2:208123138..208123394 | HeLa-S3 | FOS | uterus | ENCSR000EZE | ENCFF002CSB | 19.8552135147245 | - |
| ChIP-seq | chr2:208123318..208123638 | SK-N-SH | JUND | brain | ENCSR000BSK | ENCFF598ZLV | 19.05498 | - |
| ChIP-seq | chr2:208123375..208123631 | A549 | USF1 | lung | ENCSR000BJB | ENCFF002CGD | 18.2821813113775 | - |
| ChIP-seq | chr2:208123323..208123739 | Caco-2 | CDX2 | intestine, large intestine | TSTSR586454 | TSTFF910843 | 18 | - |
| ChIP-seq | chr2:208123114..208123530 | liver | ATF3 | liver | ENCSR480LIS | ENCFF975PTM | 17.59478 | - |
| ChIP-seq | chr2:208122865..208123405 | MCF 10A | POLR2A | mammary gland | ENCSR000DPA | ENCFF755ZCF | 17.14568 | - |
| ChIP-seq | chr2:208123302..208123558 | MCF 10A | STAT3 | mammary gland | ENCSR000DOZ | ENCFF807JAC | 14.95877 | - |
| ChIP-seq | chr2:208123094..208123714 | A549 | TCF12 | lung | ENCSR000BQQ | ENCFF672FQU | 11.69204 | - |
| ChIP-seq | chr2:208123257..208123677 | liver | JUND | liver | ENCSR837GTK | ENCFF804UDG | 10.18840 | - |

**Table S6. RegulomeDB annotation of the association lead variants near *CDKN1A***

The RegulomeDB probability score is ranging from 0 to 1, with 1 being most likely to be a regulatory variant. 'Ranking' refers to following evidence: 1a, eQTL + TF binding + matched TF motif + matched DNase Footprint + DNase peak; 1b, eQTL + TF binding + any motif + DNase Footprint + DNase peak; 1c, eQTL + TF binding + matched TF motif + DNase peak; 1d, eQTL + TF binding + any motif + DNase peak; 1e, eQTL + TF binding + matched TF motif; 1f, eQTL + TF binding / DNase peak; 2a, TF binding + matched TF motif + matched DNase Footprint + DNase peak; 2b, TF binding + any motif + DNase Footprint + DNase peak; 2c, TF binding + matched TF motif + DNase peak; 3a, TF binding + any motif + DNase peak; 3b, TF binding + matched TF motif; 4, TF binding + DNase peak; 5, TF binding or DNase peak; 6, Motif hit; 7, Other. Data was downloaded from RegulomeDB 02/10/2021.

| Rsids | Probability | Ranking | ChIP | DNase | Footprint | Footprint matched | IC matched max | IC max | PWM | PWM matched | QTL |
|---|---|---|---|---|---|---|---|---|---|---|---|
| rs6457932 | 0.60906 | 4 | TRUE | TRUE | FALSE | FALSE | 0.0 | 0.0 | FALSE | FALSE | FALSE |
| rs9349003 | 0.49417 | 3a | TRUE | TRUE | FALSE | FALSE | 0.0 | 0.930 | TRUE | FALSE | FALSE |
| rs9366912 | 0.55436 | 1f | TRUE | TRUE | FALSE | FALSE | 0.0 | 0.0 | FALSE | FALSE | TRUE |
| rs112297876 | 0.58955 | 5 | TRUE | FALSE | FALSE | FALSE | 0.0 | 0.0 | FALSE | FALSE | FALSE |
| rs12524435 | 0.38284 | 5 | TRUE | FALSE | TRUE | FALSE | 0.0 | 0.080 | TRUE | FALSE | FALSE |
| rs10947616 | 0.58955 | 5 | TRUE | FALSE | FALSE | FALSE | 0.0 | 0.0 | FALSE | FALSE | FALSE |
| rs4713993 | 0.13454 | 5 | FALSE | TRUE | FALSE | FALSE | 0.0 | 0.0 | FALSE | FALSE | FALSE |
| rs6915170 | 0.30943 | 6 | FALSE | FALSE | FALSE | FALSE | 0.0 | 1.170 | TRUE | FALSE | FALSE |
| rs4464789 | 0.6893 | 3a | TRUE | TRUE | FALSE | FALSE | 0.0 | 0.360 | TRUE | FALSE | FALSE |
| rs10456441 | 0.49737 | 3a | TRUE | TRUE | FALSE | FALSE | 0.0 | 1.650 | TRUE | FALSE | FALSE |
| rs10456442 | 0.60906 | 4 | TRUE | TRUE | FALSE | FALSE | 0.0 | 0.0 | FALSE | FALSE | FALSE |
| rs10456443 | 0.61652 | 2b | TRUE | TRUE | TRUE | FALSE | 0.0 | 1.890 | TRUE | FALSE | FALSE |
| rs12210094 | 0.60906 | 4 | TRUE | TRUE | FALSE | FALSE | 0.0 | 0.0 | FALSE | FALSE | FALSE |
| rs12528913 | 0.08 | 5 | TRUE | FALSE | FALSE | FALSE | 0.0 | 0.200 | TRUE | FALSE | FALSE |
| rs12530170 | 0.84573 | 2b | TRUE | TRUE | TRUE | FALSE | 0.0 | 1.970 | TRUE | FALSE | FALSE |
| rs12203953 | 0.96944 | 1a | TRUE | TRUE | TRUE | TRUE | 1.930 | 1.930 | TRUE | TRUE | TRUE |
| rs6920453 | 0.60906 | 4 | TRUE | TRUE | FALSE | FALSE | 0.0 | 0.0 | FALSE | FALSE | FALSE |
| rs75420281 | 0.60906 | 4 | TRUE | TRUE | FALSE | FALSE | 0.0 | 0.0 | FALSE | FALSE | FALSE |
| rs3176348 | 0.86655 | 2b | TRUE | TRUE | TRUE | FALSE | 0.0 | 0.990 | TRUE | FALSE | FALSE |
| rs150499251 | 0.58955 | 5 | TRUE | FALSE | FALSE | FALSE | 0.0 | 0.0 | FALSE | FALSE | FALSE |
| rs9357241 | 0.04578 | 5 | TRUE | FALSE | TRUE | FALSE | 0.0 | 0.0 | FALSE | FALSE | FALSE |
| rs6921165 | 0.60906 | 4 | TRUE | TRUE | FALSE | FALSE | 0.0 | 0.0 | FALSE | FALSE | FALSE |
| rs9394413 | 0.98162 | 5 | TRUE | FALSE | FALSE | FALSE | 0.0 | 1.490 | TRUE | FALSE | FALSE |
| rs7741267 | 0.60906 | 4 | TRUE | TRUE | FALSE | FALSE | 0.0 | 0.0 | FALSE | FALSE | FALSE |
| rs7751412 | 0.30476 | 3a | TRUE | TRUE | FALSE | FALSE | 0.0 | 0.340 | TRUE | FALSE | FALSE |

**Table S7. eQTLs for the association lead variants listed in Table 1.**

| Locus | Lead variant | Gene | Tissue | P-value | NES | Variant ID in GTEx |
|---|---|---|---|---|---|---|
| 1q43 | rs4149909 | *no significant eQTLs* | | | | |
| 1q44 | rs2183478 | *no significant eQTLs* | | | | |
| 1q44 | rs4335411 | ZNF692 | Adipose - Subcutaneous | 0,000041 | -0,075 | chr1_248897507_G_A_b38 |
| 2p14 | rs17631680 | *no significant eQTLs* | | | | |
| 2q11.2 | rs13392042 | C2orf15 | Spleen | 0,000002 | 0,31 | chr2_99454113_A_G_b38 |
| 2q11.2 | rs13392042 | C2orf15 | Whole Blood | 0,0000089 | 0,21 | chr2_99454113_A_G_b38 |
| 2q11.2 | rs13392042 | C2orf15 | Adrenal Gland | 0,000086 | -0,30 | chr2_99454113_A_G_b38 |
| 2q11.2 | rs13392042 | EIF5B | Testis | 0,0000069 | -0,26 | chr2_99454113_A_G_b38 |
| 2q11.2 | rs13392042 | EIF5B | Esophagus - Muscularis | 0,00013 | -0,14 | chr2_99454113_A_G_b38 |
| 2q11.2 | rs13392042 | LIPT1 | Skin - Sun Exposed (Lower leg) | 1,60E-25 | -0,4 | chr2_99454113_A_G_b38 |
| 2q11.2 | rs13392042 | LIPT1 | Skin - Not Sun Exposed (Suprapubic) | 1,20E-19 | -0,35 | chr2_99454113_A_G_b38 |
| 2q11.2 | rs13392042 | LIPT1 | Thyroid | 4,10E-18 | -0,32 | chr2_99454113_A_G_b38 |
| 2q11.2 | rs13392042 | LIPT1 | Artery - Tibial | 4,50E-18 | -0,29 | chr2_99454113_A_G_b38 |
| 2q11.2 | rs13392042 | LIPT1 | Muscle - Skeletal | 3,50E-17 | -0,28 | chr2_99454113_A_G_b38 |
| 2q11.2 | rs13392042 | LIPT1 | Nerve - Tibial | 9,60E-17 | -0,29 | chr2_99454113_A_G_b38 |
| 2q11.2 | rs13392042 | LIPT1 | Adipose - Subcutaneous | 1,20E-16 | -0,31 | chr2_99454113_A_G_b38 |
| 2q11.2 | rs13392042 | LIPT1 | Testis | 2,80E-16 | -0,47 | chr2_99454113_A_G_b38 |
| 2q11.2 | rs13392042 | LIPT1 | Breast - Mammary Tissue | 6,60E-15 | -0,33 | chr2_99454113_A_G_b38 |
| 2q11.2 | rs13392042 | LIPT1 | Adipose - Visceral (Omentum) | 3,50E-14 | -0,27 | chr2_99454113_A_G_b38 |
| 2q11.2 | rs13392042 | LIPT1 | Stomach | 2,90E-12 | -0,34 | chr2_99454113_A_G_b38 |
| 2q11.2 | rs13392042 | LIPT1 | Artery - Aorta | 1,10E-11 | -0,28 | chr2_99454113_A_G_b38 |
| 2q11.2 | rs13392042 | LIPT1 | Esophagus - Mucosa | 1,30E-11 | -0,26 | chr2_99454113_A_G_b38 |
| 2q11.2 | rs13392042 | LIPT1 | Colon - Sigmoid | 1,30E-11 | -0,29 | chr2_99454113_A_G_b38 |
| 2q11.2 | rs13392042 | LIPT1 | Esophagus - Muscularis | 5,70E-11 | -0,26 | chr2_99454113_A_G_b38 |
| 2q11.2 | rs13392042 | LIPT1 | Heart - Atrial Appendage | 8,20E-11 | -0,34 | chr2_99454113_A_G_b38 |
| 2q11.2 | rs13392042 | LIPT1 | Lung | 9,90E-11 | -0,18 | chr2_99454113_A_G_b38 |
| 2q11.2 | rs13392042 | LIPT1 | Colon - Transverse | 1,90E-10 | -0,28 | chr2_99454113_A_G_b38 |
| 2q11.2 | rs13392042 | LIPT1 | Esophagus - Gastroesophageal Junction | 8,20E-10 | -0,29 | chr2_99454113_A_G_b38 |
| 2q11.2 | rs13392042 | LIPT1 | Pituitary | 3,30E-09 | -0,31 | chr2_99454113_A_G_b38 |
| 2q11.2 | rs13392042 | LIPT1 | Prostate | 1,30E-08 | -0,3 | chr2_99454113_A_G_b38 |
| 2q11.2 | rs13392042 | LIPT1 | Cells - Cultured fibroblasts | 2,90E-08 | -0,19 | chr2_99454113_A_G_b38 |
| 2q11.2 | rs13392042 | LIPT1 | Adrenal Gland | 4,60E-08 | -0,47 | chr2_99454113_A_G_b38 |
| 2q11.2 | rs13392042 | LIPT1 | Whole Blood | 2,10E-07 | -0,17 | chr2_99454113_A_G_b38 |
| 2q11.2 | rs13392042 | LIPT1 | Brain - Caudate (basal ganglia) | 3,90E-07 | -0,35 | chr2_99454113_A_G_b38 |
| 2q11.2 | rs13392042 | LIPT1 | Brain - Cerebellum | 0,0000011 | -0,4 | chr2_99454113_A_G_b38 |
| 2q11.2 | rs13392042 | LIPT1 | Artery - Coronary | 0,0000017 | -0,32 | chr2_99454113_A_G_b38 |
| 2q11.2 | rs13392042 | LIPT1 | Heart - Left Ventricle | 0,0000019 | -0,25 | chr2_99454113_A_G_b38 |
| 2q11.2 | rs13392042 | LIPT1 | Kidney - Cortex | 0,0000039 | -0,48 | chr2_99454113_A_G_b38 |
| 2q11.2 | rs13392042 | LIPT1 | Liver | 0,000021 | -0,29 | chr2_99454113_A_G_b38 |
| 2q11.2 | rs13392042 | LIPT1 | Brain - Amygdala | 0,000026 | -0,41 | chr2_99454113_A_G_b38 |
| 2q11.2 | rs13392042 | LIPT1 | Brain - Nucleus accumbens (basal ganglia) | 0,00003 | -0,31 | chr2_99454113_A_G_b38 |

| Locus | Lead variant | Gene | Tissue | P-value | NES | Variant ID in GTEx |
|---|---|---|---|---|---|---|
| 2q11.2 | rs13392042 | LIPT1 | Brain - Cerebellar Hemisphere | 0,00003 | -0,32 | chr2_99454113_A_G_b38 |
| 2q11.2 | rs13392042 | LIPT1 | Brain - Frontal Cortex (BA9) | 0,000037 | -0,28 | chr2_99454113_A_G_b38 |
| 2q11.2 | rs13392042 | LYG1 | Nerve - Tibial | 1,10E-11 | 0,25 | chr2_99454113_A_G_b38 |
| 2q11.2 | rs13392042 | LYG1 | Adipose - Subcutaneous | 2,10E-08 | 0,25 | chr2_99454113_A_G_b38 |
| 2q11.2 | rs13392042 | LYG1 | Skin - Sun Exposed (Lower leg) | 1,40E-07 | 0,2 | chr2_99454113_A_G_b38 |
| 2q11.2 | rs13392042 | LYG1 | Heart - Atrial Appendage | 0,000002 | 0,22 | chr2_99454113_A_G_b38 |
| 2q11.2 | rs13392042 | LYG1 | Testis | 0,0000022 | -0,23 | chr2_99454113_A_G_b38 |
| 2q11.2 | rs13392042 | LYG1 | Breast - Mammary Tissue | 0,000017 | 0,16 | chr2_99454113_A_G_b38 |
| 2q11.2 | rs13392042 | LYG1 | Heart - Left Ventricle | 0,000026 | 0,18 | chr2_99454113_A_G_b38 |
| 2q11.2 | rs13392042 | LYG1 | Cells - Cultured fibroblasts | 0,000031 | 0,2 | chr2_99454113_A_G_b38 |
| 2q11.2 | rs13392042 | LYG1 | Colon - Transverse | 0,00018 | 0,18 | chr2_99454113_A_G_b38 |
| 2q11.2 | rs13392042 | MITD1 | Adipose - Visceral (Omentum) | 4,00E-11 | 0,2 | chr2_99454113_A_G_b38 |
| 2q11.2 | rs13392042 | MITD1 | Nerve - Tibial | 2,00E-08 | 0,21 | chr2_99454113_A_G_b38 |
| 2q11.2 | rs13392042 | MITD1 | Cells - Cultured fibroblasts | 2,70E-08 | 0,15 | chr2_99454113_A_G_b38 |
| 2q11.2 | rs13392042 | MITD1 | Heart - Atrial Appendage | 2,70E-08 | 0,26 | chr2_99454113_A_G_b38 |
| 2q11.2 | rs13392042 | MITD1 | Heart - Left Ventricle | 1,00E-07 | 0,22 | chr2_99454113_A_G_b38 |
| 2q11.2 | rs13392042 | MITD1 | Artery - Aorta | 1,60E-07 | 0,22 | chr2_99454113_A_G_b38 |
| 2q11.2 | rs13392042 | MITD1 | Breast - Mammary Tissue | 1,60E-07 | 0,21 | chr2_99454113_A_G_b38 |
| 2q11.2 | rs13392042 | MITD1 | Stomach | 3,20E-07 | 0,21 | chr2_99454113_A_G_b38 |
| 2q11.2 | rs13392042 | MITD1 | Adipose - Subcutaneous | 0,0000021 | 0,17 | chr2_99454113_A_G_b38 |
| 2q11.2 | rs13392042 | MITD1 | Esophagus - Mucosa | 0,0000042 | 0,15 | chr2_99454113_A_G_b38 |
| 2q11.2 | rs13392042 | MITD1 | Artery - Tibial | 0,000021 | 0,13 | chr2_99454113_A_G_b38 |
| 2q11.2 | rs13392042 | MITD1 | Muscle - Skeletal | 0,000037 | 0,13 | chr2_99454113_A_G_b38 |
| 2q11.2 | rs13392042 | MITD1 | Colon - Transverse | 0,000038 | 0,16 | chr2_99454113_A_G_b38 |
| 2q11.2 | rs13392042 | MITD1 | Brain - Frontal Cortex (BA9) | 0,000058 | 0,22 | chr2_99454113_A_G_b38 |
| 2q11.2 | rs13392042 | MITD1 | Whole Blood | 0,000087 | 0,09 | chr2_99454113_A_G_b38 |
| 2q11.2 | rs13392042 | MITD1 | Skin - Sun Exposed (Lower leg) | 0,00016 | 0,1 | chr2_99454113_A_G_b38 |
| 2q11.2 | rs13392042 | MRPL30 | Thyroid | 6,50E-09 | -0,18 | chr2_99454113_A_G_b38 |
| 2q11.2 | rs13392042 | MRPL30 | Colon - Transverse | 0,00002 | -0,16 | chr2_99454113_A_G_b38 |
| 2q11.2 | rs13392042 | MRPL30 | Artery - Tibial | 0,000063 | -0,1 | chr2_99454113_A_G_b38 |
| 2q11.2 | rs13392042 | REV1 | Pancreas | 2,40E-20 | -0,44 | chr2_99454113_A_G_b38 |
| 2q11.2 | rs13392042 | REV1 | Testis | 3,10E-16 | 0,23 | chr2_99454113_A_G_b38 |
| 2q11.2 | rs13392042 | REV1 | Thyroid | 1,10E-14 | -0,2 | chr2_99454113_A_G_b38 |
| 2q11.2 | rs13392042 | REV1 | Cells - Cultured fibroblasts | 4,20E-13 | -0,23 | chr2_99454113_A_G_b38 |
| 2q11.2 | rs13392042 | REV1 | Whole Blood | 2,40E-09 | -0,11 | chr2_99454113_A_G_b38 |
| 2q11.2 | rs13392042 | REV1 | Esophagus - Mucosa | 3,00E-09 | 0,15 | chr2_99454113_A_G_b38 |
| 2q11.2 | rs13392042 | REV1 | Adipose - Subcutaneous | 5,60E-07 | -0,13 | chr2_99454113_A_G_b38 |
| 2q11.2 | rs13392042 | REV1 | Spleen | 0,0000061 | -0,21 | chr2_99454113_A_G_b38 |
| 2q11.2 | rs13392042 | REV1 | Skin - Not Sun Exposed (Suprapubic) | 0,000013 | 0,12 | chr2_99454113_A_G_b38 |
| 2q11.2 | rs13392042 | REV1 | Pituitary | 0,00008 | -0,2 | chr2_99454113_A_G_b38 |
| 2q11.2 | rs13392042 | RP11-527J8.1 | Skin - Not Sun Exposed (Suprapubic) | 5,00E-07 | 0,18 | chr2_99454113_A_G_b38 |
| 2q11.2 | rs13392042 | RP11-527J8.1 | Nerve - Tibial | 8,00E-07 | 0,2 | chr2_99454113_A_G_b38 |
| 2q11.2 | rs13392042 | RP11-527J8.1 | Esophagus - Mucosa | 0,0000011 | 0,22 | chr2_99454113_A_G_b38 |

| Locus | Lead variant | Gene | Tissue | P-value | NES | Variant ID in GTEx |
|---|---|---|---|---|---|---|
| 2q11.2 | rs13392042 | RP11-527J8.1 | Artery - Tibial | 0,000013 | 0,17 | chr2_99454113_A_G_b38 |
| 2q11.2 | rs13392042 | RP11-527J8.1 | Skin - Sun Exposed (Lower leg) | 0,000014 | 0,17 | chr2_99454113_A_G_b38 |
| 2q11.2 | rs13392042 | RP11-527J8.1 | Adipose - Visceral (Omentum) | 0,00002 | 0,18 | chr2_99454113_A_G_b38 |
| 2q11.2 | rs13392042 | RP11-527J8.1 | Lung | 0,000048 | 0,16 | chr2_99454113_A_G_b38 |
| 2q11.2 | rs13392042 | RP11-527J8.1 | Colon - Transverse | 0,000071 | 0,19 | chr2_99454113_A_G_b38 |
| 2q11.2 | rs13392042 | RP11-527J8.1 | Testis | 0,000077 | 0,25 | chr2_99454113_A_G_b38 |
| 2q11.2 | rs13392042 | RP11-527J8.1 | Thyroid | 0,00031 | 0,13 | chr2_99454113_A_G_b38 |
| 2q11.2 | rs13392042 | TSGA10 | Thyroid | 9,50E-34 | -0,43 | chr2_99454113_A_G_b38 |
| 2q11.2 | rs13392042 | TSGA10 | Skin - Not Sun Exposed (Suprapubic) | 1,10E-22 | -0,38 | chr2_99454113_A_G_b38 |
| 2q11.2 | rs13392042 | TSGA10 | Esophagus - Mucosa | 2,20E-20 | -0,43 | chr2_99454113_A_G_b38 |
| 2q11.2 | rs13392042 | TSGA10 | Skin - Sun Exposed (Lower leg) | 4,30E-18 | -0,28 | chr2_99454113_A_G_b38 |
| 2q11.2 | rs13392042 | TSGA10 | Pancreas | 2,00E-17 | -0,55 | chr2_99454113_A_G_b38 |
| 2q11.2 | rs13392042 | TSGA10 | Whole Blood | 1,00E-15 | -0,35 | chr2_99454113_A_G_b38 |
| 2q11.2 | rs13392042 | TSGA10 | Spleen | 2,60E-12 | -0,57 | chr2_99454113_A_G_b38 |
| 2q11.2 | rs13392042 | TSGA10 | Pituitary | 5,40E-12 | -0,35 | chr2_99454113_A_G_b38 |
| 2q11.2 | rs13392042 | TSGA10 | Adrenal Gland | 4,30E-11 | -0,5 | chr2_99454113_A_G_b38 |
| 2q11.2 | rs13392042 | TSGA10 | Brain - Cerebellar Hemisphere | 6,70E-09 | -0,59 | chr2_99454113_A_G_b38 |
| 2q11.2 | rs13392042 | TSGA10 | Colon - Transverse | 8,10E-09 | -0,2 | chr2_99454113_A_G_b38 |
| 2q11.2 | rs13392042 | TSGA10 | Cells - EBV-transformed lymphocytes | 1,60E-08 | -0,52 | chr2_99454113_A_G_b38 |
| 2q11.2 | rs13392042 | TSGA10 | Lung | 4,90E-08 | -0,14 | chr2_99454113_A_G_b38 |
| 2q11.2 | rs13392042 | TSGA10 | Breast - Mammary Tissue | 7,50E-08 | -0,23 | chr2_99454113_A_G_b38 |
| 2q11.2 | rs13392042 | TSGA10 | Stomach | 9,80E-08 | -0,2 | chr2_99454113_A_G_b38 |
| 2q11.2 | rs13392042 | TSGA10 | Small Intestine - Terminal Ileum | 1,20E-07 | -0,35 | chr2_99454113_A_G_b38 |
| 2q11.2 | rs13392042 | TSGA10 | Prostate | 2,00E-07 | -0,27 | chr2_99454113_A_G_b38 |
| 2q11.2 | rs13392042 | TSGA10 | Brain - Cerebellum | 2,10E-07 | -0,53 | chr2_99454113_A_G_b38 |
| 2q11.2 | rs13392042 | TSGA10 | Liver | 0,0000018 | -0,39 | chr2_99454113_A_G_b38 |
| 2q11.2 | rs13392042 | TSGA10 | Brain - Cortex | 0,0000025 | -0,35 | chr2_99454113_A_G_b38 |
| 2q11.2 | rs13392042 | TSGA10 | Brain - Putamen (basal ganglia) | 0,000011 | -0,43 | chr2_99454113_A_G_b38 |
| 2q11.2 | rs13392042 | TSGA10 | Brain - Nucleus accumbens (basal ganglia) | 0,000022 | -0,3 | chr2_99454113_A_G_b38 |
| 2q11.2 | rs13392042 | TSGA10 | Testis | 0,000025 | -0,081 | chr2_99454113_A_G_b38 |
| 2q11.2 | rs13392042 | TSGA10 | Brain - Frontal Cortex (BA9) | 0,00003 | -0,36 | chr2_99454113_A_G_b38 |
| 2q11.2 | rs13392042 | TSGA10 | Adipose - Visceral (Omentum) | 0,000061 | -0,14 | chr2_99454113_A_G_b38 |
| 2q11.2 | rs13392042 | TSGA10 | Artery - Tibial | 0,00015 | -0,18 | chr2_99454113_A_G_b38 |
| 2q11.2 | rs13392042 | TXNDC9 | Testis | 1,90E-22 | -0,48 | chr2_99454113_A_G_b38 |
| 2q11.2 | rs13392042 | TXNDC9 | Muscle - Skeletal | 0,000034 | -0,098 | chr2_99454113_A_G_b38 |
| 2q33.3 | rs10804157 | *no significant eQTLs* | | | | |
| 2q37.3 | rs34766121 | AC114730.11 | Thyroid | 1,40E-45 | -0,5 | chr2_241720139_C_T_b38 |
| 2q37.3 | rs34766121 | AC114730.11 | Skin - Sun Exposed (Lower leg) | 1,40E-15 | -0,32 | chr2_241720139_C_T_b38 |
| 2q37.3 | rs34766121 | AC114730.11 | Pituitary | 2,90E-11 | -0,4 | chr2_241720139_C_T_b38 |
| 2q37.3 | rs34766121 | AC114730.11 | Skin - Not Sun Exposed (Suprapubic) | 1,10E-09 | -0,26 | chr2_241720139_C_T_b38 |
| 2q37.3 | rs34766121 | AC114730.11 | Nerve - Tibial | 8,40E-09 | -0,2 | chr2_241720139_C_T_b38 |
| 2q37.3 | rs34766121 | AC114730.11 | Lung | 1,30E-08 | -0,21 | chr2_241720139_C_T_b38 |
| 2q37.3 | rs34766121 | AC114730.11 | Artery - Tibial | 8,40E-08 | -0,2 | chr2_241720139_C_T_b38 |

| Locus | Lead variant | Gene | Tissue | P-value | NES | Variant ID in GTEx |
|---|---|---|---|---|---|---|
| 2q37.3 | rs34766121 | AC114730.11 | Testis | 2,90E-07 | -0,34 | chr2_241720139_C_T_b38 |
| 2q37.3 | rs34766121 | AC114730.11 | Esophagus - Muscularis | 0,000001 | -0,19 | chr2_241720139_C_T_b38 |
| 2q37.3 | rs34766121 | AC114730.11 | Breast - Mammary Tissue | 0,0000022 | -0,22 | chr2_241720139_C_T_b38 |
| 2q37.3 | rs34766121 | AC114730.11 | Heart - Atrial Appendage | 0,0000066 | -0,21 | chr2_241720139_C_T_b38 |
| 2q37.3 | rs34766121 | AC114730.11 | Colon - Transverse | 0,0000066 | -0,24 | chr2_241720139_C_T_b38 |
| 2q37.3 | rs34766121 | AC114730.11 | Muscle - Skeletal | 0,000018 | -0,19 | chr2_241720139_C_T_b38 |
| 2q37.3 | rs34766121 | AC114730.11 | Adipose - Subcutaneous | 0,000054 | -0,18 | chr2_241720139_C_T_b38 |
| 2q37.3 | rs34766121 | AC114730.11 | Brain - Cerebellum | 0,000091 | -0,3 | chr2_241720139_C_T_b38 |
| 2q37.3 | rs34766121 | BOK | Adipose - Subcutaneous | 8,10E-09 | -0,1 | chr2_241720139_C_T_b38 |
| 2q37.3 | rs34766121 | BOK | Whole Blood | 0,000043 | 0,16 | chr2_241720139_C_T_b38 |
| 2q37.3 | rs34766121 | DTYMK | Adipose - Subcutaneous | 4,30E-07 | 0,14 | chr2_241720139_C_T_b38 |
| 2q37.3 | rs34766121 | DTYMK | Testis | 0,000001 | 0,18 | chr2_241720139_C_T_b38 |
| 2q37.3 | rs34766121 | DTYMK | Cells - Cultured fibroblasts | 0,0000012 | 0,081 | chr2_241720139_C_T_b38 |
| 2q37.3 | rs34766121 | DTYMK | Nerve - Tibial | 0,0000037 | 0,12 | chr2_241720139_C_T_b38 |
| 2q37.3 | rs34766121 | DTYMK | Artery - Tibial | 0,000014 | 0,12 | chr2_241720139_C_T_b38 |
| 2q37.3 | rs34766121 | DTYMK | Brain - Nucleus accumbens (basal ganglia) | 0,000014 | 0,19 | chr2_241720139_C_T_b38 |
| 2q37.3 | rs34766121 | DTYMK | Brain - Hypothalamus | 0,000026 | 0,24 | chr2_241720139_C_T_b38 |
| 2q37.3 | rs34766121 | DTYMK | Brain - Cortex | 0,000053 | 0,25 | chr2_241720139_C_T_b38 |
| 2q37.3 | rs34766121 | DTYMK | Artery - Aorta | 0,00013 | 0,14 | chr2_241720139_C_T_b38 |
| 2q37.3 | rs34766121 | FARP2 | Adipose - Subcutaneous | 0,000099 | -0,15 | chr2_241720139_C_T_b38 |
| 2q37.3 | rs34766121 | ING5 | Thyroid | 2,20E-39 | -0,46 | chr2_241720139_C_T_b38 |
| 2q37.3 | rs34766121 | ING5 | Pituitary | 2,60E-14 | -0,39 | chr2_241720139_C_T_b38 |
| 2q37.3 | rs34766121 | ING5 | Testis | 6,50E-12 | -0,26 | chr2_241720139_C_T_b38 |
| 2q37.3 | rs34766121 | ING5 | Nerve - Tibial | 3,80E-11 | -0,18 | chr2_241720139_C_T_b38 |
| 2q37.3 | rs34766121 | ING5 | Lung | 5,10E-10 | -0,2 | chr2_241720139_C_T_b38 |
| 2q37.3 | rs34766121 | ING5 | Adipose - Visceral (Omentum) | 5,70E-09 | -0,19 | chr2_241720139_C_T_b38 |
| 2q37.3 | rs34766121 | ING5 | Skin - Not Sun Exposed (Suprapubic) | 1,30E-08 | -0,16 | chr2_241720139_C_T_b38 |
| 2q37.3 | rs34766121 | ING5 | Esophagus - Muscularis | 1,50E-08 | -0,15 | chr2_241720139_C_T_b38 |
| 2q37.3 | rs34766121 | ING5 | Artery - Tibial | 6,00E-07 | -0,13 | chr2_241720139_C_T_b38 |
| 2q37.3 | rs34766121 | ING5 | Skin - Sun Exposed (Lower leg) | 9,10E-07 | -0,14 | chr2_241720139_C_T_b38 |
| 2q37.3 | rs34766121 | ING5 | Prostate | 0,0000012 | -0,26 | chr2_241720139_C_T_b38 |
| 2q37.3 | rs34766121 | ING5 | Adipose - Subcutaneous | 0,0000056 | -0,14 | chr2_241720139_C_T_b38 |
| 2q37.3 | rs34766121 | ING5 | Adrenal Gland | 0,000026 | -0,22 | chr2_241720139_C_T_b38 |
| 2q37.3 | rs34766121 | ING5 | Spleen | 0,00003 | -0,3 | chr2_241720139_C_T_b38 |
| 2q37.3 | rs34766121 | ING5 | Artery - Aorta | 0,00004 | -0,13 | chr2_241720139_C_T_b38 |
| 2q37.3 | rs34766121 | ING5 | Heart - Atrial Appendage | 0,000067 | -0,15 | chr2_241720139_C_T_b38 |
| 2q37.3 | rs34766121 | ING5 | Esophagus - Mucosa | 0,000083 | -0,11 | chr2_241720139_C_T_b38 |
| 2q37.3 | rs34766121 | THAP4 | Thyroid | 0,00018 | -0,12 | chr2_241720139_C_T_b38 |
| 3p26.1 | rs3804984 | ITPR | Esophagus - Mucosa | 0,000051 | 0,098 | chr3_4674530_T_C_b38 |
| 3p24.2 | rs1010961 | THRB | Artery - Aorta | 0,0000084 | -0,099 | chr3_24213259_T_A_b38 |
| 3q27.2 | rs13060777 | IGF2BP2 | Thyroid | 5,30E-08 | 0,22 | chr3_185807411_A_G_b38 |
| 4q23 | rs1037475 | ADH4 | Spleen | 1,20E-08 | -0,45 | chr4_99031559_A_G_b38 |
| 4q23 | rs1037475 | ADH4 | Esophagus - Gastroesophageal Junction | 8,30E-07 | 0,31 | chr4_99031559_A_G_b38 |

| Locus | Lead variant | Gene | Tissue | P-value | NES | Variant ID in GTEx |
|---|---|---|---|---|---|---|
| 4q23 | rs1037475 | ADH4 | Small Intestine - Terminal Ileum | 0,0000016 | -0,44 | chr4_99031559_A_G_b38 |
| 4q23 | rs1037475 | ADH4 | Esophagus - Muscularis | 0,0000053 | 0,24 | chr4_99031559_A_G_b38 |
| 4q23 | rs1037475 | ADH4 | Esophagus - Mucosa | 0,000006 | 0,24 | chr4_99031559_A_G_b38 |
| 4q23 | rs1037475 | ADH4 | Muscle - Skeletal | 0,000058 | 0,17 | chr4_99031559_A_G_b38 |
| 4q23 | rs1037475 | ADH4 | Cells - Cultured fibroblasts | 0,000073 | -0,21 | chr4_99031559_A_G_b38 |
| 4q23 | rs1037475 | ADH5 | Muscle - Skeletal | 9,50E-08 | -0,17 | chr4_99031559_A_G_b38 |
| 4q23 | rs1037475 | ADH5 | Whole Blood | 4,90E-07 | -0,11 | chr4_99031559_A_G_b38 |
| 4q23 | rs1037475 | ADH5 | Esophagus - Mucosa | 0,000043 | -0,08 | chr4_99031559_A_G_b38 |
| 4q23 | rs1037475 | ADH6 | Esophagus - Muscularis | 0,00022 | -0,18 | chr4_99031559_A_G_b38 |
| 4q23 | rs1037475 | EIF4E | Esophagus - Mucosa | 4,30E-09 | 0,11 | chr4_99031559_A_G_b38 |
| 4q23 | rs1037475 | EIF4E | Cells - Cultured fibroblasts | 0,0002 | 0,047 | chr4_99031559_A_G_b38 |
| 4q23 | rs1037475 | METAP1 | Artery - Tibial | 1,90E-09 | 0,16 | chr4_99031559_A_G_b38 |
| 4q23 | rs1037475 | METAP1 | Colon - Sigmoid | 0,0000013 | 0,19 | chr4_99031559_A_G_b38 |
| 4q23 | rs1037475 | METAP1 | Artery - Aorta | 0,0000028 | 0,21 | chr4_99031559_A_G_b38 |
| 4q23 | rs1037475 | METAP1 | Adipose - Subcutaneous | 0,000015 | 0,14 | chr4_99031559_A_G_b38 |
| 4q23 | rs1037475 | MTTP | Thyroid | 0,0002 | 0,21 | chr4_99031559_A_G_b38 |
| 4q23 | rs1037475 | RP11-571L19.8 | Liver | 0,000012 | -0,28 | chr4_99031559_A_G_b38 |
| 4q23 | rs1037475 | RP11-571L19.8 | Adipose - Visceral (Omentum) | 0,000089 | -0,19 | chr4_99031559_A_G_b38 |
| 4q23 | rs1037475 | RP11-571L19.8 | Muscle - Skeletal | 0,00016 | -0,17 | chr4_99031559_A_G_b38 |
| 4q23 | rs1037475 | RP11-696N14.1 | Adipose - Subcutaneous | 8,80E-09 | 0,25 | chr4_99031559_A_G_b38 |
| 4q23 | rs1037475 | RP11-696N14.1 | Skin - Sun Exposed (Lower leg) | 2,50E-08 | 0,2 | chr4_99031559_A_G_b38 |
| 4q23 | rs1037475 | RP11-696N14.1 | Skin - Not Sun Exposed (Suprapubic) | 7,90E-07 | 0,21 | chr4_99031559_A_G_b38 |
| 4q23 | rs1037475 | RP11-696N14.1 | Lung | 0,000016 | 0,18 | chr4_99031559_A_G_b38 |
| 4q23 | rs1037475 | RP11-696N14.1 | Pancreas | 0,000057 | 0,25 | chr4_99031559_A_G_b38 |
| 4q23 | rs1037475 | RP11-696N14.1 | Colon - Transverse | 0,000076 | 0,23 | chr4_99031559_A_G_b38 |
| 5q31.1 | rs4367292 | HSPA4 | Muscle - Skeletal | 3,90E-21 | 0,26 | chr5_133099880_C_T_b38 |
| 5q31.1 | rs4367292 | HSPA4 | Nerve - Tibial | 1,40E-14 | 0,17 | chr5_133099880_C_T_b38 |
| 5q31.1 | rs4367292 | HSPA4 | Testis | 3,30E-14 | 0,25 | chr5_133099880_C_T_b38 |
| 5q31.1 | rs4367292 | HSPA4 | Brain - Nucleus accumbens (basal ganglia) | 1,00E-09 | 0,28 | chr5_133099880_C_T_b38 |
| 5q31.1 | rs4367292 | HSPA4 | Thyroid | 8,50E-09 | 0,1 | chr5_133099880_C_T_b38 |
| 5q31.1 | rs4367292 | HSPA4 | Brain - Cortex | 7,10E-08 | 0,25 | chr5_133099880_C_T_b38 |
| 5q31.1 | rs4367292 | HSPA4 | Whole Blood | 1,90E-07 | -0,13 | chr5_133099880_C_T_b38 |
| 5q31.1 | rs4367292 | HSPA4 | Brain - Cerebellum | 3,00E-07 | 0,25 | chr5_133099880_C_T_b38 |
| 5q31.1 | rs4367292 | HSPA4 | Brain - Caudate (basal ganglia) | 3,20E-07 | 0,3 | chr5_133099880_C_T_b38 |
| 5q31.1 | rs4367292 | HSPA4 | Brain - Cerebellar Hemisphere | 0,0000037 | 0,24 | chr5_133099880_C_T_b38 |
| 5q31.1 | rs4367292 | HSPA4 | Brain - Putamen (basal ganglia) | 0,000008 | 0,27 | chr5_133099880_C_T_b38 |
| 5q31.1 | rs4367292 | HSPA4 | Colon - Sigmoid | 0,000095 | 0,11 | chr5_133099880_C_T_b38 |
| 6p21.2 | rs10456443 | ETV7 | Whole Blood | 3,60E-09 | -0,23 | chr6_36653670_G_A_b38 |
| 6p21.2 | rs10456443 | RAB44 | Cells - Cultured fibroblasts | 0,0000011 | 0,28 | chr6_36653670_G_A_b38 |
| 6p21.2 | rs10456443 | RAB44 | Whole Blood | 0,0000094 | 0,12 | chr6_36653670_G_A_b38 |
| 6p21.2 | rs10456443 | RP1-50J22.4 | Testis | 0,000094 | -0,34 | chr6_36653670_G_A_b38 |
| 6q21 | rs11153158 | ARMC2 | Testis | 1,30E-07 | -0,17 | chr6_109054915_T_C_b38 |
| 6q21 | rs11153158 | CCDC162P | Thyroid | 1,60E-07 | -0,39 | chr6_109054915_T_C_b38 |

| Locus | Lead variant | Gene | Tissue | P-value | NES | Variant ID in GTEx |
|---|---|---|---|---|---|---|
| 6q21 | rs11153158 | CCDC162P | Esophagus - Muscularis | 0,000003 | -0,38 | chr6_109054915_T_C_b38 |
| 6q21 | rs11153158 | CCDC162P | Colon - Sigmoid | 0,000028 | -0,38 | chr6_109054915_T_C_b38 |
| 6q21 | rs11153158 | CCDC162P | Nerve - Tibial | 0,000068 | -0,3 | chr6_109054915_T_C_b38 |
| 6q21 | rs11153158 | CD164 | Muscle - Skeletal | 0,0000018 | 0,2 | chr6_109054915_T_C_b38 |
| 6q21 | rs11153158 | RP11-787I22.3 | Testis | 1,40E-08 | 0,53 | chr6_109054915_T_C_b38 |
| 6q21 | rs11153158 | RP11-787I22.3 | Skin - Sun Exposed (Lower leg) | 0,00032 | 0,16 | chr6_109054915_T_C_b38 |
| 6q21 | rs11153158 | SESN1 | Prostate | 0,0000019 | 0,41 | chr6_109054915_T_C_b38 |
| 6q21 | rs11153158 | SESN1 | Esophagus - Muscularis | 0,0000099 | 0,16 | chr6_109054915_T_C_b38 |
| 6q21 | rs11153158 | SESN1 | Esophagus - Mucosa | 0,000027 | 0,21 | chr6_109054915_T_C_b38 |
| 6q21 | rs11153158 | SESN1 | Artery - Tibial | 0,00021 | 0,12 | chr6_109054915_T_C_b38 |
| 7p14.3 | rs4723230 | BBS9 | Thyroid | 0,00011 | -0,15 | chr7_33008785_C_T_b38 |
| 7p14.3 | rs4723230 | NT5C3A | Thyroid | 1,40E-35 | -0,6 | chr7_33008785_C_T_b38 |
| 7p14.3 | rs4723230 | NT5C3A | Pituitary | 2,00E-08 | -0,46 | chr7_33008785_C_T_b38 |
| 7p14.3 | rs4723230 | NT5C3A | Brain - Cerebellum | 7,90E-07 | -0,4 | chr7_33008785_C_T_b38 |
| 7p14.3 | rs4723230 | NT5C3A | Liver | 0,0000038 | -0,41 | chr7_33008785_C_T_b38 |
| 7p14.3 | rs4723230 | NT5C3A | Lung | 0,000015 | -0,19 | chr7_33008785_C_T_b38 |
| 7p14.3 | rs4723230 | RP9P | Nerve - Tibial | 0,00013 | -0,18 | chr7_33008785_C_T_b38 |
| 7q31.31 | rs12706314 | CPED1 | Cells - Cultured fibroblasts | 1,40E-12 | 0,32 | chr7_121132432_G_A_b38 |
| 7q31.31 | rs12706314 | CPED1 | Esophagus - Muscularis | 0,000093 | 0,11 | chr7_121132432_G_A_b38 |
| 7q31.31 | rs12706314 | WNT16 | Adipose - Subcutaneous | 0,0000044 | 0,21 | chr7_121132432_G_A_b38 |
| 7q31.31 | rs12706314 | WNT16 | Brain - Putamen (basal ganglia) | 0,000014 | -0,3 | chr7_121132432_G_A_b38 |
| 7q32.3 | rs35908158 | *no significant eQTLs* | | | | |
| 8p12 | rs13275869 | GTF2E2 | Esophagus - Muscularis | 2,40E-12 | -0,17 | chr8_30452819_T_C_b38 |
| 8p12 | rs13275869 | GTF2E2 | Artery - Tibial | 0,00011 | -0,069 | chr8_30452819_T_C_b38 |
| 8p12 | rs13275869 | PPP2CB | Whole Blood | 0,000033 | -0,097 | chr8_30452819_T_C_b38 |
| 8p12 | rs13275869 | RBPMS | Brain - Cerebellum | 0,00002 | 0,23 | chr8_30452819_T_C_b38 |
| 8p12 | rs13275869 | SMIM18 | Cells - EBV-transformed lymphocytes | 0,00003 | 0,43 | chr8_30452819_T_C_b38 |
| 8q24.21 | rs1516980 | *no significant eQTLs* | | | | |
| 9q22.2 | rs28508285 | UNQ6494 | Muscle - Skeletal | 1,00E-63 | -0,68 | chr9_89639982_A_G_b38 |
| 9q22.2 | rs28508285 | UNQ6494 | Artery - Tibial | 4,20E-39 | -0,63 | chr9_89639982_A_G_b38 |
| 9q22.2 | rs28508285 | UNQ6494 | Esophagus - Muscularis | 5,10E-20 | -0,47 | chr9_89639982_A_G_b38 |
| 9q22.2 | rs28508285 | UNQ6494 | Adipose - Subcutaneous | 4,30E-17 | -0,52 | chr9_89639982_A_G_b38 |
| 9q22.2 | rs28508285 | UNQ6494 | Nerve - Tibial | 1,00E-16 | -0,43 | chr9_89639982_A_G_b38 |
| 9q22.2 | rs28508285 | UNQ6494 | Esophagus - Gastroesophageal Junction | 4,40E-13 | -0,48 | chr9_89639982_A_G_b38 |
| 9q22.2 | rs28508285 | UNQ6494 | Heart - Atrial Appendage | 2,60E-12 | -0,55 | chr9_89639982_A_G_b38 |
| 9q22.2 | rs28508285 | UNQ6494 | Artery - Aorta | 8,70E-10 | -0,44 | chr9_89639982_A_G_b38 |
| 9q22.2 | rs28508285 | UNQ6494 | Lung | 1,80E-09 | -0,36 | chr9_89639982_A_G_b38 |
| 9q22.2 | rs28508285 | UNQ6494 | Artery - Coronary | 1,60E-07 | -0,51 | chr9_89639982_A_G_b38 |
| 9q22.2 | rs28508285 | UNQ6494 | Uterus | 2,90E-07 | -0,78 | chr9_89639982_A_G_b38 |
| 9q22.2 | rs28508285 | UNQ6494 | Testis | 5,80E-07 | -0,53 | chr9_89639982_A_G_b38 |
| 9q22.2 | rs28508285 | UNQ6494 | Skin - Not Sun Exposed (Suprapubic) | 0,000057 | -0,26 | chr9_89639982_A_G_b38 |
| 9q22.2 | rs28508285 | UNQ6494 | Skin - Sun Exposed (Lower leg) | 0,000075 | -0,23 | chr9_89639982_A_G_b38 |
| 10p12.31 | rs946711 | CASC10 | Skin - Sun Exposed (Lower leg) | 3,70E-07 | 0,16 | chr10_21517903_A_C_b38 |

| Locus | Lead variant | Gene | Tissue | P-value | NES | Variant ID in GTEx |
|---|---|---|---|---|---|---|
| 10p12.31 | rs946711 | CASC10 | Brain - Cerebellum | 0,0000099 | 0,28 | chr10_21517903_A_C_b38 |
| 10p12.31 | rs946711 | CASC10 | Esophagus - Muscularis | 0,00002 | 0,18 | chr10_21517903_A_C_b38 |
| 10p12.31 | rs946711 | CASC10 | Nerve - Tibial | 0,00014 | 0,093 | chr10_21517903_A_C_b38 |
| 10p12.31 | rs946711 | CASC10 | Artery - Tibial | 0,00028 | 0,12 | chr10_21517903_A_C_b38 |
| 10p12.31 | rs946711 | MLLT10 | Skin - Sun Exposed (Lower leg) | 0,000021 | -0,1 | chr10_21517903_A_C_b38 |
| 10p12.31 | rs946711 | MLLT10 | Thyroid | 0,00028 | 0,12 | chr10_21517903_A_C_b38 |
| 10q23.31 | rs1426619 | ANKRD22 | Esophagus - Mucosa | 0,000048 | 0,13 | chr10_88331783_C_T_b38 |
| 11q23.2 | rs10891420 | NCAM1 | Muscle - Skeletal | 2,50E-07 | -0,17 | chr11_112703765_T_C_b38 |
| 11q23.2 | rs10891420 | NCAM1 | Skin - Sun Exposed (Lower leg) | 0,0000069 | -0,16 | chr11_112703765_T_C_b38 |
| 11q23.2 | rs10891420 | NCAM1 | Artery - Aorta | 0,000057 | -0,22 | chr11_112703765_T_C_b38 |
| 11q23.2 | rs10891420 | RP11-356J5.12 | Muscle - Skeletal | 0,000013 | -0,1 | chr11_112703765_T_C_b38 |
| 15q23 | rs12148374 | CALML4 | Nerve - Tibial | 0,00017 | 0,13 | chr15_67922458_T_C_b38 |
| 15q23 | rs12148374 | IQCH | Esophagus - Mucosa | 0,00021 | 0,22 | chr15_67922458_T_C_b38 |
| 15q23 | rs12148374 | PIAS1 | Esophagus - Muscularis | 1,50E-15 | 0,22 | chr15_67922458_T_C_b38 |
| 15q23 | rs12148374 | PIAS1 | Artery - Tibial | 4,60E-14 | 0,17 | chr15_67922458_T_C_b38 |
| 15q23 | rs12148374 | PIAS1 | Nerve - Tibial | 7,70E-14 | 0,19 | chr15_67922458_T_C_b38 |
| 15q23 | rs12148374 | PIAS1 | Artery - Aorta | 2,10E-09 | 0,17 | chr15_67922458_T_C_b38 |
| 15q23 | rs12148374 | PIAS1 | Muscle - Skeletal | 2,80E-09 | 0,14 | chr15_67922458_T_C_b38 |
| 15q23 | rs12148374 | PIAS1 | Esophagus - Gastroesophageal Junction | 3,30E-07 | 0,18 | chr15_67922458_T_C_b38 |
| 15q23 | rs12148374 | PIAS1 | Heart - Atrial Appendage | 4,40E-07 | 0,13 | chr15_67922458_T_C_b38 |
| 15q23 | rs12148374 | PIAS1 | Colon - Sigmoid | 0,000002 | 0,12 | chr15_67922458_T_C_b38 |
| 15q23 | rs12148374 | PIAS1 | Adipose - Visceral (Omentum) | 0,0000025 | 0,15 | chr15_67922458_T_C_b38 |
| 15q23 | rs12148374 | PIAS1 | Testis | 0,000017 | 0,16 | chr15_67922458_T_C_b38 |
| 15q23 | rs12148374 | PIAS1 | Adipose - Subcutaneous | 0,000051 | 0,12 | chr15_67922458_T_C_b38 |
| 15q23 | rs12148374 | RP11-315D16.4 | Thyroid | 0,0000017 | 0,27 | chr15_67922458_T_C_b38 |
| 15q23 | rs12148374 | RP11-315D16.4 | Esophagus - Mucosa | 0,0000045 | 0,27 | chr15_67922458_T_C_b38 |
| 15q23 | rs12148374 | RP11-315D16.4 | Breast - Mammary Tissue | 0,000017 | 0,3 | chr15_67922458_T_C_b38 |
| 15q23 | rs12148374 | RP11-315D16.4 | Colon - Sigmoid | 0,000096 | 0,28 | chr15_67922458_T_C_b38 |
| 15q23 | rs12148374 | RP11-315D16.4 | Esophagus - Muscularis | 0,00013 | 0,24 | chr15_67922458_T_C_b38 |
| 15q23 | rs12148374 | RP11-315D16.4 | Nerve - Tibial | 0,00016 | 0,22 | chr15_67922458_T_C_b38 |
| 16q12.1 | rs12599260 | CNEP1R1 | Skin - Sun Exposed (Lower leg) | 0,00002 | -0,12 | chr16_50059327_G_A_b38 |
| 16q12.1 | rs12599260 | HEATR3 | Lung | 1,40E-114 | 0,7 | chr16_50059327_G_A_b38 |
| 16q12.1 | rs12599260 | HEATR3 | Nerve - Tibial | 1,00E-112 | 0,82 | chr16_50059327_G_A_b38 |
| 16q12.1 | rs12599260 | HEATR3 | Thyroid | 3,30E-111 | 0,71 | chr16_50059327_G_A_b38 |
| 16q12.1 | rs12599260 | HEATR3 | Artery - Tibial | 7,00E-102 | 0,65 | chr16_50059327_G_A_b38 |
| 16q12.1 | rs12599260 | HEATR3 | Esophagus - Muscularis | 7,80E-92 | 0,79 | chr16_50059327_G_A_b38 |
| 16q12.1 | rs12599260 | HEATR3 | Adipose - Subcutaneous | 3,80E-91 | 0,57 | chr16_50059327_G_A_b38 |
| 16q12.1 | rs12599260 | HEATR3 | Whole Blood | 1,90E-77 | 0,43 | chr16_50059327_G_A_b38 |
| 16q12.1 | rs12599260 | HEATR3 | Artery - Aorta | 5,70E-75 | 0,64 | chr16_50059327_G_A_b38 |
| 16q12.1 | rs12599260 | HEATR3 | Esophagus - Gastroesophageal Junction | 1,10E-69 | 0,81 | chr16_50059327_G_A_b38 |
| 16q12.1 | rs12599260 | HEATR3 | Testis | 5,60E-65 | 0,87 | chr16_50059327_G_A_b38 |
| 16q12.1 | rs12599260 | HEATR3 | Adipose - Visceral (Omentum) | 4,60E-64 | 0,53 | chr16_50059327_G_A_b38 |
| 16q12.1 | rs12599260 | HEATR3 | Skin - Not Sun Exposed (Suprapubic) | 1,80E-63 | 0,51 | chr16_50059327_G_A_b38 |

| Locus | Lead variant | Gene | Tissue | P-value | NES | Variant ID in GTEx |
|---|---|---|---|---|---|---|
| 16q12.1 | rs12599260 | HEATR3 | Colon - Transverse | 2,10E-61 | 0,63 | chr16_50059327_G_A_b38 |
| 16q12.1 | rs12599260 | HEATR3 | Brain - Cerebellum | 2,60E-61 | 1,1 | chr16_50059327_G_A_b38 |
| 16q12.1 | rs12599260 | HEATR3 | Skin - Sun Exposed (Lower leg) | 3,30E-59 | 0,44 | chr16_50059327_G_A_b38 |
| 16q12.1 | rs12599260 | HEATR3 | Colon - Sigmoid | 7,40E-52 | 0,77 | chr16_50059327_G_A_b38 |
| 16q12.1 | rs12599260 | HEATR3 | Stomach | 3,50E-51 | 0,64 | chr16_50059327_G_A_b38 |
| 16q12.1 | rs12599260 | HEATR3 | Breast - Mammary Tissue | 4,20E-51 | 0,53 | chr16_50059327_G_A_b38 |
| 16q12.1 | rs12599260 | HEATR3 | Pancreas | 6,20E-50 | 0,69 | chr16_50059327_G_A_b38 |
| 16q12.1 | rs12599260 | HEATR3 | Heart - Atrial Appendage | 1,80E-49 | 0,57 | chr16_50059327_G_A_b38 |
| 16q12.1 | rs12599260 | HEATR3 | Pituitary | 8,10E-47 | 0,94 | chr16_50059327_G_A_b38 |
| 16q12.1 | rs12599260 | HEATR3 | Spleen | 3,80E-46 | 0,89 | chr16_50059327_G_A_b38 |
| 16q12.1 | rs12599260 | HEATR3 | Brain - Cerebellar Hemisphere | 8,80E-45 | 1,1 | chr16_50059327_G_A_b38 |
| 16q12.1 | rs12599260 | HEATR3 | Brain - Cortex | 1,40E-43 | 0,94 | chr16_50059327_G_A_b38 |
| 16q12.1 | rs12599260 | HEATR3 | Muscle - Skeletal | 8,00E-42 | 0,39 | chr16_50059327_G_A_b38 |
| 16q12.1 | rs12599260 | HEATR3 | Heart - Left Ventricle | 2,00E-39 | 0,62 | chr16_50059327_G_A_b38 |
| 16q12.1 | rs12599260 | HEATR3 | Esophagus - Mucosa | 5,10E-36 | 0,37 | chr16_50059327_G_A_b38 |
| 16q12.1 | rs12599260 | HEATR3 | Brain - Nucleus accumbens (basal ganglia) | 1,40E-30 | 0,63 | chr16_50059327_G_A_b38 |
| 16q12.1 | rs12599260 | HEATR3 | Adrenal Gland | 3,50E-28 | 0,76 | chr16_50059327_G_A_b38 |
| 16q12.1 | rs12599260 | HEATR3 | Small Intestine - Terminal Ileum | 4,10E-28 | 0,79 | chr16_50059327_G_A_b38 |
| 16q12.1 | rs12599260 | HEATR3 | Artery - Coronary | 4,30E-28 | 0,61 | chr16_50059327_G_A_b38 |
| 16q12.1 | rs12599260 | HEATR3 | Brain - Caudate (basal ganglia) | 3,90E-26 | 0,7 | chr16_50059327_G_A_b38 |
| 16q12.1 | rs12599260 | HEATR3 | Cells - Cultured fibroblasts | 2,00E-25 | 0,19 | chr16_50059327_G_A_b38 |
| 16q12.1 | rs12599260 | HEATR3 | Prostate | 8,00E-25 | 0,59 | chr16_50059327_G_A_b38 |
| 16q12.1 | rs12599260 | HEATR3 | Brain - Frontal Cortex (BA9) | 2,10E-24 | 0,76 | chr16_50059327_G_A_b38 |
| 16q12.1 | rs12599260 | HEATR3 | Brain - Putamen (basal ganglia) | 1,40E-23 | 0,68 | chr16_50059327_G_A_b38 |
| 16q12.1 | rs12599260 | HEATR3 | Ovary | 7,90E-23 | 0,52 | chr16_50059327_G_A_b38 |
| 16q12.1 | rs12599260 | HEATR3 | Brain - Hypothalamus | 1,70E-20 | 0,91 | chr16_50059327_G_A_b38 |
| 16q12.1 | rs12599260 | HEATR3 | Brain - Anterior cingulate cortex (BA24) | 5,00E-20 | 0,86 | chr16_50059327_G_A_b38 |
| 16q12.1 | rs12599260 | HEATR3 | Brain - Spinal cord (cervical c-1) | 9,90E-20 | 0,87 | chr16_50059327_G_A_b38 |
| 16q12.1 | rs12599260 | HEATR3 | Brain - Hippocampus | 3,60E-19 | 0,79 | chr16_50059327_G_A_b38 |
| 16q12.1 | rs12599260 | HEATR3 | Brain - Substantia nigra | 4,80E-17 | 0,72 | chr16_50059327_G_A_b38 |
| 16q12.1 | rs12599260 | HEATR3 | Cells - EBV-transformed lymphocytes | 4,70E-15 | 0,41 | chr16_50059327_G_A_b38 |
| 16q12.1 | rs12599260 | HEATR3 | Uterus | 6,10E-15 | 0,48 | chr16_50059327_G_A_b38 |
| 16q12.1 | rs12599260 | HEATR3 | Liver | 7,10E-14 | 0,35 | chr16_50059327_G_A_b38 |
| 16q12.1 | rs12599260 | HEATR3 | Brain - Amygdala | 1,10E-12 | 0,71 | chr16_50059327_G_A_b38 |
| 16q12.1 | rs12599260 | HEATR3 | Minor Salivary Gland | 1,40E-12 | 0,61 | chr16_50059327_G_A_b38 |
| 16q12.1 | rs12599260 | HEATR3 | Vagina | 6,80E-10 | 0,42 | chr16_50059327_G_A_b38 |
| 16q12.1 | rs12599260 | HEATR3 | Kidney - Cortex | 5,80E-07 | 0,74 | chr16_50059327_G_A_b38 |
| 16q12.1 | rs12599260 | RP11-429P3.8 | Brain - Cerebellum | 8,00E-14 | 0,61 | chr16_50059327_G_A_b38 |
| 16q12.1 | rs12599260 | RP11-429P3.8 | Lung | 5,30E-13 | 0,38 | chr16_50059327_G_A_b38 |
| 16q12.1 | rs12599260 | RP11-429P3.8 | Pituitary | 1,40E-11 | 0,53 | chr16_50059327_G_A_b38 |
| 16q12.1 | rs12599260 | RP11-429P3.8 | Artery - Tibial | 2,30E-11 | 0,33 | chr16_50059327_G_A_b38 |
| 16q12.1 | rs12599260 | RP11-429P3.8 | Breast - Mammary Tissue | 3,70E-11 | 0,37 | chr16_50059327_G_A_b38 |
| 16q12.1 | rs12599260 | RP11-429P3.8 | Stomach | 4,20E-11 | 0,45 | chr16_50059327_G_A_b38 |

| Locus | Lead variant | Gene | Tissue | P-value | NES | Variant ID in GTEx |
|---|---|---|---|---|---|---|
| 16q12.1 | rs12599260 | RP11-429P3.8 | Thyroid | 6,70E-10 | 0,3 | chr16_50059327_G_A_b38 |
| 16q12.1 | rs12599260 | RP11-429P3.8 | Skin - Not Sun Exposed (Suprapubic) | 8,80E-10 | 0,3 | chr16_50059327_G_A_b38 |
| 16q12.1 | rs12599260 | RP11-429P3.8 | Esophagus - Muscularis | 1,10E-09 | 0,36 | chr16_50059327_G_A_b38 |
| 16q12.1 | rs12599260 | RP11-429P3.8 | Brain - Cerebellar Hemisphere | 1,30E-08 | 0,39 | chr16_50059327_G_A_b38 |
| 16q12.1 | rs12599260 | RP11-429P3.8 | Artery - Aorta | 1,80E-08 | 0,34 | chr16_50059327_G_A_b38 |
| 16q12.1 | rs12599260 | RP11-429P3.8 | Esophagus - Gastroesophageal Junction | 2,00E-08 | 0,41 | chr16_50059327_G_A_b38 |
| 16q12.1 | rs12599260 | RP11-429P3.8 | Adipose - Subcutaneous | 2,40E-08 | 0,24 | chr16_50059327_G_A_b38 |
| 16q12.1 | rs12599260 | RP11-429P3.8 | Skin - Sun Exposed (Lower leg) | 4,40E-08 | 0,29 | chr16_50059327_G_A_b38 |
| 16q12.1 | rs12599260 | RP11-429P3.8 | Nerve - Tibial | 5,00E-08 | 0,28 | chr16_50059327_G_A_b38 |
| 16q12.1 | rs12599260 | RP11-429P3.8 | Spleen | 1,40E-07 | 0,33 | chr16_50059327_G_A_b38 |
| 16q12.1 | rs12599260 | RP11-429P3.8 | Colon - Sigmoid | 4,00E-07 | 0,38 | chr16_50059327_G_A_b38 |
| 16q12.1 | rs12599260 | RP11-429P3.8 | Adipose - Visceral (Omentum) | 4,10E-07 | 0,24 | chr16_50059327_G_A_b38 |
| 16q12.1 | rs12599260 | RP11-429P3.8 | Esophagus - Mucosa | 7,50E-07 | 0,26 | chr16_50059327_G_A_b38 |
| 16q12.1 | rs12599260 | RP11-429P3.8 | Adrenal Gland | 0,0000025 | 0,37 | chr16_50059327_G_A_b38 |
| 16q12.1 | rs12599260 | RP11-429P3.8 | Colon - Transverse | 0,0000073 | 0,32 | chr16_50059327_G_A_b38 |
| 16q12.1 | rs12599260 | RP11-429P3.8 | Heart - Atrial Appendage | 0,000016 | 0,31 | chr16_50059327_G_A_b38 |
| 16q12.1 | rs12599260 | RP11-429P3.8 | Artery - Coronary | 0,000017 | 0,37 | chr16_50059327_G_A_b38 |
| 16q12.1 | rs66998222 | *no significant eQTLs* | | | | |
| 17p12 | rs12601765 | *no significant eQTLs* | | | | |
| 19p12 | rs8105767 | AC025811.3 | Thyroid | 0,000014 | 0,17 | chr19_22032639_A_G_b38 |
| 19p12 | rs8105767 | BNIP3P30 | Thyroid | 0,00004 | 0,22 | chr19_22032639_A_G_b38 |
| 19p12 | rs8105767 | CTD-3030D20.1 | Brain - Putamen (basal ganglia) | 0,000057 | -0,43 | chr19_22032639_A_G_b38 |
| 19p12 | rs8105767 | MTDHP3 | Whole Blood | 0,000014 | -0,22 | chr19_22032639_A_G_b38 |
| 19p12 | rs8105767 | RP11-157B13.7 | Testis | 1,70E-08 | -0,2 | chr19_22032639_A_G_b38 |
| 19p12 | rs8105767 | RP11-678G14.2 | Thyroid | 0,000018 | -0,25 | chr19_22032639_A_G_b38 |
| 19p12 | rs8105767 | RP11-678G14.3 | Whole Blood | 0,0000015 | -0,22 | chr19_22032639_A_G_b38 |
| 19p12 | rs8105767 | RP11-678G14.3 | Muscle - Skeletal | 0,0000062 | -0,23 | chr19_22032639_A_G_b38 |
| 19p12 | rs8105767 | RP11-678G14.3 | Adipose - Visceral (Omentum) | 0,000027 | -0,28 | chr19_22032639_A_G_b38 |
| 19p12 | rs8105767 | RP11-678G14.3 | Brain - Cerebellar Hemisphere | 0,000087 | -0,47 | chr19_22032639_A_G_b38 |
| 19p12 | rs8105767 | VN1R85P | Thyroid | 4,70E-15 | 0,46 | chr19_22032639_A_G_b38 |
| 19p12 | rs8105767 | VN1R85P | Esophagus - Muscularis | 1,60E-07 | 0,35 | chr19_22032639_A_G_b38 |
| 19p12 | rs8105767 | VN1R85P | Artery - Aorta | 0,00004 | 0,28 | chr19_22032639_A_G_b38 |
| 19p12 | rs8105767 | VN1R85P | Heart - Left Ventricle | 0,00013 | 0,27 | chr19_22032639_A_G_b38 |
| 19p12 | rs8105767 | ZNF257 | Skin - Sun Exposed (Lower leg) | 6,10E-50 | 0,77 | chr19_22032639_A_G_b38 |
| 19p12 | rs8105767 | ZNF257 | Adipose - Subcutaneous | 1,20E-40 | 0,74 | chr19_22032639_A_G_b38 |
| 19p12 | rs8105767 | ZNF257 | Lung | 3,00E-37 | 0,64 | chr19_22032639_A_G_b38 |
| 19p12 | rs8105767 | ZNF257 | Esophagus - Mucosa | 3,60E-37 | 0,73 | chr19_22032639_A_G_b38 |
| 19p12 | rs8105767 | ZNF257 | Nerve - Tibial | 4,70E-33 | 0,66 | chr19_22032639_A_G_b38 |
| 19p12 | rs8105767 | ZNF257 | Artery - Tibial | 5,20E-32 | 0,63 | chr19_22032639_A_G_b38 |
| 19p12 | rs8105767 | ZNF257 | Skin - Not Sun Exposed (Suprapubic) | 1,30E-27 | 0,61 | chr19_22032639_A_G_b38 |
| 19p12 | rs8105767 | ZNF257 | Colon - Transverse | 3,60E-27 | 0,79 | chr19_22032639_A_G_b38 |
| 19p12 | rs8105767 | ZNF257 | Whole Blood | 1,60E-25 | 0,46 | chr19_22032639_A_G_b38 |
| 19p12 | rs8105767 | ZNF257 | Cells - Cultured fibroblasts | 1,30E-24 | 0,63 | chr19_22032639_A_G_b38 |

| Locus | Lead variant | Gene | Tissue | P-value | NES | Variant ID in GTEx |
|---|---|---|---|---|---|---|
| 19p12 | rs8105767 | ZNF257 | Thyroid | 4,20E-22 | 0,47 | chr19_22032639_A_G_b38 |
| 19p12 | rs8105767 | ZNF257 | Breast - Mammary Tissue | 1,20E-20 | 0,53 | chr19_22032639_A_G_b38 |
| 19p12 | rs8105767 | ZNF257 | Adipose - Visceral (Omentum) | 1,50E-20 | 0,49 | chr19_22032639_A_G_b38 |
| 19p12 | rs8105767 | ZNF257 | Stomach | 8,30E-20 | 0,65 | chr19_22032639_A_G_b38 |
| 19p12 | rs8105767 | ZNF257 | Artery - Aorta | 3,60E-19 | 0,59 | chr19_22032639_A_G_b38 |
| 19p12 | rs8105767 | ZNF257 | Heart - Left Ventricle | 7,50E-19 | 0,61 | chr19_22032639_A_G_b38 |
| 19p12 | rs8105767 | ZNF257 | Esophagus - Muscularis | 3,80E-18 | 0,54 | chr19_22032639_A_G_b38 |
| 19p12 | rs8105767 | ZNF257 | Prostate | 2,80E-15 | 0,69 | chr19_22032639_A_G_b38 |
| 19p12 | rs8105767 | ZNF257 | Pancreas | 3,60E-15 | 0,67 | chr19_22032639_A_G_b38 |
| 19p12 | rs8105767 | ZNF257 | Heart - Atrial Appendage | 2,60E-14 | 0,51 | chr19_22032639_A_G_b38 |
| 19p12 | rs8105767 | ZNF257 | Brain - Hypothalamus | 1,70E-13 | 0,72 | chr19_22032639_A_G_b38 |
| 19p12 | rs8105767 | ZNF257 | Spleen | 4,00E-13 | 0,59 | chr19_22032639_A_G_b38 |
| 19p12 | rs8105767 | ZNF257 | Brain - Cerebellar Hemisphere | 7,70E-13 | 0,77 | chr19_22032639_A_G_b38 |
| 19p12 | rs8105767 | ZNF257 | Brain - Cerebellum | 9,30E-13 | 0,77 | chr19_22032639_A_G_b38 |
| 19p12 | rs8105767 | ZNF257 | Artery - Coronary | 1,10E-12 | 0,62 | chr19_22032639_A_G_b38 |
| 19p12 | rs8105767 | ZNF257 | Small Intestine - Terminal Ileum | 7,60E-12 | 0,43 | chr19_22032639_A_G_b38 |
| 19p12 | rs8105767 | ZNF257 | Esophagus - Gastroesophageal Junction | 4,80E-11 | 0,47 | chr19_22032639_A_G_b38 |
| 19p12 | rs8105767 | ZNF257 | Brain - Spinal cord (cervical c-1) | 6,20E-11 | 0,76 | chr19_22032639_A_G_b38 |
| 19p12 | rs8105767 | ZNF257 | Colon - Sigmoid | 1,10E-10 | 0,51 | chr19_22032639_A_G_b38 |
| 19p12 | rs8105767 | ZNF257 | Brain - Putamen (basal ganglia) | 1,90E-10 | 0,66 | chr19_22032639_A_G_b38 |
| 19p12 | rs8105767 | ZNF257 | Adrenal Gland | 6,20E-09 | 0,55 | chr19_22032639_A_G_b38 |
| 19p12 | rs8105767 | ZNF257 | Brain - Nucleus accumbens (basal ganglia) | 7,40E-09 | 0,48 | chr19_22032639_A_G_b38 |
| 19p12 | rs8105767 | ZNF257 | Ovary | 3,00E-08 | 0,48 | chr19_22032639_A_G_b38 |
| 19p12 | rs8105767 | ZNF257 | Vagina | 8,00E-08 | 0,57 | chr19_22032639_A_G_b38 |
| 19p12 | rs8105767 | ZNF257 | Brain - Caudate (basal ganglia) | 8,20E-08 | 0,46 | chr19_22032639_A_G_b38 |
| 19p12 | rs8105767 | ZNF257 | Brain - Hippocampus | 7,80E-07 | 0,49 | chr19_22032639_A_G_b38 |
| 19p12 | rs8105767 | ZNF257 | Brain - Substantia nigra | 8,50E-07 | 0,59 | chr19_22032639_A_G_b38 |
| 19p12 | rs8105767 | ZNF257 | Pituitary | 0,0000021 | 0,35 | chr19_22032639_A_G_b38 |
| 19p12 | rs8105767 | ZNF257 | Brain - Cortex | 0,000005 | 0,5 | chr19_22032639_A_G_b38 |
| 19p12 | rs8105767 | ZNF257 | Cells - EBV-transformed lymphocytes | 0,0000065 | 0,62 | chr19_22032639_A_G_b38 |
| 19p12 | rs8105767 | ZNF257 | Uterus | 0,0000072 | 0,66 | chr19_22032639_A_G_b38 |
| 19p12 | rs8105767 | ZNF257 | Brain - Frontal Cortex (BA9) | 0,0000098 | 0,52 | chr19_22032639_A_G_b38 |
| 19p12 | rs8105767 | ZNF257 | Minor Salivary Gland | 0,000011 | 0,55 | chr19_22032639_A_G_b38 |
| 19p12 | rs8105767 | ZNF257 | Brain - Anterior cingulate cortex (BA24) | 0,000018 | 0,51 | chr19_22032639_A_G_b38 |
| 19p12 | rs8105767 | ZNF429 | Whole Blood | 0,0000065 | -0,15 | chr19_22032639_A_G_b38 |
| 19p12 | rs8105767 | ZNF493 | Whole Blood | 0,000033 | -0,14 | chr19_22032639_A_G_b38 |
| 19p12 | rs8105767 | ZNF676 | Testis | 1,20E-11 | -0,36 | chr19_22032639_A_G_b38 |
| 19p12 | rs8105767 | ZNF676 | Thyroid | 5,50E-11 | 0,35 | chr19_22032639_A_G_b38 |
| 19p12 | rs8105767 | ZNF676 | Lung | 0,000067 | 0,19 | chr19_22032639_A_G_b38 |
| 19p12 | rs8105767 | ZNF708 | Whole Blood | 0,0000014 | -0,16 | chr19_22032639_A_G_b38 |
| 19p12 | rs8105767 | ZNF708 | Skin - Not Sun Exposed (Suprapubic) | 0,00028 | -0,15 | chr19_22032639_A_G_b38 |
| 19p12 | rs8105767 | ZNF729 | Testis | 0,0001 | -0,21 | chr19_22032639_A_G_b38 |
| 19p12 | rs8105767 | ZNF98 | Heart - Left Ventricle | 0,000083 | 0,3 | chr19_22032639_A_G_b38 |

| Locus | Lead variant | Gene | Tissue | P-value | NES | Variant ID in GTEx |
|---|---|---|---|---|---|---|
| 20q13.31 | rs13039273 | BMP7 | Thyroid | 0,000081 | -0,071 | chr20_57441016_T_C_b38 |
| 20q13.31 | rs13039273 | RBM38 | Pancreas | 2,80E-16 | -0,59 | chr20_57441016_T_C_b38 |
| 20q13.31 | rs13039273 | RBM38 | Heart - Atrial Appendage | 3,10E-07 | -0,16 | chr20_57441016_T_C_b38 |
| 20q13.31 | rs13039273 | RBM38 | Ovary | 0,0000087 | -0,23 | chr20_57441016_T_C_b38 |
| 20q13.33 | rs75691080 | RTEL1 | Adipose - Visceral (Omentum) | 3,00E-09 | -0,27 | chr20_63638397_C_T_b38 |
| 20q13.33 | rs75691080 | RTEL1 | Heart - Atrial Appendage | 0,0000041 | -0,21 | chr20_63638397_C_T_b38 |
| 20q13.33 | rs75691080 | RTEL1 | Muscle - Skeletal | 0,000008 | -0,19 | chr20_63638397_C_T_b38 |
| 20q13.33 | rs75691080 | RTEL1 | Breast - Mammary Tissue | 0,000013 | -0,21 | chr20_63638397_C_T_b38 |
| 20q13.33 | rs75691080 | RTEL1 | Cells - Cultured fibroblasts | 0,00011 | -0,14 | chr20_63638397_C_T_b38 |
| 20q13.33 | rs75691080 | STMN3 | Cells - Cultured fibroblasts | 3,70E-11 | -0,48 | chr20_63638397_C_T_b38 |
| 20q13.33 | rs75691080 | STMN3 | Artery - Aorta | 0,000064 | -0,25 | chr20_63638397_C_T_b38 |
| 20q13.33 | rs75691080 | TNFRSF6B | Cells - Cultured fibroblasts | 0,0000025 | -0,27 | chr20_63638397_C_T_b38 |
| 21q22.12 | rs2834747 | *no significant eQTLs* | | | | |
| 22q12.3 | rs9610482 | *no significant eQTLs* | | | | |
| 2p11.2 (META-2) | rs573520030 | *no significant eQTLs* | | | | |
| 4p11 (META-2) | rs538533131 | *no significant eQTLs* | | | | |
| 10q25.1 (META-2) | rs12247648 | *no significant eQTLs* | | | | |
| Xp11.23 (META-2) | rs6611312 | CHST7 | Brain - Cortex | 3,60E-07 | 0,18 | chrX_46904059_T_G_b38 |
| Xp11.23 (META-2) | rs6611312 | CHST7 | Brain - Cerebellum | 0,0000041 | 0,2 | chrX_46904059_T_G_b38 |
| Xp11.23 (META-2) | rs6611312 | CHST7 | Brain - Putamen (basal ganglia) | 0,0000053 | 0,21 | chrX_46904059_T_G_b38 |
| Xp11.23 (META-2) | rs6611312 | CHST7 | Brain - Frontal Cortex (BA9) | 0,000011 | 0,17 | chrX_46904059_T_G_b38 |
| Xp11.23 (META-2) | rs6611312 | JADE3 | Esophagus - Muscularis | 1,40E-07 | -0,083 | chrX_46904059_T_G_b38 |
| Xp11.23 (META-2) | rs6611312 | JADE3 | Esophagus - Mucosa | 0,000065 | -0,11 | chrX_46904059_T_G_b38 |
| Xp11.23 (META-2) | rs6611312 | SLC9A7 | Artery - Tibial | 8,40E-41 | -0,4 | chrX_46904059_T_G_b38 |
| Xp11.23 (META-2) | rs6611312 | SLC9A7 | Nerve - Tibial | 3,40E-38 | -0,39 | chrX_46904059_T_G_b38 |
| Xp11.23 (META-2) | rs6611312 | SLC9A7 | Esophagus - Muscularis | 2,50E-29 | -0,38 | chrX_46904059_T_G_b38 |
| Xp11.23 (META-2) | rs6611312 | SLC9A7 | Colon - Sigmoid | 4,00E-22 | -0,37 | chrX_46904059_T_G_b38 |
| Xp11.23 (META-2) | rs6611312 | SLC9A7 | Lung | 3,80E-21 | -0,27 | chrX_46904059_T_G_b38 |
| Xp11.23 (META-2) | rs6611312 | SLC9A7 | Artery - Aorta | 4,40E-18 | -0,38 | chrX_46904059_T_G_b38 |
| Xp11.23 (META-2) | rs6611312 | SLC9A7 | Adipose - Subcutaneous | 2,10E-17 | -0,22 | chrX_46904059_T_G_b38 |
| Xp11.23 (META-2) | rs6611312 | SLC9A7 | Esophagus - Mucosa | 8,40E-16 | -0,25 | chrX_46904059_T_G_b38 |
| Xp11.23 (META-2) | rs6611312 | SLC9A7 | Esophagus - Gastroesophageal Junction | 3,40E-14 | -0,29 | chrX_46904059_T_G_b38 |
| Xp11.23 (META-2) | rs6611312 | SLC9A7 | Muscle - Skeletal | 4,20E-13 | -0,15 | chrX_46904059_T_G_b38 |
| Xp11.23 (META-2) | rs6611312 | SLC9A7 | Artery - Coronary | 4,30E-13 | -0,35 | chrX_46904059_T_G_b38 |
| Xp11.23 (META-2) | rs6611312 | SLC9A7 | Skin - Sun Exposed (Lower leg) | 5,50E-11 | -0,15 | chrX_46904059_T_G_b38 |
| Xp11.23 (META-2) | rs6611312 | SLC9A7 | Colon - Transverse | 4,80E-08 | -0,17 | chrX_46904059_T_G_b38 |
| Xp11.23 (META-2) | rs6611312 | SLC9A7 | Heart - Left Ventricle | 0,0000025 | -0,13 | chrX_46904059_T_G_b38 |
| Xp11.23 (META-2) | rs6611312 | SLC9A7 | Liver | 0,0000026 | -0,24 | chrX_46904059_T_G_b38 |
| Xp11.23 (META-2) | rs6611312 | SLC9A7 | Thyroid | 0,0000094 | -0,088 | chrX_46904059_T_G_b38 |
| Xp11.23 (META-2) | rs6611312 | SLC9A7 | Skin - Not Sun Exposed (Suprapubic) | 0,000011 | -0,11 | chrX_46904059_T_G_b38 |
| Xp11.23 (META-2) | rs6611312 | SLC9A7 | Cells - Cultured fibroblasts | 0,000055 | 0,085 | chrX_46904059_T_G_b38 |

**Table S8. Genetic correlations of UL with other traits in LD Hub database.**
The analyses were completed using an automated LD score regression pipeline[6] available at http://ldsc.broadinstitute.org/. After correcting for 830 traits tested, we considered statistical significance at $p<6x10^{-5}$ (highlighted with asterisk). Genetic correlations ($r_g$) with $p<0.05$ are shown.

| trait1 | trait2 | PMID | Category | ethnicity | $r_g$ | se | p |
|---|---|---|---|---|---|---|---|
| UL | Non-cancer illness code, self-reported: uterine fibroids | 0 | ukbb | European | 0.9668 | 0.0761 | 5.9077e-37* |
| UL | Diagnoses - main ICD10: D25 Leiomyoma of uterus | 0 | ukbb | European | 0.9069 | 0.0795 | 3.8946e-30* |
| UL | Bilateral oophorectomy (both ovaries removed) | 0 | ukbb | European | 0.7434 | 0.0709 | 1.0499e-25* |
| UL | Ever had hysterectomy (womb removed) | 0 | ukbb | European | 0.6233 | 0.0696 | 3.2388e-19* |
| UL | Number of operations, self-reported | 0 | ukbb | European | 0.2951 | 0.0428 | 5.0895e-12* |
| UL | Age when periods started (menarche) | 0 | ukbb | European | -0.1802 | 0.0298 | 1.3983e-09* |
| UL | Age at Menarche | 25231870 | reproductive | European | -0.1981 | 0.0332 | 2.3017e-09* |
| UL | Number of self-reported non-cancer illnesses | 0 | ukbb | European | 0.19 | 0.0363 | 1.5993e-07* |
| UL | Frequency of tiredness / lethargy in last 2 weeks | 0 | ukbb | European | 0.175 | 0.0343 | 3.3458e-07* |
| UL | Diagnoses - main ICD10: N92 Excessive, frequent and irregular menstruation | 0 | ukbb | European | 0.5086 | 0.1017 | 5.6598e-07* |
| UL | Vascular/heart problems diagnosed by doctor: None of the above | 0 | ukbb | European | -0.1536 | 0.0328 | 2.8358e-06* |
| UL | Years of schooling 2016 | 27225129 | education | European | -0.1255 | 0.0271 | 3.762e-06* |
| UL | Vascular/heart problems diagnosed by doctor: High blood pressure | 0 | ukbb | European | 0.1474 | 0.0322 | 4.579e-06* |
| UL | Impedance of arm (right) | 0 | ukbb | European | -0.1372 | 0.0301 | 5.1161e-06* |
| UL | Impedance of arm (left) | 0 | ukbb | European | -0.1364 | 0.0304 | 7.0462e-06* |
| UL | Non-cancer illness code, self-reported: hypertension | 0 | ukbb | European | 0.1445 | 0.0322 | 7.4186e-06* |
| UL | Number of treatments/medications taken | 0 | ukbb | European | 0.158 | 0.0358 | 1.0016e-05* |
| UL | Arm predicted mass (right) | 0 | ukbb | European | 0.1236 | 0.0281 | 1.0712e-05* |
| UL | Neuroticism | 27089181 | personality | European | 0.1712 | 0.0389 | 1.0729e-05* |
| UL | Arm predicted mass (left) | 0 | ukbb | European | 0.1221 | 0.0279 | 1.199e-05* |
| UL | Hip circumference | 25673412 | anthropometric | European | 0.1649 | 0.0379 | 1.3608e-05* |
| UL | Arm fat-free mass (right) | 0 | ukbb | European | 0.1207 | 0.0278 | 1.4525e-05* |
| UL | Arm fat-free mass (left) | 0 | ukbb | European | 0.1202 | 0.0278 | 1.526e-05* |
| UL | Overall health rating | 0 | ukbb | European | 0.1335 | 0.0314 | 2.1308e-05* |
| UL | Waist circumference | 25673412 | anthropometric | European | 0.1555 | 0.0367 | 2.2591e-05* |
| UL | Had menopause | 0 | ukbb | European | -0.3269 | 0.0772 | 2.3094e-05* |
| UL | Frequency of depressed mood in last 2 weeks | 0 | ukbb | European | 0.1761 | 0.0419 | 2.6306e-05* |
| UL | Sensitivity / hurt feelings | 0 | ukbb | European | 0.1475 | 0.0353 | 2.932e-05* |
| UL | Illnesses of siblings: None of the above (group 1) | 0 | ukbb | European | -0.1804 | 0.0433 | 3.1229e-05* |
| UL | Basal metabolic rate | 0 | ukbb | European | 0.1142 | 0.0275 | 3.2734e-05* |
| UL | Depressive symptoms | 27089181 | psychiatric | European | 0.2047 | 0.0493 | 3.3117e-05* |
| UL | Waist circumference | 0 | ukbb | European | 0.1182 | 0.0285 | 3.4278e-05* |
| UL | Whole body water mass | 0 | ukbb | European | 0.1146 | 0.0278 | 3.7429e-05* |
| UL | Neuroticism score | 0 | ukbb | European | 0.1358 | 0.033 | 3.8753e-05* |
| UL | Trunk fat-free mass | 0 | ukbb | European | 0.1123 | 0.0273 | 3.8954e-05* |
| UL | Whole body fat-free mass | 0 | ukbb | European | 0.1132 | 0.0276 | 4.2057e-05* |
| UL | Miserableness | 0 | ukbb | European | 0.1393 | 0.0341 | 4.5168e-05* |
| UL | Triglycerides | 20686565 | lipids | European | 0.1595 | 0.0392 | 4.8016e-05* |
| UL | Trunk predicted mass | 0 | ukbb | European | 0.1102 | 0.0272 | 5.1615e-05* |
| UL | Ever used hormone-replacement therapy (HRT) | 0 | ukbb | European | 0.202 | 0.0501 | 5.538e-05* |

| trait1 | trait2 | PMID | Category | ethnicity | $r_g$ | se | p |
|---|---|---|---|---|---|---|---|
| UL | Impedance of whole body | 0 | ukbb | European | -0.1242 | 0.0309 | 5.81e-05* |
| UL | Seen doctor (GP) for nerves, anxiety, tension or depression | 0 | ukbb | European | 0.15 | 0.0373 | 5.8163e-05* |
| UL | Mood swings | 0 | ukbb | European | 0.1555 | 0.0387 | 5.9597e-05* |
| UL | Hearing difficulty/problems with background noise | 0 | ukbb | European | 0.1248 | 0.0317 | 8.0633e-05 |
| UL | Had other major operations | 0 | ukbb | European | 0.211 | 0.0539 | 9.0346e-05 |
| UL | Comparative height size at age 10 | 0 | ukbb | European | 0.1045 | 0.0274 | 1,00e-04 |
| UL | Fed-up feelings | 0 | ukbb | European | 0.1473 | 0.0387 | 1,00e-04 |
| UL | Medication for cholesterol, blood pressure, diabetes, or take exogenous hormones: None of the above | 0 | ukbb | European | -0.1681 | 0.0441 | 1,00e-04 |
| UL | Medication for cholesterol, blood pressure, diabetes, or take exogenous hormones: Hormone replacement therapy | 0 | ukbb | European | 0.2758 | 0.0717 | 1,00e-04 |
| UL | Alcohol intake frequency. | 0 | ukbb | European | 0.1178 | 0.0318 | 2,00e-04 |
| UL | Diabetes diagnosed by doctor | 0 | ukbb | European | 0.1553 | 0.0413 | 2,00e-04 |
| UL | Leg fat-free mass (right) | 0 | ukbb | European | 0.1058 | 0.0285 | 2,00e-04 |
| UL | Leg predicted mass (right) | 0 | ukbb | European | 0.1058 | 0.0285 | 2,00e-04 |
| UL | Leg fat-free mass (left) | 0 | ukbb | European | 0.107 | 0.0287 | 2,00e-04 |
| UL | Leg predicted mass (left) | 0 | ukbb | European | 0.1066 | 0.0287 | 2,00e-04 |
| UL | Illnesses of siblings: High blood pressure | 0 | ukbb | European | 0.1743 | 0.0476 | 2,00e-04 |
| UL | Medication for cholesterol, blood pressure or diabetes: Blood pressure medication | 0 | ukbb | European | 0.1478 | 0.04 | 2,00e-04 |
| UL | Diagnoses - main ICD10: R07 Pain in throat and chest | 0 | ukbb | European | 0.2303 | 0.0627 | 2,00e-04 |
| UL | Difference in height between adolescence and adulthood; age 14 | 23449627 | anthropometric | European | -0.3142 | 0.0863 | 3,00e-04 |
| UL | Loneliness, isolation | 0 | ukbb | European | 0.1521 | 0.0418 | 3,00e-04 |
| UL | Weight | 0 | ukbb | European | 0.0966 | 0.027 | 3,00e-04 |
| UL | Heel bone mineral density (BMD) T-score, automated | 0 | ukbb | European | 0.1187 | 0.0345 | 6,00e-04 |
| UL | Qualifications: College or University degree | 0 | ukbb | European | -0.0983 | 0.0288 | 6,00e-04 |
| UL | Medication for pain relief, constipation, heartburn: Aspirin | 0 | ukbb | European | 0.1762 | 0.0512 | 6,00e-04 |
| UL | Frequency of tenseness / restlessness in last 2 weeks | 0 | ukbb | European | 0.146 | 0.0431 | 7,00e-04 |
| UL | Pain type(s) experienced in last month: None of the above | 0 | ukbb | European | -0.1225 | 0.0361 | 7,00e-04 |
| UL | Taking other prescription medications | 0 | ukbb | European | 0.136 | 0.0406 | 8,00e-04 |
| UL | Body mass index (BMI) | 0 | ukbb | European | 0.0949 | 0.0284 | 8,00e-04 |
| UL | Femoral Neck bone mineral density | 26367794 | bone | Mixed | 0.1803 | 0.0544 | 9,00e-04 |
| UL | Cancer diagnosed by doctor | 0 | ukbb | European | 0.2839 | 0.0854 | 9,00e-04 |
| UL | Number of self-reported cancers | 0 | ukbb | European | 0.2891 | 0.0886 | 0.0011 |
| UL | Medication for cholesterol, blood pressure or diabetes: None of the above | 0 | ukbb | European | -0.1429 | 0.0437 | 0.0011 |
| UL | Body mass index | 20935630 | anthropometric | European | 0.1097 | 0.0339 | 0.0012 |
| UL | Non-cancer illness code, self-reported: heart attack/myocardial infarction | 0 | ukbb | European | 0.1831 | 0.0565 | 0.0012 |
| UL | Waist-to-hip ratio | 25673412 | anthropometric | European | 0.1164 | 0.0366 | 0.0015 |
| UL | Vascular/heart problems diagnosed by doctor: Angina | 0 | ukbb | European | 0.1564 | 0.0491 | 0.0015 |
| UL | Impedance of leg (left) | 0 | ukbb | European | -0.0994 | 0.0316 | 0.0017 |
| UL | Subjective well being | 27089181 | psychiatric | European | -0.1683 | 0.054 | 0.0018 |
| UL | Non-cancer illness code, self-reported: diabetes | 0 | ukbb | European | 0.1376 | 0.044 | 0.0018 |
| UL | Diastolic blood pressure, automated reading | 0 | ukbb | European | 0.0953 | 0.0306 | 0.0019 |
| UL | Illness, injury, bereavement, stress in last 2 years: Financial difficulties | 0 | ukbb | European | 0.1319 | 0.0424 | 0.0019 |
| UL | Pain type(s) experienced in last month: Stomach or abdominal pain | 0 | ukbb | European | 0.1559 | 0.0503 | 0.0019 |

| trait1 | trait2 | PMID | Category | ethnicity | $r_g$ | se | p |
|---|---|---|---|---|---|---|---|
| UL | Leg fat mass (left) | 0 | ukbb | European | 0.0854 | 0.0278 | 0.0021 |
| UL | Arm fat mass (left) | 0 | ukbb | European | 0.0856 | 0.028 | 0.0023 |
| UL | Impedance of leg (right) | 0 | ukbb | European | -0.0946 | 0.0312 | 0.0024 |
| UL | Long-standing illness, disability or infirmity | 0 | ukbb | European | 0.1207 | 0.0399 | 0.0025 |
| UL | Frequency of unenthusiasm / disinterest in last 2 weeks | 0 | ukbb | European | 0.1293 | 0.043 | 0.0026 |
| UL | Medication for cholesterol, blood pressure, diabetes, or take exogenous hormones: Blood pressure medication | 0 | ukbb | European | 0.1148 | 0.0381 | 0.0026 |
| UL | Medication for pain relief, constipation, heartburn: None of the above | 0 | ukbb | European | -0.1133 | 0.0376 | 0.0026 |
| UL | Age at Menopause | 26414677 | reproductive | European | 0.1745 | 0.0582 | 0.0027 |
| UL | Leg fat mass (right) | 0 | ukbb | European | 0.0842 | 0.0281 | 0.0027 |
| UL | Obesity class 1 | 23563607 | anthropometric | European | 0.1135 | 0.0379 | 0.0028 |
| UL | Diagnoses - main ICD10: R10 Abdominal and pelvic pain | 0 | ukbb | European | 0.216 | 0.0722 | 0.0028 |
| UL | Vascular/heart problems diagnosed by doctor: Heart attack | 0 | ukbb | European | 0.1679 | 0.0564 | 0.0029 |
| UL | Arm fat mass (right) | 0 | ukbb | European | 0.0826 | 0.0278 | 0.003 |
| UL | Irritability | 0 | ukbb | European | 0.1139 | 0.0385 | 0.0031 |
| UL | Diagnoses - main ICD10: R55 Syncope and collapse | 0 | ukbb | European | 0.2936 | 0.0995 | 0.0032 |
| UL | Medication for cholesterol, blood pressure, diabetes, or take exogenous hormones: Cholesterol lowering medication | 0 | ukbb | European | 0.1456 | 0.0496 | 0.0033 |
| UL | Diagnoses - main ICD10: M17 Gonarthrosis [arthrosis of knee] | 0 | ukbb | European | 0.1823 | 0.0626 | 0.0036 |
| UL | Non-cancer illness code, self-reported: hiatus hernia | 0 | ukbb | European | 0.2265 | 0.0797 | 0.0045 |
| UL | Illnesses of mother: None of the above (group 1) | 0 | ukbb | European | -0.1843 | 0.0648 | 0.0045 |
| UL | Creatinine (enzymatic) in urine | 0 | ukbb | European | 0.1081 | 0.0382 | 0.0047 |
| UL | Leptin,not,adjBMI | 26833098 | hormone | European | 0.2152 | 0.0763 | 0.0048 |
| UL | Pain type(s) experienced in last month: Headache | 0 | ukbb | European | 0.1126 | 0.0402 | 0.0051 |
| UL | Nap during day | 0 | ukbb | European | 0.0919 | 0.0329 | 0.0053 |
| UL | Non-cancer illness code, self-reported: angina | 0 | ukbb | European | 0.1363 | 0.0489 | 0.0053 |
| UL | Health satisfaction | 0 | ukbb | European | 0.1322 | 0.0475 | 0.0054 |
| UL | Age at first live birth | 0 | ukbb | European | -0.0986 | 0.0355 | 0.0055 |
| UL | Illnesses of siblings: Heart disease | 0 | ukbb | European | 0.1698 | 0.0612 | 0.0055 |
| UL | Non-cancer illness code, self-reported: type 2 diabetes | 0 | ukbb | European | 0.2661 | 0.0961 | 0.0056 |
| UL | Age at last live birth | 0 | ukbb | European | -0.1215 | 0.044 | 0.0058 |
| UL | Hip circumference | 0 | ukbb | European | 0.0761 | 0.0277 | 0.0059 |
| UL | Medication for pain relief, constipation, heartburn: Paracetamol | 0 | ukbb | European | 0.1158 | 0.0422 | 0.006 |
| UL | Neuroticism | 24828478 | personality | European | 0.2302 | 0.0839 | 0.0061 |
| UL | Job involves heavy manual or physical work | 0 | ukbb | European | 0.1073 | 0.0392 | 0.0062 |
| UL | Overweight | 23563607 | anthropometric | European | 0.1108 | 0.0405 | 0.0063 |
| UL | Non-cancer illness code, self-reported: gout | 0 | ukbb | European | 0.1598 | 0.0585 | 0.0063 |
| UL | Forced vital capacity | 0 | lung,function | European | -0.1384 | 0.0507 | 0.0063 |
| UL | Whole body fat mass | 0 | ukbb | European | 0.0742 | 0.0272 | 0.0064 |
| UL | Illnesses of mother: High blood pressure | 0 | ukbb | European | 0.1358 | 0.0498 | 0.0064 |
| UL | Diagnoses - main ICD10: R35 Polyuria | 0 | ukbb | European | 0.4404 | 0.1637 | 0.0071 |
| UL | Pulse wave Arterial Stiffness index | 0 | ukbb | European | 0.176 | 0.0656 | 0.0073 |
| UL | Non-cancer illness code, self-reported: osteoarthritis | 0 | ukbb | European | 0.1543 | 0.0578 | 0.0077 |

| trait1 | trait2 | PMID | Category | ethnicity | $r_g$ | se | p |
|---|---|---|---|---|---|---|---|
| UL | Non-cancer illness code, self-reported: varicose veins | 0 | ukbb | European | 0.2939 | 0.1104 | 0.0078 |
| UL | Heel bone mineral density (BMD) T-score, automated (left) | 0 | ukbb | European | 0.1031 | 0.0388 | 0.0079 |
| UL | Pain type(s) experienced in last month: Back pain | 0 | ukbb | European | 0.1136 | 0.0429 | 0.0081 |
| UL | Types of physical activity in last 4 weeks: Heavy DIY (eg: weeding, lawn mowing, carpentry, digging) | 0 | ukbb | European | -0.1187 | 0.0449 | 0.0082 |
| UL | Why stopped smoking: Doctors advice | 0 | ukbb | European | 0.267 | 0.1012 | 0.0083 |
| UL | Diagnoses - main ICD10: I20 Angina pectoris | 0 | ukbb | European | 0.192 | 0.0728 | 0.0083 |
| UL | Forced expiratory volume in 1 second (FEV1) | 21946350 | lung function | European | -0.2041 | 0.0776 | 0.0085 |
| UL | Forced expiratory volume in 1 second | 0 | lung function | European | -0.125 | 0.0476 | 0.0087 |
| UL | Other serious medical condition/disability diagnosed by doctor | 0 | ukbb | European | 0.1257 | 0.048 | 0.0089 |
| UL | Forced Vital capacity(FVC) | 28166213 | lung function | European | -0.11 | 0.0423 | 0.0093 |
| UL | Shortness of breath walking on level ground | 0 | ukbb | European | 0.1508 | 0.0582 | 0.0095 |
| UL | Pain type(s) experienced in last month: Neck or shoulder pain | 0 | ukbb | European | 0.1125 | 0.0435 | 0.0098 |
| UL | Fathers age at death | 0 | ukbb | European | -0.1419 | 0.055 | 0.0099 |
| UL | Relative age of first facial hair | 0 | ukbb | European | -0.0931 | 0.0363 | 0.0102 |
| UL | Snoring | 0 | ukbb | European | -0.099 | 0.0386 | 0.0104 |
| UL | Illnesses of mother: Breast cancer | 0 | ukbb | European | 0.2278 | 0.089 | 0.0105 |
| UL | Medication for cholesterol, blood pressure or diabetes: Insulin | 0 | ukbb | European | 0.3305 | 0.1291 | 0.0105 |
| UL | Eye problems/disorders: Diabetes related eye disease | 0 | ukbb | European | 0.2156 | 0.0845 | 0.0107 |
| UL | Time spent watching television (TV) | 0 | ukbb | European | 0.0805 | 0.0316 | 0.0108 |
| UL | Tense / highly strung | 0 | ukbb | European | 0.0909 | 0.0358 | 0.011 |
| UL | Illnesses of mother: None of the above (group 2) | 0 | ukbb | European | -0.2189 | 0.0861 | 0.011 |
| UL | Qualifications: None of the above | 0 | ukbb | European | 0.0807 | 0.0318 | 0.0111 |
| UL | Femoral neck bone mineral density | 22504420 | bone | Mixed | 0.1103 | 0.0435 | 0.0112 |
| UL | Relative age voice broke | 0 | ukbb | European | -0.11 | 0.0436 | 0.0118 |
| UL | Fasting insulin main effect | 22581228 | glycemic | European | 0.1854 | 0.0739 | 0.0121 |
| UL | Average weekly red wine intake | 0 | ukbb | European | -0.1034 | 0.0412 | 0.0121 |
| UL | Diagnoses - main ICD10: K52 Other non-infective gastro-enteritis and colitis | 0 | ukbb | European | 0.2529 | 0.1008 | 0.0121 |
| UL | Pulse wave peak to peak time | 0 | ukbb | European | -0.1579 | 0.063 | 0.0123 |
| UL | Trunk fat mass | 0 | ukbb | European | 0.0672 | 0.027 | 0.0127 |
| UL | Mono-unsaturated fatty acids | 27005778 | metabolites | European | 0.2561 | 0.1039 | 0.0137 |
| UL | Guilty feelings | 0 | ukbb | European | 0.0894 | 0.0364 | 0.0139 |
| UL | Urinary albumin-to-creatinine ratio | 26631737 | kidney | European | 0.2176 | 0.0888 | 0.0143 |
| UL | Diagnoses - main ICD10: R14 Flatulence and related conditions | 0 | ukbb | European | 0.333 | 0.1359 | 0.0143 |
| UL | Lung adenocarcinoma | 27488534 | cancer | European | 0.3038 | 0.1241 | 0.0144 |
| UL | Duration of light DIY | 0 | ukbb | European | 0.1299 | 0.0533 | 0.0148 |
| UL | Diagnoses - main ICD10: J34 Other disorders of nose and nasal sinuses | 0 | ukbb | European | 0.4087 | 0.1682 | 0.0151 |
| UL | Mean Caudate | 25607358 | brain,volume | European | -0.1641 | 0.0677 | 0.0153 |
| UL | Major depressive disorder | 22472876 | psychiatric | European | 0.1797 | 0.0743 | 0.0156 |
| UL | Phospholipids in large VLDL | 27005778 | metabolites | European | 0.1923 | 0.0797 | 0.0158 |
| UL | Acetate | 27005778 | metabolites | European | -0.2911 | 0.1217 | 0.0167 |
| UL | Daytime dozing / sleeping (narcolepsy) | 0 | ukbb | European | 0.0954 | 0.04 | 0.0172 |
| UL | Wheeze or whistling in the chest in last year | 0 | ukbb | European | 0.0888 | 0.0373 | 0.0173 |
| UL | Forced vital capacity | 0 | lung function | European | -0.0708 | 0.0298 | 0.0174 |

| trait1 | trait2 | PMID | Category | ethnicity | $r_g$ | se | p |
|---|---|---|---|---|---|---|---|
| UL | Triglycerides in very large HDL | 27005778 | metabolites | European | 0.2449 | 0.104 | 0.0186 |
| UL | Qualifications: Other professional qualifications eg: nursing, teaching | 0 | ukbb | European | -0.0886 | 0.0379 | 0.0195 |
| UL | Chest pain or discomfort | 0 | ukbb | European | 0.1059 | 0.0454 | 0.0197 |
| UL | Types of physical activity in last 4 weeks: Other exercises (eg: swimming, cycling, keep fit, bowling) | 0 | ukbb | European | -0.094 | 0.0405 | 0.0202 |
| UL | Total lipids in large VLDL | 27005778 | metabolites | European | 0.1867 | 0.0808 | 0.0209 |
| UL | Time spent driving | 0 | ukbb | European | 0.1053 | 0.0457 | 0.0213 |
| UL | Had major operations | 0 | ukbb | European | 0.1539 | 0.0671 | 0.0219 |
| UL | Heel bone mineral density (BMD) T-score, automated (right) | 0 | ukbb | European | 0.0914 | 0.0403 | 0.0232 |
| UL | Forced expiratory volume in 1 second (FEV1) | 26635082 | lung function | European | -0.1372 | 0.0605 | 0.0234 |
| UL | Tinnitus: Yes, now some of the time | 0 | ukbb | European | 0.331 | 0.1466 | 0.024 |
| UL | Diagnoses - main ICD10: K57 Diverticular disease of intestine | 0 | ukbb | European | 0.1326 | 0.059 | 0.0245 |
| UL | Illnesses of father: High blood pressure | 0 | ukbb | European | 0.1337 | 0.0596 | 0.025 |
| UL | Leg fat percentage (left) | 0 | ukbb | European | 0.0628 | 0.028 | 0.0251 |
| UL | Duration of heavy DIY | 0 | ukbb | European | 0.1326 | 0.0594 | 0.0257 |
| UL | Forced expiratory volume in 1 second | 0 | lung function | European | -0.067 | 0.0302 | 0.0263 |
| UL | Total lipids in very large VLDL | 27005778 | metabolites | European | 0.1818 | 0.0822 | 0.027 |
| UL | Type 2 Diabetes | 22885922 | glycemic | European | 0.1178 | 0.0533 | 0.0272 |
| UL | Noisy workplace | 0 | ukbb | European | 0.1153 | 0.0523 | 0.0275 |
| UL | Hand grip strength (left) | 0 | ukbb | European | -0.0699 | 0.0317 | 0.0276 |
| UL | Alcohol intake versus 10 years previously | 0 | ukbb | European | 0.1042 | 0.0475 | 0.0283 |
| UL | Serum total triglycerides | 27005778 | metabolites | European | 0.1666 | 0.076 | 0.0285 |
| UL | Leg fat percentage (right) | 0 | ukbb | European | 0.0619 | 0.0283 | 0.0288 |
| UL | Home area population density - urban or rural: Scotland - Large Urban Area | 0 | ukbb | European | -0.2139 | 0.0995 | 0.0315 |
| UL | Total lipids in chylomicrons and largest VLDL particles | 27005778 | metabolites | European | 0.1949 | 0.0911 | 0.0324 |
| UL | Ever had bowel cancer screening | 0 | ukbb | European | 0.1424 | 0.0666 | 0.0325 |
| UL | Job involves mainly walking or standing | 0 | ukbb | European | 0.0884 | 0.0414 | 0.0327 |
| UL | Usual walking pace | 0 | ukbb | European | -0.0704 | 0.033 | 0.0329 |
| UL | Obesity class 2 | 23563607 | anthropometric | European | 0.1035 | 0.0488 | 0.034 |
| UL | Insomnia | 28604731 | sleeping | European | 0.1228 | 0.0581 | 0.0345 |
| UL | Diagnoses - main ICD10: Z09 Follow-up examination after treatment for conditions other than malignant neoplasms | 0 | ukbb | European | 0.2268 | 0.1075 | 0.0349 |
| UL | Mean diameter for VLDL particles | 27005778 | metabolites | European | 0.165 | 0.0784 | 0.0353 |
| UL | Total lipids in medium VLDL | 27005778 | metabolites | European | 0.1631 | 0.0783 | 0.0372 |
| UL | Triglycerides in very small VLDL | 27005778 | metabolites | European | 0.1613 | 0.0775 | 0.0373 |
| UL | Types of transport used (excluding work): Cycle | 0 | ukbb | European | -0.0961 | 0.0462 | 0.0376 |
| UL | Illness, injury, bereavement, stress in last 2 years: Serious illness, injury or assault to yourself | 0 | ukbb | European | 0.1302 | 0.0627 | 0.0377 |
| UL | Pain type(s) experienced in last month: Facial pain | 0 | ukbb | European | 0.1782 | 0.0859 | 0.038 |
| UL | Types of transport used (excluding work): Walk | 0 | ukbb | European | -0.0908 | 0.0438 | 0.0382 |
| UL | Potassium in urine | 0 | ukbb | European | 0.0915 | 0.0442 | 0.0386 |
| UL | Hand grip strength (right) | 0 | ukbb | European | -0.0639 | 0.0309 | 0.0388 |
| UL | Concentration of very large VLDL particles | 27005778 | metabolites | European | 0.1767 | 0.0861 | 0.04 |
| UL | Triglycerides in large VLDL | 27005778 | metabolites | European | 0.1667 | 0.0812 | 0.0402 |
| UL | Medication for pain relief, constipation, heartburn: Omeprazole (e.g. Zanprol) | 0 | ukbb | European | 0.1275 | 0.0624 | 0.0411 |

| trait1 | trait2 | PMID | Category | ethnicity | $r_g$ | se | p |
|---|---|---|---|---|---|---|---|
| UL | Free cholesterol in medium VLDL | 27005778 | metabolites | European | 0.1665 | 0.0816 | 0.0415 |
| UL | Multiple sclerosis | 21833088 | autoimmune | European | 0.2827 | 0.1388 | 0.0417 |
| UL | Illnesses of father: Diabetes | 0 | ukbb | European | 0.1104 | 0.0545 | 0.0429 |
| UL | Total cholesterol in medium VLDL | 27005778 | metabolites | European | 0.1576 | 0.0779 | 0.0431 |
| UL | Average weekly spirits intake | 0 | ukbb | European | 0.1045 | 0.0519 | 0.0439 |
| UL | Alcohol drinker status: Previous | 0 | ukbb | European | 0.1119 | 0.0559 | 0.0454 |
| UL | Falls in the last year | 0 | ukbb | European | 0.092 | 0.0461 | 0.0461 |
| UL | Free cholesterol in large VLDL | 27005778 | metabolites | European | 0.1555 | 0.0785 | 0.0475 |
| UL | Concentration of small VLDL particles | 27005778 | metabolites | European | 0.1521 | 0.0771 | 0.0486 |
| UL | Arm fat percentage (left) | 0 | ukbb | European | 0.0558 | 0.0283 | 0.0486 |
| UL | Concentration of large VLDL particles | 27005778 | metabolites | European | 0.1651 | 0.0838 | 0.0489 |
| UL | Exposure to tobacco smoke at home | 0 | ukbb | European | 0.1276 | 0.0649 | 0.0492 |

**Table S9. Results of bi-directional two-sample Mendelian randomization.**

Two-sample Mendelian randomization was performed using 'TwoSampleMR' R library[7]. Genetic instruments for UL were extracted from the GWAS results obtained in FinnGen. For other traits, the instruments were extracted from the GWAS database provided by the MRC Integrative Epidemiology Unit available at https://gwas.mrcieu.ac.uk/. LD pruning was completed using standard thresholds (r2=0.001, clumping window=10kb).

| Analysis direction | Trait | Method | $N_{snp}$ | Causal estimate scale | Causal estimate | Lo CI | Up CI | P | $P_{HET}$ | Egger intercept | $P_{PLE}$ |
|---|---|---|---|---|---|---|---|---|---|---|---|
| trait -> UL | Age when periods started (menarche) \|\| id:ukb-b-3768 | Inverse variance weighted | 175 | OR | 0.730 | 0.635 | 0.839 | 9.05E-06 | 2.02E-05 | | |
| trait -> UL | Age when periods started (menarche) \|\| id:ukb-b-3768 | MR Egger | 175 | OR | 0.844 | 0.570 | 1.250 | 0.399 | 1.91E-05 | -0.0030 | 0.439 |
| UL -> trait | Age when periods started (menarche) \|\| id:ukb-b-3768 | Inverse variance weighted | 26 | BETA | 0.016 | 0.0043 | 0.028 | 0.0073 | 0.0018 | | |
| UL -> trait | Age when periods started (menarche) \|\| id:ukb-b-3768 | MR Egger | 26 | BETA | 0.020 | -0.0013 | 0.041 | 0.079 | 0.0013 | -0.00063 | 0.675 |
| trait -> UL | Basal metabolic rate \|\| id:ukb-b-16446 | Inverse variance weighted | 487 | OR | 1.243 | 1.082 | 1.429 | 0.0022 | 2.00E-39 | | |
| trait -> UL | Basal metabolic rate \|\| id:ukb-b-16446 | MR Egger | 487 | OR | 1.359 | 0.979 | 1.887 | 0.067 | 1.67E-39 | -0.0014 | 0.556 |
| UL -> trait | Basal metabolic rate \|\| id:ukb-b-16446 | Inverse variance weighted | 26 | BETA | 0.020 | 0.0009 | 0.039 | 0.040 | 5.49E-55 | | |
| UL -> trait | Basal metabolic rate \|\| id:ukb-b-16446 | MR Egger | 26 | BETA | 0.039 | 0.0055 | 0.072 | 0.032 | 6.66E-51 | -0.0031 | 0.192 |
| trait -> UL | Bilateral oophorectomy (both ovaries removed) \|\| id:ukb-b-17868 | Inverse variance weighted | 5 | OR | 3661619 | 64082 | 2.09E+8 | 2.44E-13 | 0.0043 | | |
| trait -> UL | Bilateral oophorectomy (both ovaries removed) \|\| id:ukb-b-17868 | MR Egger | 5 | OR | 115063 | 0.0054 | 2.47E+12 | 0.269 | 0.0025 | 0.024 | 0.705 |
| UL -> trait | Bilateral oophorectomy (both ovaries removed) \|\| id:ukb-b-17868 | Inverse variance weighted | 22 | BETA | 0.024 | 0.016 | 0.032 | 1.82E-09 | 1.09E-10 | | |
| UL -> trait | Bilateral oophorectomy (both ovaries removed) \|\| id:ukb-b-17868 | MR Egger | 22 | BETA | 0.041 | 0.018 | 0.063 | 0.0023 | 1.92E-09 | -0.0020 | 0.150 |
| trait -> UL | Body mass index (BMI) \|\| id:ukb-b-19953 | Inverse variance weighted | 399 | OR | 1.133 | 1.033 | 1.243 | 0.0084 | 3.42E-06 | | |
| trait -> UL | Body mass index (BMI) \|\| id:ukb-b-19953 | MR Egger | 399 | OR | 1.139 | 0.890 | 1.459 | 0.301 | 2.92E-06 | -0.00011 | 0.961 |
| UL -> trait | Body mass index (BMI) \|\| id:ukb-b-19953 | Inverse variance weighted | 26 | BETA | 0.004 | -0.017 | 0.025 | 0.722 | 2.15E-24 | | |
| UL -> trait | Body mass index (BMI) \|\| id:ukb-b-19953 | MR Egger | 26 | BETA | 0.025 | -0.011 | 0.062 | 0.185 | 2.49E-22 | -0.0036 | 0.173 |
| UL -> trait | Depressive symptoms \|\| id:ieu-a-1000 | Inverse variance weighted | 21 | BETA | -0.005 | -0.024 | 0.013 | 0.573 | 0.909 | | |
| UL -> trait | Depressive symptoms \|\| id:ieu-a-1000 | MR Egger | 21 | BETA | 0.005 | -0.054 | 0.065 | 0.863 | 0.883 | -0.0012 | 0.716 |
| UL -> trait | Diagnoses - main ICD10: N92.0 Excessive and frequent menstruation with regular cycle \|\| id:ukb-b-11572 | Inverse variance weighted | 20 | BETA | 0.004 | 0.002 | 0.005 | 1.07E-04 | 0.00273 | | |
| UL -> trait | Diagnoses - main ICD10: N92.0 Excessive and frequent menstruation with regular cycle \|\| id:ukb-b-11572 | MR Egger | 20 | BETA | 0.003 | -0.002 | 0.009 | 0.280 | 0.00176 | 4.80E-05 | 0.881 |
| trait -> UL | diastolic blood pressure \|\| id:ieu-b-39 | Inverse variance weighted | 415 | OR | 1.016 | 1.006 | 1.027 | 0.0026 | 4.75E-21 | | |
| trait -> UL | diastolic blood pressure \|\| id:ieu-b-39 | MR Egger | 415 | OR | 1.012 | 0.987 | 1.039 | 0.348 | 3.70E-21 | 0.00081 | 0.732 |

| Analysis direction | Trait | Method | $N_{snp}$ | Causal estimate scale | Causal estimate | Lo CI | Up CI | P | $P_{HET}$ | Egger intercept | $P_{PLE}$ |
|---|---|---|---|---|---|---|---|---|---|---|---|
| UL -> trait | diastolic blood pressure ‖ id:ieu-b-39 | Inverse variance weighted | 24 | BETA | 0.369 | 0.149 | 0.588 | 0.0010 | 2.08E-30 | | |
| UL -> trait | diastolic blood pressure ‖ id:ieu-b-39 | MR Egger | 24 | BETA | 0.625 | 0.202 | 1.049 | 0.0084 | 9.05E-28 | -0.039 | 0.181 |
| trait -> UL | Ever had hysterectomy (womb removed) ‖ id:ukb-b-6445 | Inverse variance weighted | 3 | OR | 2092679472 | 394 | 1.11E+16 | 0.0066 | 4.01E-14 | | |
| trait -> UL | Ever had hysterectomy (womb removed) ‖ id:ukb-b-6445 | MR Egger | 3 | OR | 3.96E+16 | 3.92E-41 | 4.00E+73 | 0.670 | 2.66E-14 | -0.105 | 0.842 |
| UL -> trait | Ever had hysterectomy (womb removed) ‖ id:ukb-b-6445 | Inverse variance weighted | 21 | BETA | 0.022 | 0.016 | 0.028 | 9.81E-14 | 0.0028 | | |
| UL -> trait | Ever had hysterectomy (womb removed) ‖ id:ukb-b-6445 | MR Egger | 21 | BETA | 0.031 | 0.013 | 0.048 | 0.0028 | 0.0034 | -0.0010 | 0.330 |
| trait -> UL | Ever used hormone-replacement therapy (HRT) ‖ id:ukb-b-18541 | Inverse variance weighted | 5 | OR | 0.124 | 0.008 | 1.873 | 0.132 | 0.00014 | | |
| trait -> UL | Ever used hormone-replacement therapy (HRT) ‖ id:ukb-b-18541 | MR Egger | 5 | OR | 0.494 | 0.00041 | 598 | 0.858 | 8.25E-05 | -0.020 | 0.701 |
| UL -> trait | Ever used hormone-replacement therapy (HRT) ‖ id:ukb-b-18541 | Inverse variance weighted | 25 | BETA | 0.0029 | -0.0053 | 0.011 | 0.493 | 0.00060 | | |
| UL -> trait | Ever used hormone-replacement therapy (HRT) ‖ id:ukb-b-18541 | MR Egger | 25 | BETA | 0.0079 | -0.0068 | 0.023 | 0.303 | 0.00060 | -0.00084 | 0.425 |
| trait -> UL | Frequency of tiredness / lethargy in last 2 weeks ‖ id:ukb-b-929 | Inverse variance weighted | 38 | OR | 1.065 | 0.719 | 1.576 | 0.754 | 0.413 | | |
| trait -> UL | Frequency of tiredness / lethargy in last 2 weeks ‖ id:ukb-b-929 | MR Egger | 38 | OR | 0.330 | 0.049 | 2.209 | 0.261 | 0.437 | 0.014 | 0.225 |
| UL -> trait | Frequency of tiredness / lethargy in last 2 weeks ‖ id:ukb-b-929 | Inverse variance weighted | 26 | BETA | -0.0033 | -0.013 | 0.0060 | 0.483 | 0.0088 | | |
| UL -> trait | Frequency of tiredness / lethargy in last 2 weeks ‖ id:ukb-b-929 | MR Egger | 26 | BETA | 0.0017 | -0.015 | 0.018 | 0.841 | 0.0079 | -0.00084 | 0.480 |
| trait -> UL | Impedance of whole body ‖ id:ukb-b-19921 | Inverse variance weighted | 463 | OR | 0.795 | 0.692 | 0.912 | 0.0010 | 3.80E-44 | | |
| trait -> UL | Impedance of whole body ‖ id:ukb-b-19921 | MR Egger | 463 | OR | 0.584 | 0.408 | 0.836 | 0.0034 | 1.89E-43 | 0.0049 | 0.069 |
| UL -> trait | Impedance of whole body ‖ id:ukb-b-19921 | Inverse variance weighted | 26 | BETA | -0.013 | -0.039 | 0.012 | 0.297 | 1.62E-77 | | |
| UL -> trait | Impedance of whole body ‖ id:ukb-b-19921 | MR Egger | 26 | BETA | -0.034 | -0.079 | 0.011 | 0.149 | 7.37E-74 | 0.0034 | 0.286 |
| trait -> UL | Neuroticism score ‖ id:ukb-b-4630 | Inverse variance weighted | 107 | OR | 1.050 | 0.988 | 1.115 | 0.114 | 0.011 | | |
| trait -> UL | Neuroticism score ‖ id:ukb-b-4630 | MR Egger | 107 | OR | 1.158 | 0.835 | 1.607 | 0.380 | 0.0096 | -0.0053 | 0.550 |
| UL -> trait | Neuroticism score ‖ id:ukb-b-4630 | Inverse variance weighted | 26 | BETA | 0.011 | -0.022 | 0.043 | 0.517 | 0.215 | | |
| UL -> trait | Neuroticism score ‖ id:ukb-b-4630 | MR Egger | 26 | BETA | 0.050 | -0.006 | 0.105 | 0.093 | 0.298 | -0.0065 | 0.110 |
| trait -> UL | systolic blood pressure ‖ id:ieu-b-38 | Inverse variance weighted | 405 | OR | 1.007 | 1.001 | 1.013 | 0.026 | 1.34E-14 | | |
| trait -> UL | systolic blood pressure ‖ id:ieu-b-38 | MR Egger | 405 | OR | 1.015 | 1.000 | 1.030 | 0.054 | 1.58E-14 | -0.0027 | 0.257 |
| UL -> trait | systolic blood pressure ‖ id:ieu-b-38 | Inverse variance weighted | 23 | BETA | 0.346 | -0.085 | 0.777 | 0.116 | 1.41E-38 | | |
| UL -> trait | systolic blood pressure ‖ id:ieu-b-38 | MR Egger | 23 | BETA | -0.086 | -0.917 | 0.745 | 0.841 | 4.29E-36 | 0.065 | 0.248 |

| Analysis direction | Trait | Method | $N_{snp}$ | Causal estimate scale | Causal estimate | Lo CI | Up CI | P | $P_{HET}$ | Egger intercept | $P_{PLE}$ |
|---|---|---|---|---|---|---|---|---|---|---|---|
| trait -> UL | Triglycerides ǁ id:ieu-a-302 | Inverse variance weighted | 53 | OR | 1.088 | 0.973 | 1.217 | 0.139 | 2.17E-05 | | |
| trait -> UL | Triglycerides ǁ id:ieu-a-302 | MR Egger | 53 | OR | 1.119 | 0.936 | 1.338 | 0.221 | 1.63E-05 | -0.0018 | 0.690 |
| UL -> trait | Triglycerides ǁ id:ieu-a-302 | Inverse variance weighted | 17 | BETA | 0.0024 | -0.031 | 0.035 | 0.886 | 0.076 | | |
| UL -> trait | Triglycerides ǁ id:ieu-a-302 | MR Egger | 17 | BETA | 0.064 | -0.074 | 0.203 | 0.377 | 0.076 | -0.0066 | 0.381 |
| trait -> UL | Waist circumference ǁ id:ukb-b-9405 | Inverse variance weighted | 326 | OR | 1.194 | 1.054 | 1.352 | 0.0052 | 2.61E-09 | | |
| trait -> UL | Waist circumference ǁ id:ukb-b-9405 | MR Egger | 326 | OR | 1.250 | 0.881 | 1.775 | 0.212 | 2.14E-09 | -0.0008 | 0.782 |
| UL -> trait | Waist circumference ǁ id:ukb-b-9405 | Inverse variance weighted | 26 | BETA | 0.013 | -0.0028 | 0.028 | 0.109 | 4.49E-14 | | |
| UL -> trait | Waist circumference ǁ id:ukb-b-9405 | MR Egger | 26 | BETA | 0.029 | 0.0016 | 0.056 | 0.049 | 7.41E-13 | -0.0027 | 0.174 |
| trait -> UL | Whole body fat mass ǁ id:ukb-b-19393 | Inverse variance weighted | 387 | OR | 1.036 | 0.943 | 1.137 | 0.463 | 5.68E-05 | | |
| trait -> UL | Whole body fat mass ǁ id:ukb-b-19393 | MR Egger | 387 | OR | 1.170 | 0.902 | 1.519 | 0.238 | 5.77E-05 | -0.0023 | 0.326 |
| UL -> trait | Whole body fat mass ǁ id:ukb-b-19393 | Inverse variance weighted | 26 | BETA | 0.0056 | -0.010 | 0.021 | 0.481 | 1.41E-10 | | |
| UL -> trait | Whole body fat mass ǁ id:ukb-b-19393 | MR Egger | 26 | BETA | 0.024 | -0.0023 | 0.051 | 0.086 | 3.41E-09 | -0.0031 | 0.108 |
| trait -> UL | Whole body fat-free mass ǁ id:ukb-b-13354 | Inverse variance weighted | 486 | OR | 1.239 | 1.079 | 1.423 | 0.0024 | 6.68E-33 | | |
| trait -> UL | Whole body fat-free mass ǁ id:ukb-b-13354 | MR Egger | 486 | OR | 1.398 | 1.013 | 1.929 | 0.042 | 6.53E-33 | -0.0018 | 0.419 |
| UL -> trait | Whole body fat-free mass ǁ id:ukb-b-13354 | Inverse variance weighted | 26 | BETA | 0.020 | 0.0014 | 0.039 | 0.035 | 7.97E-62 | | |
| UL -> trait | Whole body fat-free mass ǁ id:ukb-b-13354 | MR Egger | 26 | BETA | 0.038 | 0.0052 | 0.071 | 0.033 | 1.26E-57 | -0.0030 | 0.210 |
| trait -> UL | Whole body water mass ǁ id:ukb-b-14540 | Inverse variance weighted | 500 | OR | 1.215 | 1.058 | 1.396 | 0.0058 | 3.19E-35 | | |
| trait -> UL | Whole body water mass ǁ id:ukb-b-14540 | MR Egger | 500 | OR | 1.334 | 0.965 | 1.843 | 0.081 | 2.74E-35 | -0.0014 | 0.532 |
| UL -> trait | Whole body water mass ǁ id:ukb-b-14540 | Inverse variance weighted | 26 | BETA | 0.020 | 0.0013 | 0.040 | 0.037 | 1.13E-63 | | |
| UL -> trait | Whole body water mass ǁ id:ukb-b-14540 | MR Egger | 26 | BETA | 0.039 | 0.0050 | 0.072 | 0.034 | 2.24E-59 | -0.0030 | 0.211 |
| trait -> UL | Years of schooling ǁ id:ieu-a-1239 | Inverse variance weighted | 287 | OR | 0.914 | 0.792 | 1.055 | 0.222 | 0.00017 | | |
| trait -> UL | Years of schooling ǁ id:ieu-a-1239 | MR Egger | 287 | OR | 0.777 | 0.445 | 1.356 | 0.375 | 0.00016 | 0.0023 | 0.553 |
| UL -> trait | Years of schooling ǁ id:ieu-a-1239 | Inverse variance weighted | 26 | BETA | -0.0051 | -0.014 | 0.0041 | 0.280 | 0.0069 | | |
| UL -> trait | Years of schooling ǁ id:ieu-a-1239 | MR Egger | 26 | BETA | 0.0050 | -0.011 | 0.021 | 0.540 | 0.014 | -0.0017 | 0.144 |
